# Integration of human organoids single-cell transcriptomic profiles and human genetics repurposes critical cell type-specific drug targets for severe COVID-19

**DOI:** 10.1101/2023.07.03.23292161

**Authors:** Yunlong Ma, Yijun Zhou, Dingping Jiang, Wei Dai, Jingjing Li, Chunyu Deng, Cheng Chen, Gongwei Zheng, Yaru Zhang, Fei Qiu, Haojun Sun, Shilai Xing, Haijun Han, Jia Qu, Nan Wu, Yinghao Yao, Jianzhong Su

## Abstract

Human organoids recapitulate the cell type diversity and function of their primary organs holding tremendous potentials for basic and translational research. Advances in single-cell RNA sequencing (scRNA-seq) technology and genome-wide association study (GWAS) have accelerated the biological and therapeutic interpretation of trait-relevant cell types or states. Here, we constructed a computational framework to integrate atlas-level organoid scRNA-seq data, GWAS summary statistics, expression quantitative trait loci, and gene-drug interaction data for distinguishing critical cell populations and drug targets relevant to COVID-19 severity. We found that 39 cell types across eight kinds of organoids were significantly associated with COVID-19 outcomes. Notably, subset of lung mesenchymal stem cells (MSCs) increased proximity with fibroblasts predisposed to repair COVID-19-damaged lung tissue. Brain endothelial cell subset exhibited significant associations with severe COVID-19, and this cell subset showed a notable increase in cell-to-cell interactions with other brain cell types, including microglia. We repurposed 33 druggable genes, including *IFNAR2*, *TYK2*, and *VIPR2*, and their interacting drugs for COVID-19 in a cell-type-specific manner. Overall, our results showcase that host genetic determinants have cellular specific contribution to COVID-19 severity, and identification of cell type-specific drug targets may facilitate to develop effective therapeutics for treating severe COVID-19 and its complications.

## Introduction

The coronavirus disease 2019 (COVID-19), caused by the novel severe acute respiratory syndrome coronavirus 2 (SARS-CoV-2), is characterized by heterogeneous clinical manifestations ranging from asymptomatic to severe disruptions [1]. Multiple lines of evidence have demonstrated that increased number of severe COVID-19 patients have significant extrapulmonary complications [2, 3], which deteriorate the condition of infected patients. Although vaccines have now been developed for preventing COVID-19 infection, it is unclear how long it will take to gain herd immunity, or if novel mutations will enable SARS-CoV-2 to escape the protection from current vaccines [4]. To date, there is still no specific antiviral drugs to target SARS-CoV-2 for alleviating established diseases [5]. Thus, it is an urgent need to rapidly highlight existing drugs that can be repurposed for management in severe COVID-19 and its complications.

Human organoids, self-organizing three-dimensional (3D) cultured systems, recapitulate numerous core features of human organ development and biological functions. Hence, these 3D *in vitro* structures hold tremendous potential as avatars for preclinical drug developments and interventional experiments that are difficult or impossible to carry out in human subjects [6, 7]. Although having incredibly powerful capabilities, human organoids are biomimetic and heterogeneous model systems with complicated cell types and states, which is intractable to analyze through the conventional technologies, e.g., immunohistochemistry. Advancing single-cell RNA sequencing (scRNA-seq) technique provides an unprecedented opportunity to dissect the cellular and molecular heterogeneity in primary human organs/tissues [8–10]. Compared with transcriptome measurements from bulk samples, single-cell sequencing methods not only generate cell states and transcription regulatory programs in these 3D model systems at single-cell resolution, but also gain insights into the disease-related processes and complex cellular interactions [11–13]. Since the COVID-19 outbreak, many scRNA-seq studies have demonstrated that numerous types of organoids, including lung, intestinal, kidney, brain, and choroid plexus organoids, enable to investigate the tropism of SARS-CoV-2 infection [11, 14, 15].

Genome-wide association studies (GWAS) have been widely used for identifying significant genotype-phenotype associations for complex diseases or traits [16]. To date, several GWASs have reported that a large amount of genetic variants show notable associations with COVID-19 severities [17–20]. Integrating GWAS summary statistics and expression quantitative trait loci (eQTL) data, recent studies have distinguished several candidates as putative drug targets for treating COVID-19 [4, 21, 22]. Moreover, linking genome-wide polygenic signals with single-cell expression measurements from scRNA-seq data has considerable potential to unveil critical cell types or subpopulations relevant to complex diseases [23]. Our and other recent studies [24–26] have identified numerous immune and lung cell types that are impacted by genetic variants associated with COVID-19; e.g., alveolar type 2 cells and CD8+T cells in lung [26], and CD16+ monocytes, megakaryocytes and memory CD8+ T cells in peripheral blood [24]. Nevertheless, these reported studies largely focused on predefined cell type annotations, which considerably ignored the intra-heterogeneity within cell types. To date, no atlas-level analysis of combining scRNA-seq data across multiple tissues and organs with GWAS summary statistics to systematically identify COVID-19-relevant cell populations and drug targets at a single-cell resolution.

In light of the vital role of human organoids in drug developments, we collected and unifiedly processed numerous scRNA-seq datasets across 10 kinds of human organoids with more than one million cells, and developed a computational framework to integrate these human organoids scRNA-seq data, GWAS summary statistics, eQTL data, and gene-drug interaction data for distinguishing critical cell types/subpopulations and drug targets relevant to severe COVID-19. We found that numerous cell types across different human organoids were remarkably associated with COVID-19 severities. Notably, we showed that prioritizing COVID-19-relevant cell type-specific gene-drug interacting pairs in lung MSCs, intestinal tuft cells, and brain endothelial cells might conduce to repurpose drugs for treating severe COVID-19 and accompanied complications.

## Results

### Computational framework of COVID-19-relevant cell types and drug repositioning

To facilitate the data integration and minimize the batch effects, we have built a unified pipeline to conduct re-alignment, quality control, and standard analysis of all human organoids (n = 1,159,206 cells) and fetal scRNA-seq datasets (n =223,334 cells, Supplementary Figure S1 and Table S1). To distinguish critical cell types/subpopulations and repurpose potential drugs and interacting targets for the treatment of severe COVID-19, we devise a computational framework to incorporate these organoids and fetal scRNA-seq data and large-scale meta-GWAS summary statistics on three COVID-19 phenotypes (i.e., very severe, hospitalized, and susceptible COVID-19; Figure 1, and Supplementary Table S2 and Figures S2-S3). There are three main sections: (1) integrating GWAS summary statistics with human organoids scRNA-seq datasets to genetically map trait-relevant single-cell landscapes for three COVID-19 outcomes (Figure 1A); (2) combining GWAS summary statistics with eQTL data in the GTEx database to identify putative risk genes and critical pathways associated with COVID-19 severities (Figure 1B); and (3) prioritization of cell type-specific gene-drug interaction pairs for treating severe COVID-19 and related complications at a fine-grained resolution (Figure 1C).

**Figure 1.**
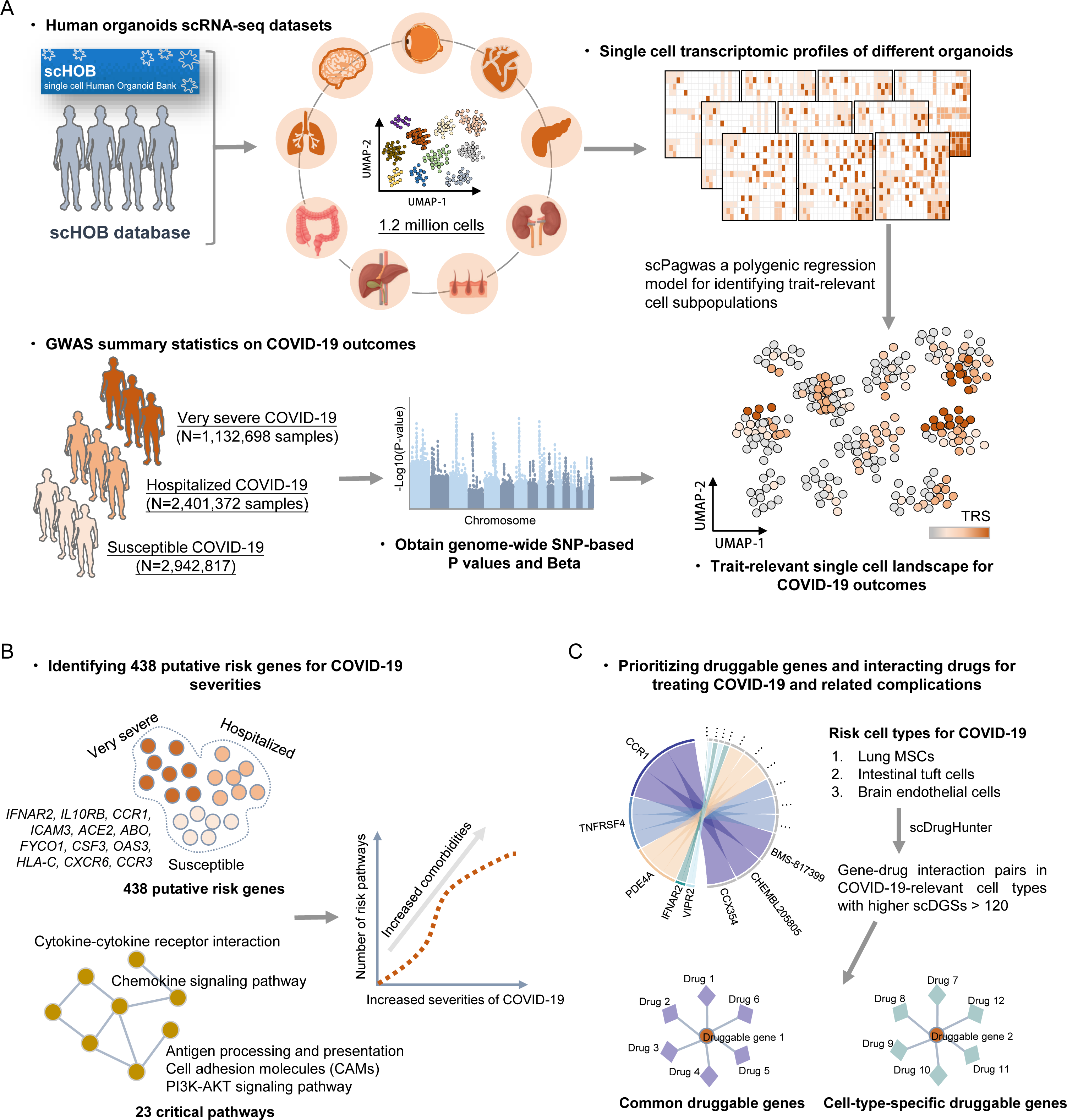
The workflow of integrative genomics analyses for COVID-19-relevant drug repositioning. A. Integration analysis of single-cell transcriptomic profiles in the scHOB database with GWAS summary statistics on three COVID-19 phenotypes. There were approximate 1.2 million cells from 10 kinds of human organoids (i.e., brain, eye, heart, lung, liver & bile duct, pancreas, kidney, intestine, and skin), and three GWAS datasets with more than two million samples downloaded from the COVID-19 Host Genetics Initiative. B. An increase in genetics-risk pathways and comorbidities for COVID-19 severities. C. Prioritization of druggable genes and interacting drugs for treating COVID-19 using the scDrugHunter method. Three COVID-19-relevant risk cell types (i.e, lung mesenchymal stem cell (MSCs), intestinal tuft cells, and brain endothelial cells) were leveraged as representative examples for searching druggable genes and interacting drugs, and comparison analysis were performed to find cell-type-common and cell-type-specific druggable genes for severe COVID-19.

### Systematic integrative analysis for discerning COVID-19-relevant cell types

We initially applied the scPagwas-based polygenic regression model [27] to incorporate genetic signals from GWAS summary statistics on three COVID-19 outcomes with single-cell transcriptomic profiles from 10 kinds of human organoids scRNA-seq data for identifying critical cell types relevant to COVID-19 severities. Among them, 39 cell types in 8 human organoids showed notable associations with at least one COVID-19-related phenotype (Figure 2A and Supplementary Table S3). Notably, there existed highly consistent results among very severe, hospitalized, and susceptible COVID-19 (rho = 0.99 and P = 2.58×10^-6^, rho = 0.948 and P = 3.4×10^-4^; Figure 2B-C and Supplementary Figure S4). As for lung organoids, the cell type of mesenchymal stem cells (MSCs) was significantly enriched for all three COVID-19 phenotypes (Figure 2A). Previous studies [28–30] have suggested that MSCs have a substantially therapeutic potential to improve the outcomes of COVID-19 patients by facilitating to repair lung-tissue injury for relieving acute pulmonary edema. Several recent clinical trials have been conducted to determine the positive effects of MSCs on the treatment of critically ill patients with coronavirus infection (Identifiers: NCT04898088, NCT04336254, and NCT04573270).

**Figure 2.**
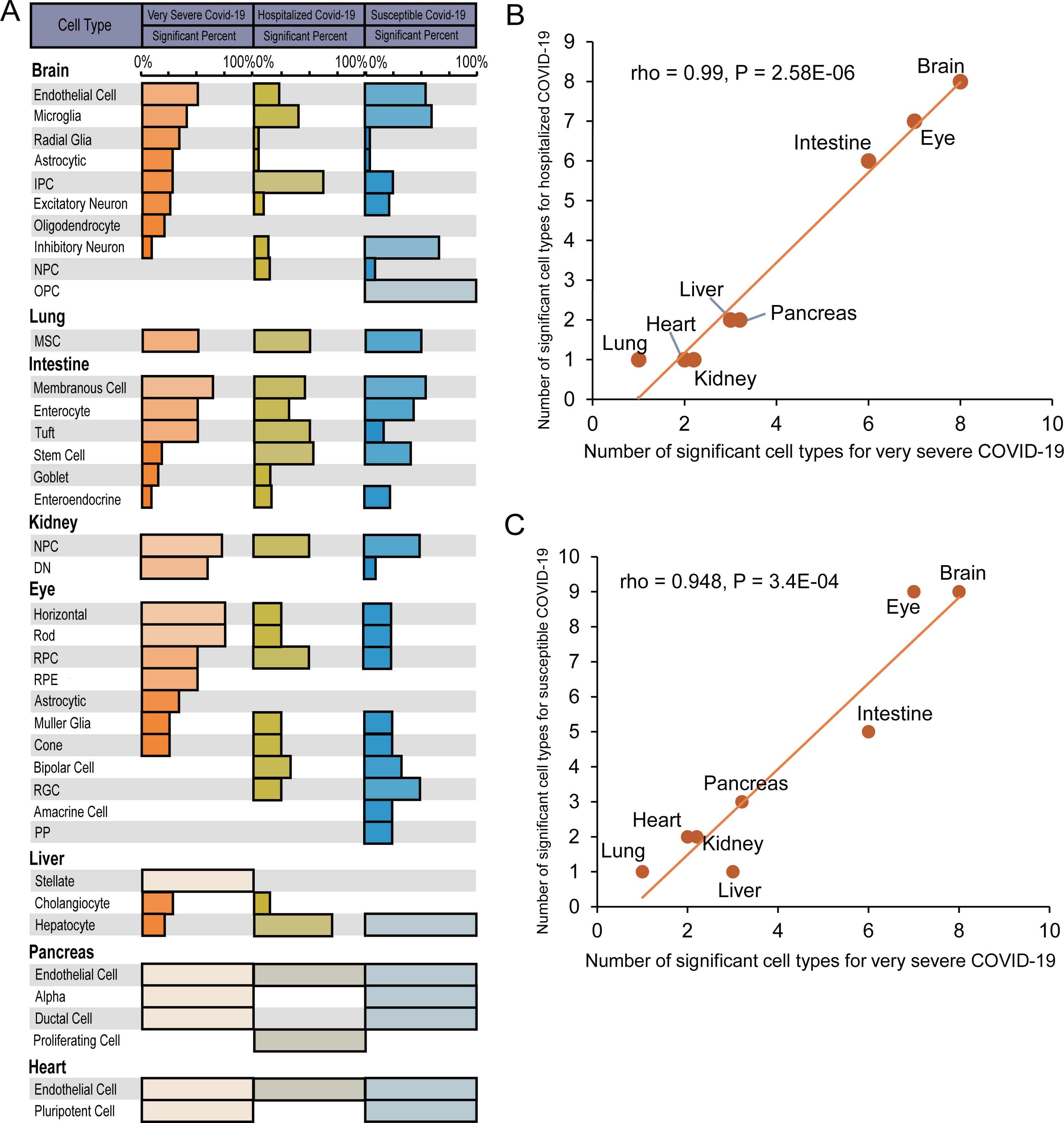
Significant associations between human organoids cell types and COVID-19 severities. A. Summary of 39 significant cell types in eight kinds of human organoids for three COVID-19 phenotypes. Bar plot represents the significant percent of each cell type in corresponding organoid with different scRNA-seq datasets. B. Correlation results of the number of significant cell types in eight human organoids between very severe COVID-19 (x axis) and hospitalized COVID-19 (y axis). C. Correlation results of the number of significant cell types in eight human organoids between very severe COVID-19 (x axis) and susceptible COVID-19 (y axis). The Pearson correlation analysis was used to calculate the correlation coefficients (rho). See also Supplementary Table S3.

There were six cell types including membranous cell, enterocyte, and tuft in intestine organoids associated with three COVID-19 phenotypes. Earlier studies have demonstrated that the angiotensin-converting enzyme 2 (ACE2) as a direct mediator regulate the SARS-CoV-2 entry into enterocytes in the gastrointestinal tract [31, 32], and COVID-19 patients often show gastrointestinal symptoms including vomiting, belly pain and diarrhea [33, 34]. For brain organoids, eight cell types, including endothelial cell and microglia, exhibited notable associations with severe COVID-19. Previous evidence have documented that cerebral endothelial dysfunction may be the cause of increased rates of cerebrovascular pathology relevant to COVID-19 [35], and severe COVID-19 patients experiencing a severe cytokine storm have considerable potential to induce microglia activation that lead to neurotoxicity [36, 37]. In addition, there existed seven cell types in eye organoids significantly associated with very severe COVID-19, including horizontal cells, rod, RPC, and cone. Our recent study [38] has indicated that host genetic factors play critical roles in facilitating SARS-CoV-2 infection in the ocular surface cells. For other organoids, we found that two cell types of nephron progenitor cell (NPC) and differentiating nephron (DN) in kidney organoids, three cell types of stellate, cholangiocyte, and hepatocyte in liver organoid, three cell types including endothelial cell and alpha in pancreas organoid, and two cell types of endothelial cell and pluripotent cell in heart organoids were significantly associated with COVID-19 severities (Figure 2A).

For validation, we used the RISmed method [39] that performs a PubMed search for resorting to reported evidence concerning the association between the trait of interest and a particular cell type. By counting the number of reported publications using the keyword pairs between COVID-19 and specific cell type, we computed the correlation between the number of publications and the significant percent of each cell type identified by scPagwas, and found significantly or suggestively positive correlations between scPagwas-identified cell-type results and PubMed search results across three COVID-19 phenotypes (Supplementary Figure S5A-F). Moreover, to replicate the biological findings from human organoids, we applied the same regression model to integrate GWAS summary data on very severe COVID-19 with human fetal scRNA-seq data with multiple tissues. The aforementioned observations remained reproducible in analyzing human fetal scRNAs-seq data (Supplementary Figure S6). For example, lung MSCs, intestinal tuft and enterocyte cells, eye cone and horizontal cells, and brain endothelial cells and microglia were notably associated with very severe COVID-19 in human fetal tissues. Taken together, we provide new insights for inferring critical cell types by which genetic variants influence COVID-19 severities.

### Transcriptome-wide association analysis identifies causal genes for three COVID-19 outcomes

To identify putative causal genes for COVID-19 severities, we applied the S-MultiXcan method [40] to integrate GWAS summary statistics and eQTL datasets based on 49 GTEx tissues. There were 243, 277, and 158 genes identified to be significantly associated with susceptible, hospitalized, and very severe COVID-19, respectively (total N = 438 genes, FDR < 0.05, Figure 3A, and Supplementary Figure S7 and Tables S4-S6). Many of these identified genes, including *ACE2*, *SLC6A20*, *OAS3*, *CCR1*, *CXCR6*, *IFNAR2*, *IL10RB*, and *DPP9*, have been reported to be associated with COVID-19 susceptibility in previous studies [20, 24, 41–45]. By overlapping these three COVID-19-associated gene sets, we found that 67 common genes whose genetically regulated expression have potentially important roles in COVD-19 initiation and progression (FDR < 0.05, Figure 3B and Supplementary Figure S8A and Table S7).

**Figure 3.**
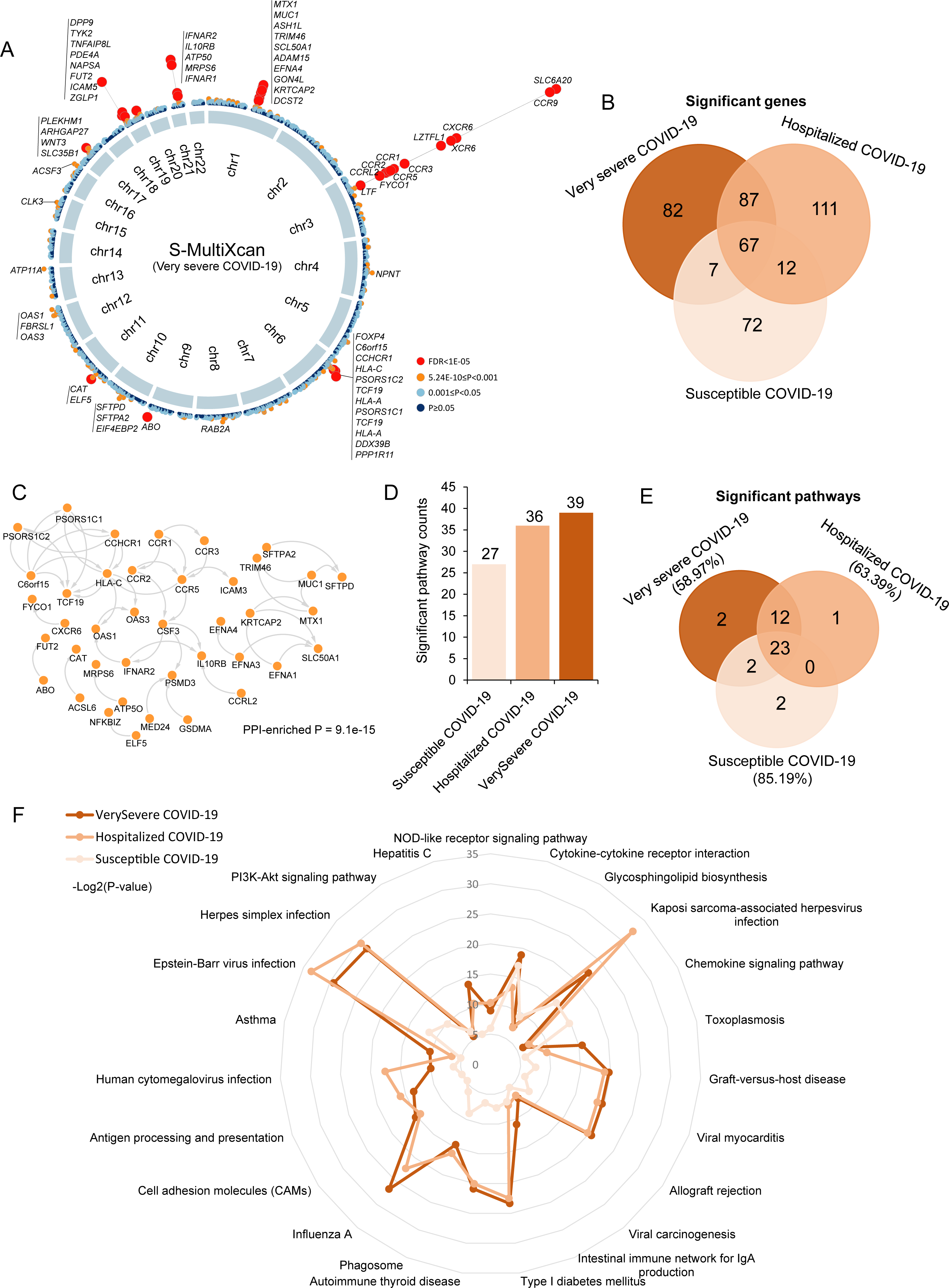
Risk genes and pathways associated with COVID-19 severities. A. Circus plot showing the results of the S-MultiXcan-based integrative analysis. The inner ring represents the 22 autosomal chromosomes (Chr1-22). In the out ring, a circular symbol demonstrates a specific gene with different color to mark the statistical significance of the gene for very severe COVID-19 (red marks FDR < 1E-05, orange marks 5.24E-10 ≤ P < 0.001, light blue marks 0.001 ≤ P ≤ 0.05, and dark blue marks P > 0.05). B. Venn diagram showing the overlapped risk genes across three COVID-19 phenotypes. C. Protein-protein interaction (PPI) network of 67 common risk genes based on the STRING database (v11.5, https://string-db.org/). D. Bar plot showing the counts of significant pathways enriched by using S-MultiXcan-identified risk genes in three COVID-19 phenotypes. E. Venn plot indicating the overlapped significant pathways across three COVID-19 phenotypes. F. Radar plot showing the significant level of 23 common pathways across three COVID-19 phenotypes. The P value of each pathway was negatively log-transformed (-Log2(P)) for visualization. See also Supplementary Tables S4-S6.

Network enrichment analysis exhibited that 40 of 67 common genes were significantly enriched in a protein-protein interaction (PPI) subnetwork (enriched P = 9.1×10^-15^, Figure 3C and Supplementary Figure S8B-C), which is in line with the consensus that disease-causing genes are more likely to be interacted [46, 47]. By conducting S-PrediXcan analyses of lung and blood tissues that were most relevant to SARS-CoV-2 infection, 280 of 438 risk genes (63.93%) identified from S-MultiXcan-based analyses were validated to be relevant to at least one COVID-19 outcome (P < 0.05, Supplementary Figure S9 and Tables S8-S9). Moreover, using MAGMA as an independent technique for validation (see Supplementary Methods), we found that there was a high consistence between results from MAGMA and S-MultiXcan analyses for three COVID-19 phenotypes (JSI = 0.28∼0.31, empirical P < 1×10^-5^, Supplementary Figures S10-S12 and Table S10).

Furthermore, we performed pathway-based enrichment analyses for three S-MultiXcan-identified gene sets to enrich critical pathways implicated in COVID-19 severities. We observed that the number of significant pathways was elevated with increased severities of COVID-19 (Figure 3D and Supplementary Figure S13A-C), which is consistent with the findings in an earlier study [24]. There was a large proportion of significant pathways (n = 23) in common among susceptible, hospitalized, and very severe COVID-19 (Figure 3E and Supplementary Table S11). We also noticed that the significant level of these common pathways showed an increased notable pattern with the increase of COVID-19 severities (FDR < 0.05, Figure 3F). Several of these pathways, including cytokine-cytokine receptor interaction and chemokine signaling pathway, have been documented to involve in the COVID-19 susceptibility in previous studies [20, 24, 48]. In sum, our integrative genomic analysis identifies that 438 risk genes involved in critical biological pathways show notable associations with COVID-19 severities.

### Genetic correlations between three COVID-19 outcomes and complex diseases

Previous epidemiologic and clinical studies have documented that the clinical manifestations of COVID-19 are heterogeneous, and many of COVD-19 cases are identified as having at least one comorbidity, including hypertension, diabetes, and other cerebrovascular, cardiovascular, and gastrointestinal complications, which may lead to poorer clinical outcomes [49, 50]. Given the high genetic heritability of these putative complications, we calculated the genetic correlations of 66 diseases/traits from six main disease categories with three COVID-19 phenotypes using the LDSC method[51]. We found that 29 of them (43.94%), including anorexia nervosa, attention deficit hyperactivity disorder, multiple sclerosis, neuroticism, ischemic stroke, cognitive performance, hypertension, type 2 diabetes, and pulmonary embolism, exhibited significantly genetic correlations with COVID-19 severities (P < 0.05, Figure 4, and Supplementary Table S12), suggesting that the shared genetic risk factors of these comorbidities may aggravate the severities of COVID-19.

**Figure 4.**
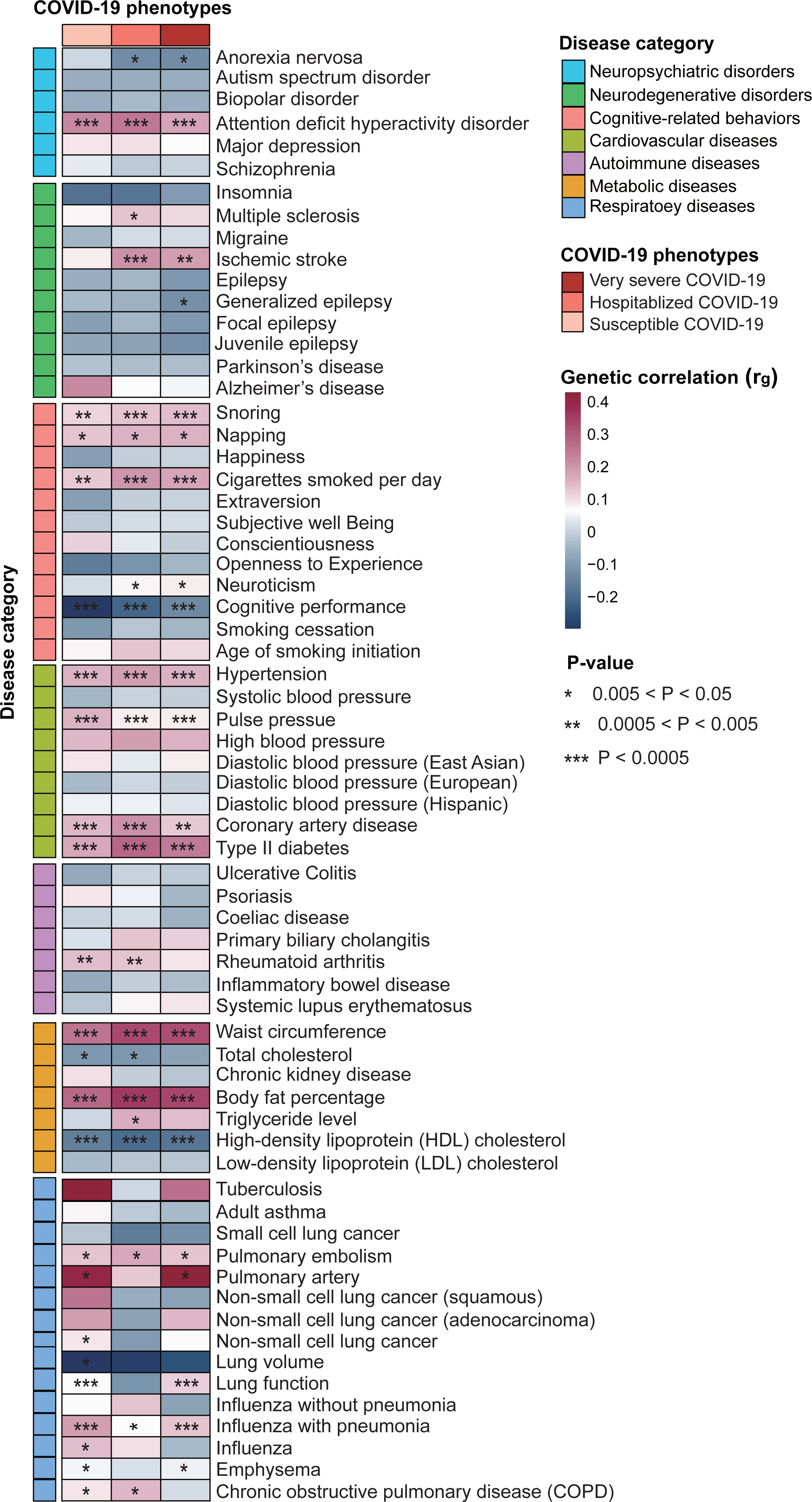
LDSC analysis identifies the genetic correlations between three COVID-19 phenotypes and complex diseases. Heatmap plot showing the results of genetic correlations between 66 diseases or traits from six main disease categories (i.e., Neuropsychiatric disorders, neurodegenerative disorders, cognitive-related behaviors, cardiovascular diseases, autoimmune diseases, metabolic diseases, and respiratory diseases) and three COVID-19 outcomes (i.e., very severe COVID-19, hospitalized COVID-19, and susceptible COVID-19 using the LDSC method. The asterisk represents the significance of genetic correlation between COVID-19 and complex disease. See also Supplementary Table S12.

Given that the primary goal of current study was to characterize the context-specific genetic etiology of COVID-19 severities, we concentrated the subsequent analyses on identifying severe COVID-19-relevant cell subpopulations across three main human organoids (i.e., lung, intestine, and brain), and used these 438 risk genes to reposition drug targets for treating severe COVID-19 and related complications.

### Identifying severe COVID-19-relevant cell subpopulations in lung organoids

Respiratory failure is the leading cause of death in severe COVID-19 patients [52, 53]. It is important to study pathologic cells associated with COVID-19 in human lung organoids for facilitating to explore key features of viral biology and drug repositioning [54]. Thus, we sought to identify severe COVID-19-relevant cell subpopulations by integrating GWAS summary statistics with human lung organoid scRNA-seq data [55] using the scPagwas method. Among three main cell types, we found that the MSCs with higher trait-relevant scores (TRSs) exhibited striking enrichments in very severe COVID-19 (Figure 5A-C and Supplementary Figure S14), reminiscing that the cell type of MSCs was identified to be associated with COVID-19 severities in human fetal lung tissue (Supplementary Figure S6). There was a prominently higher proportion of scPagwas positive cells in MSCs (42.14%) compared to other two cell types (Figure 5D). Because of the binary trait settings of very severe COVID-19 and healthy population, these scPagwas positive cells should be associated with COVID-19 severity, and scPagwas negative cells should be relevant to the normal phenotype. Moreover, we used the recent cell-scoring method, scDRS[56], to re-analyze the same data, and found that these results were remarkably consistent (rho = 0.926, P < 2.2×10^-16^, Supplementary Figure S15A-B).

**Figure 5.**
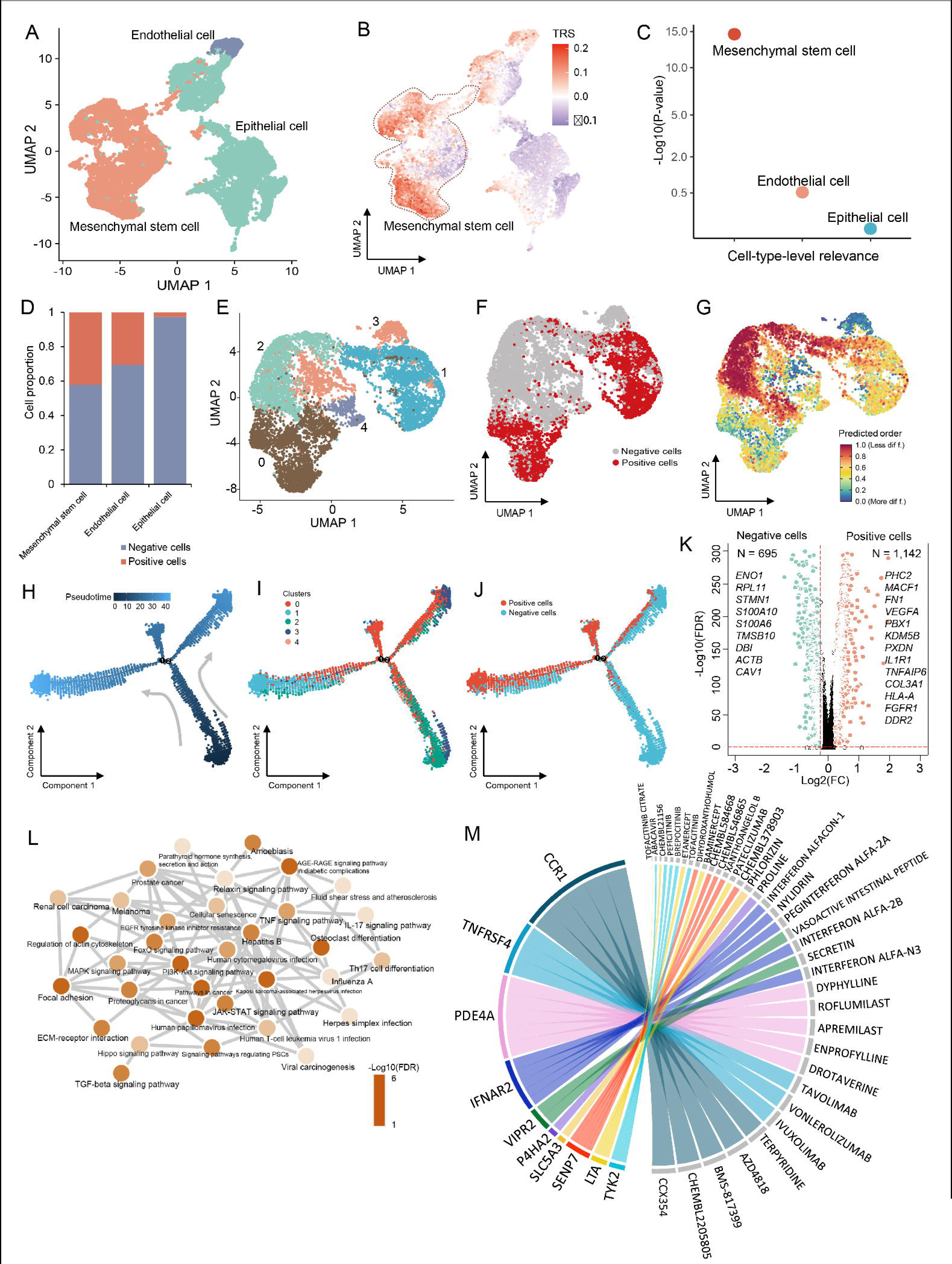
Identification of lung MSCs associated with COVID-19 severities. A. UAMP projections of human lung organoids cells colored by three annotated cell types. B. UMAP embedding of all cells among three cell types in lung organoids colored by the TRSs for the phenotype of very severe COVID-19. C. Dotplot showing the significant associations of three cell types in lung organoids for very severe COVID-19. y axis indicates the log-transformed P value (-Log10(P)), and x axis indicates the cell-type-level inference using the scPagwas method. D. Bar plot showing the proportion of positive cells in three lung organoid cell types. E. UMAP projections of lung MSCs colored by five cell clusters. F. UMAP plot showing the distribution of lung MSC positive cells and negative cells. The C’ value significantly lower than 1 indicates a high level of disease-association heterogeneity across the set of cells (C’ value = 0.924, heterogeneity FDR = 3.332×10^-4^). G. CytoTRACE differentiation continuum across the lung MSCs. The color legend indicates the degree of differentiation that is gradually increased from more differentiation (blue) to less differentiation (red). H. Unsupervised trajectory inference of lung MSCs functional state transitions. Color legend indicates the pseudotimes of individual cells calculated by using the Monocle2 method. I-J) Visualization of the distribution of five cell clusters (I) and MSC positive cells (J) in the inferred trajectory. K. Volcano plot showing significantly up-regulated genes between MSC positive cells and negative cells. A two-side Wilcoxon test was used for assessing the significance. L. Notably enriched pathways by 1,142 up-regulated genes in MSC positive cells. Color legend represents the log-transformed FDR value (-Log10(FDR)). M. Chord diagram of scDrugHunter-identified top 10 druggable genes and relevant interacting drugs for very severe COVID-19 in lung MSCs. The width of each line is determined by the number of drugs (n =1∼5) known to interact with each gene. Genes are ordered by the degree of scDGS at the top of the diagram. See also Supplementary Tables S13-S14.

As shown in Figure 5E, MSCs were clustered into five cell clusters. Among the 9,795 MSCs cells, scPagwas identified 4,128 positive cells that are most relevant to severe COVID-19 (Bonferroni-corrected P < 0.05, Figure 5F). These severe COVID-19-relevant positive cells with higher TRSs were overrepresented in clusters 0 and 1, whereas cluster 2 exhibited the lowest TRSs (heterogeneous FDR = 3.332×10^-4^, *C’* value = 0.924; Figure 5F and Supplementary Figure S16A-B), which is consistent with the results from the scDRS analysis (concordance rate = 88.25%, Supplementary Figure S15C-E). Furthermore, the per-cell genetic risk scores using the 438 COVID-19-relevant genes showed a notable correlation with scPagwas TRSs across all MSCs (P < 2.2×10^-16^, Supplementary Figure S16C-D). On CytoTRACE analysis [57] for predicting differentiation states from MSCs, we found that cells in clusters 0 and 1 were predicted to be more differentiated than that in cluster 2 (Figure 5G and Supplementary Figure S16E). By performing an unsupervised trajectory inference analysis [58], MSC positive cells in clusters 0 and 1 were largely distributed in the middle and end positions of the trajectory (Figure 5H-J). The pseudotime of MSCs were positively correlated with corresponding TRSs (rho = 0.664, P < 2.2×10^-16^, Supplementary Figure S16F-G). Notably, these top branch-dependent genes related to MSC positive cells exhibited notable enrichments in several critical biological processes, which are relevant to lung and respiratory proliferation and growth (Supplementary Figure S16H).

Recent evidence [59] suggested that increased numbers of MSCs and fibroblasts concomitant with increased proximity between these two cell types during the COVID-19 progresses, which probably reflects a response to repair the damaged lung tissue. Thus, we further sought to examine whether MSC positive cells have higher proximity with fibroblasts than negative cells. As expected, we found that the fibroblast-relevant cell state scores by collapsing the expression levels of fibroblast marker genes were significantly higher among MSC positive cells compared to negative cells (P < 2.2×10^-16^, Supplementary Figure S17A-C). These results indicate that MSC positive cells tend to have differentiation potentials for facilitating to repair COVID-19-induced lung-tissue injury. Compared with negative cells, there were 1,142 significantly up-expressed genes in MSC positive cells, such as *FN1*, *VEGFA, IL1R1, TNFAIP6*, and *PHC2* (Figure 5K and Supplementary Figure S16I). The gene of *FN1*, known to be a driver of pulmonary fibrosis, was reported to be upregulated in COVID-19 survivors [60]. Functionally, these up-regulated genes were significantly overrepresented in 40 biological pathways (FDR < 0.05, Figure 5L and Supplementary Table S13), including human papillomavirus infection, PI3K-AKT signaling pathway, and JAK-STAT signaling pathway, recalling that many of them have been strikingly enriched in aforementioned genetics-based pathway analyses (Figure 3F).

To prioritize critical gene-drug pairs, we applied the scDrugHunter method [61] to reposition MSC-specific druggable genes and interacting drugs for treating severe COVID-19. Among 438 genetic risk genes (Supplementary Table S8), we found that 98 genes (22.4%) were targeted at least one known drug, and 15.3% of these 98 genes were documented to be targets for potential COVID-19-relevant drugs based on registers of clinical trials for COVID-19 [62], which is notably higher than that from random selections based on *in silico* permutation analysis (permuted P < 0.001, Supplementary Figure S18A-B). Of note, there were 19 druggable genes with 117 targeting drugs yielding remarkably higher single-cell druggable gene scores (scDGSs > 120 and FDR < 0.05) in lung MSCs for treating severe COVID-19, including *CCR1*, *TNFRSF4*, *PDE4A*, and *IFNAR2* (Figure 5M, Supplementary Figure S19A-B and Table S14). Notably, we found that 12 of these interacting drugs, including IBUDILAST, ILOPROST, INTERFERON ALFA-2B, and INTERFERON BETA-1B, were tested in 60 double-blind and placebo-controlled clinical trials for the treatment of COVID-19 (<u>Clinicaltrials.gov</u>, Supplementary Figure S20A). Consistently, we performed evidence-driven analysis using the RISmed method[39], and found that a high proportion of these prioritized drugs have been associated with COVID-19 (proportion = 42.74%, Supplementary Figure S20B). Collectively, these results demonstrate that cell subsets of MSCs are highly relevant to severe COVID-19, and these highlighted druggable genes potentially have therapeutic functions in MSCs for severe COVID-19.

### Discerning severe COVID-19-relevant cell subpopulations in intestine organoids

Although COVID-19 primarily manifests pulmonary infection, it has significant extrapulmonary complications to damage other organ systems, including the intestinal tract [63]. Due to the extensive surface area of intestinal capillaries, intestinal epithelial cells are more likely to be infected by SARS-CoV-2 than other extrapulmonary organs [64]. To understand the mechanism underlying severe COVID-19-associated intestinal injury, we performed an integrative analysis by incorporating the GWAS summary dataset and human intestinal organoids scRNA-seq data [65]. Among the five cell types, we found that severe COVID-19-relevant cells with higher TRSs were mainly from tuft cells (n = 2,167 cells, Figure 6A-C and Supplementary Figures S21-S22). At cell-type level inference, two cell types of tuft cells and membranous cells (M cells) demonstrated a significant association with severe COVID-19 (Figure 6D), which is consistent with the results based on human fetal intestine tissue (Supplementary Figure S6). This observation remained reproducible by using the scDRS method [56] with the inclusion of the same single-cell dataset (rho = 0.981, P < 2.2×10^-16^, Supplementary Figure S23A-B). While tuft cells are chemosensory epithelial cells, they serve as the primary physiologic target of viral infection and drive an inflammatory adaptive immune response, which is classically correlated with allergy and parasitic infection [66, 67].

**Figure 6.**
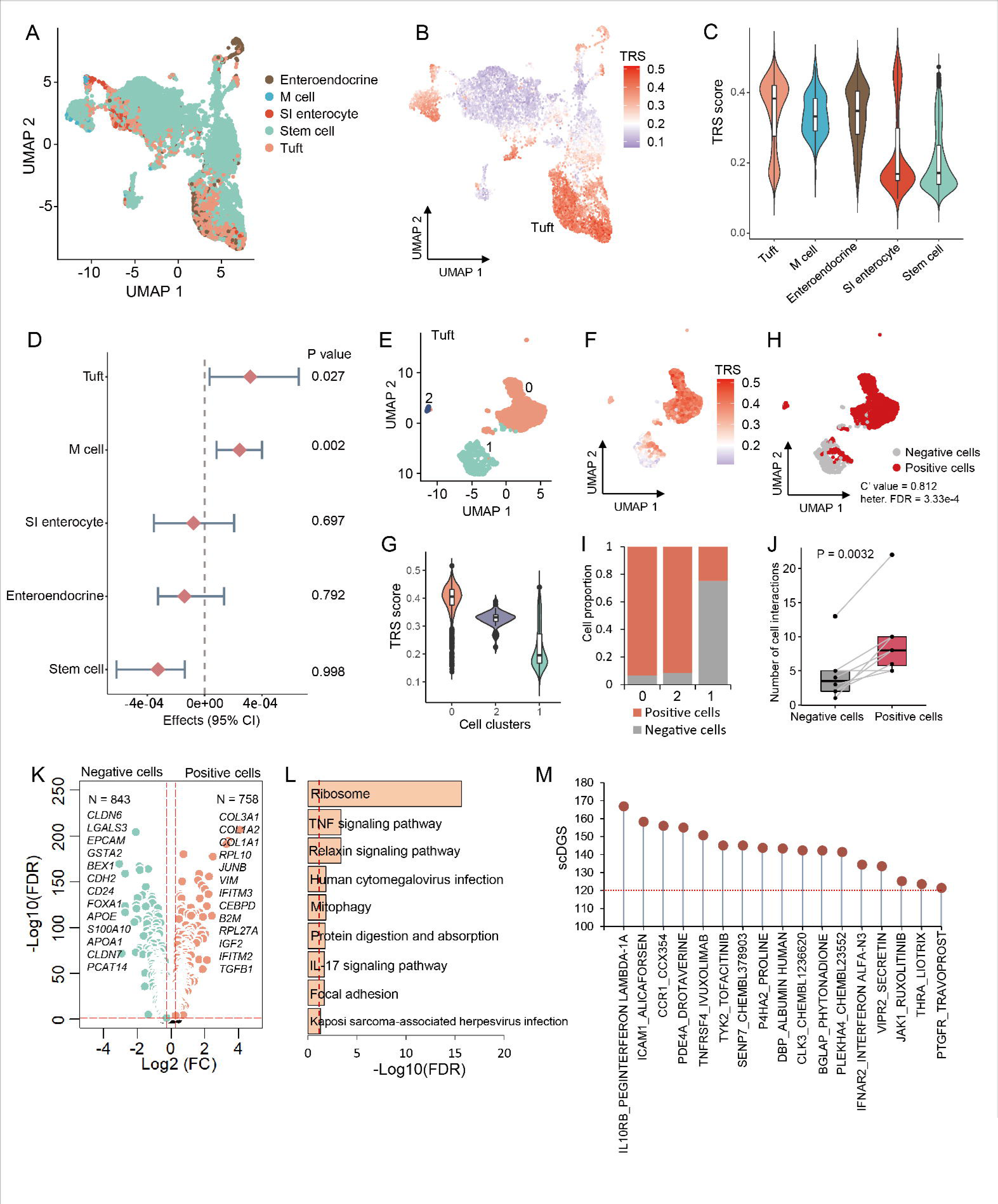
Discerning intestinal tuft cells relevant to COVID-19 severities. A. UAMP projections of human intestine organoids cells colored by five predefined cell types. B. UMAP embedding of all cells among five cell types in intestine organoids colored by the TRSs for the phenotype of very severe COVID-19. C. Violin plot showing the TRSs in five cell types among intestine organoids. D. Forest plot showing the associations of intestinal cell types with very severe COVID-19. Effect parameter indicates the strength of association, and range specifies the empirical bounds of the 95% confidence interval. The P value of each cell type is shown in the right panel. E. UMAP showing three cell clusters of intestinal tuft cells. F. UMAP visualization of intestinal tuft cells colored by TRSs. G. Violin plot showing the TRSs in three cell clusters among intestinal tuft cells. H. UMAP visualization of intestinal tuft cells colored by tuft positive and negative cells. The C’ value significantly lower than 1 indicates a high level of disease-association heterogeneity across the set of cells (C’ value = 0.812, heterogeneity FDR = 3.33×10^-4^). I. Bar plot showing the proportion of positive cells in three cell clusters of intestinal tuft cells. J. Boxplot showing a notable increase in cellular interactions of tuft positive cells with other cells among intestinal organoids compared to tuft negative cells. K. Volcano plot showing significantly up-regulated genes between tuft positive cells and negative cells. A two-side Wilcoxon test was used for assessing the significance. L. Notably enriched pathways by 758 up-regulated genes in tuft positive cells. x axis indicates the log-transformed FDR value (-Log10(FDR)). M. Dotplot showing the results of scDrugHunter-identified 17 druggable genes and interacting drugs with high scDGS > 120 in intestinal tuft cells. See also Supplementary Tables S15-S16.

As shown in Figure 6E, tuft cells were grouped into three cell clusters. Among them, we found that severe COVID-19-associated genetic signals were highly enriched in cluster 0 (heterogeneous FDR = 3.33×10^-4^, *C’* value = 0.812, Figure 6F-G). Consistently, clusters 0 and 2 had a higher proportion of positive cells relevant to COVID-19 severities than that in cluster 1 (Figure 6H-I), which is in concordance with tuft positive cells identified using the scDRS method (concordance rate = 0.984, Supplementary Figure S23C-E). Moreover, this result was also validated by using the per-cell genetic risk scores of 438 COVID-19-relevant genes (P =2.86×10^-8^, Supplementary Figure S24A-B). Cellular communication analysis indicated that tuft positive cells had a significantly higher number of receptor-ligand interactions with other intestinal cell types than that of tuft negative cells (P = 0.0032, Figure 6J, Supplementary Figure S24C-E). For example, tuft positive cells showed relatively high communications with M cells, containing 32 significant receptor-ligand interactions; for example, several unique interacted pairs of WNT5A-FZD5, SEMA3A-(NRP1+PLXNA3), and PTN-SDC3 (Supplementary Figure S24F-G).

By performing a differential expression analysis, we found that 758 genes showed significantly higher expressions in tuft positive cells compared to negative cells, including *COL3A1*, *COL1A2*, *IFITM3*, *RPL10*, *VIM*, and *LGALS1* (Figure 6K). These extracellular matrix genes, including *COL3A1* and *COL1A2*, were reported to be up-regulated in COVID-19 microvessels and lung lower lobes [52, 68, 69]. Genetic variants in the interferon-induced transmembrane protein (*IFITM3*) have been demonstrated to be associated with SARS-CoV-2 infection and COVID-19 severities [70–72]. Functionally, these highly expressed genes showed notable enrichments in several critical pathways, including ribosome, TNF signaling pathway, and relaxin signaling pathway (Figure 6L and Supplementary Table S15), of which several have been reported to be implicated in COVID-19 infection [24, 73]. For example, previous evidence has suggested that ribosomal proteins potentially play crucial roles in blocking viral replication by binding to the specific phosphoproteins for the host immune factors [74], and the immunosuppression and low expression of ribosomal protein genes were related to the persistence of the viral infection in COVID-19 patients [75].

Moreover, we also repurposed tuft-specific druggable genes and interacting drugs for treating severe COVID-19 and intestinal comorbidities. Among 438 genetic risk genes, we found that 17 druggable genes with 151 interacting drugs yielded higher scDGSs (> 120, and FDR < 0.05) in tuft cells for treating severe COVID-19, including *IL10RB*, *ICAM1*, *TYK2, SENP7*, and *VIPR2* (Figure 6M and Supplementary Figures S25-S26 and Table S16). Among these identified gene-drug pairs, 14 drugs, including PEGINTERFERON LAMBDA-1A, TOFACITINIB, TADALAFIL, and PENTOXIFYLLINE, have been examined in 89 clinical trials for treating COVID-19 patients (<u>Clinicaltrials.gov</u>, Supplementary Figure S27A). Furthermore, the RISmed analysis consistently demonstrated that a large number of these identified drugs (n = 64) were relevant to the treatment of COVID-19 (proportion = 42.38%, Supplementary Figure S27B). Together, our results indicate that subset of tuft cells exhibit notable associations with severe COVID-19, and critical drug targets, including *IL10RB*, *ICAM1*, and *VIPR2*, are prioritized for treating severe COVID-19 and concomitant intestinal symptoms.

### Distinguishing severe COVID-19-relevant cell subpopulations in brain organoids

Accompanied with respiratory and gastrointestinal symptoms, severe COVID-19 patients often present with short- and long-term neuropsychiatric symptoms and brain sequelae [36, 76]. Brain organoids provide a promising tool for uncovering the pathophysiologic mechanisms and potential therapeutic options for neuropsychiatric complications of severe COVID-19 [77]. We leveraged the scPagwas method [27] to integrate the GWAS summary dataset on very severe COVID-19 and human cerebral organoids scRNA-seq data [78]. Among eight main cell types, we identified that both endothelial cells (P = 6.96×10^-6^) and microglia (P = 5.29×10^-5^) yielding higher TRSs were significantly associated with very severe COVID-19 compared to other cell populations (Figure 7A-C), recalling that these two cell types were identified to be associated with COVID-19 severities in human fetal brain tissue (Supplementary Figure S6). Consistently, these results were notably reproduced by using the scDRS method [56] in the same dataset (rho = 0.98, P < 2.2×10^-16^, Supplementary Figure S28A-B). Earlier studies [36, 79] have indicated that SARS-CoV-2 invade into central nervous system via endothelial cells resulting in inflammation, thrombi, and brain damage.

**Figure 7.**
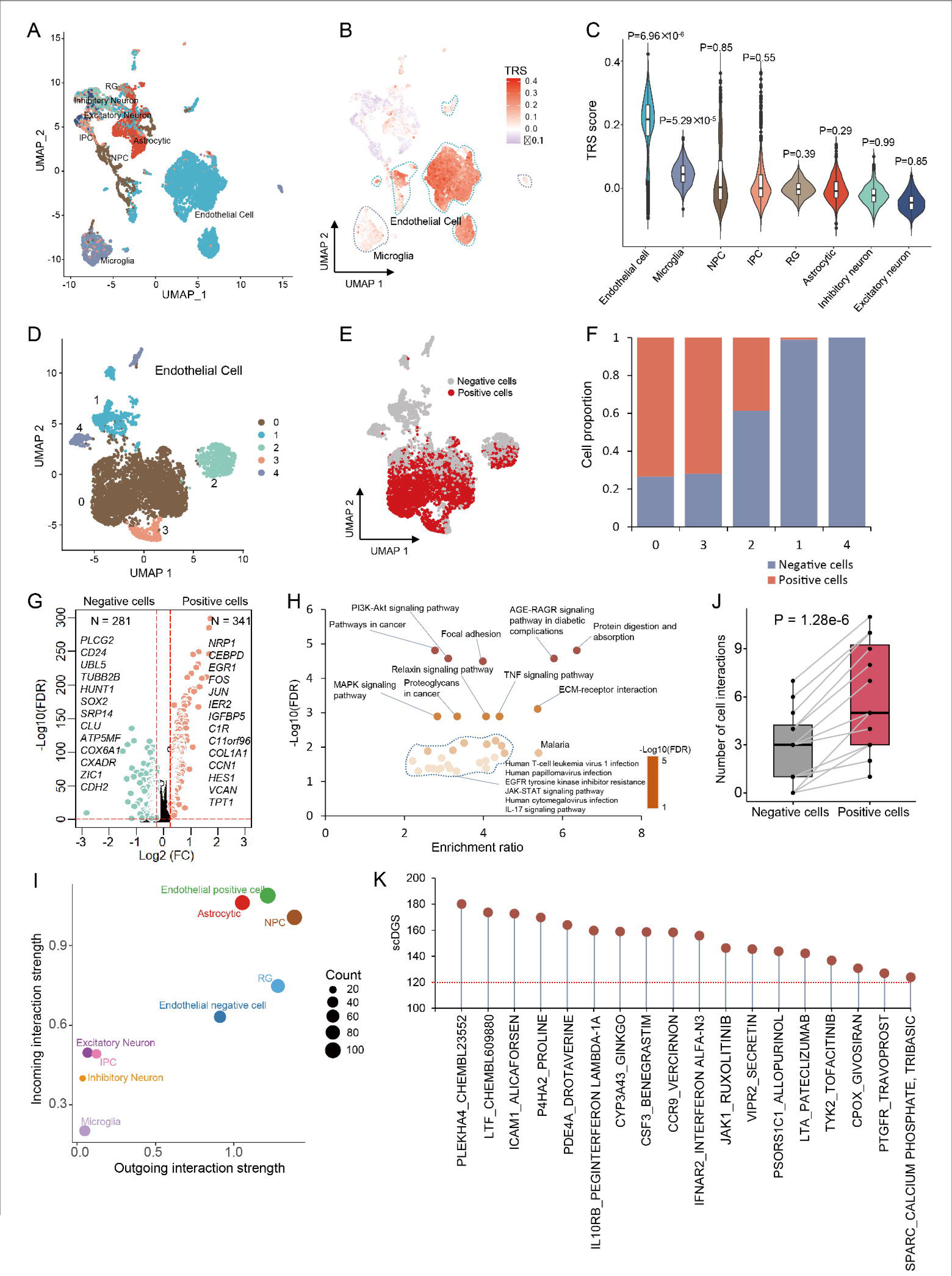
Distinguishing brain endothelial cells contribute risk to COVID-19 severities. A. UAMP projections of all cells colored by eight predefined cell types in human brain organoids. B. UMAP embedding of all cells in brain organoids colored by the TRSs for the phenotype of very severe COVID-19. C. Violin plot showing the TRSs in eight cell types among intestine organoids. The significant level (P values) of associations of brain cell types with very severe COVID-19 is shown in top-panel of the violin plot. D. UMAP visualization of five cell clusters in brain endothelial cells. E. UMAP plot highlighting the brain endothelial positive cells. The C’ value significantly lower than 1 indicates a high level of disease-association heterogeneity across the set of cells (C’ value = 0.841, heterogeneity FDR = 3.33×10^-4^). F. Bar plot showing the proportion of positive cells in five cell clusters of brain endothelial cells. G. Volcano plot showing significantly up-regulated genes between endothelial positive cells and negative cells. A two-side Wilcoxon test was used. H. Notably enriched pathways by 341 up-regulated genes in endothelial positive cells. y axis indicates the log-transformed FDR value (-Log10(FDR)), and x axis indicates the enrichment ratio of each pathway. I. Scatter plot exhibiting the dominant senders (sources) and receivers (targets) in a 2D space. y axis represents incoming interaction strength, and x axis represents outgoing interaction strength. The size of each node indicates the count of cellular interactions. J. A notable increase in cellular interactions of endothelial positive cells with other cells among brain organoids compared to endothelial negative cells. K. Dotplot exhibiting the results of scDrugHunter-identified 18 druggable genes and interacting drugs with high scDGS > 120 in brain endothelial cells. See also Supplementary Tables S17-S19.

Among endothelial cells with five clusters, we identified 3,443 positive cells that were significantly associated with very severe COVID-19 (Bonferroni-corrected P < 0.05, proportion = 56.6%, Figure 7D-E and Supplementary Figure S29). Remarkably heterogeneous associations between brain endothelial cells and severe COVID-19 were uncovered (heterogeneous FDR = 3.33×10^-4^, *C’* value = 0.841, Figure 7E). Of note, clusters 0 and 3 exhibited a higher proportion of positive cells than other clusters (Figure 7F and Supplementary Figure S30A-B), which is in accordance with the results from the scDRS analysis (concordance rate = 0.74, Supplementary Figure S28C-E). Compared with endothelial negative cells, we found that 341 genes, including *NRP1*, *CEBPD*, and *EGR1*, were significantly up-regulated in positive cells (Figure 7G). The cell surface receptor of neuropilin-1 (NRP1) was reported to serve as an entry factor and potentiate SARS-CoV-2 infectivity, and it up-regulated expression is critical in angiogenesis, viral entry, immune function, and axonal guidance [80, 81]. Functionally, these highly up-regulated genes were enriched in multiple critical pathways and biological processes, including PI3K-AKT signaling pathway, focal adhesion, TNF signaling pathway, ECM-receptor interaction, and angiogenesis (Figure 7H, and Supplementary Figure S30C and Tables S17-S18).

To gain refined insights into endothelial positive cells, we conducted a cell-to-cell interaction analysis among cell populations in human brain organoids. Through constructing the aggregated cellular interaction network based on the count of receptor-ligand pairs, endothelial positive cells exhibited the highest incoming interaction strength than other cell types (Figure 7I). Compared to endothelial negative cells, we found a significant increase in cell-to-cell interactions with other brain cell types (P = 1.28×10^-6^, Figure 7J and Supplementary Figure S31A-B). By summarizing the communication probability among cellular interactions, there were 25 significant ligand-receptor interactions of endothelial positive cells, including *CXCL12-CXCR4*, *FGF7-FGFR1/2*, *PTN-NCL*, and *MDK-NCL* (Supplementary Figure S31C-D). For communicating with microglia, three unique ligand-receptor pairs of *MIF-ACKR3*, *NAMPT-INSR*, and *NAMPT-(ITGA5+ITGB1)* were detected in endothelial positive cells compared to negative cells. SARS-CoV-2 infection enable to damage endothelial cells leading to inflammation that further induce the activation of microglia, which may result in region- and neurotransmitter-specific neuropsychiatric symptoms [36, 82, 83]. Collectively, our results indicate that both endothelial cells and microglia have considerable potential to contribute risk to severe COVID-19.

Subsequently, the scDrugHunter method was used to discern brain endothelial cell-specific druggable genes and interacting drugs for treating severe COVID-19 and corresponding neuropsychiatric complications. Among these putative COVID-19-risk genes, we uncovered that 18 druggable genes with 154 interacting drugs obtained notably higher scDGSs (> 120, FDR < 0.05) in brain endothelial cells, including top-ranked genes of *PLRKHA4*, *LTF*, *ICMA1*, and *P4HA2* (Figure 7K, Supplementary Figures S32-S33 and Table S19). Of note, 16 of these prioritized drugs have been demonstrated to be tested in 96 clinical trials for the treatment of COVID-19 (<u>Clinicaltrials.gov</u>, Supplementary Figure S34A). Consistently, the RISmed analysis indicated that 74 drugs have been associated with the treatment of COVID-19 (48.05%, Supplementary Figure S34B).

By performing a comparison analysis, we further found that seven druggable genes of *IFNAR2*, *TYK2*, *VIPR2*, *PLEKHA4*, *PDE4A*, *P4HA2*, and *PTGFR* were identified to be common targets across three COVID-19-relevant cell types of lung MSCs, intestinal tufts, and brain endothelial cells (Supplementary Figure S35A). Eight druggable genes of *COL11A2*, *SACM1L*, *HCN3*, *CA11*, *SLC22A4*, *CLK2*, *IMPG3*, and *SLC5A3* were specific to lung MSCs, four druggable genes of *DBP*, *CLK3*, *BGLAP*, and *THRA* were specific to intestinal tufts, and seven brain endothelial cell-specific druggable genes of *CSF3*, *LTF*, *PSORS1C1*, *SPARC*, *CCR9*, *CPOX*, and *CYP3A43*. Collectively, we repurposed 33 putative druggable genes and 215 interacting drugs for the treatment of severe COVID-19 and corresponding complications, and these 33 druggable genes were jointly enriched in a functional subnetwork (Supplementary Figure S35B).

## Discussion

Multiple lines of evidence [23, 24, 27, 39, 56] have demonstrated that integrating scRNA-seq data and polygenic risk signals from GWAS is a promising approach to uncover the cellular mechanisms through which these variants drive complex diseases. In the current study, we sought to identify critical cell types/subpopulations relevant to COVID-19 severities by combining large-scale GWAS summary statistics and human organoids single-cell sequencing data. Crucially, 39 main cell types in eight kinds of organoids were identified to be associated with COVID-19 severities. We further concentrated on unveiling the functions of COVD-19-relevant cell subpopulations across three main organoids of lung, intestine, and brain, which contribute to characterize important features of viral biology and facilitate to the identification of repurposable drug candidates against SARS-CoV-2 infection and its related comorbidities.

Although vaccines have been developed to prevent SARS-CoV-2 infection, no specific antiviral drug exist to mitigate the established disease of severe COVID-19 [5]. As developing a new drug takes years to a decade and substantial cost, drug repurposing is an effective way that can notably accelerate the development cycle of therapeutic strategies for treating COVID-19 [4]. There are two main approaches, virus-based and host-based treatment options, to test candidate targets in clinical trials. Of them, the host-based approaches target critical host factors that are used by SARS-CoV-2 for viral replication or stimulate host innate antiviral responses [84]. The key to host-based drug repurposing for the treatment of COVID-19 infection is to distinguish the true host risk genes. GWAS-identified disease risk genes were more prone to code for proteins that are “biopharmable” or “druggable” than the rest of the human genome [85]. In the present investigation, we leveraged integrative genomic analyses to analyze large-scale GWAS data and prioritized 438 COVID-19-relevant risk genes, including *IFNAR2*, *CCR1*, *ICAM1*, *VIPR2*, and *IL10RB*, which are attributable to search genuine drug targets for COVID-19 severities.

Despite the success of GWASs, nearly 90% of disease-associated variants are identified to be located in the non-coding regions, which are enriched in cell-type-specific transcriptional regulatory elements relevant to disease risk [86–88]. Integration of GWAS summary data and eQTL data has been extensively used to discern novel candidate genes and yield functional insights into disease-relevant pharmacological effects [4, 20, 21], however, few of these insights has considered the cell-type-specific effects of drug targets. Thus, in the present study, we repositioned drugs and their interacting targets for treating severe COVID-19 in a cell-type-specific context. Collectively, we found that 33 druggable genes and 215 interacting drugs were considered as putative candidates for severe COVID-19 and relevant complications. Large proportions of these drugs have been experimented for the treatment of severe COVID-19. For example, the FDA-approved drugs of INTERFERON ALFA-2B and INTERFERON BETA-1B exhibited agonist-receptor interactions with *IFNAR2*, which could be used alone or in conjunction with other anti-virus drugs for against COVID-19 initiation and progression [89, 90].

Several limitations of this study should be cautious. First, the power of the cell-type-level integration analysis is limited by the lack of scRNA-seq data and matched genetic information in each sample for discerning COVID-19-relevant cell types. To diminish the impact of this limitation, we adopted a powerful approach by incorporating a large-scale GWAS summary dataset and human organoids scRNA-seq data with a large amount of cells, as reference to previous studies [8, 23, 24, 27, 56]. Second, the identification of COVID-19-relevant cell types or subpopulations does not imply causality but may reflect indirect discovery of causal phenotype-cell associations, analogous to earlier studies [24, 56]. Third, we removed the MHC region from all genomic analyses to reduce the influence of the complex genetic architecture and extensively high levels of LD, parallel to previous studies [8, 24]. However, it should be noted that COVID-19-relevent genetic signals in this locus might be ignored. Finally, we adopted a default strategy that linking SNPs into genes based on the proximal distance of a 20kb window. Other powerful strategies, including the enhancer-gene linking approaches from Roadmap and Activity-By-Contact models [23, 91], can also be used to establish the link between SNPs and genes.

## Conclusions

In summary, we provide systematic insights that the effects of host genetic factors on COVID-19 initiation and progression in a cellular context, and first repurpose COVID-19-relevant cell-type-specific druggable targets and interacting drugs. Numerous critical cell types or subpopulations, including lung MSCs, intestinal tuft cells, and brain endothelial cells, contribute higher risk to COVID-19 severities. The integration of human genetics, single-cell transcriptomic data, and large-scale compound resources should improve *in silico* pharmacology for drug repurposing, which will provide novel insights in therapy discovery and development for the infection pandemic.

## Methods

### Human organoids scRNA-seq datasets

In this study, we collected and curated 93 independent scRNA-seq datasets of 10 kinds of widely-adopted human organoids (i.e., brain, lung, intestine, heart, eye, liver & bile duct, pancreas, kidney, and skin) spanning 1,159,206 cells with 62 main cell types from two widely-used databases of Gene Expression Omnibus (GEO) [92] and ArrayExpress [93]. Only datasets with publically available raw reads (e.g., SAR, bam file, or fastq) were included. We leveraged a unified pipeline to conduct re-alignment, quality control, and standard analysis for facilitating the data integration and minimize the batch effects (Supplementary Figure S1). Human cancer-derived organoid scRNA-seq datasets were excluded from our current analyses. A common dictionary of gene symbols was used to annotate genes for allowing comparison analysis across samples and datasets, and these unrecognized symbols were removed.

### Human fetal scRNA-seq datasets

To validate the reliability of human organoids-based significant results, we also collected nine independent scRNA-seq datasets containing eight kinds of *de facto* human fetal organs (i.e., brain, lung, intestine, liver, kidney, eye, pancreas, and skin) across 48 samples from the Gene Expression Omnibus (GEO) [92] and ArrayExpress [93] databases. Analogue to organoids scRNA-seq data, we only included datasets with publically available raw reads (e.g., .SAR, .bam file, or .fastq) and used the unified pipeline to carry out re-alignment, quality control, and standard analysis (Supplementary Figure S1). In total, there were 223,334 cells across all human fetal organs, ranging from 1,745 to 63,020 cells in each dataset.

### Single-cell RNA sequencing data processing

We initially applied two widely-used tools of SRA-toolkit (version 3.0.5) [94] and bamtofastq (version 2.31.0)[95] to convert single-cell transcriptomic profiles in .SRA and .bam format to .fastq format. The CellRanger (version 6.1.2) [96] and STARsolo (version 2.7.10a) [97] were used for separately processing human organoid or fetal scRNA-seq data from 10× Genomics sequencing platform and Drop-seq sequencing platform to debarcode cells and generate a matrix of unique molecular identifiers (UMIs) for each sample. For both sequencing platforms, we used the human reference genome assembly hg38 [98] to align reads tagged with a cell barcode and UMI. Subsequently, featureCounts (version 1.22.2) [99] was used for assigning tagged reads to corresponding genes, and SCANPY (version 1.9.1) [100] was utilized for filtering out cells with < 500 or >20,000 detectable genes, >30,000 expressed gene counts, and >10% mitochondrial rate.

Moreover, we used the *FindVariableFeatures()* function in Seurat (version 4.3.0) [101] to select top 2,000 high variable genes (HVGs), and employed the *NormalizeData()* and *ScaleData()* in Seurat to transform and scale human organoid and fetal scRNA-seq data. The Harmony (version 3.8) [102] tool was adopted to integrate samples and remove batch effects, and the Principal component analysis (PCA) was applied to obtain top 30 the most different principal components (PCs), which could explain the most variance of top 2,000 HVGs in the aforementioned step of finding variable features. High quality cells were embedded into two dimensions by using the uniform manifold approximation and projection (UMAP), and annotated to specific cell types using the transfer learning method of scArches (version 0.5.1) [103] with manually validation.

### GWAS summary data on COVID-19-related phenotypes

The COVID-19 meta-analytic GWAS summary statistics were downloaded from the official website of COVID-19 Host Genetics Initiative [104] (https://www.covid19hg.org/; COVID19-hg GWAS meta-analyses round 7, released date of April 8, 2022). For the current investigation, we used three of these GWAS meta-analyses, which included 81 independent studies containing mixed population ancestries (Supplementary Figures S2-S3 and Table S2). Most cohorts were based on European ancestry. Three examined COVID-19-related phenotypes includes: 1) very severe respiratory confirmed COVID-19 (Very severe, file named A2_ALL_leave_23andme, n = 18,152 cases) vs population (n = 1,145,546 controls), 2) hospitalized COVID-19 (Hospitalization, file named B2_ALL_leave_23andme, n = 44,986 cases) vs population (n = 2,356,386 controls), and 3) susceptibility to COVID-19 (Susceptible, file named C2_ALL_leave_23andme, n = 159,840 cases) vs population (n = 2,782,977).

As referenced to a previous study [17], very severe COVID-19 patients were defined as hospitalized COVID-19 patients as the primary reason for hospital admission with laboratory-confirmed SARS-CoV-2 infection and death or respiratory support. Simple supplementary oxygen (e.g., 2L min^-1^ through nasal cannula) did not meet the definition of very severe status. Hospitalized COVID-19 patients were defined as individuals hospitalized with laboratory-confirmed SARS-CoV-2 infection, where the hospitalization of patients because of COVID-19-relevant symptoms. Susceptibility to COVID-19 patients were defined as individuals with self-reported infection, health-record infections, or laboratory-confirmed SARS-CoV-2 infection. In comparison, controls were defined as those individuals in the participating studies who did not qualify the definition of cases. The meta-GWAS summary datasets contained p-value for each single nucleotide polymorphism (SNP), effect size on log(OR) scale, standard error of effect size, minor allele frequency (MAF), and p-value from Cochran’s Q heterogeneity test. After stringent quality control, a total of 11,732,503, 12,030,868, and 14,335,927 genetic variants with MAF over 0.0001 and the imputation score (R^2^) of greater than 0.6 were satisfied in the A2, B2, and C2 meta-GWAS datasets, respectively. Results from 23&Me cohort GWAS summary statistics were removed from the current investigation. The *qqman* [105] R package was applied to visualize Manhattan plot and quantile-quantile (QQ) plot.

### Integration of GWAS summary statistics and scRNA-seq data

To distinguish critical cell types/subpopulations by which genetic variants influence COVID-19 severities, we implemented our own developed pathway-based polygenic regression method, scPagwas (version 1.1.0) [27], to integrate GWAS summary data on three COVID-19 outcomes with human organoids and fetal scRNA-seq datasets. Initially, scPagwas annotates SNPs to their proximal genes (a default window size of 20kb) of the corresponding pathways, which are based on the experimentally validated canonical pathways in the Kyoto Encyclopedia of Genes and Genomes (KEGG) database [106]. Then, scPagwas leverages the singular value decomposition algorithm to transform a scaled scRNA-seq matrix into a pathway activity score (PAS) matrix. The projection of the features of genes in a given pathway on the direction of the first principal component (PC1) eigenvalue to define PAS *s_i,j_* for the pathway *i* in cell *j*.

scPagwas assumes *a priori* that SNPs’ effect sizes ***b****_s_i__* in the pathway *i* follow the multi-variable normal distribution 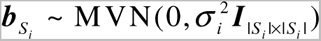, where *σ*^2^ is the variance of effect-sizes for SNPs in the pathway, and ***I*** is the | *S_i_* |× | *S_i_* | identity matrix. The notation *S_i_* ={*k* : *g*(*k*) ∈ *P_i_* } is used to indicate the set of SNPs within pathway *i*, and the notation *P_i_* indicates the set of genes in the pathway *i* . The variance *σ*^2^ is estimated by using the linear weighted sum method:

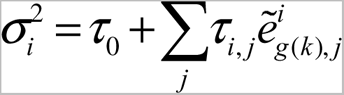

where *τ_0_* indicates an intercept term, *τ _i,j_* indicates the coefficient for the pathway *i* in cell *j*, and 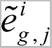 is the expression level for each gene *g* adjusted by the pathway activity *s_i, j_* in the given pathway *i* . scPagwas estimates 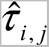 by the following equation:

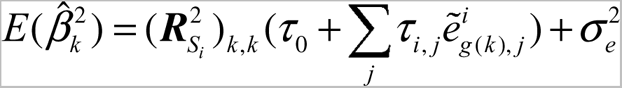

Where 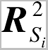 represents the *k*th diagonal element of matrix and denotes the linkage disequilibrium (LD) matrix. The 1,000 Genomes Project Phase 3 Panel [98] is used to compute the LD among SNPs extracted from COVID-19-related GWAS summary statistics.

The genetically-associated PAS (gPAS) for each pathway in a given cell is calculated by summing the product between estimated coefficient 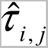 and weighted pathway activity. Then, trait-relevant genes are prioritized by ranking the Pearson correlation coefficients between the expression of each gene and the sum of gPASs over all pathways in each cell across cells. The trait-relevant score (TRS) for each individual cell is calculated using top 1,000 trait-relevant genes based on the *AddModuleScore()* function in Seurat [101]. scPagwas assesses the statistical significance of each cell by using the percent ranks of these trait-relevant genes across individual cells. In addition, scPagwas is also used to infer COVID-19-relevant predefined cell types based on the block bootstrap method [107]. We only include the SNPs on autosomes with MAF > 0.01. The major histocompatibility complex region (chr6: 25-35 Mbp) is removed because of the extensive LD in this genomic region. For more detailed information, please refers to the original paper [27].

### Assessment the heterogeneity of a given cell type relevant to COVID-19

Following a previous study [108], we adopt the Geary’s C method [109] measure the spatial autocorrelation of TRS across cells within a given cell type/subpopulation with regard to a cell-cell similarity matrix. The autocorrelation statistic C, is calculated as the following equation:

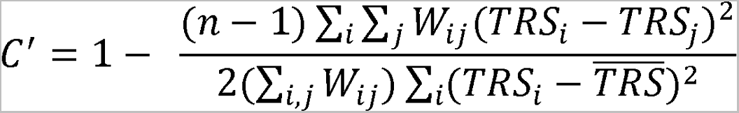

where *n* indicates the total number of cells within a given cell type/subpopulation, *TRS* indicates the TRS of each cell, 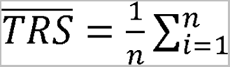 *TRS*_i_ and *W* represents the weight between cells. First, the nearest k neighbors (e.g., 5) should be determined for each cell in the latent model. Subsequently, a Gaussian kernel to the distances between nearest neighbors is used to compute the weights. Higher weights are assigned to similar cells, and zeroed weights are assigned to distant cells. In this way, the Geary’s C method provides a measure of how similar the TRS ranks for neighboring cells given a latent mapping. The C, value is defined as the autocorrelation effect size that a 1 indicates maximal autocorrelation and a 0 intuitively indicates no autocorrelation. The C, value notably close to 1 indicates strongly spatial autocorrelation, reflecting that there is a remarkable trait-association heterogeneity across the given cell type or cell cluster. The VISION R package [108] is used to evaluate the heterogeneity of cells within three COVID-19-relevant cell types of lung MSCs, intestinal tuft cells, and brain endothelial cells using default parameters.

### Transcriptome-wide association analysis

To prioritize genetically-regulatory expression of genes relevant to COVID-19, we perform an integrative genomics analysis of incorporating GWAS summary statistics on three COVID-19-related phenotypes (released round 7) with expression quantitative trait loci (eQTL) data for 49 tissues from the GTEx Project (version 8) by using the S-PrediXcan [110] method. S-PrediXcan primarily leverages two linear regression models to analyze the association between predicted gene expression and COVID-19-related phenotypes:

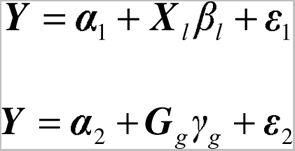

Where *α*_1_ and *α*_2_ are intercepts, *ε*_1_ and *ε*_2_ are stochastic environmental error terms, ***Y*** is the *n* dimensional vector for *n* individuals, ***X*** *_l_* indicates the allelic dosage for SNP *l* in *n* individuals, *β_l_* indicates the effect size of SNP *l*, ***G****_g_* = ∑_*i*∈*gene*(*g*)_ *ω_ig_****X****_i_* indicates the predicted expression calculated by *ω_lg_* and ***X*** *_l_*, in which *ω_lg_* is generated by using the GTEx tissue-specific eQTL dataset, and *γ_g_* is the effect size of ***G****_g_* . The Z-score (Wald-statistic) of the association between predicted gene expression and COVID-19-related phenotypes can be written as:

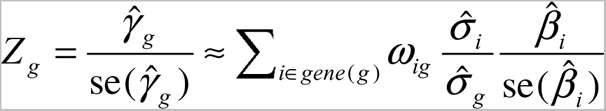

Where 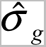 indicates the standard deviation of 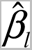 represents the effect size from GWAS on COVID-19 and 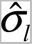 indicates the standard deviation of 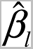. For each COVID-19-related phenotype, S-PrediXcan-based integration analysis is conducted for each of 49 tissues.

To enhance the power to distinguish potential causal genes, S-MultiXcan [40] is adopted to meta-analyze the substantial shared eQTLs across 49 GTEx tissues. By taking into account the correlation structure across multiple panels, the multivariate linear regression model of S-MultiXcan is fitted as the following equation:

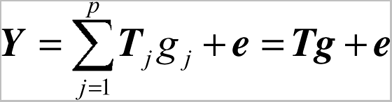

Where 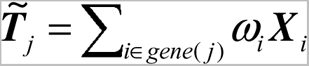 indicates the predicted expression of tissue *j*, and ***T****_j_* indicates the standardization of 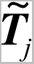 to *mean* = 0 and *standard Deviation* = 1 . *g _j_* indicates the effect size for the predicted gene expression in tissue *j*, ***e*** represents a stochastic environmental error term with variance 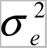, and *p* represents the count of chosen tissues. A gene with false discover rate (FDR) ≤ 0.05 is considered to be of significance.

### MAGMA-based gene-level association analysis

To conduct gene-level genetic association analyses of meta-GWAS summary statistics on three phenotypes of COVID-19, we apply the updated version SNP-wise Mean Model of the Multi-marker Analysis of GenoMic Annotation (MAGMA) [111]. Using this model, MAGMA computes a test statistic as the following algorithm:

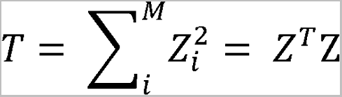

where *M* is the number of variants (e.g., SNP_1_, SNP_2_, …, SNP_i_, *i* ∈ M) in a given gene *g*(*j*), *j* ∈ *N. N* is total number of genes annotated in the GWAS summary dataset. We assign a specific SNP to a given gene *g* according to the location of the SNP whether located into the gene body or within an extended +/- 20 kb upstream or downstream region of the gene. Notably, *Z_i_* = ϕ(*p_i_*), where ϕ indicates the cumulative normal distribution function, and *p_i_* indicates the marginal p-value for a specific SNP *i*. Moreover, the gene-level converging model assumes ***Z*** ∼ MVN(***0***, ***S***), where ***S*** is the LD matrix among SNPs. The LD matrix can be diagonalized and written as ***S*** = ***QAQ****^T^*, where ***Q*** is an orthogonal matrix and ***A***= *diag*(*λ*_1_, *λ*_2,…_ *λ*_M_) with *λ_m_* being the *m*th eigenvalue of ***S*** .The 1,000 Genomes Project Phase 3 Panel[112] is adopted as a reference for calculating the LD matrix. ***D*** ∼ MVN(***0***, ***I****_K_*) indicates a random variable, where ***D*** = ***A^-0.5^Q^T^ Z*** . Thus, the sum of squared SNP Z-statistics can be calculated:

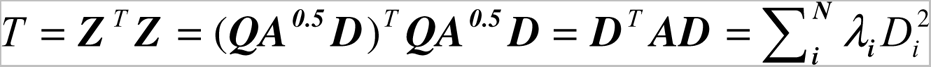

Where *D_i_* ∼ N(0,1) and 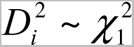 follows a mixture distribution of independent 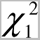 random variables. The Benjamini-Hochberg FDR method is used to adjust for multiple testing correction, and a gene with FDR ≤ 0.05 is interpreted as significance.

### In silico permutation analysis

As referenced previous methods [24, 113, 114], an *in silico* permutation analysis of 100,000 times of random selections is leveraged for assessing the concordance of findings between S-MultiXcan and MAGMA analyses across three COVID-19 outcomes. The notation of *G*_1_ represents the number of genes identified from the S-MultiXcan analysis, and *G*_2_ is the number of genes identified from the MAGMA analysis. At first, we count the overlapped genes between *G*_1_ and *G*_2_ (N*_Observation_* = *G*_1_ ∩ *G*_2_). Then, we adopt the total genes in the MAGMA analysis as background genes (*G_Background_*). By randomly selecting the same number of genes as gene set *G*_2_ from the background genes *G_Background_*, and after repeating it 100,000 times (*N_Total_*), we count the overlapped genes between gene set *G*_1_ and the sample randomly selected each time (*N_Random_*).We compute the empirically permuted P value as follows: 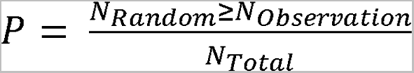, and empirical P value ≤ 0.05 is treated as significance. To measure the similarity between gene sets from S-MultiXcan and MAGMA analyses, we further leverage the Jaccard Similarity Index (JSI) [115], which is defined as the intersection size divided by the union size of both gene sets: 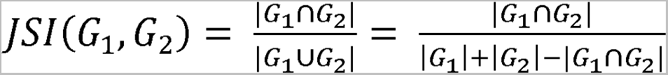, where 0 ≤ *JSI*(*G*_1_, *G*_2_) ≤1.

### Functional enrichment analysis

To elucidate the biological functions of S-MultiXcan- and MAGMA-identified risk genes for COVID-19 outcomes, we conduct functional enrichment analyses by using the WEB-based Gene SeT AnaLysis Toolkit (WebGestlat, http://www.webgestalt.org/) [116] with default parameters based on the KEGG [106] and Gene Ontology (GO) database [117]. The biological process category, which is removed the redundant GO terms, is used in the GO-based functional enrichment analysis. Moreover, we also performed KEGG pathway enrichment analyses by using significantly up-regulated genes in scPagwas-identified positive cells among lung mesenchymal stem cells (MSCs), intestinal tuft cells, and brain endothelial cells. The over-representation algorithm is leveraged to compute the significant level for each enrichment analysis, and the Benjamini-Hochberg FDR method is applied for multiple correction.

### LDSC analysis

The LDSC (version 1.0.1) method [51] is used to evaluate the genetic correlations between each of three COVID-19 phenotypes and each of 66 complex diseases/traits from six main disease categories (Supplementary Table S2). Differences in genetic correlations are computed with a block *jackknife* method to compute their corresponding standard errors. The significant association threshold is set to P < 0.00025 (0.05/198) after stringent Bonferroni correction, and P < 0.05 is considered to be suggestively significant.

### Cell-type-specific prioritization analysis of gene-drug interacting pairs for COVID-19

To identify cell type-specific drug targets relevant to severe COVID-19, we used scDrugHunter (version 1.1.0) [61] to integrate multiple layers of omics evidence, including human organoid scRNA-seq data, GWAS summary statistics on very severe COVID-19, eQTL data from the GTEx project [118], and gene-drug interactions from the Drug Gene Interaction database (DGIdb v4.2.0, https://www.dgidb.org/) [119]. In reference to previous methods [120, 121], scDrugHunter employs multiple computational algorithms to extract four dimensional features, which include cell type specificity scores of genes, gene relevance score (reflecting the relevance of genes for traits of interest in a given cellular context), gene significance scores (reflecting the association between genes whose genetically predicted expression levels and interested traits), and gene-drug interaction scores. scDrugHunter then ranks and scales the descending order of gene-specific scores for each feature in a particular cell type and uses a synthetic measures method [122] to combine the scaled ranks from the four dimensional features to compute the area of the patch in the Radar Chart for each gene-drug pair (called the single-cell druggable gene score, scDGS), according to the following equation:

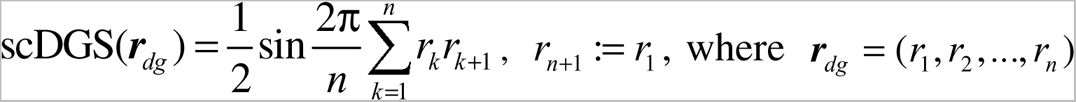 is a ranking vector for a gene-drug pair, and *n* is the number of extracted features (in this case, four-dimensional features). The threshold of scDGS ≥ 120 with permutation P value ≤ 0.05 is employed to repurpose cell-type-specific gene-drug pairs associated with the trait of interest.

### Cell-to-cell interaction analysis

To uncover potential cell-to-cell interactions of intestinal tuft positive cells and brain endothelial positive cells with other cells, we leveraged the CellChat (version 1.6.0) [123] R package to infer the predicted cellular communications based on two intestinal and brain organoids scRNA-seq datasets [65, 78]. The method of CellChat could examine the significant level of ligand-receptor interactions among different types of cells depended on the expression of soluble agonist, soluble antagonist, and stimulatory and inhibitory membrane-bound co-receptors. By summing the probabilities of these ligand-receptor pairs among a given pathway, CellChat could compute the communication probability for the pathway. The incoming (i.e., treating cells as target) and outgoing (i.e., treating cells as resource) interaction strength for each cell type was calculated by counting the number of significant ligand-receptor pairs.

### Statistical analysis

The Wilcoxon sum-rank test is utilized to calculate the significant level between positive cells and negative cells of lung MSCs, intestinal tuft cells, and brain endothelial cells. The hypergeometric test is used in KEGG-pathway-based and GO-term-based enrichment analyses to identify notable pathways and biological processes [116]. The Pearson correlation method is applied to compute the correlation coefficients of scPagwas TRSs [27] with scDRS TRSs [56], genetic risk scores, pseudotimes, and fibroblast cell scores, respectively. The paired Student’s T test is used to assess the difference in the number of ligand/receptor interactions with other cells between positive cells and negative cells in intestinal tuft and brain endothelial cells. The RISmed (version 2.3.0) [39] is used to perform a PubMed search for resorting to reported evidence supporting the association between COVID-19 and a given cell type or drug (see Supplementary Methods).

## Declarations

## Data Availability

All data produced in the present study are available upon reasonable request to the authors

https://www.covid19hg.org/results

https://zenodo.org/record/3518299#.Xv6Z6igzbgl

https://www.ncbi.nlm.nih.gov/gds

https://www.ebi.ac.uk/biostudies/arrayexpress

https://schob.su-lab.org/function

## Acknowledgements

We appreciate Dr. Hui Liu from the School of Ophthalmology & Optometry and Eye Hospital, Wenzhou Medical University for providing helpful suggestions, and our appreciation also goes to all authors from the COVID-19 Host Genetic Consortium who have deposited and shared GWAS summary statistics on various phenotypes of COVID-19 and goes to the authors who publicly released all human organoids and fetal scRNA-seq data in the public databases.

## Authors’ contributions

Y.M. and J.S. conceived and designed the study. Y.M., Y.J.Z., W.D., F.Q., C.D., J.L., Y.R.Z., D.J., G.Z., Y.Y., H.S., S.X., and H.H contributed to management of data collection. Y.M., F.Q., Y.J.Z., H.S., and Y.Y. conducted bioinformatics analysis and data interpretation. Y.M., J.S., N.W., and J.Q. wrote the manuscripts. All authors reviewed and approved the final manuscript.

## Data and materials availability

All the GWAS summary statistics used in this study can be accessed in the official websites (www.covid19hg.org/results). The GTEx eQTL data (version 8) were downloaded from Zenodo repository (https://zenodo.org/record/3518299#.Xv6Z6igzbgl). All the human Organoids scRNA-seq data were downloaded from two databases of GEO (https://www.ncbi.nlm.nih.gov/gds) and ArrayExpress (https://www.ebi.ac.uk/biostudies/arrayexpress). We have assembled a comprehensive pan-organoids single-cell RNA-seq dataset, which is available through the Curated scHOB website (https://schob.su-lab.org/function/). The code to reproduce the results is available in a dedicated GitHub repository (https://github.com/mayunlong89/scHuman_organoids_COVID19).

## Ethics approval and consent to participate

Not applicable

## Consent for publication

Not applicable

