## Supplementary Figures for "Integration of human organoids single-cell transcriptomic profiles and human genetics repurposes critical cell type-specific drug targets for severe COVID-19"

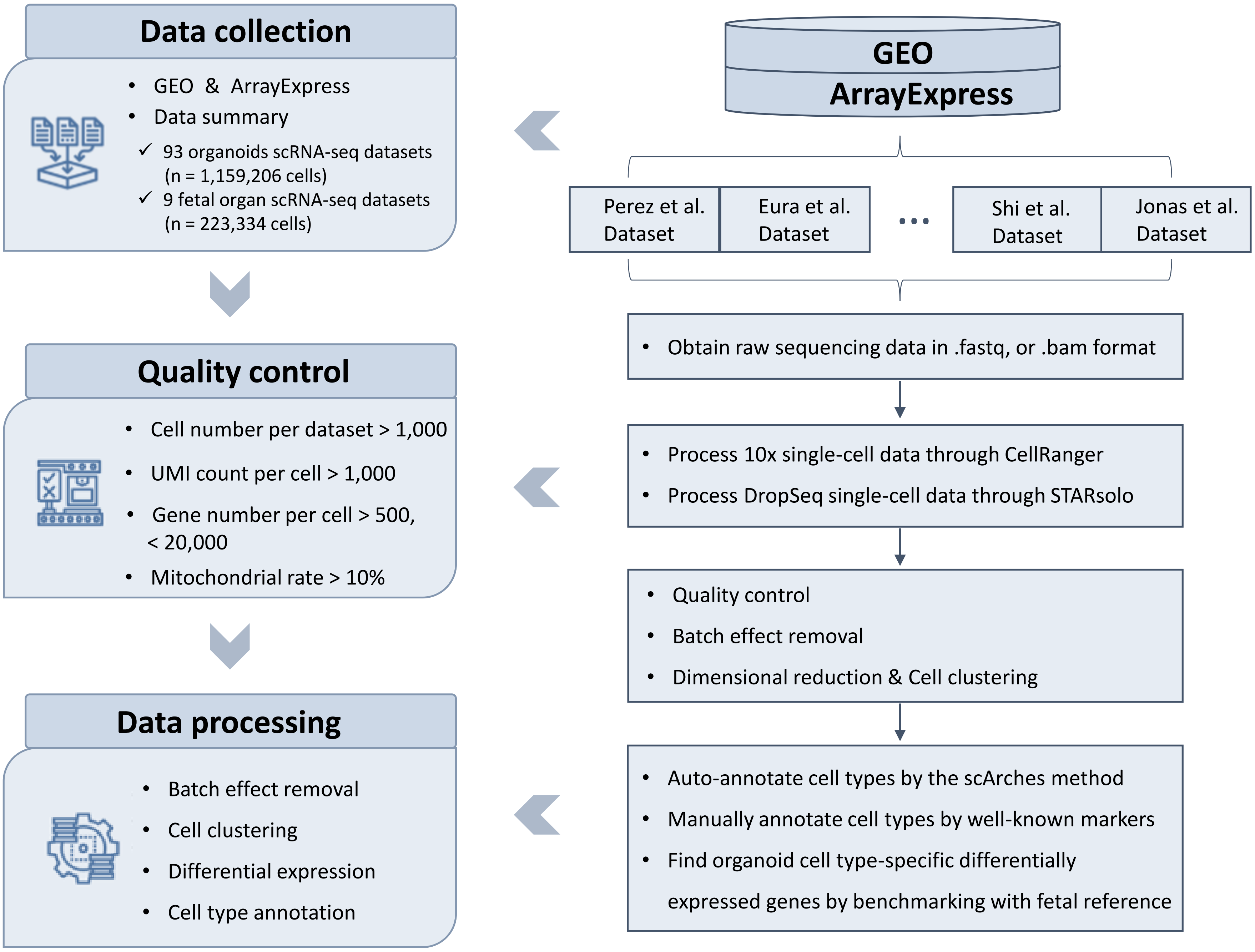


**Supplementary Figure S1. A unified pipeline for conducting re-alignment, quality control, and standard analysis of all human organoids and fetal scRNA-seq datasets.** Note: 1) Data collection, we collected human organoids and fetal scRNA-seq datasets generated from different platforms (i.e., 10x genomics and DropSeq) based on two widely-used databases of GEO and ArrayExpress. 2) Quality control, we downloaded the raw sequencing data from both databases, and used a series of tools to pre-process the data, including realignment and quality control with stringent criteria. 3) Data processing, we applied various widely-adopted bioinformatics methods to remove batch effect, reduce dimensions, cell clustering and cell type annotation.

| COVID-19 outcomes | GWAS datasets | Cases | Controls |
|---|---|---|---|
| 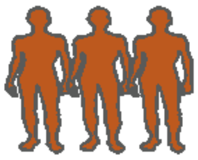 | Very severe COVID-19 vs. Population | 18,152 | 1,114,546 |
| 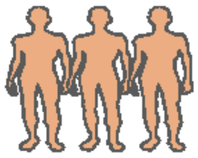 | Hospitalized COVID-19 vs. Population | 44,986 | 2,356,386 |
| 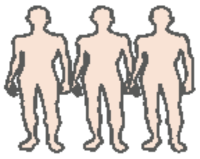 | Susceptible COVID-19 vs. Population | 159,840 | 2,782,977 |

**Supplementary Figure S2. Summary of the GWAS summary datasets on three COVID-19 outcomes.** Note, 1) The sample size of meta-GWAS on very severe respiratory confirmed COVID-19 (File name: A2_ALL_leave_23andme; n = 18,152 cases and 1,145,546 controls). 2) The sample size of meta-GWAS on hospitalized COVID-19 (File name: B2_ALL_leave_23andme; n = 44,986 cases and 2,356,386 controls). 3) The sample size of meta-GWAS on susceptible to COVID-19 (File name: C2_ALL_leave_23andme; n = 159,840 cases and 2,782,977 controls).

A

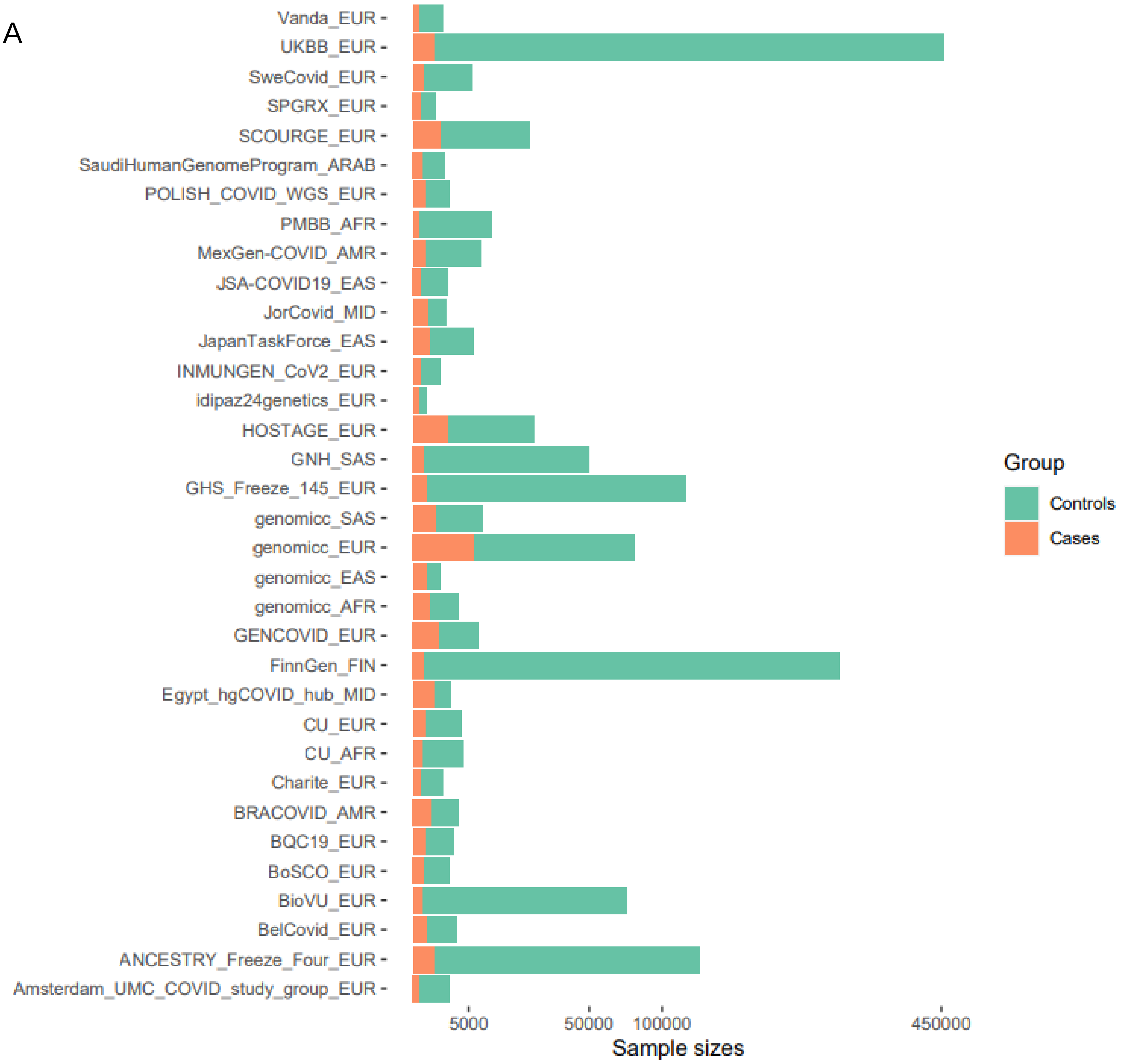


**Supplementary Figure S3A. Summary of the sample size of each cohort in COVID-19 meta-analytic GWAS summary data.** A. Bar plot shows the sample size of each cohort in meta-GWAS on very severe respiratory confirmed COVID-19 (File name: A2_ALL_leave_23andme; n = 18,152 cases and 1,145,546 controls). There were 34 individual GWAS datasets used for this meta-analysis on very severe respiratory confirmed COVID-19. All these meta-GWAS summary statistics (COVID19-hg GWAS meta-analyses round 7) were downloaded from the official website of the COVID-19 Host Genetics Initiative (https://www.covid19hg.org/results/r7/). Green color indicates the number of control samples, and orange color indicates the number of COVID-19 patient samples.

B

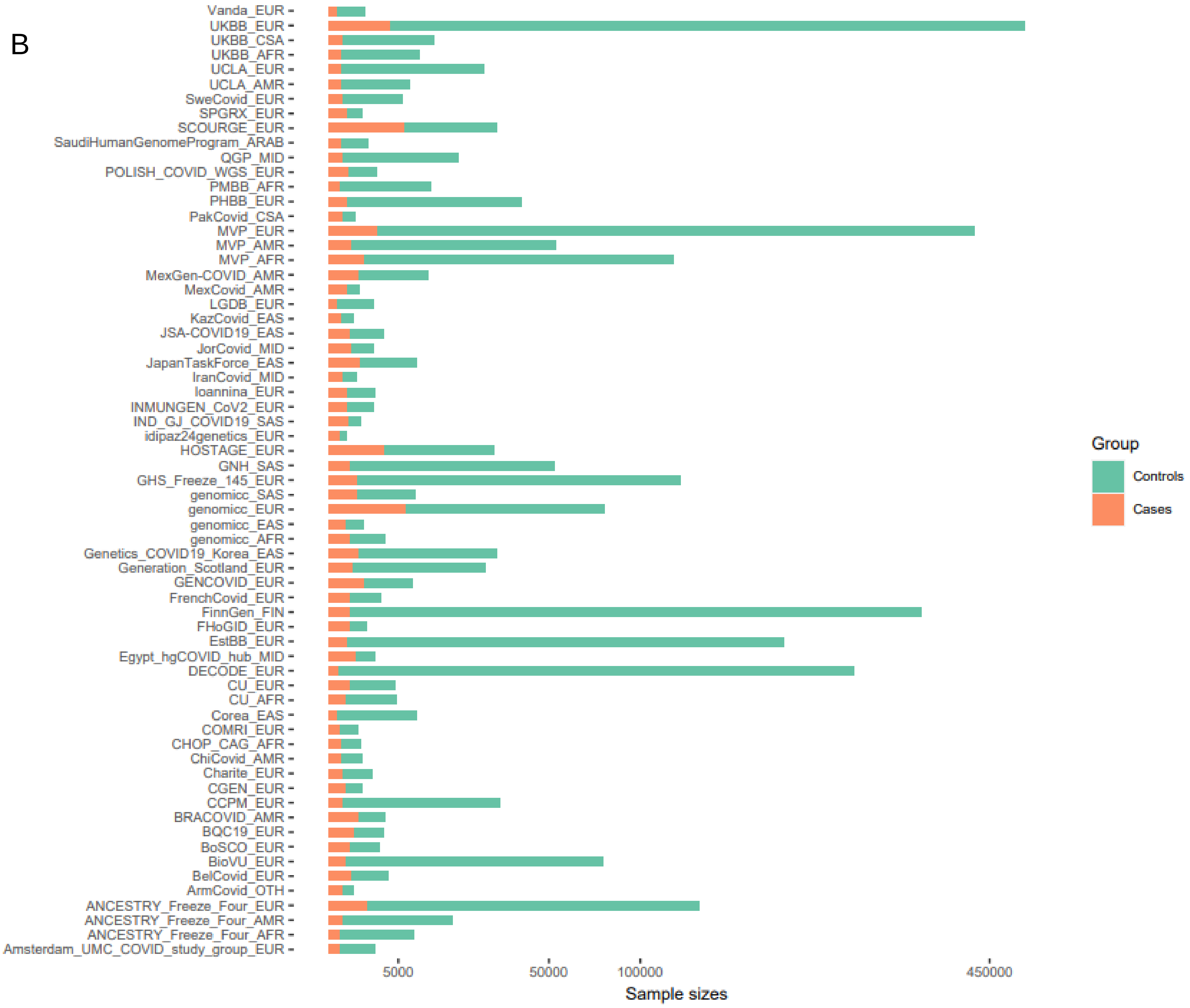


**Supplementary Figure S3B. Summary of the sample size of each cohort in COVID-19 meta-analytic GWAS summary data.** B. Bar plot shows the sample size of each cohort in meta-GWAS on hospitalized COVID-19 (File name: B2_ALL_leave_23andme; n = 44,986 cases and 2,356,386 controls). There were 65 individual GWAS datasets used for this meta-analysis on hospitalized COVID-19. All these meta-GWAS summary statistics (COVID19-hg GWAS meta-analyses round 7) were downloaded from the official website of the COVID-19 Host Genetics Initiative (https://www.covid19hg.org/results/r7/). Green color indicates the number of control samples, and orange color indicates the number of COVID-19 patient samples.

C

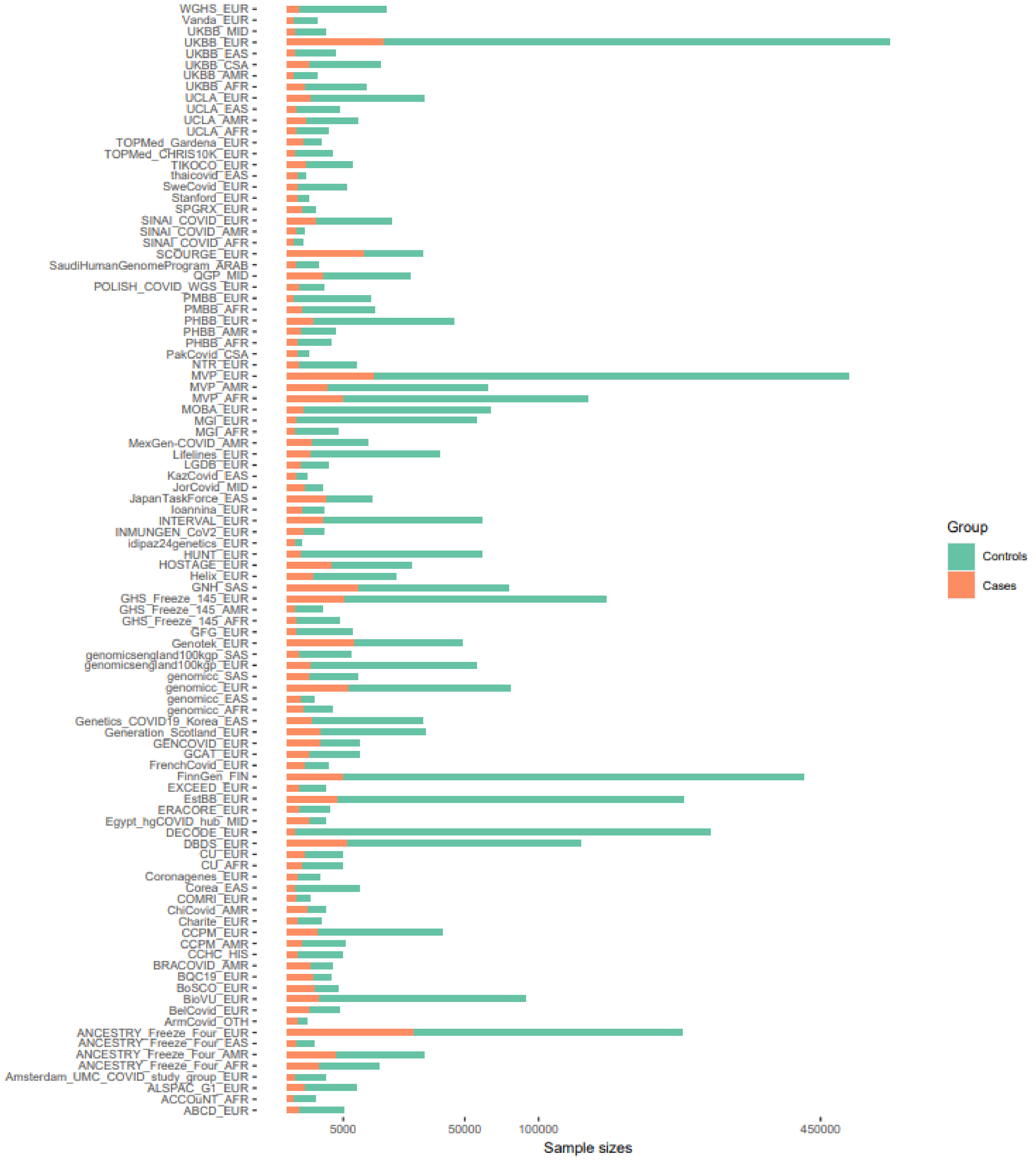


**Supplementary Figure S3C. Summary of the sample size of each cohort in COVID-19 meta-analytic GWAS summary data.** C. Bar plot shows the sample size of each cohort in meta-GWAS on susceptible to COVID-19 (File name: C2_ALL_leave_23andme; n = 159,840 cases and 2,782,977 controls). There were 100 individual GWAS datasets used for this meta-analysis on susceptible to COVID-19. All these meta-GWAS summary statistics (COVID19-hg GWAS meta-analyses round 7) were downloaded from the official website of the COVID-19 Host Genetics Initiative (https://www.covid19hg.org/results/r7/). Green color indicates the number of control samples, and orange color indicates the number of COVID-19 patient samples.

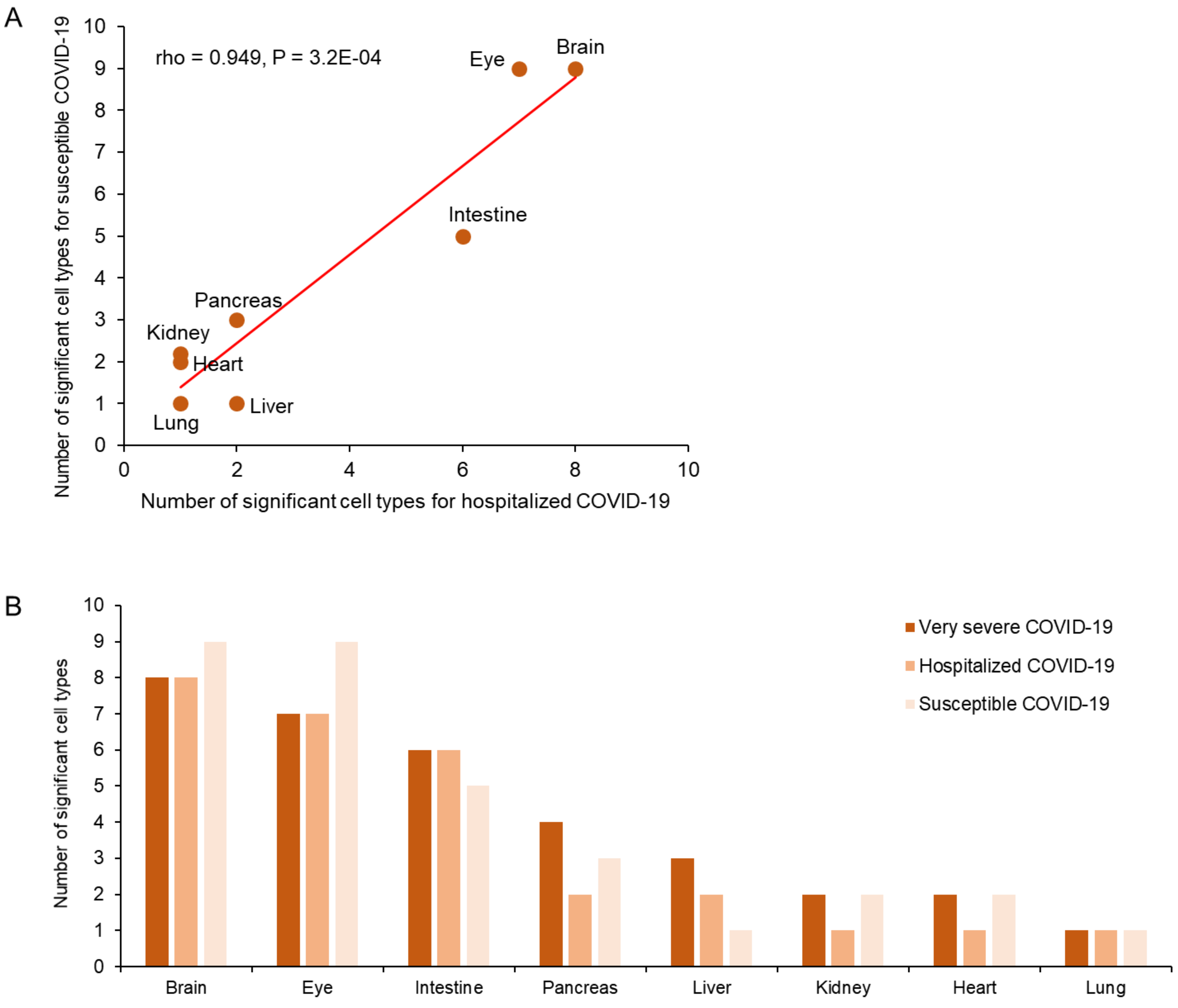


**Supplementary Figure S4. Consistent cell type enrichment results for three COVID-19 outcomes based on eight human organoids scRNA-seq data.** A. Correlation results of number of significant cell types in 8 human organoids between hospitalized COVID-19 and susceptible COVID-19. B. Barplot showing the number of significant cell types across 8 human organoids for three COVID-19 phenotypes. This result is related to Figure 3. scPagwas analysis of integrating GWAS summary statistics on three COVID-19 outcomes (i.e., very severe COVID-19, hospitalized COVID-19, and Susceptible COVID-19) and 10 kinds of human organoids scRNA-seq data in the scHOB database. There were 33 cell types in 8 human organoids (i.e., brain, eye, intestine, pancreas, liver, kidney, heart, and lung) significantly enriched by three COVID-19 phenotypes-relevant genetic association signals.

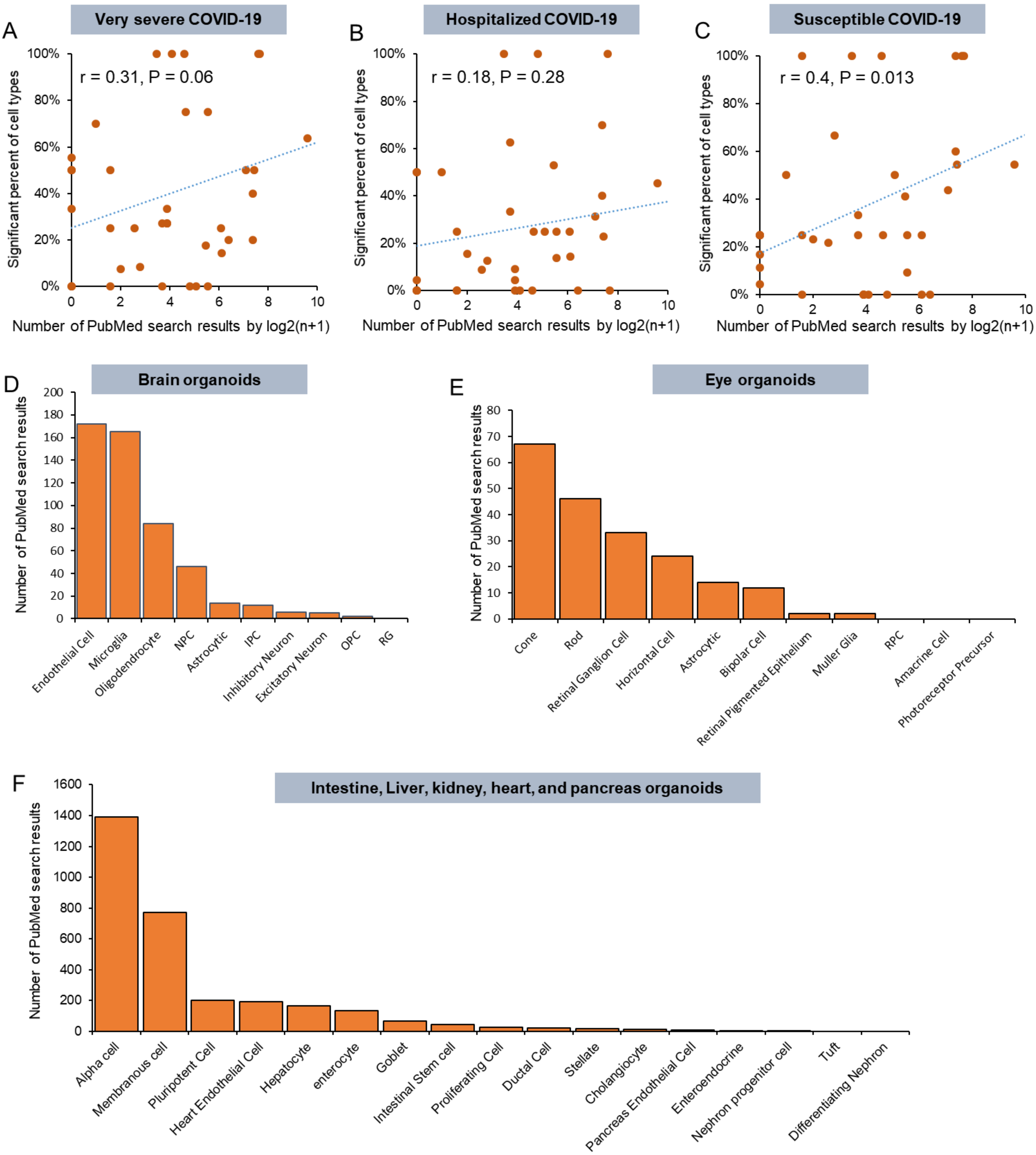


**Supplementary Figure S5. Correlations of scPagwas-identified significant cell types with PubMed Search results.** A. The Pearson correlation between the PubMed search results and scPagwas-identified significant cell types for very severe COVID-19. B. The Pearson correlation between the PubMed search results and scPagwas-identified significant cell types for hospitalized COVID-19. C. The Pearson correlation between the PubMed search results and scPagwas-identified significant cell types for susceptible COVID-19. D-F). Summary of the number of PubMed search results for each cell type among eight human organoids. Correlation coefficients and P-values from correlation analyses are included.

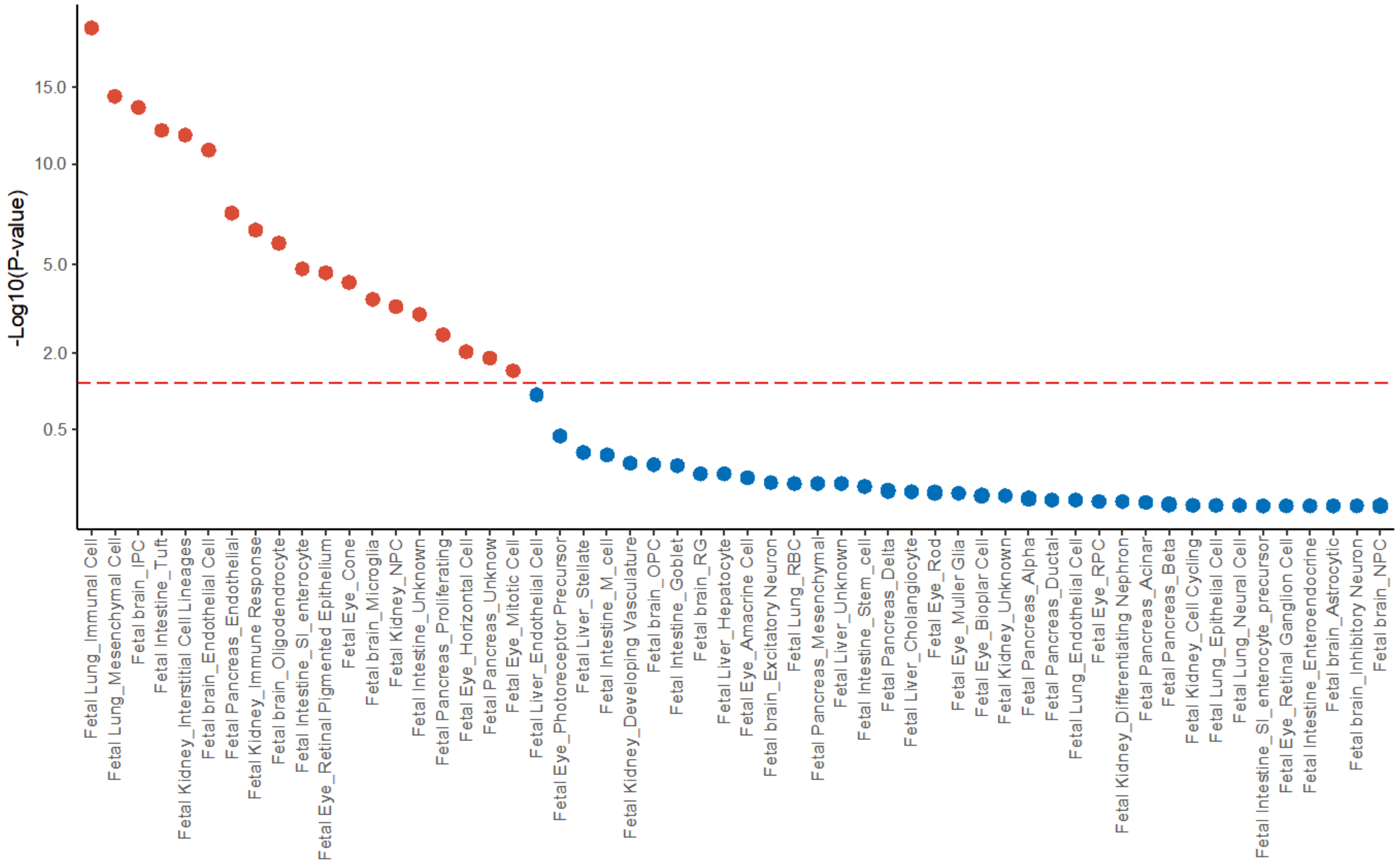


**Supplementary Figure S6. The results of scPagwas-identified cell types in fetal scRNA-seq data for very severe COVID-19.** The horizontal red dashed line represents the significant threshold ($P < 0.05$). Red dots in the plot indicated the significant cell types associated with very severe COVID-19, and blue dots indicated the non-significant cell types. The y axis shows the negative log-transformation P value (-log10(P-value), and x axis shows all annotated cell types in these included fetal scRNA-seq datasets. It should be noted that some cell types might not exist in human organoids scRNA-seq datasets. For example, immune cells in fetal lung were not in lung organoids.

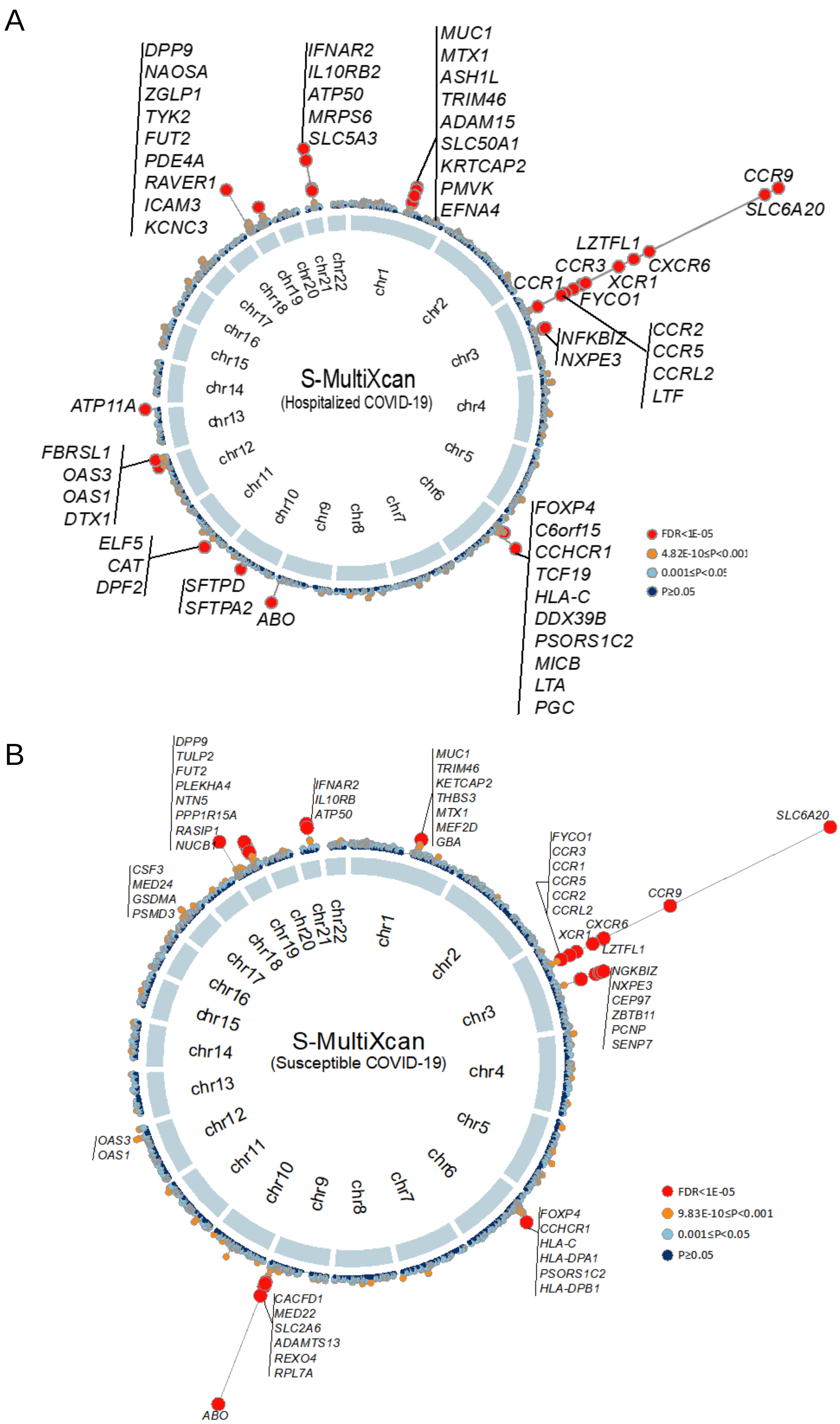


**Supplementary Figure S7. The results of S-MultiXcan-based analysis for hospitalized and susceptible COVID-19.** A. Circus plot showing the results of S-MultiXcan-based analysis for hospitalized COVID-19. B. Circus plot showing the results of S-MultiXcan-based analysis for susceptible COVID-19. The inner ring indicates the 22 autosomal chromosomes (Chr1-22). In the outer ring, a circular symbol indicates a specific gene and color marks the statistical significance of the gene for hospitalized or susceptible COVID-19 (Red color marks FDR < 1E-05, orange color indicates P < 0.001, light blue marks 0.001 ≤ P < 0.05, and dark blue indicates P > 0.05)

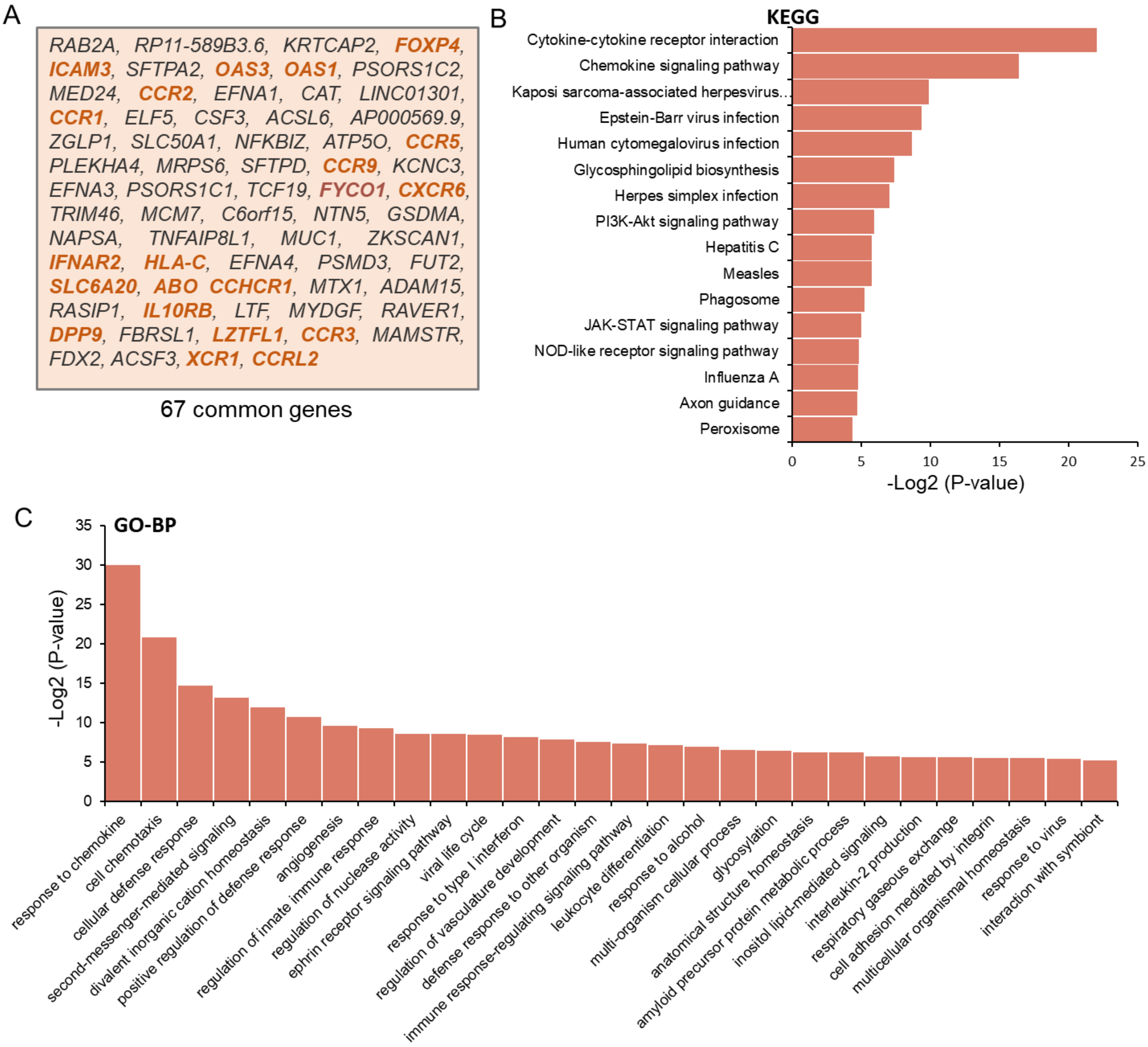


**Supplementary Figure S8. Functional annotation of common causal genes across three COVID-19 outcomes identified by S-MultiXcan analyses.** A. Summary of the 67 common causal genes among very severe COVID-19, hospitalized COVID-19, and susceptible COVID-19. B. Barplot showing the top-ranked KEGG pathways enriched by these 67 common genes. C. Barplot showing the top-ranked biological processes (GO-term) enriched by these 67 common genes. These functional enrichment analyses were performed by using the web-accessed tool of WebGestlat (http://www.webgestalt.org/) based on the KEGG pathway database and GO-term biological process terms with no redundant.

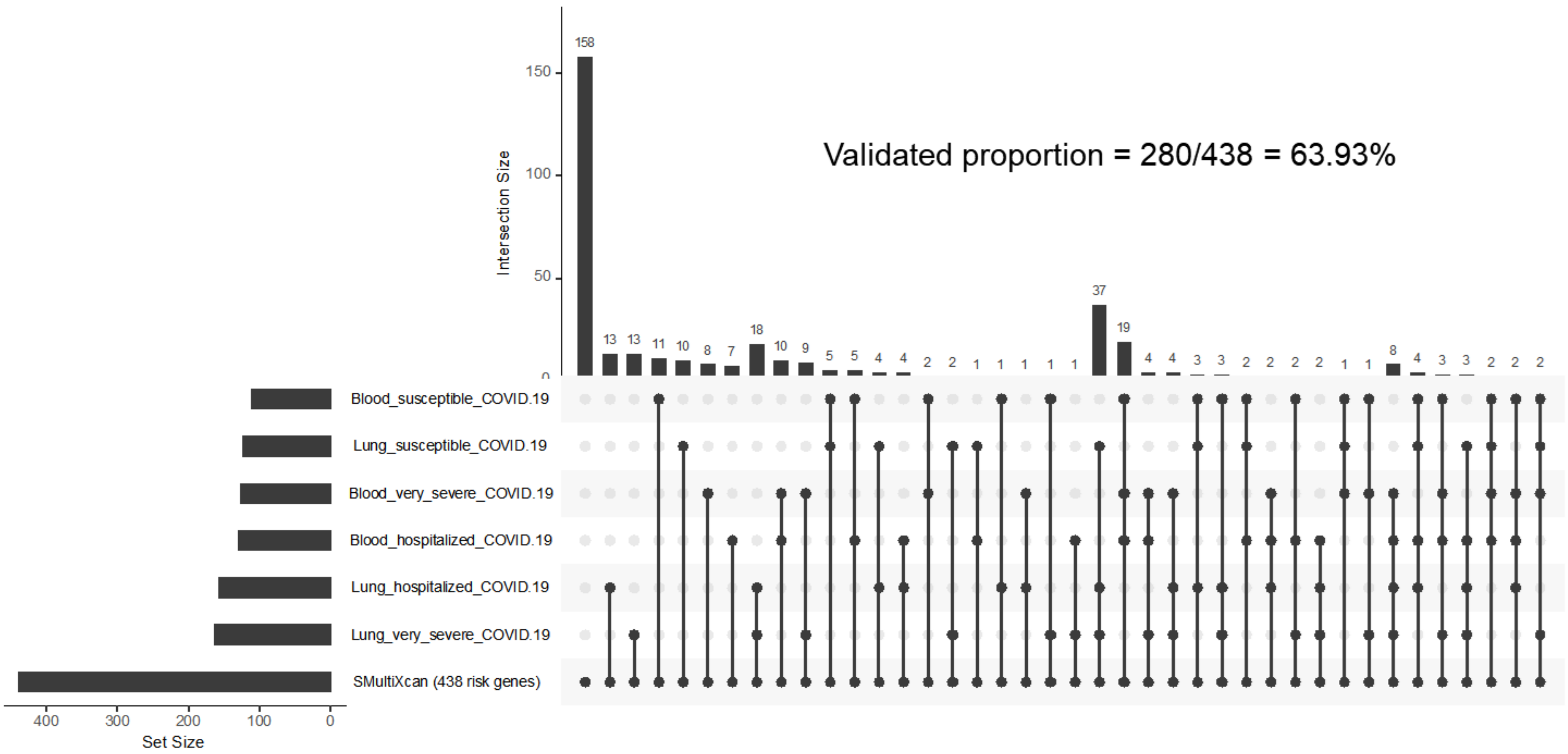


**Supplementary Figure S9. An UpSetR plot of COVID-19-risk genes across seven gene sets from both S-PrediXcan and S-MultiXcan integrative analyses.** For the seven gene sets, three blood-based and three lung-based S-PrediXcan analyses for three COVID-19 outcomes (i.e., very severe, hospitalized, and susceptible COVID-19, P < 0.05), and one S-MultiXcan-based gene set across three COVID-19 phenotypes (438 risk genes in total, FDR < 0.05). For each set that is part of a given intersection, a black filled circle is placed in the corresponding matrix cell, and a light gray circle is placed when a set is not part of the intersection. In each column, the topmost black circle is connected with the bottommost black circle by using a vertical black line for emphasizing the column-based relationships. The interaction sizes among gene sets are shown as a bar chart placed on the top of the matrix, and the gene set sizes are also shown as a bar chart placed on the left of the matrix.

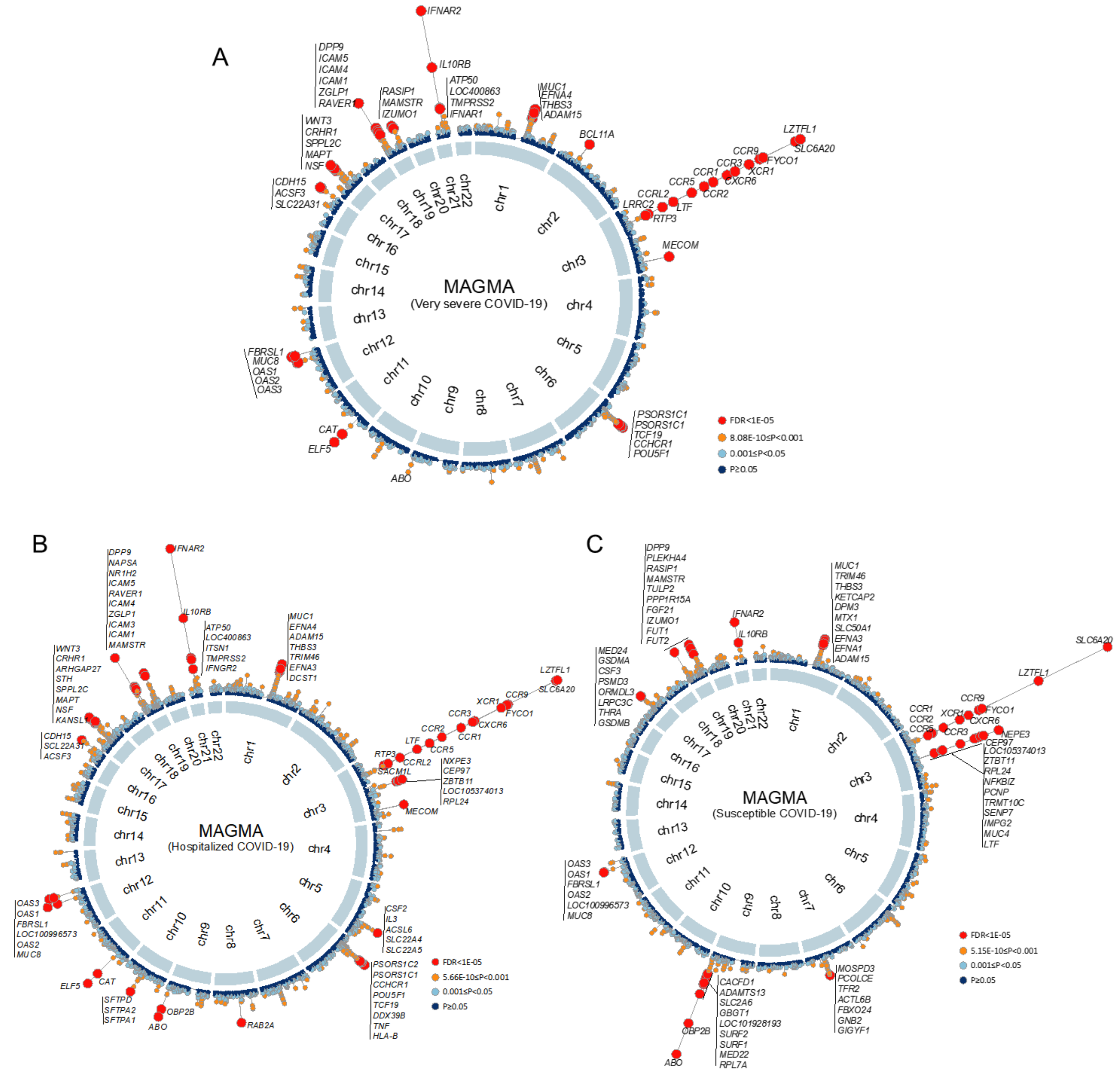


**Supplementary Figure S10. The results of MAGMA-based analysis three COVID-19 outcomes.** A. Circus plot showing the results of MAGMA-based analysis for very severe COVID-19. B. Circus plot showing the results of MAGMA-based analysis for hospitalized COVID-19. C. Circus plot showing the results of MAGMA-based analysis for susceptible COVID-19. Note: The inner ring indicates the 22 autosomal chromosomes (Chr1-22). In the outer ring, a circular symbol indicates a specific gene and color marks the statistical significance of the gene for hospitalized or susceptible COVID-19 (Red color marks FDR < 1E-05, orange color indicates P < 0.001, light blue marks 0.001 < P ≤ 0.05, and dark blue indicates P > 0.05). There were 391, 574, and 321 MAGMA-identified risk genes significantly associated with very severe, hospitalized, and susceptible COVID-19, respectively (FDR ≤ 0.05).

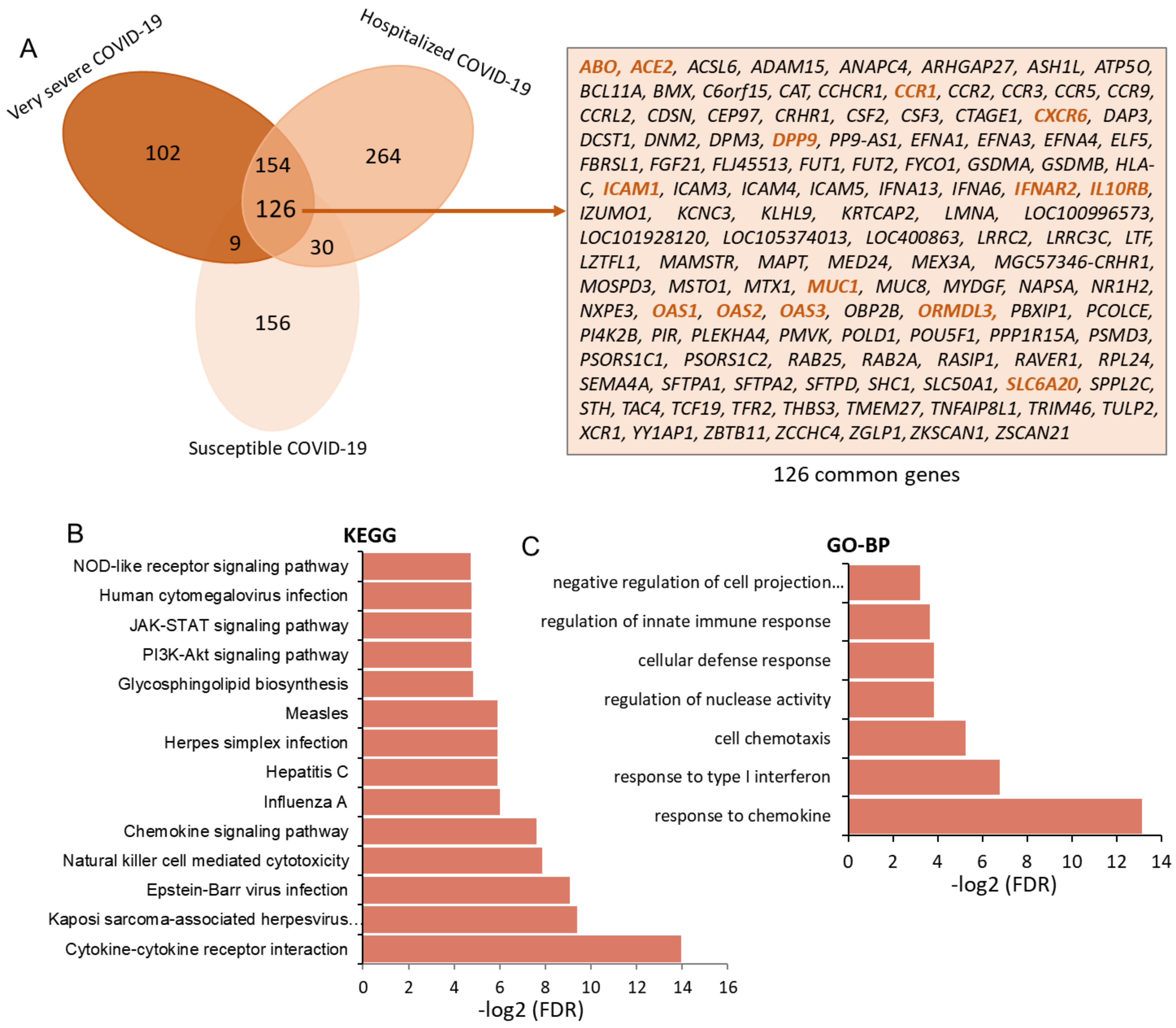


**Supplementary Figure S11. Functional annotation of 126 common genetic risk genes identified by MAGMA analyses across three COVID-19 outcomes.** A. Venn diagram showing the overlapped genes across very severe COVID-19, hospitalized COVID-19, and susceptible COVID-19. Right panel: Summary of the 126 common genetic risk genes across three COVID-19 outcomes. B. Barplot showing the top-ranked KEGG pathways enriched by these 126 common genes. C. Barplot showing the top-ranked biological processes enriched by these 126 common genes. These functional enrichment analyses were performed by using the web-accessed tool of WebGestlat (http://www.webgestalt.org/) based on the KEGG pathway database and GO-term biological process terms with no redundant.

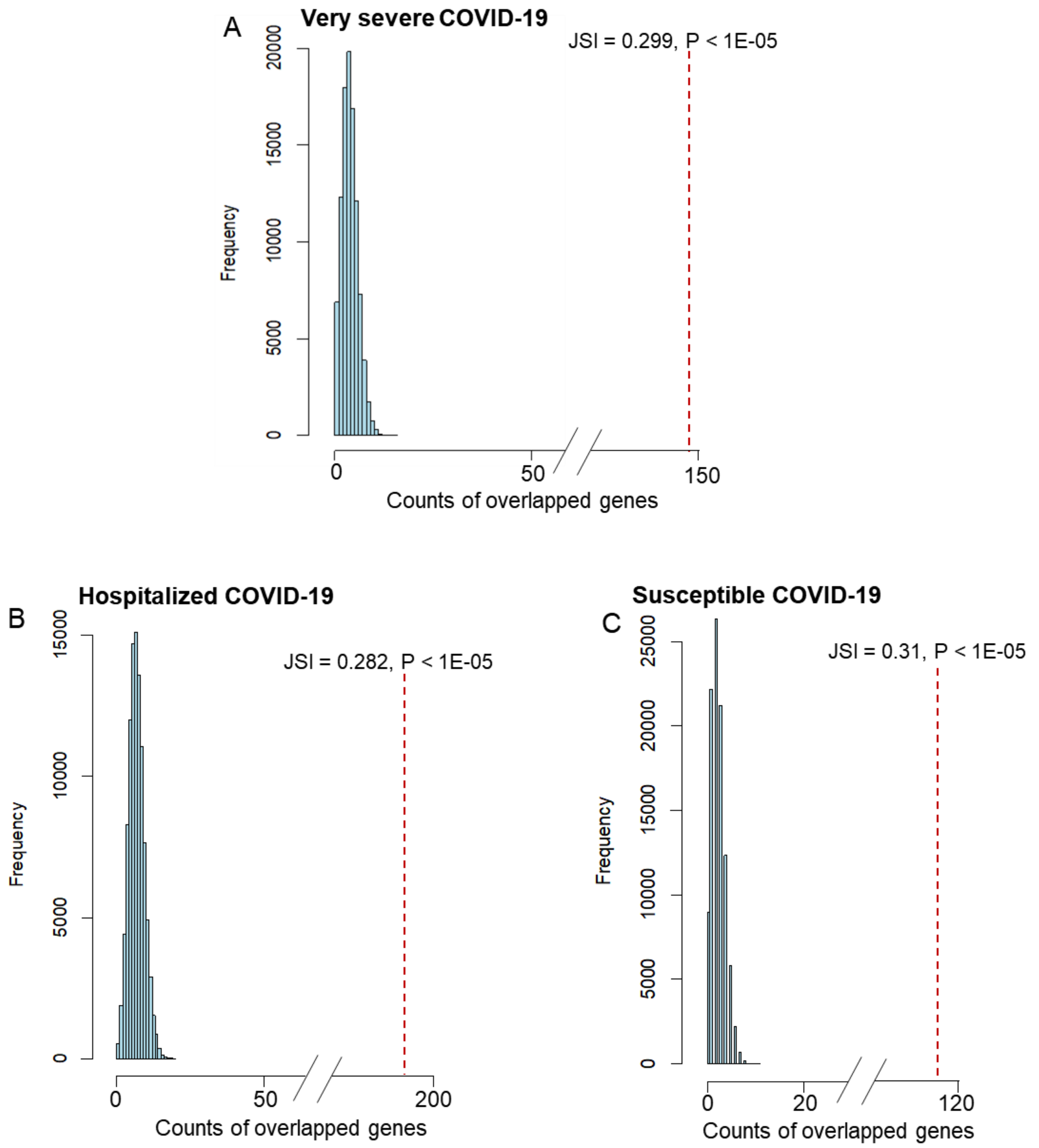


**Supplementary Figure S12. *In silico* permutation analysis of 100,000 times of random selections.** A. The comparison of top-ranked genes from MAGMA gene-based analysis (FDR < 0.05) with that from S-MultiXcan-based analysis (FDR < 0.05) on very severe COVID-19. B. The comparison of top-ranked genes from MAGMA gene-based analysis (FDR < 0.05) with that from S-MultiXcan-based analysis (FDR < 0.05) on hospitalized COVID-19. C. The comparison of top-ranked genes from MAGMA gene-based analysis (FDR < 0.05) with that from S-MultiXcan-based analysis (FDR < 0.05) on susceptible COVID-19. The vertical red line represents the observed overlapped count of genes between MAGMA and S-MultiXcan analyses on three COVID-19 outcomes. Permuted P value is calculated for assessing the significant level of the overlap, and Jaccard Similarity Index (JSI) is used for assessing the degree of similarity between two gene sets identified from MAGMA and S-MultiXcan analyses.

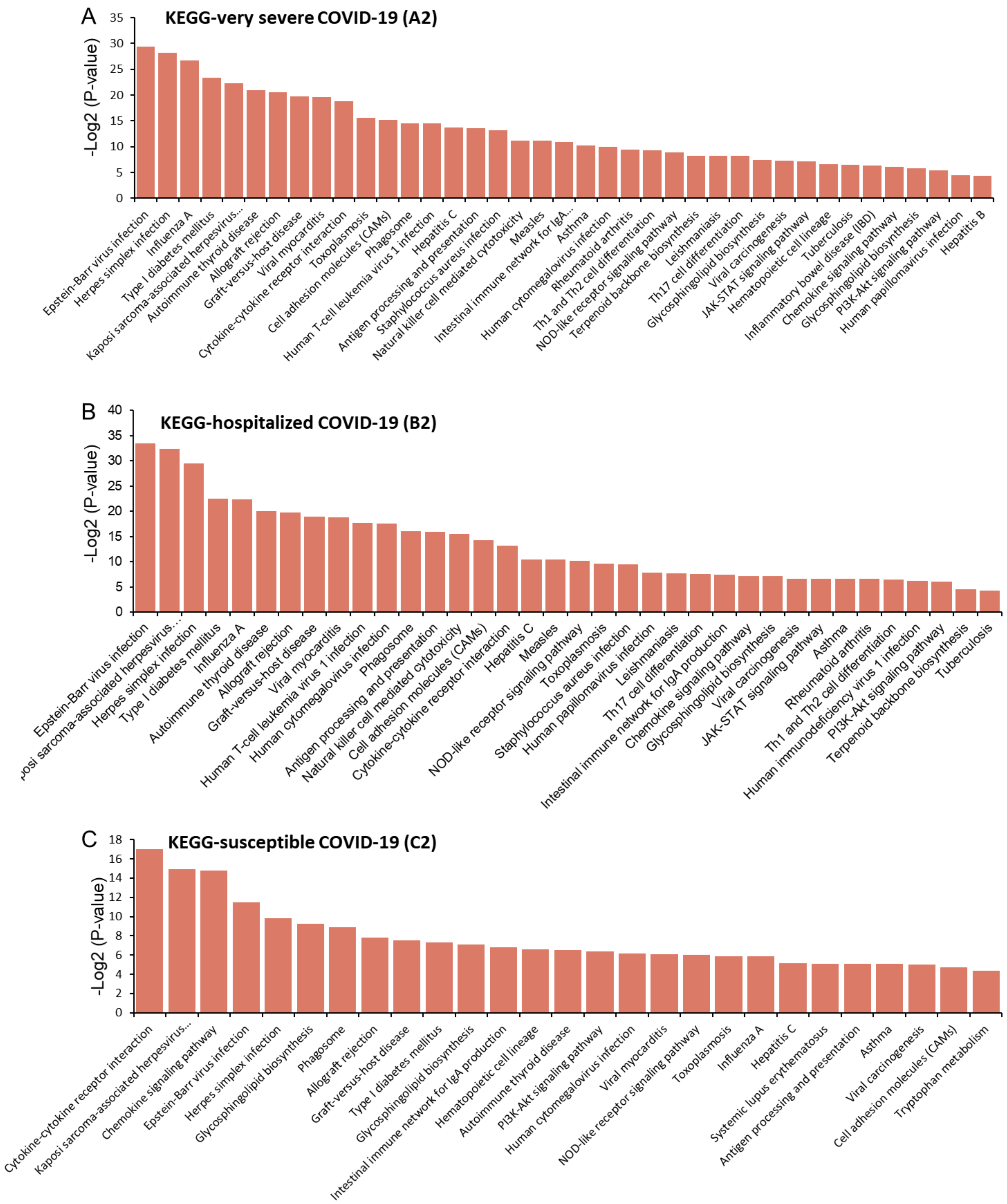


**Supplementary Figure S13. Pathway enrichment analysis for three S-MultiXcan-identified gene sets across three COVID-19 outcomes.** A. Barplot showing the significant KEGG pathways enriched by these 243 risk genes for very severe COVID-19 (A2). B. Barplot showing the significant KEGG pathways enriched by these 277 risk genes for hospitalized COVID-19 (B2). C. Barplot showing the significant KEGG pathways enriched by these 158 risk genes for susceptible COVID-19 (C2). These pathway enrichment analyses were performed by using the web-accessed tool of WebGestlat (http://www.webgestalt.org/) based on the KEGG pathway database.

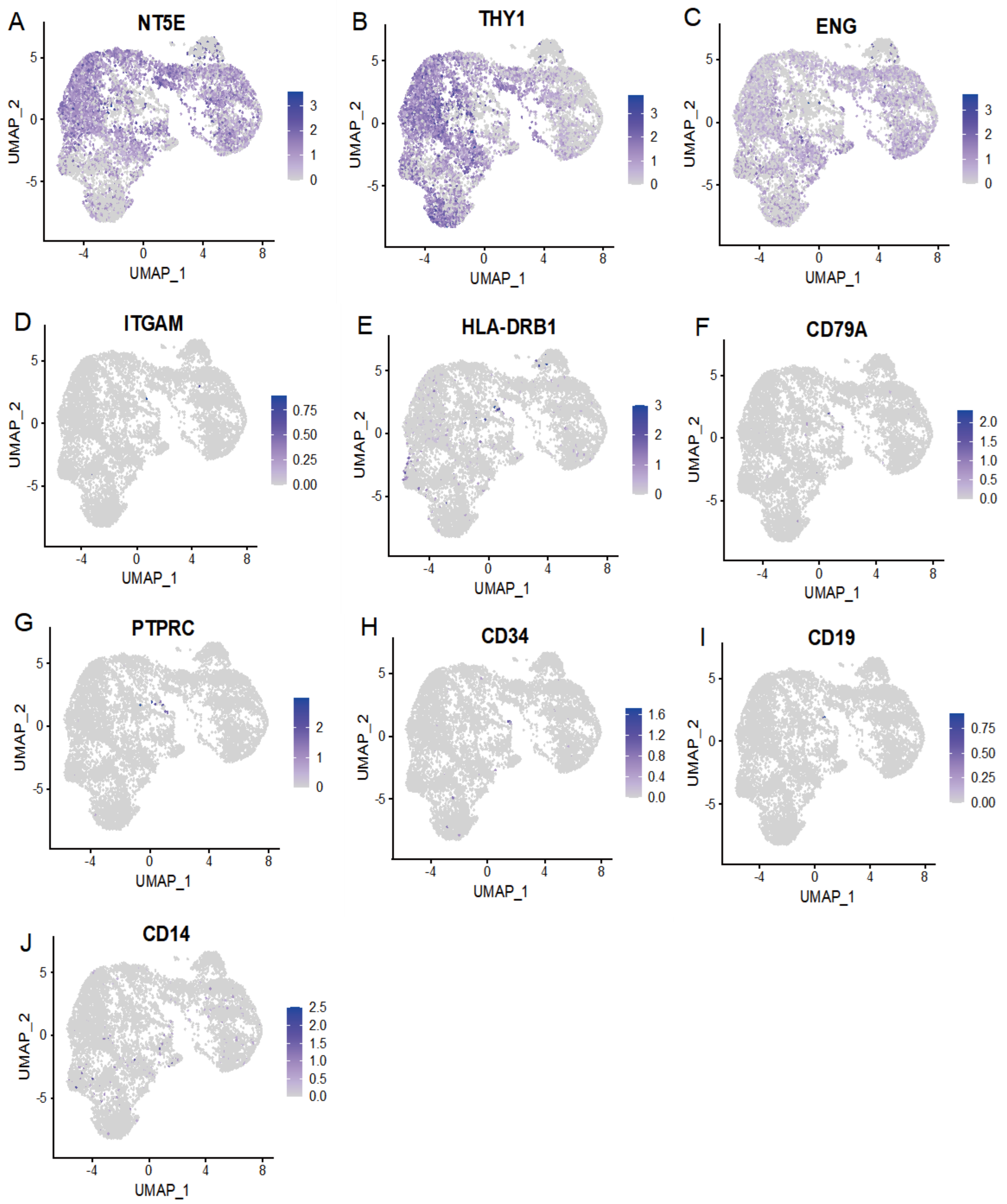


**Supplementary Figure S14. Normalized expression of common markers for human lung mesenchymal stem cells.** In the current study, we first used the scArches method to auto-annotate the cell type of mesenchymal stem cells (MSCs, n = 9,795 cells) in human lung organoids based on the human fetal lung scRNA-seq data as the reference. Then, to further annotate the cell type of MSCs, sets of cell surface markers must be expressed or absent from MSCs, which have been recognized by the International Society for Cellular Therapy (ISCT) as one of the minimal criteria for MSC identification. Expressed markers includes CD73 (NT5E), CD90 (THY1), and CD105 (ENG), and unexpressed markers includes CD11b (ITGAM), CD14, CD19, CD34, CD45 (PTPRC), CD79a (CD79A), and HLA-DR (HLA-DRB1).

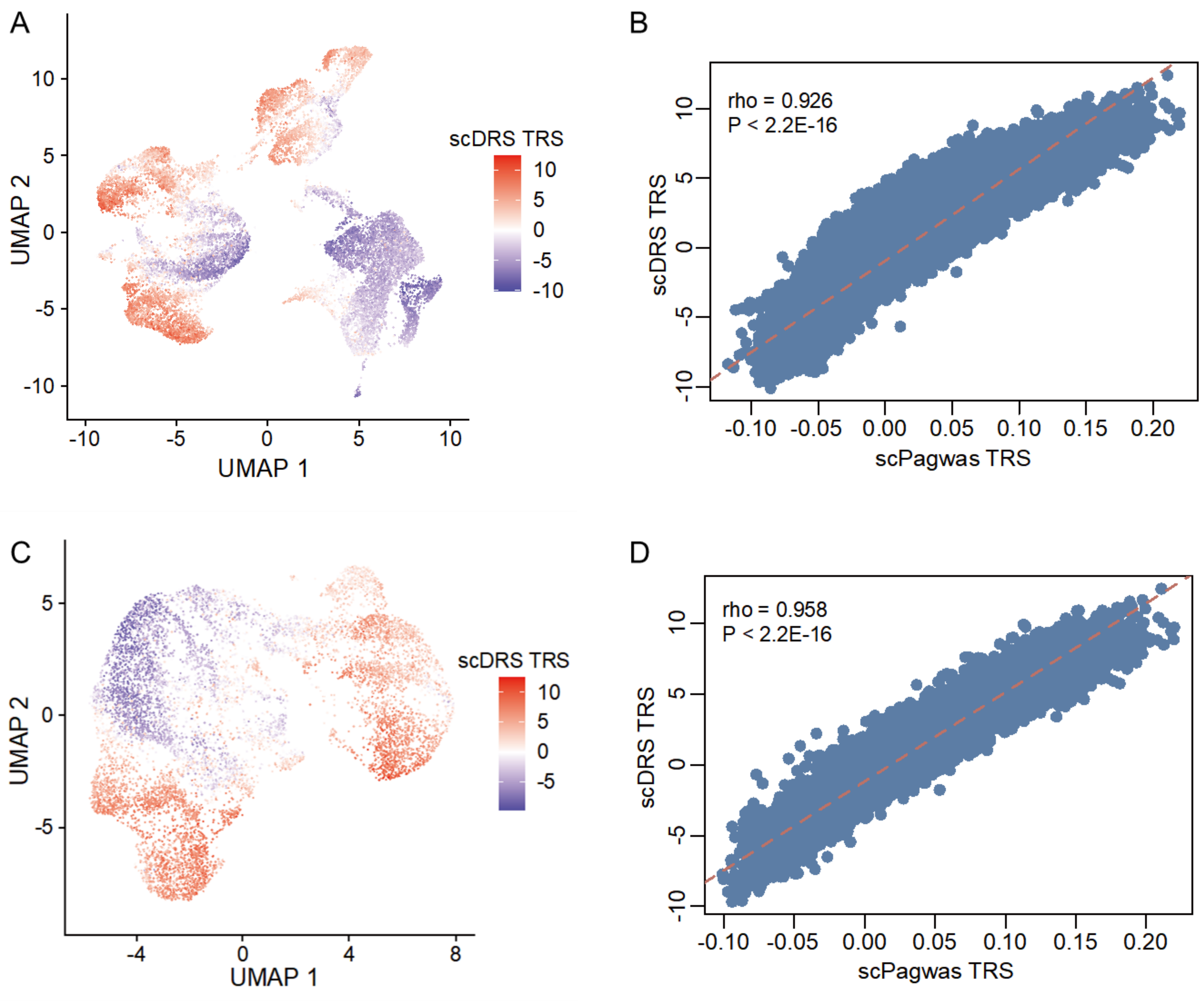


E

| | | scDRS | |
|---|---|---|---|
| | Proportion | Negative cells | Positive cells |
| scPagwas | Negative cells | 0.840 | 0.160 |
| | Positive cells | 0.075 | 0.925 |

Concordance rate = (0.84+0.925)/(0.84+0.16+0.075+0.925) = 0.8825

**Supplementary Figure S15. Concordance analysis of the single-cell association results between scDRS and scPagwas in lung MSCs.** A. UMAP plot shows that the scDRS TRSs for the phenotype of very severe COVID-19 is shown in lung organoids scRNA-seq data (n = 20,602 cells). Three main cell types of lung organoids included mesenchymal stem cells (MSCs, n = 9,795 cells), epithelial cells (n = 10,125 cells), and endothelial cells (n = 682 cells; Figure 5A). B. The Spearman correlation analysis of TRSs from between scDRS-based and scPagwas-based method in lung organoids. C. UMAP plot shows that the scDRS TRSs for the phenotype of very severe COVID-19 is shown in lung MSCs (n = 9,795 cells). D. The Spearman correlation analysis of TRSs from between scDRS-based and scPagwas-based method in lung MSCs. E. Concordance assessment of the positive cell results from scDRS and scPagwas in lung MSCs. scDRS estimates the significant level for each cell by using the Monte Carlo (MC) sampling method, and FDR < 0.05 is considered to be statistically significant (see Supplemental Methods for details).

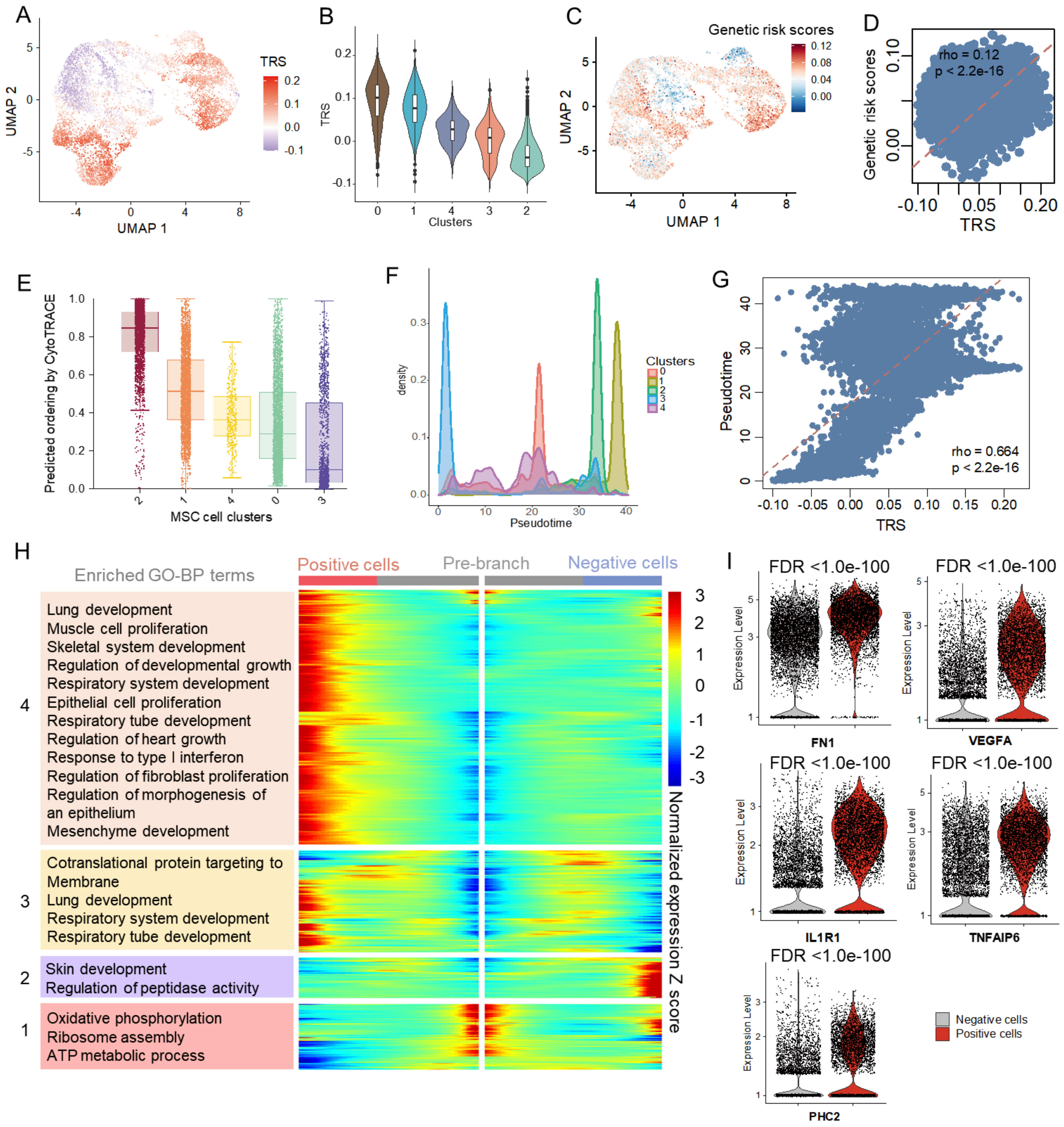


**Supplementary Figure S16. The characteristics of mesenchymal stem cells (MSCs) in human lung organoids.** A. UMAP plot shows that scPagwas TRSs for the phenotype of very severe COVID-19 is shown in lung MSCs (n = 9,795 cells). B. Violin plot showing the scPagwas TRSs in five clusters among MSCs. C. UMAP visualization of lung MSCs with five cell clusters colored by the genetic risk scores (GRSs) of 438 COVID-19-relevant genes. D. The Spearman correlation between the GRSs of 438 genes and scPagwas TRSs in lung MSCs. E. Boxplots showing CytoTRACE-predicted values for five cell clusters in lung MSCs. F. Density plots showing the distribution of pseudotimes for five cell clusters in lung MSCs. The pseudotime of each cell was calculated by using the Monocle 2 R package. G. The Spearman correlation of scPagwas trait-relevant scores (TRSs) with pseudotimes across all lung MSCs. H. Heatmap showing relative expressions of genes that are significantly branch dependent in lung MSCs. Columns are points in pseudotimes, rows are genes, and the beginning of pseudotime is in the middle of the heatmap. The left branch (red branch) shows the pseudotimes of cells related to positive cells, and the right branch (blue branch) shows the pseudotimes of cells related to negative cells. We used the branch-dependent genes to perform GO-term-biological process (BP) enrichment analysis. I. Violin plots show the expression levels of important up-regulated genes (*FN1, VEGFA, IL1R1, TNFAIP6*, and *PHC2*) in scPagwas-identified MSC positive cells. The Wilcoxon rank test was used to examine the significance between positive and negative cells, and FDR was used for multiple correction.

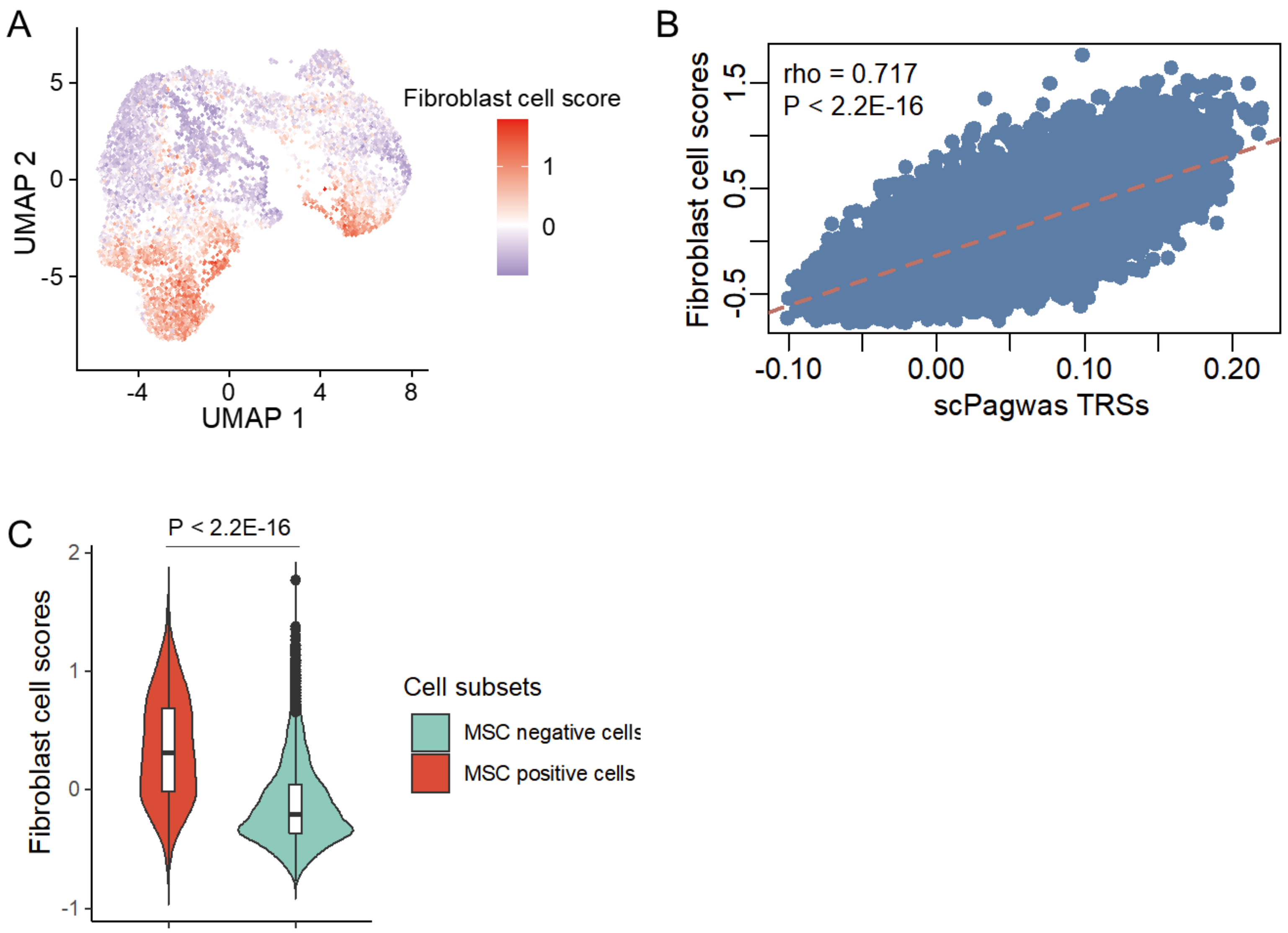


**Supplementary Figure S17. The cell state scores relevant to fibroblast calculated by using the expression levels of common fibroblast markers in lung MSCs.** A. UMAP embedding of lung MSCs with five cell clusters colored by the fibroblast cell state scores calculated by using the expression levels of common fibroblast markers in lung MSCs. We used the short-list of common fibroblast markers, including many extracellular matrix genes (ECM), i.e., *COL1A1, COL1A2* , *COL5A1, LOXL1, LUM, FBLN1, FBLN2*, and the cell surface receptors, i.e., *CD34* and *PDGFRA*, to calculate fibroblast cell state scores using the *AddModuleScore* function in the Seurat software. These fibroblast markers were collected from Muhl et al. study (PMID: 32769974). B. The Spearman correlation between the fibroblast cell state scores and scPagwas TRSs in lung MSCs. C. Violin plot showing the difference in fibroblast cell state scores between MSC positive cells and negative cells. A two-side Wilcoxon sum-rank test was used to assess the significance.

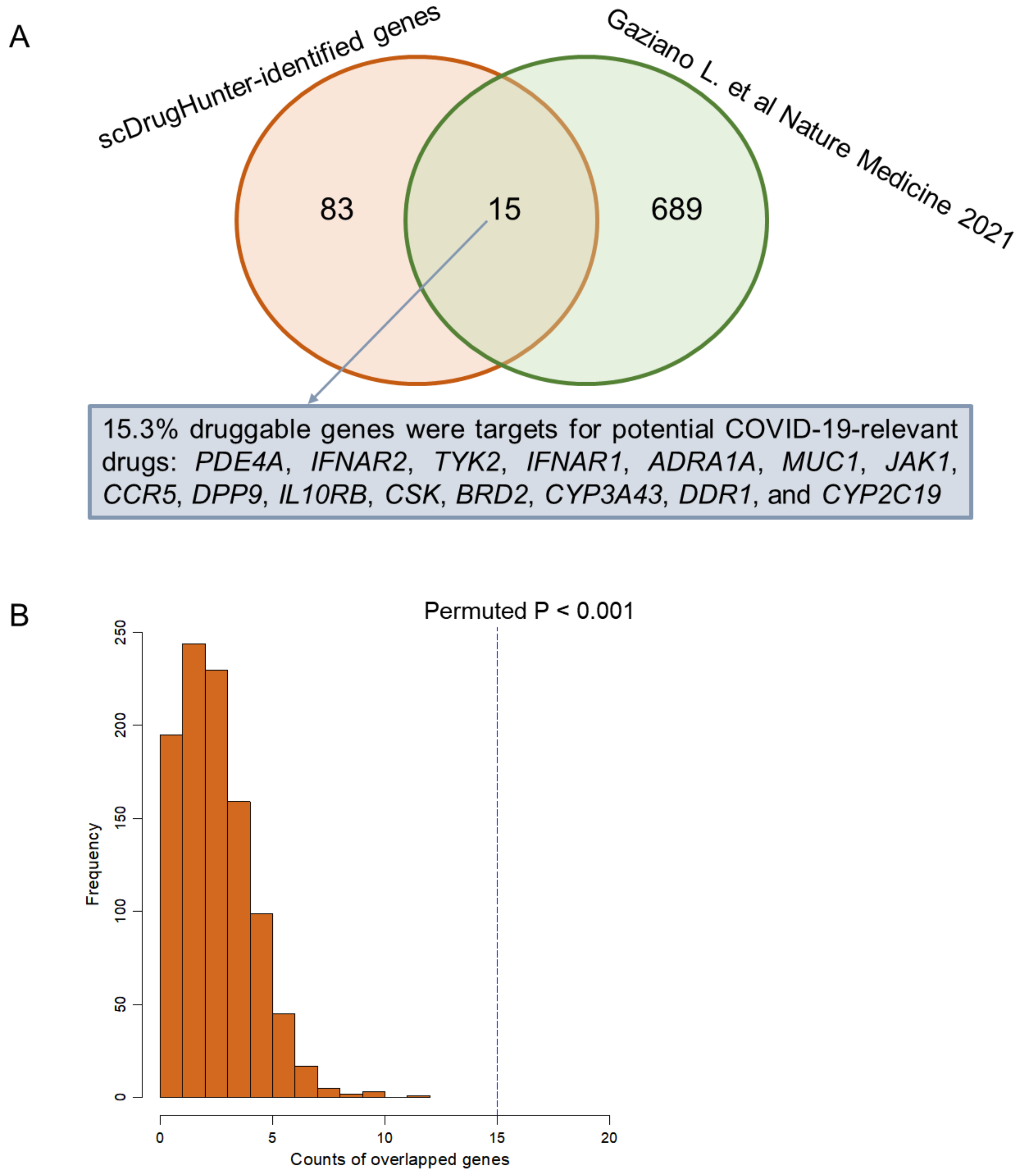


**Supplementary Figure S18. Permutation analysis for assessing the performance of scDrugHunter to identify MSC-specific druggable genes for a trait of interest.** A. The proportion of scDrugHunter-identified 98 druggable genes overlapped with 704 genes that are targets for potential COVID-19-relevant drugs based on registers of clinical trials for COVID-19, approved immunomodulatory/anticoagulant drugs, or have biological functions associated with SARS-CoV-2 infection reported by Gaziano L. et al. *Nature Medicine*, 2021 (PMID: 33837377). These 98 druggable genes that have at least one known drugs in the DGIdb database (DGIdb v4.2.0, https://www.dgidb.org/). B. *In silico* permutation analysis of 1,000 times of random selections from all background genes in the SMultiXcan analyses (N = 22,341 genes) to overlap with the 704 genes that are potential targets for COVID-19-relevant drugs. The vertical blue line represents the observed overlapped count of genes identified by scDrugHunter in MSCs.

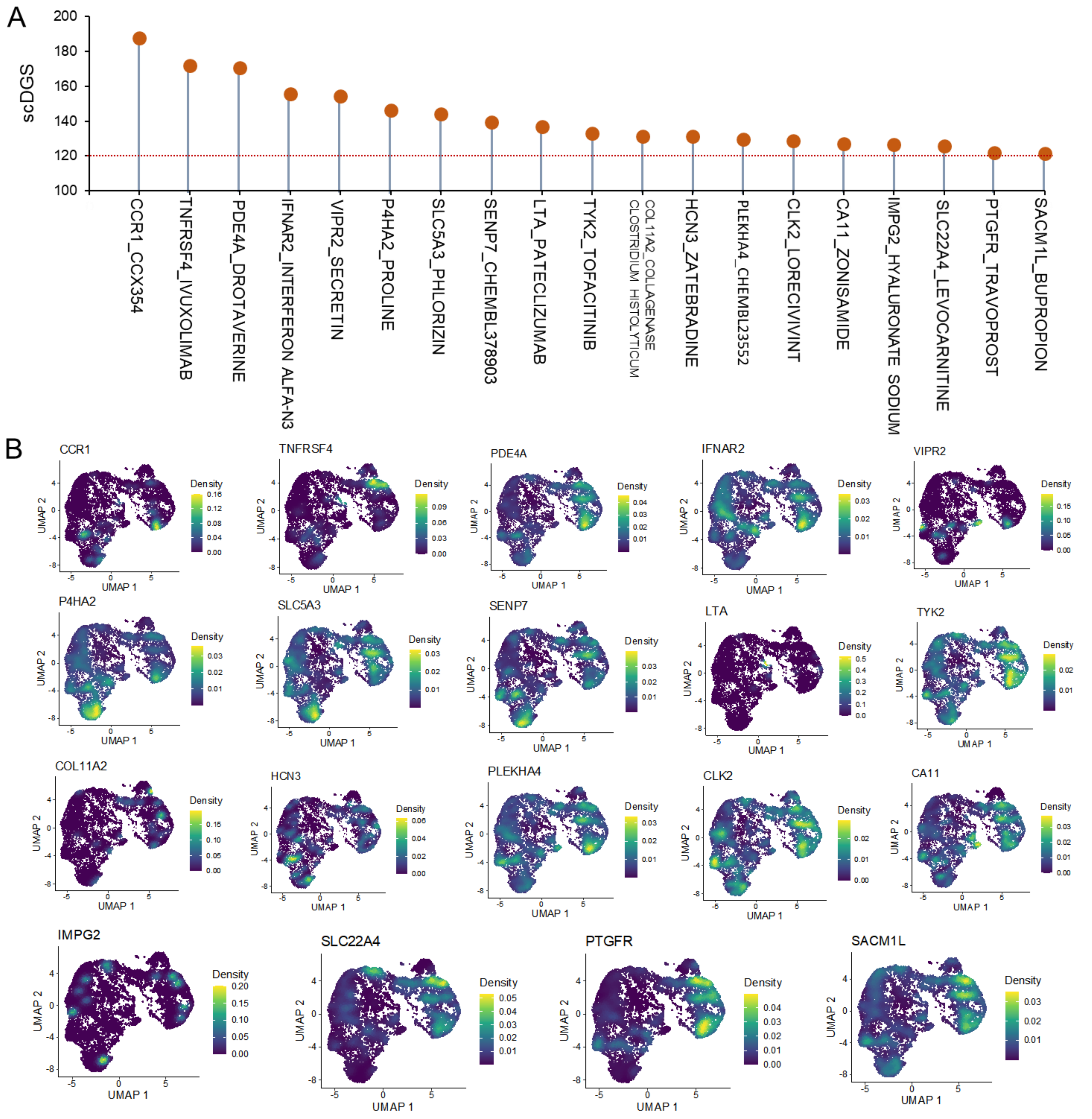


**Supplementary Figure S19. scDrugHunter-identified 19 druggable genes in lung MSCs.** A. The dot plot showing the result of scDrugHunter-identified 19 druggable genes and interacting drugs with high scDGS > 120 in lung MSCs. B. UMAP embedding plot showing the expression density of 19 druggable genes in lung MSCs. The colored bar represent the density of normalized expression of these 19 druggable genes identified by scDrugHunter in lung MSCs. A-Q). *CCR1, TNFRSF4, PDE4A, IFNAR2, VIPR2, P4HA2, SLC5A3, SENP7, LTA, TYK2, COL11A2, HCN3, PLEKHA4, CLK2, CA11, IMPG2, SLC22A4, PTGFR*, and *SACM1L*.

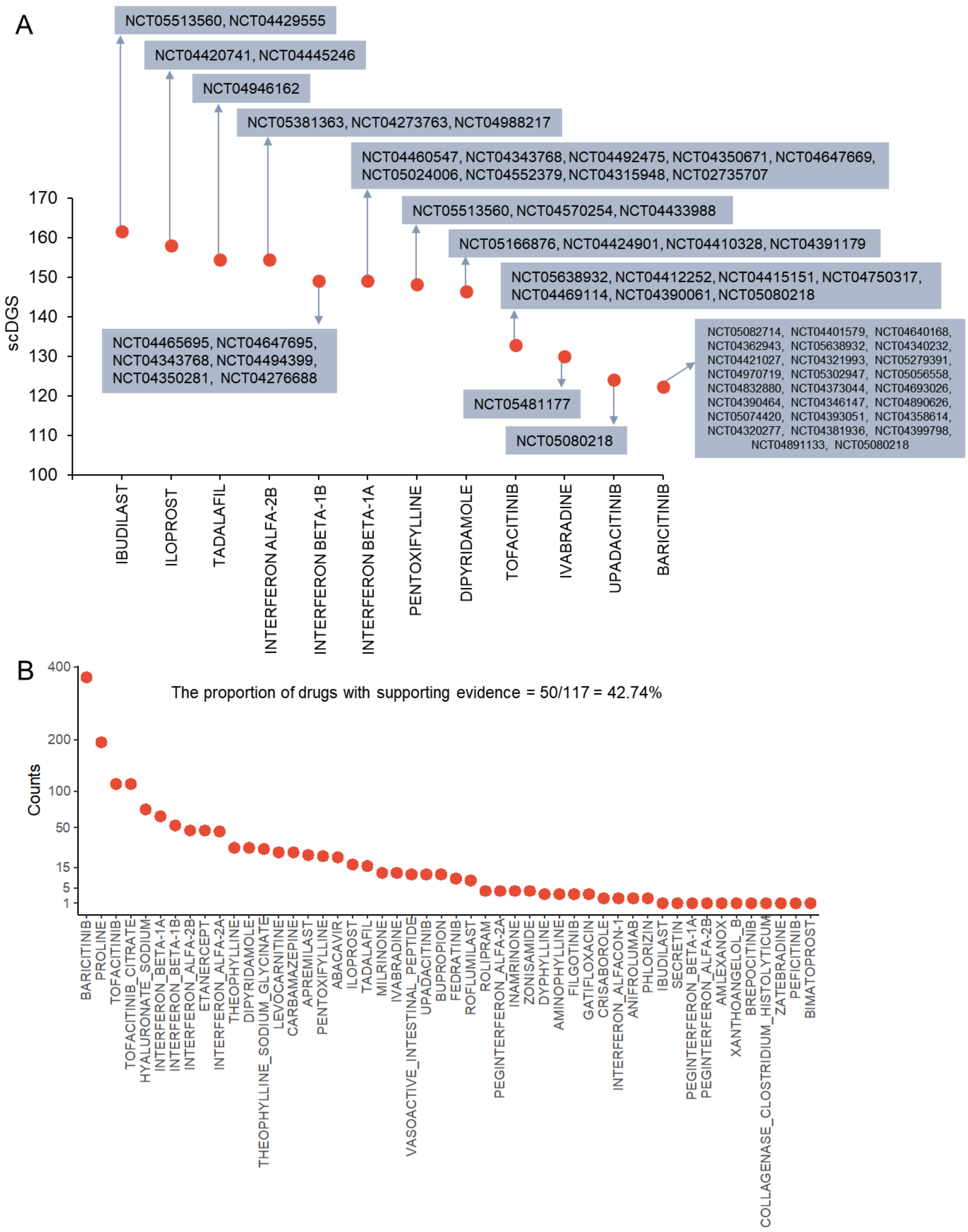


**Supplementary Figure S20. Summary of supporting evidence for scDrugHunter-identified drugs related to treating severe COVID-19 in lung MSCs.** A. Dot plot showing that 12 COVID-19-relevant drugs that have at least one clinical trial identified by the scDrugHunter method. These 12 drugs had been tested in 60 double-blind and placebo-controlled clinical trials for the treatment of COVID-19 (Clinicaltrials.gov). The identifier for each clinical trial (e.g., NCT05513560 for IBUDILAST) is marked in the plot. B. Dot plot showing the results of RISmed to identify supporting evidence reported in the PubMed database for scDrugHunter-identified 117 drugs relevant for treating severe COVID-19. We found 50 drugs with at least one piece of supporting evidence (proportion = 50/117 = 42.74%).

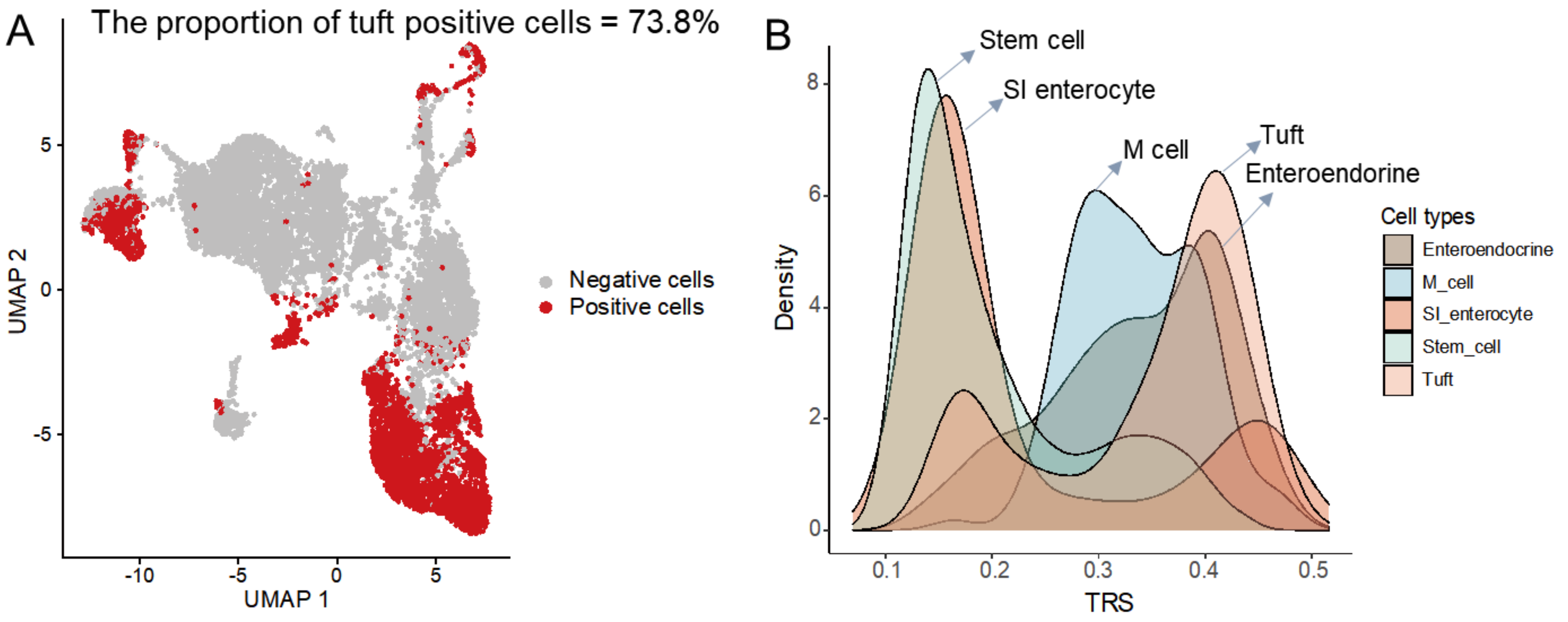


**Supplementary Figure S21. scPagwas-identified positive cells in human intestinal organoids.** A. The UMAP visualization of scPagwas-identified positive cells in human intestine organoids (n = 12,097 cells). The proportion of tuft positive cells is 73.8% (1,599/2,167). Positive cells are shown in red color, and negative cells are shown in gray color. B. Density plots showing the distribution of scPagwas trait-relevant scores (TRSs) for five cell populations in human intestine organoids. Different color represent corresponding cell types, including enteroendocrine (n = 398 cells), membranous cells (M cells, n = 112 cells), small intestine (SI) enterocytes (n = 525 cells), stem cells (n = 8,895 cells), and tuft cells (n = 2,167 cells).

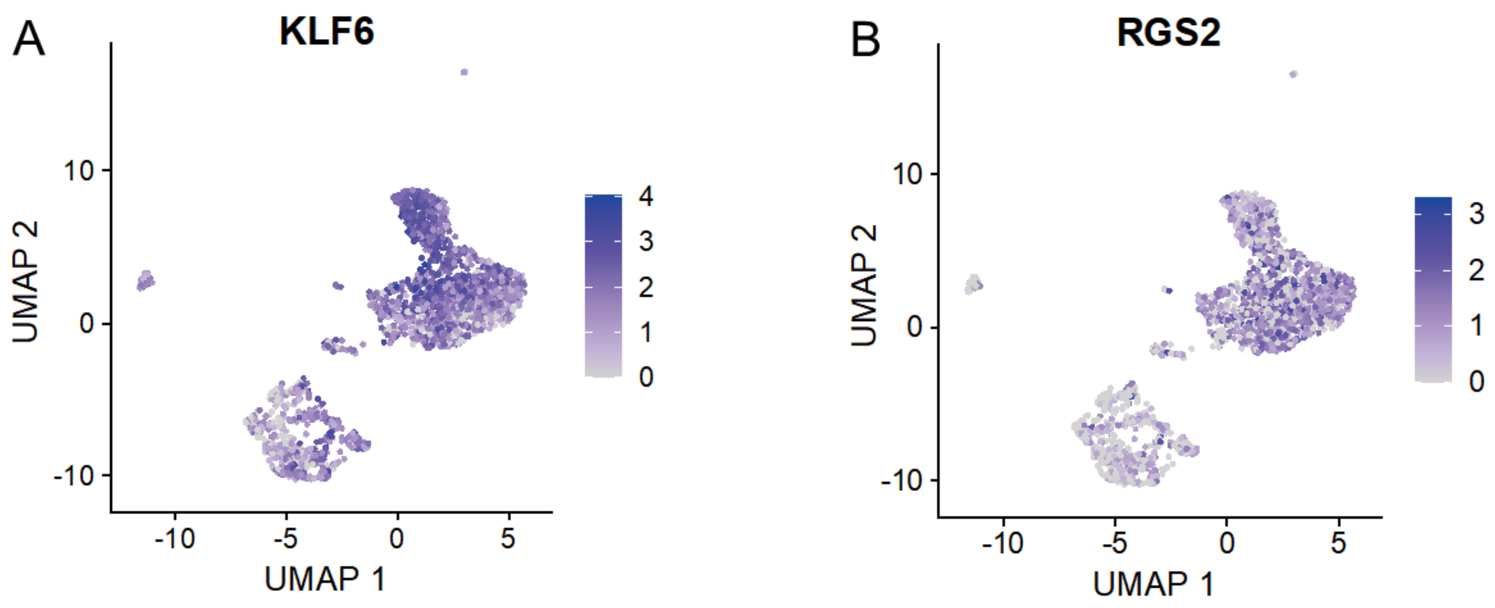


**Supplementary Figure S22. Normalized expression of common markers for human intestinal tuft cells.** In the present investigation, we first applied the scArches method to auto-annotate the cell type of tuft cells (n = 2,167 cells) in human intestine organoids based on the human fetal intestine scRNA-seq data as the reference. Then, to further confirm the annotation of intestinal tuft cells, we used the expressed markers of (A) *KLF6* and (B) *RGS2* for annotation and visualization. These two gene expression markers were based on the single-cell gene expression resource of PanglaoDB (https://www.panglaodb.se/index.html).

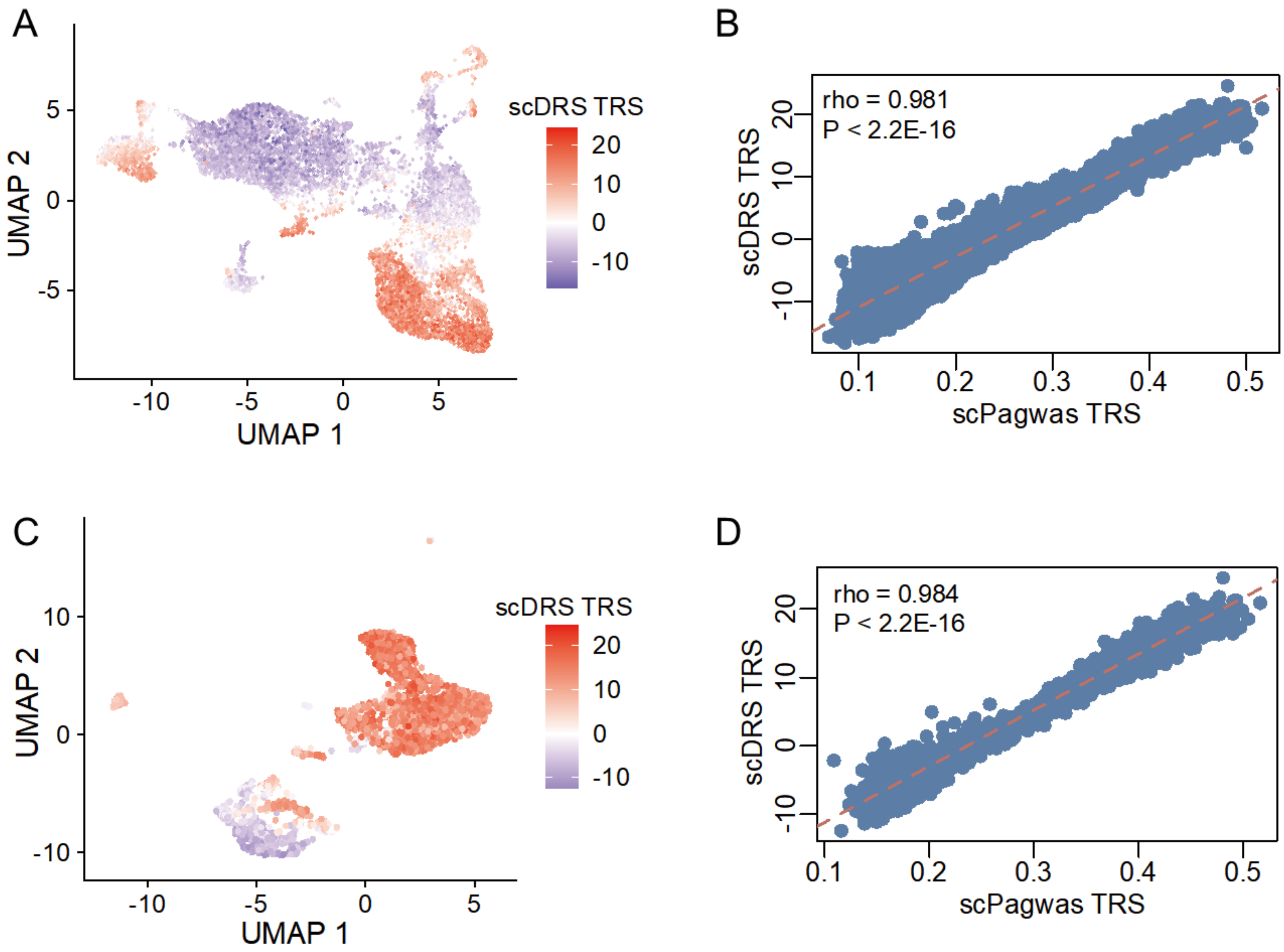


E

scDRS (columns) vs scPagwas (rows)

| Proportion | Negative cells | Positive cells |
|---|---|---|
| Negative cells | 0.901 | 0.0986 |
| Positive cells | 0.0056 | 0.994 |

Concordance rate = (0.901+0.994)/(0.901+0.0986+0.0056+0.994) = 0.948

**Supplementary Figure S23. Concordance analysis of the individual cellular association results between scDRS and scPagwas in intestinal tuft cells**. A. UMAP plot shows that the scDRS TRSs for the phenotype of very severe COVID-19 is shown in intestine organoids scRNA-seq data (n = 12,097 cells). Five main cell types of intestine organoids included tuft cells (n = 2,167 cells), membranous cells (M cells, n = 112 cells), enteroendocrines (n = 398 cells), small intestine (SI) enterocytes (n = 525 cells), and stem cells (n = 8,895 cells; Figure 6A). B. The Spearman correlation analysis of TRSs from between scDRS-based and scPagwas-based method in intestine organoids. C. UMAP visualizes that the scDRS TRSs for the phenotype of very severe COVID-19 is shown in intestinal tuft cells (n = 2,167 cells). D. The Spearman correlation analysis of TRSs from between scDRS-based and scPagwas-based method in intestinal tuft cells. E. E. Concordance assessment of the positive cell results from scDRS and scPagwas in intestinal tuft cells. scDRS estimates the significant level for each cell by using the Monte Carlo (MC) sampling method, and FDR < 0.05 is considered to be statistically significant (see Supplemental Methods for details).

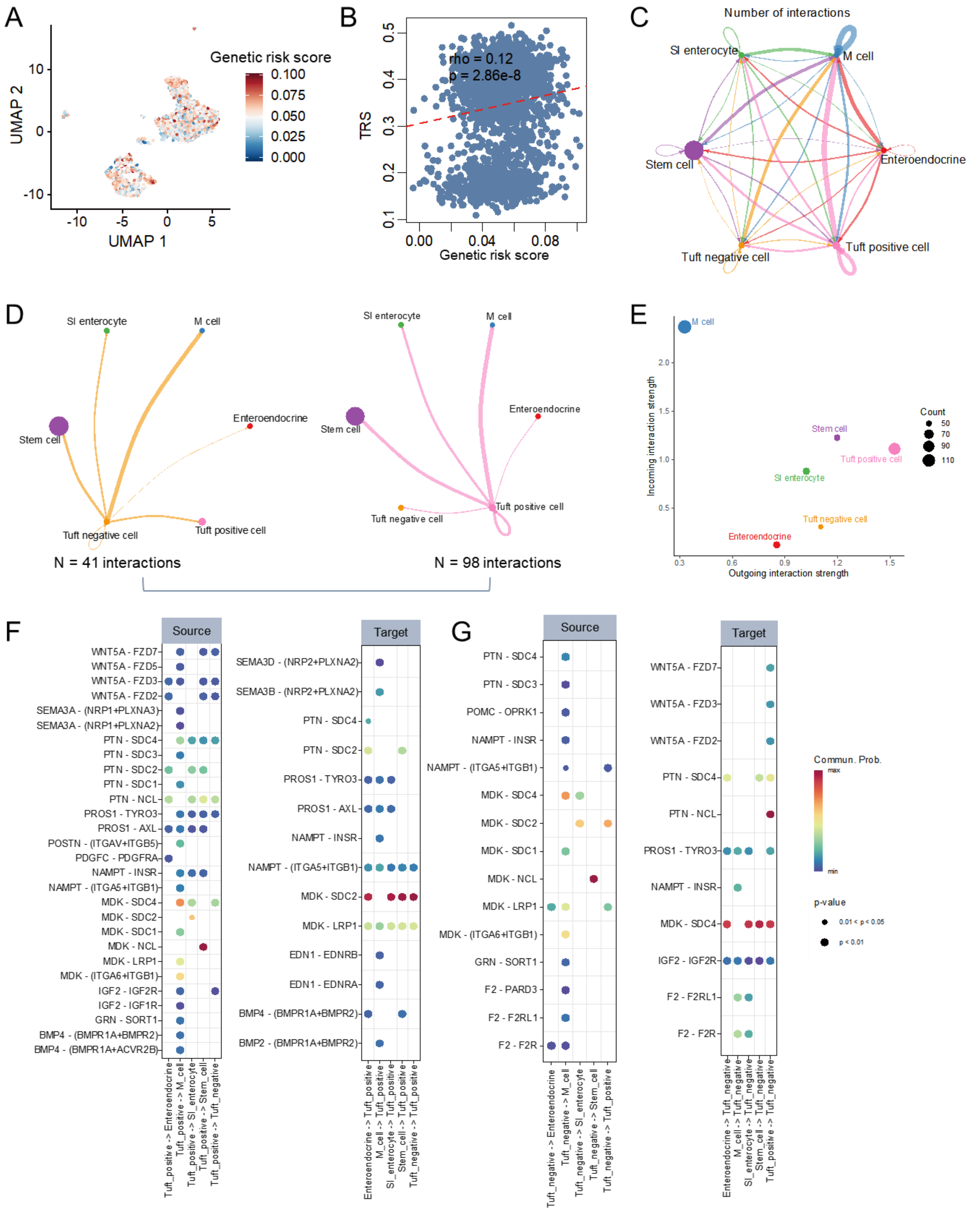


**Supplementary Figure S24. 438 genetic risk gene scores and cellular communications of intestinal tuft positive cells.** A. UMAP embedding of intestinal tuft cells among three cell clusters colored by the genetic risk scores (GRS) of 438 COVID-19-relevant genes. B. The correlation between the genetic risk scores of 438 genes and scPagwas TRSs across all intestinal tuft cells. C. The aggregated cell-to-cell communication network among cell types in human intestine organoids. D. The cellular communications of tuft negative cells (left panel) and tuft positive cells (right panel) with other cell types in human intestine organoids. The width represents the number of cellular interactions, and color represents each cell type. E. Scatter plot showing the dominant senders (sources) and receivers (targets) in a 2D space. y axis represents incoming interaction strength, and x axis represents outgoing interaction strength. The size of each node indicates the count of cellular interactions. F-G) Dot plots showing that predicted cellular interactions of tuft positive cells (F) and tuft negative cells (G) with other cell populations in human intestine organoids. Left panel indicates tuft positive cells (F) and tuft negative cells (G) as a source (outgoing) communicating with other cell types, and right panel indicates tuft positive cells (F) and tuft negative cells (G) as a target (incoming) communicating with other cell types. The circular size represents the statistical significance of each ligand-receptor pair, and color represents the communication probability.

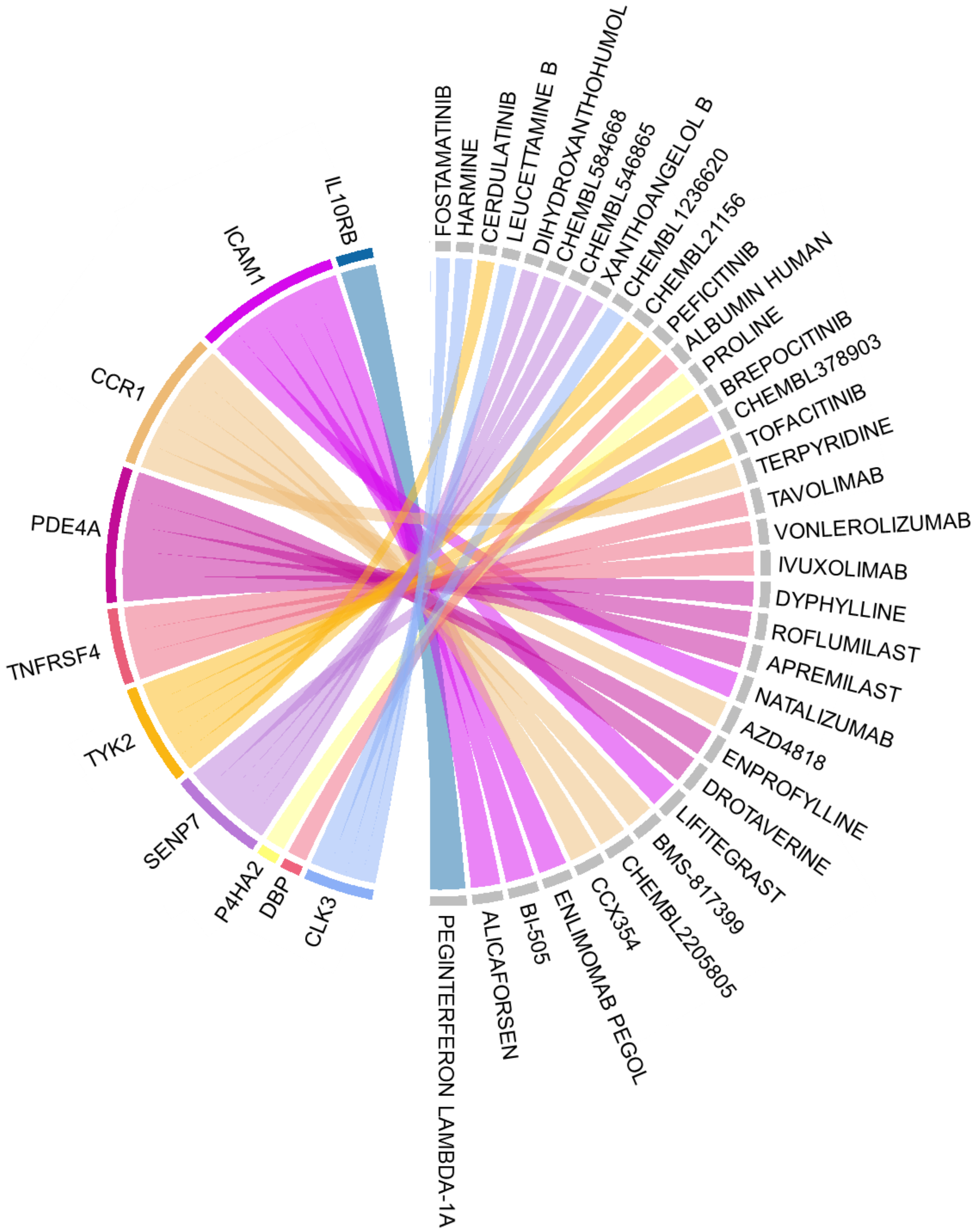


**Supplementary Figure S25. Chord diagram of scDrugHunter-identified top 10 druggable genes and relevant interacting drugs for very severe COVID-19 in intestinal tuft cells**. The width of each line is determined by the number of drugs (n = 1~5 drugs) known to interact with each gene. Genes are ordered by the degree of single-cell druggable score (scDGS) at the top of the diagram. The top-ranked 10 genes contained *IL10RB, ICAM1, CCR1, PDE4A, TNFRSF4, TYK2, SENP7, P4HA2, DBP,* and *CLK3*. The interacting drugs were analyzed based on the DGIdb database (DGIdb v4.2.0, https://www.dgidb.org/). Visualization of the number of interactions between druggable genes relevant to very severe COVID-19 and their interacting drugs was undertaken using the R package *circlize* 0.4.15.

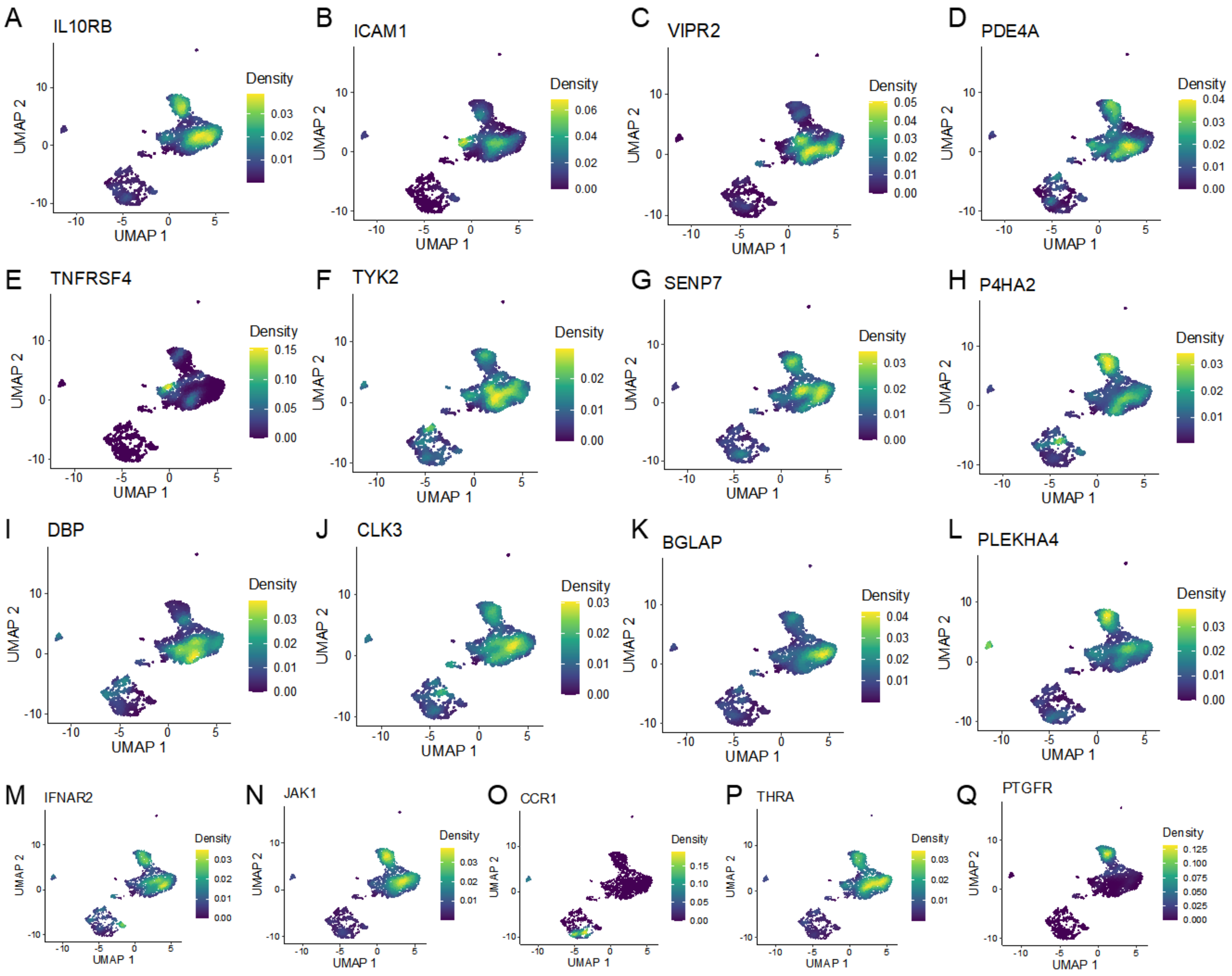


**Supplementary Figure S26. UMAP embedding plot showing the expression density of 17 scDrugHunter-identified druggable genes in intestinal tuft cells**. The colored bar represent the density of normalized expression of these 17 druggable genes with higher scDGSs (>120) identified by scDrugHunter in intestinal tuft cells. A-Q). *IL10RB, ICAM1, VIPR2, PDE4A, TNFRSF4, TYK2, SENP7, P4HA2, DBP, CLK3, BGLAP, PLEKHA4, IFNAR2, JAK1, CCR1, THRA*, and *PTGFR*.

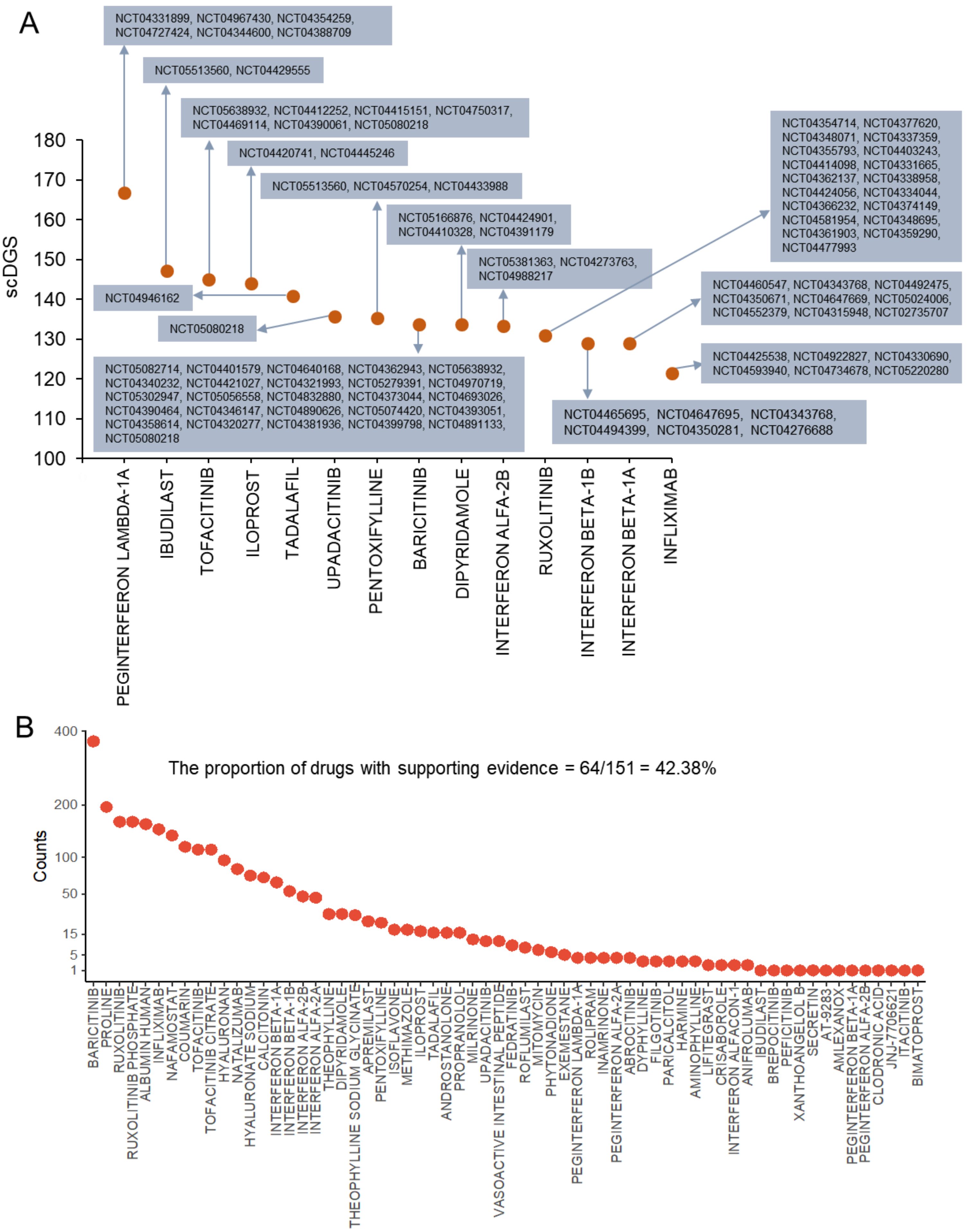


**Supplementary Figure S27. Summary of supporting evidence for scDrugHunter-identified drugs related to treating severe COVID-19 in intestinal tuft cells.** A. Dot plot showing that 14 COVID-19-relevant drugs that have at least one clinical trial identified by the scDrugHunter method. These 14 drugs had been tested in 89 double-blind and placebo-controlled clinical trials for the treatment of COVID-19 (Clinicaltrials.gov). The identifier for each clinical trial (e.g., NCT04331899 for PEGINTERFERON LAMBDA-1A) is marked in the plot. B. Dot plot showing the results of RISmed to identify supporting evidence reported in the PubMed database for scDrugHunter-identified 151 drugs relevant for treating severe COVID-19. We found 64drugs with at least one piece of supporting evidence (proportion = 64/151 = 42.38%).

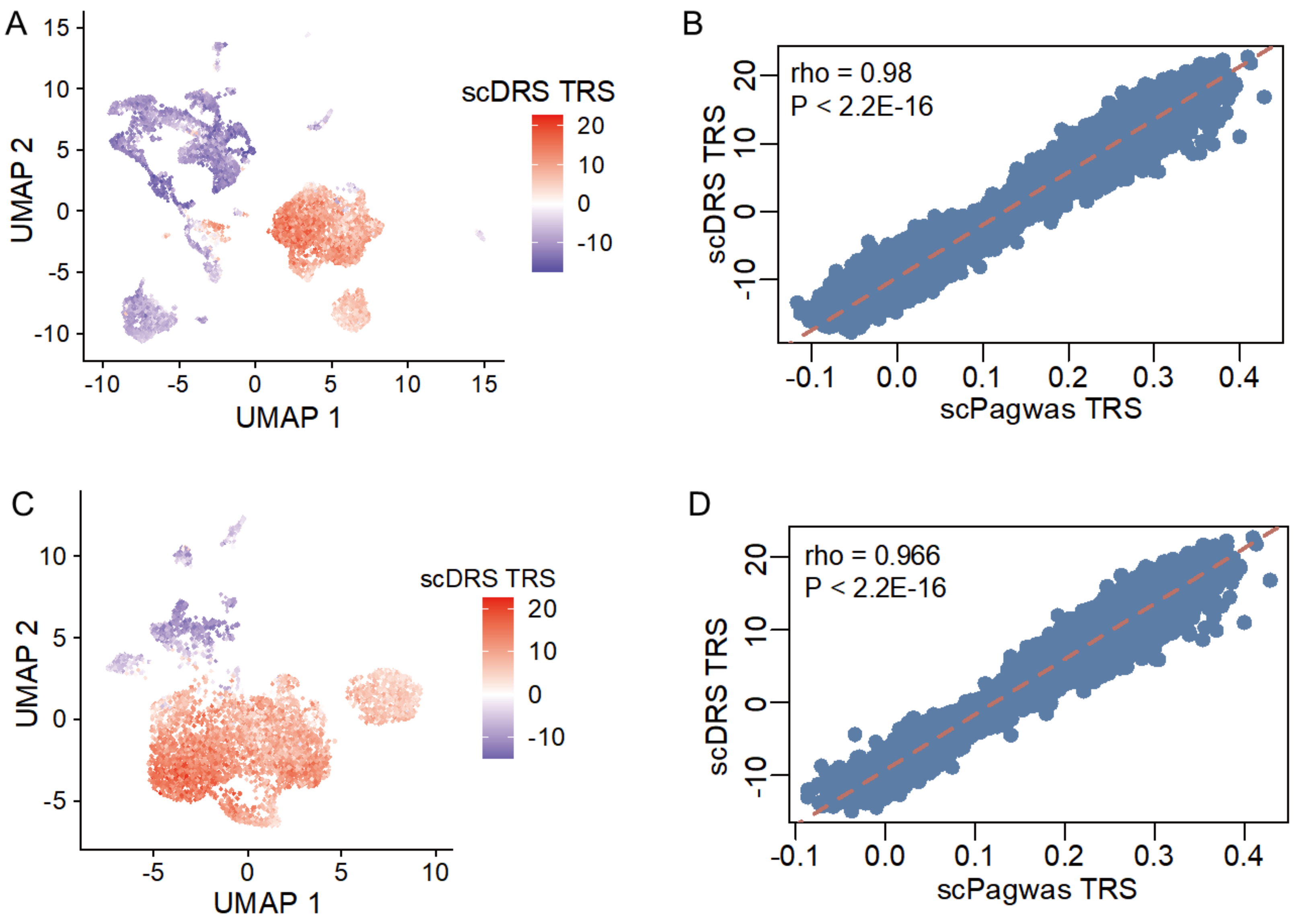


E

| | | scDRS | |
|---|---|---|---|
| | Proportion | Negative cells | Positive cells |
| scPagwas | Negative cells | 0.476 | 0.524 |
| scPagwas | Positive cells | 0.0032 | 0.997 |

Concordance rate = (0.997+0.476)/(0.476+0.524+0.0032+0.997) = 0.74

**Supplementary Figure S28. Concordance analysis of the individual cellular association results between scDRS and scPagwas in brain endothelial cells.** A. UMAP plot shows that the scDRS TRSs for the phenotype of very severe COVID-19 is shown in brain organoids scRNA-seq data (n = 10,677 cells). Eight main cell types in brain organoids included eight cell types of excitatory neuron (n = 194 cells), inhibitory neuron (n = 426 cells), microglia (n = 899 cells), astrocytic cells (n = 1,209 cells), endothelial cells (n = 6,088 cells), neural progenitor cells (NPC, n = 805 cells), intermediate progenitor cells (IPC, n = 871 cells), and radial glia (RG, n = 185 cells, see Figure 7A). B. The Spearman correlation analysis of TRSs from between scDRS-based and scPagwas-based method in brain organoids. C. UMAP plot shows that the scDRS TRSs for the phenotype of very severe COVID-19 is shown in brain endothelial cells (n = 6,088 cells). D. The Spearman correlation analysis of TRSs from between scDRS-based and scPagwas-based method in brain endothelial cells. E. Concordance assessment of the positive cell results from scDRS and scPagwas in brain endothelial cells. scDRS estimates the significant level for each cell by using the Monte Carlo (MC) sampling method, and FDR < 0.05 is considered to be statistically significant (see Supplemental Methods for details).

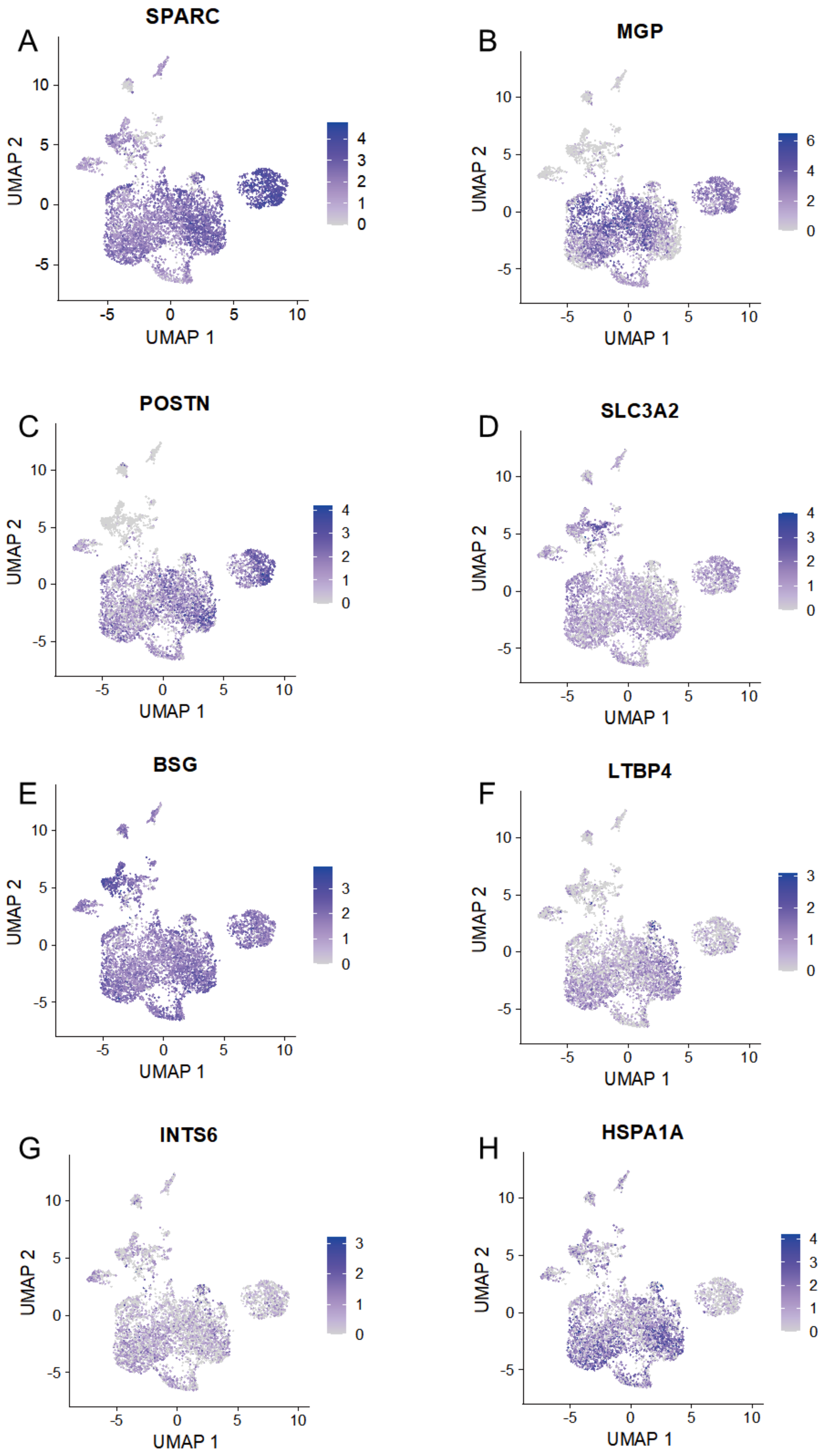


**Supplementary Figure S29. UMAP visualization of normalized expression of common markers for human brain endothelial cells.** In the present investigation, we first applied the scArches method to auto-annotate the cell type of endothelial cells (n = 6,088 cells) in human brain organoids based on the human fetal brain scRNA-seq data as the reference. Then, to further confirm the annotation of brain endothelial cells, we used the expressed markers of (A) *SPARC,* (B) *MGP,* (C) *POSTN,* (D) *SLC3A2,* (E) *BSG,* (F) *LTBP4,* (G) *INTS6*, and (H) *HSPA1A* for annotation and visualization. These gene expression markers were based on the single-cell gene expression resource of PanglaoDB (https://www.panglaodb.se/index.html) and one reported study (Zhong et al. Nature 2020, PMID: 31942070).

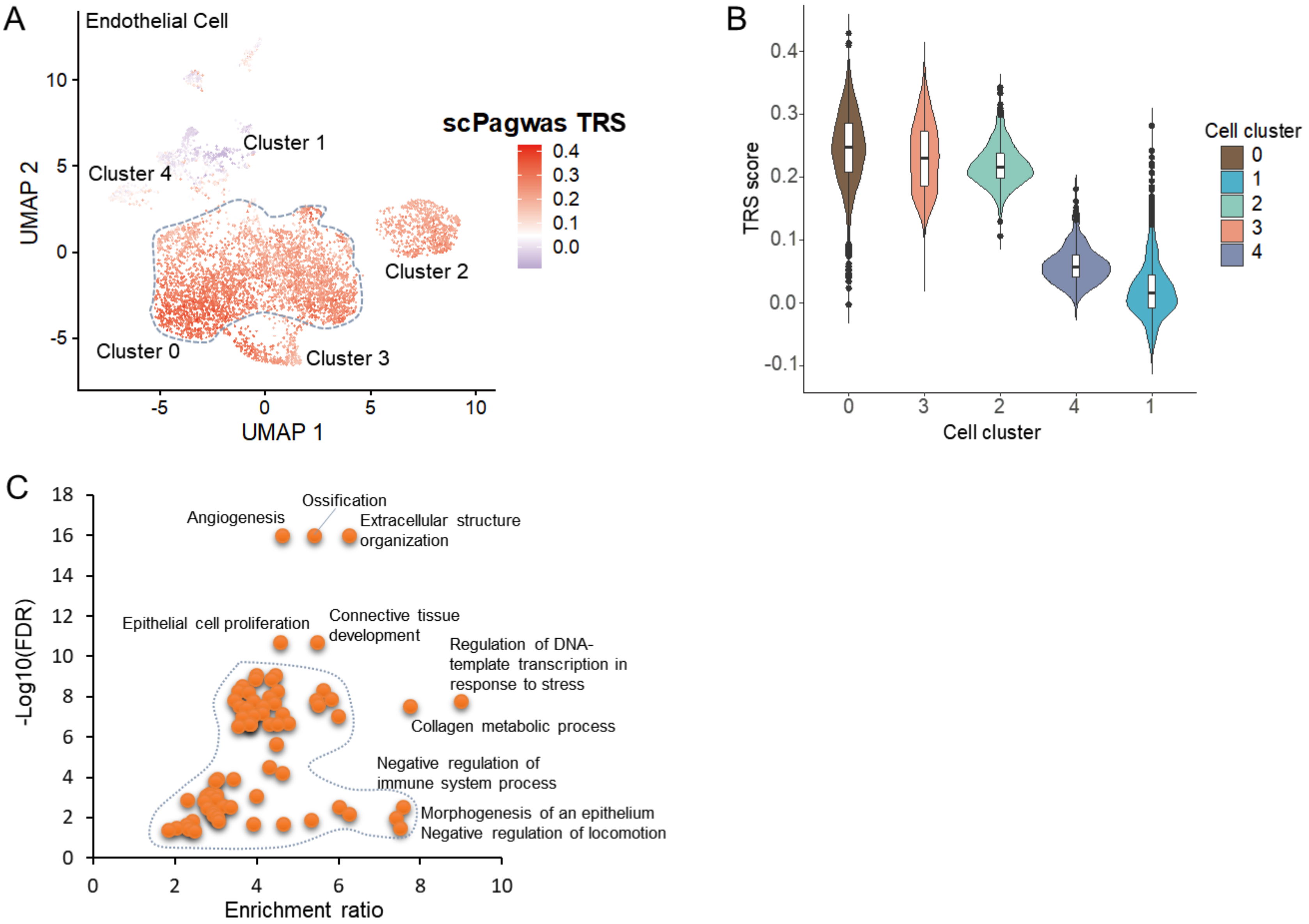


**Supplementary Figure S30. The scPagwas TRS scores and druggable gene scores of five clusters in brain endothelial cells for very severe COVID-19.** A. UMAP plot shows that scPagwas TRSs for the phenotype of very severe COVID-19 is shown in brain endothelial cells. B. Violin plot exhibiting the scPagwas TRS scores in five cell clusters among brain endothelial cells. C. GO-BP-term enrichment analysis on 341 significantly up-regulated genes in endothelial positive cells by using the WebGestalt. The y-axis indicates –Log10(FDR), and the x-axis indicates the enrichment ratio.

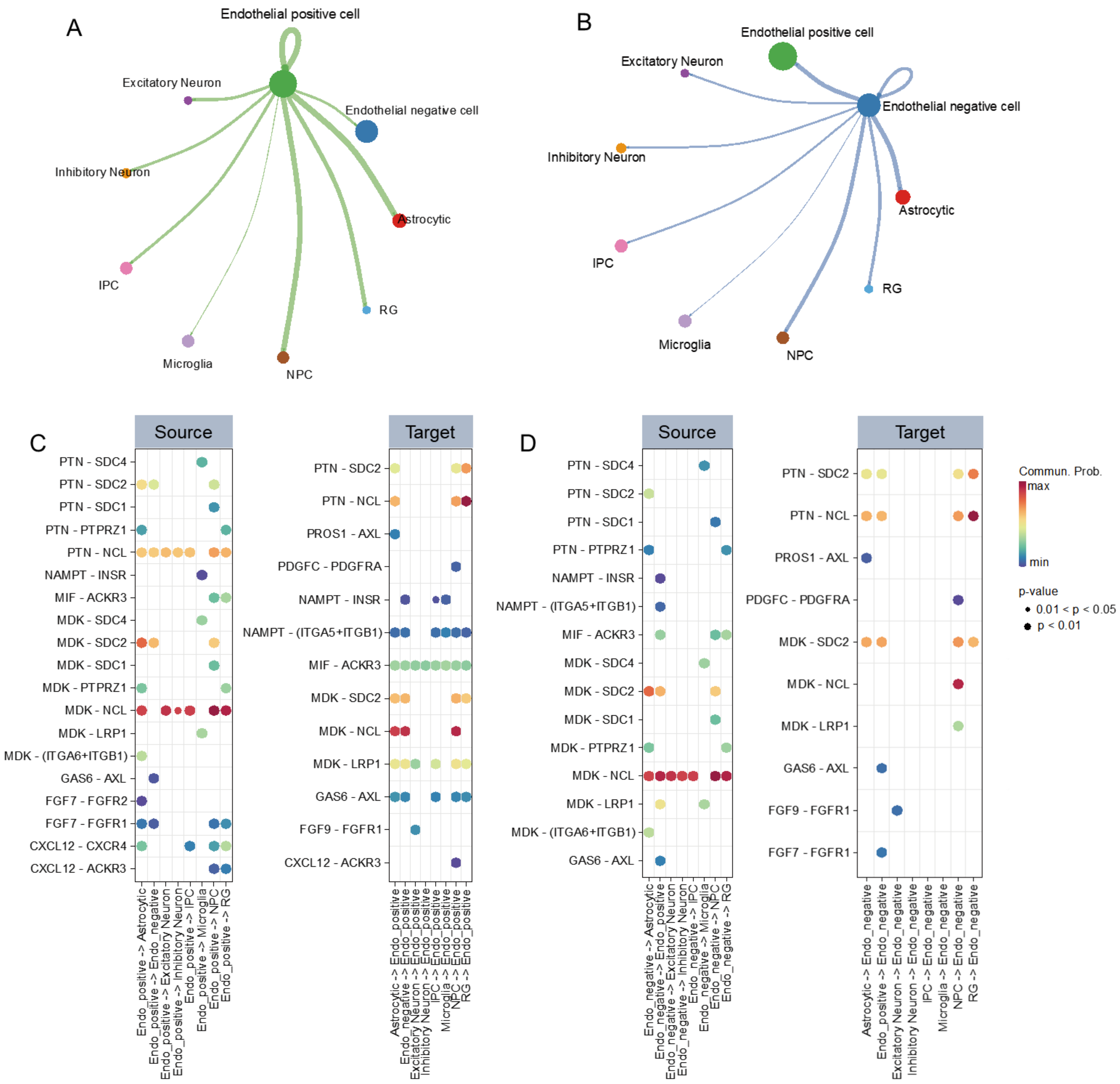


**Supplementary Figure S31. Cellular communications of endothelial positive cells with other cell types.** A. The cellular communications of endothelial positive cells with other cell populations in human brain organoids. B. The cellular communications of endothelial negative cells with other cell populations in human brain organoids. The edge width represents the count of cellular interactions, the color represents different cell types, and the dot size represents the total count of cellular interactions. C. Dot plots showing that predicted cellular interactions of endothelial positive cells with other cell populations in brain organoids. Right panel: endothelial positive cells (endo_positive) as a target (incoming) communicating with other cells; Left panel: endothelial positive cells (endo_positive) as a source (outgoing) communicating with other cells; D. Dot plots showing that predicted cellular interactions of endothelial negative cells with other cell populations in brain organoids. Right panel: endothelial negative cells (endo_negative) as a target (incoming) communicating with other cells; Left panel: endothelial negative cells (endo_negative) as a source (outgoing) communicating with other cells. The dot size represents the statistical significance of each ligand-receptor pair, and color represents the communication probability.

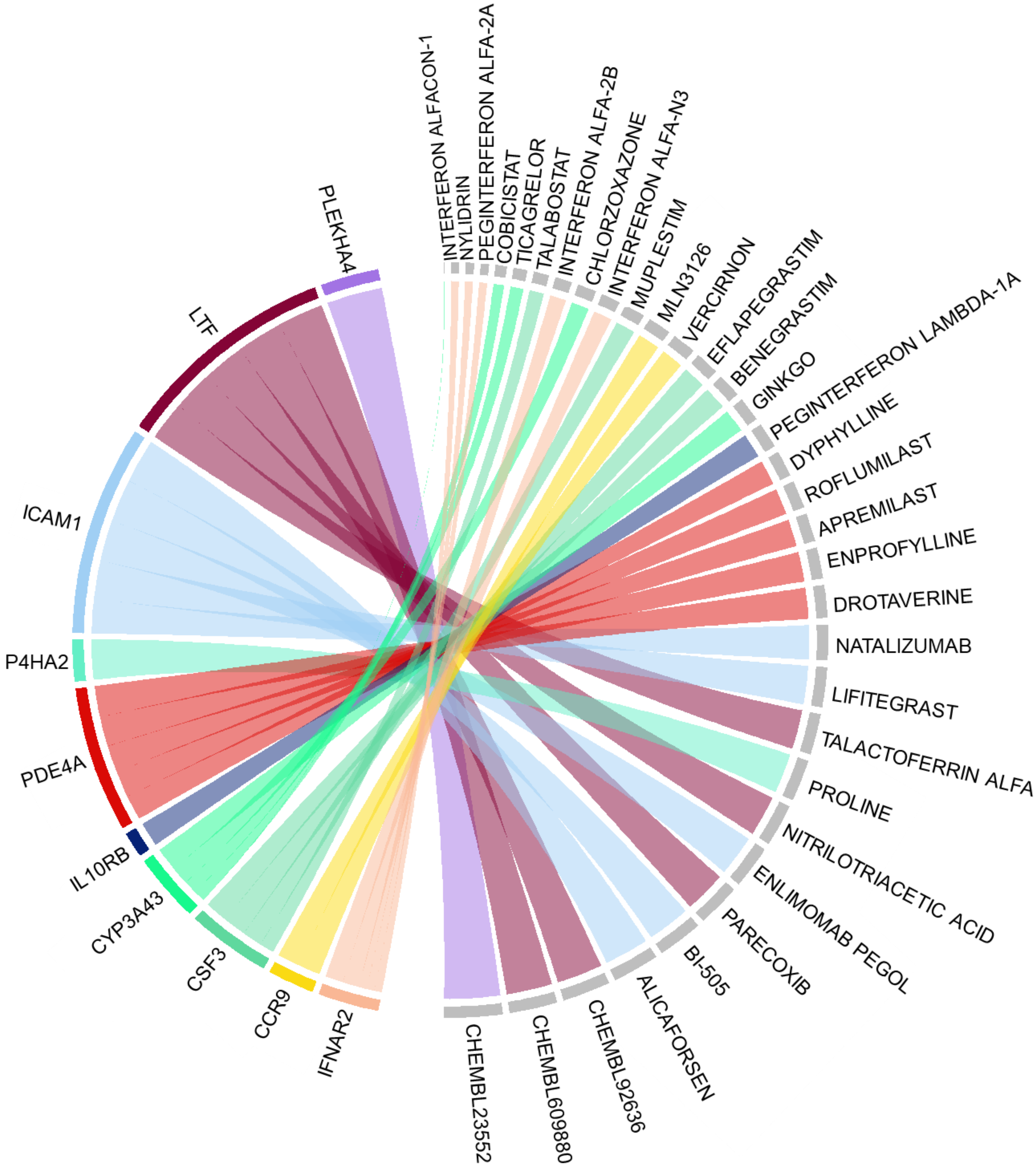


**Supplementary Figure S32. Chord diagram of scDrugHunter-identified top 10 druggable genes and relevant interacting drugs for very severe COVID-19 in intestinal tuft cells.** The width of each line is determined by the number of drugs (n = 1~5 drugs) known to interact with each gene. Genes are ordered by the degree of single-cell druggable score (scDGS) at the top of the diagram. The top-ranked 10 genes contained *PLEKHA4, LTF, ICAM1, P4HA2, PDE4A, IL10RB, CYP3A43, CSF3, CCR9,* and *IFNAR2*. The interacting drugs were analyzed based on the DGIdb database (DGIdb v4.2.0, https://www.dgidb.org/). Visualization of the number of interactions between druggable genes relevant to very severe COVID-19 and their interacting drugs was undertaken using the R package circlize 0.4.15.

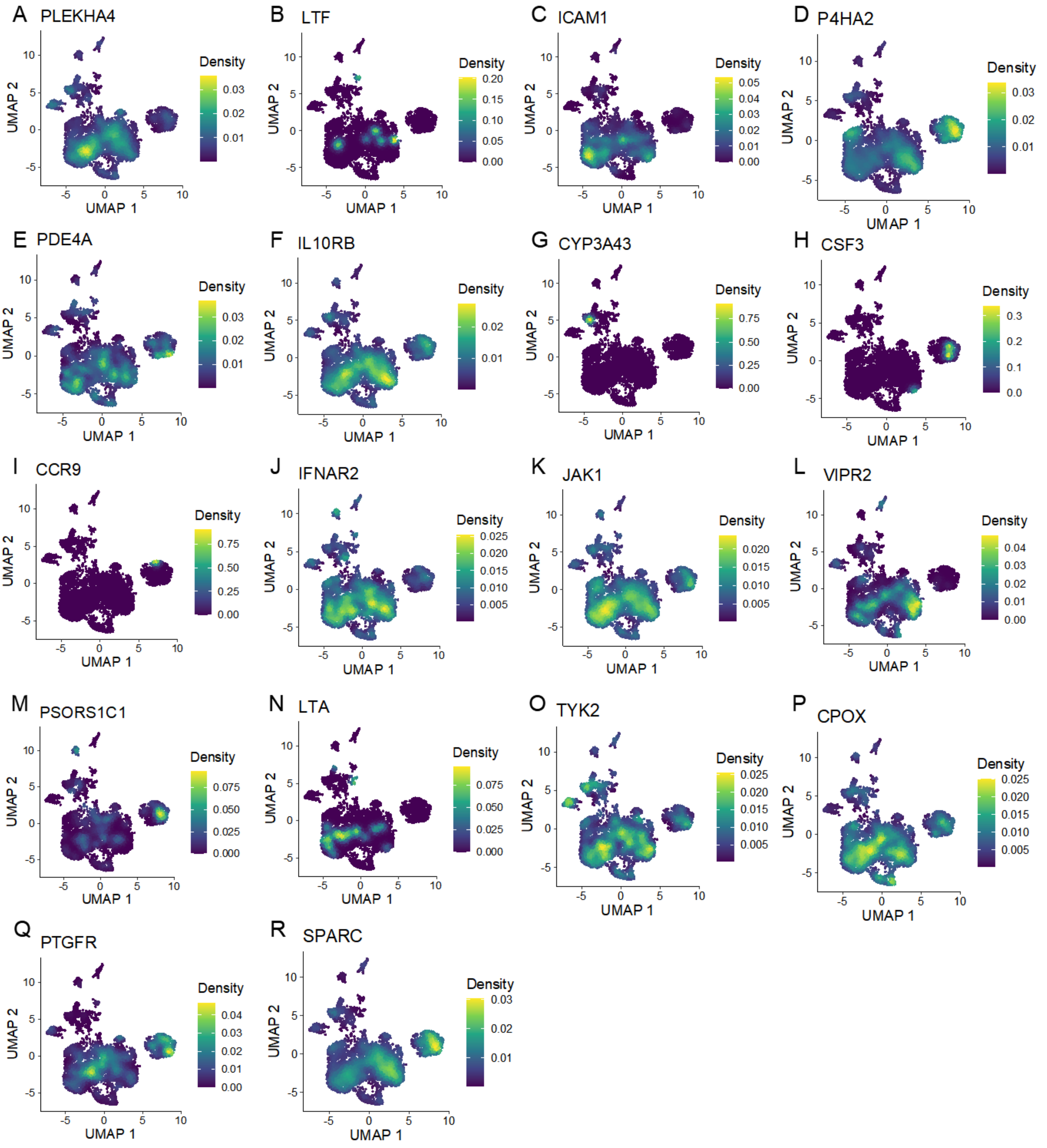


**Supplementary Figure S33. UMAP embedding plot showing the expression density of 18 scDrugHunter-identified druggable genes in brain endothelial cells.** The colored bar represent the density of normalized expression of these 18 druggable genes with higher scDGSs (>120) identified by scDrugHunter in brain endothelia cells. A-Q). *PLEKHA4, LTF, ICAM1, P4HA2, PDE4A, IL10RB, CYP3A43, CSF3, CCR9, IFNAR2, JAK1, VIPR2, PSORS1C1, LTA, TYK2, CPOX, PTGFR*, and *SPARC*.

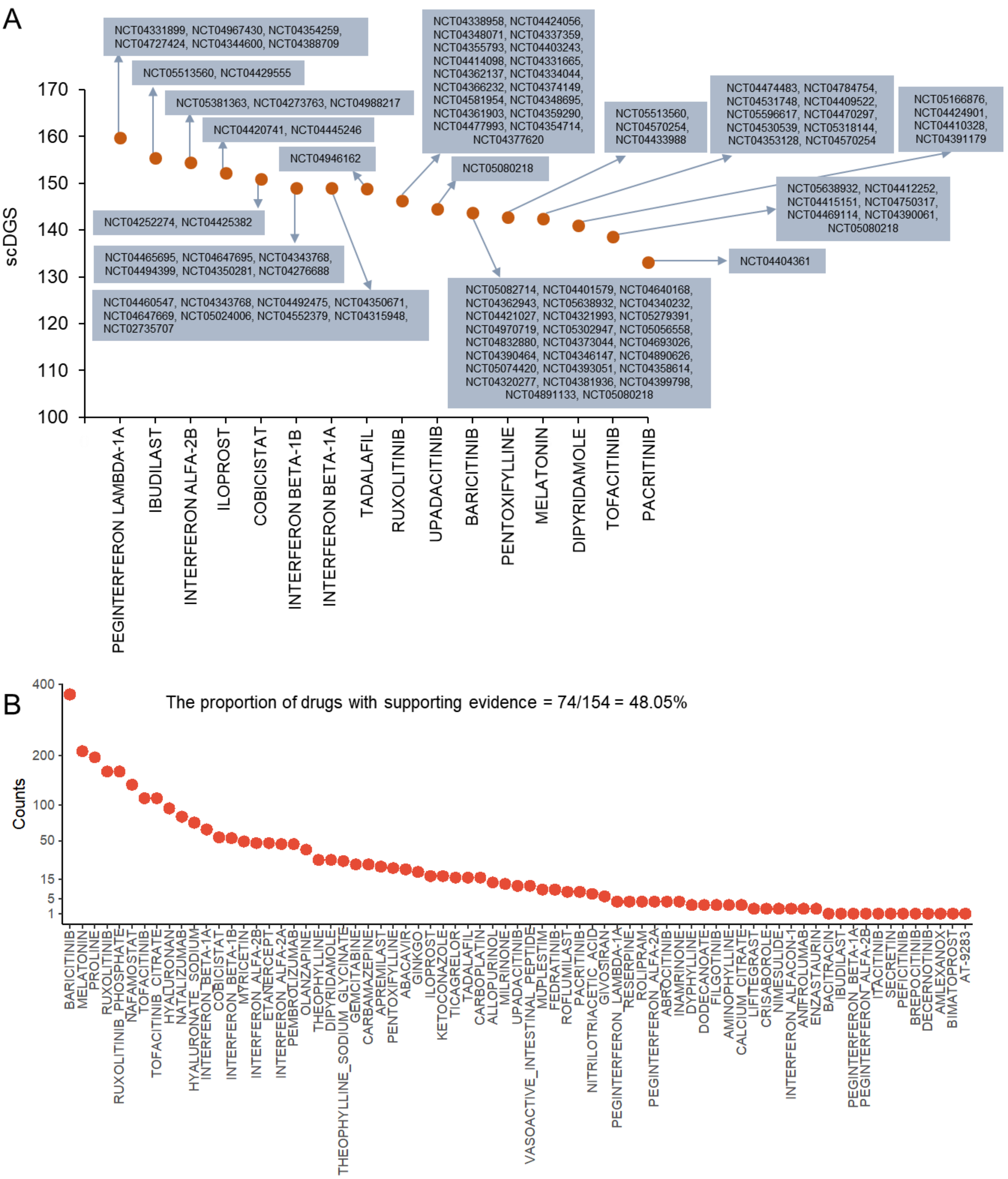


**Supplementary Figure S34. Summary of supporting evidence for scDrugHunter-identified drugs related to treating severe COVID-19 in intestinal tuft cells.** A. Dot plot showing that 14 COVID-19-relevant drugs that have at least one clinical trial identified by the scDrugHunter method. These 14 drugs had been tested in 89 double-blind and placebo-controlled clinical trials for the treatment of COVID-19 (Clinicaltrials.gov). The identifier for each clinical trial (e.g., NCT04331899 for PEGINTERFERON LAMBDA-1A) is marked in the plot. B. Dot plot showing the results of RISmed to identify supporting evidence reported in the PubMed database for scDrugHunter-identified 151 drugs relevant for treating severe COVID-19. We found 64 drugs with at least one piece of supporting evidence (proportion = 74/154 = 48.05%).

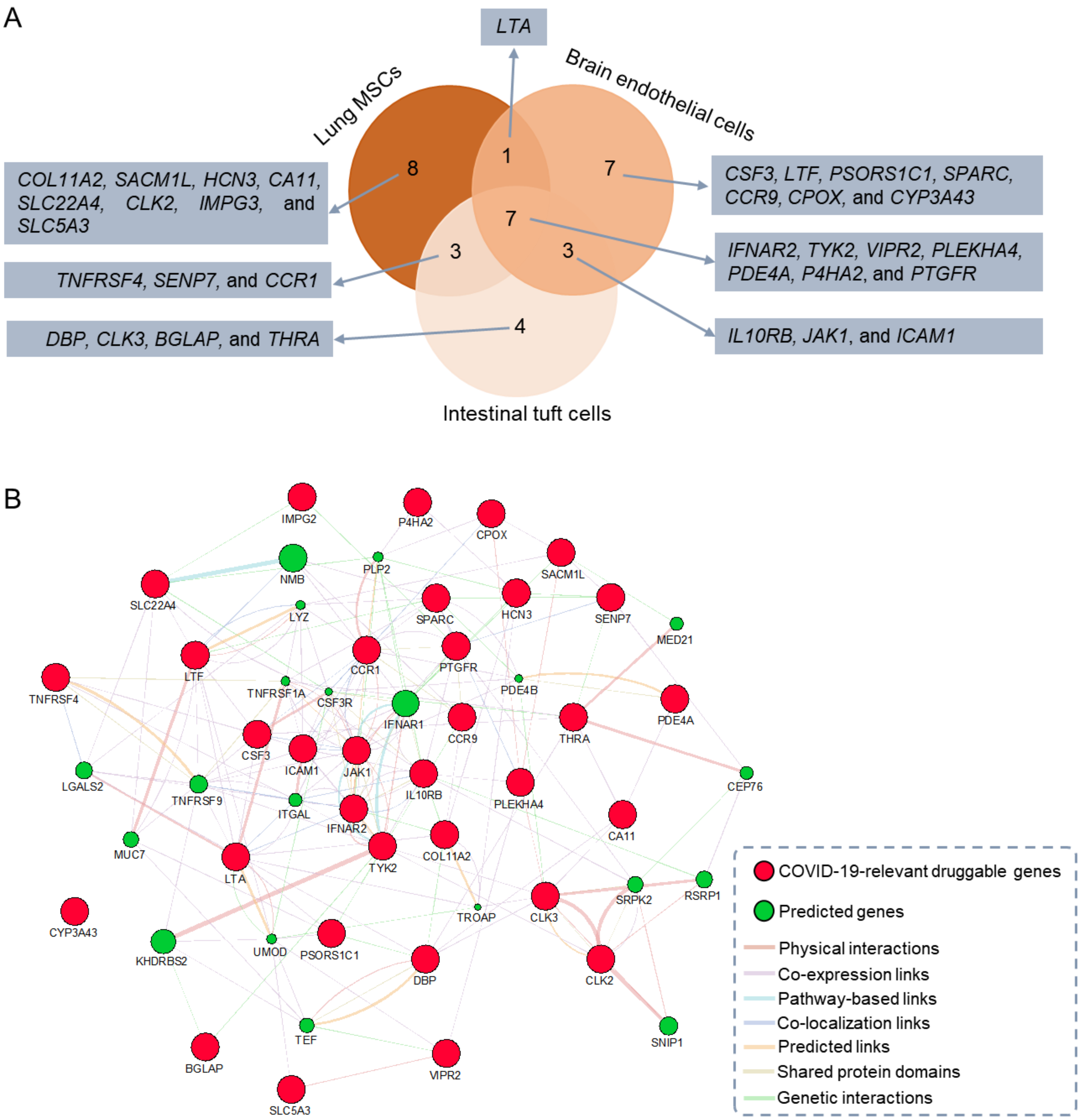


**Supplementary Figure S35. Comparison analysis of scDrugHunter-identified druggable genes across three cell types of lung MSCs, intestinal tuft cells, and brain endothelial cells.** A. The overlapped druggable genes across lung MSCs (n = 19 druggable genes), intestinal tuft cells (n = 17 druggable genes), and brain endothelial cells (n = 18 druggable genes). There were 33 druggable genes in total identified by scDrugHunter in a given cell type based on the DGIdb database (DGIdb v4.2.0, https://www.dgidb.org/). B. Protein-protein interaction network of 33 druggable genes. The 33 druggable genes are shown in red color and the predicted genes are shown in green color. The predicted attributes included physical interactions, co-expression links, pathway-based links, co-localization links, predicted links, shared protein domains, and genetic interactions using the GeneMANIA tool (http://www.genemania.org).
