## Supplementary Tables for "Integration of human organoids single-cell transcriptomic profiles and human genetics repurposes critical cell type-specific drug targets for severe COVID-19"

### Supplementary Table S1. Summary of scRNA-seq datasets on human fetal organs

| Fetal Organs | Samples | Cell Count | Sequencing Platform | Accession No. | Data Source |
|---|---|---|---|---|---|
| Brain | 8 | 46,985 | 10X Genomics | GSE119212 | GEO |
| Kidney | 1 | 4,176 | 10X Genomics | GSE102596 | GEO |
| Liver | 1 | 1,745 | 10X Genomics | E-MTAB-8210 | ArrayExpress |
| Lung | 11 | 35,848 | 10X Genomics | E-MTAB-8221 | ArrayExpress |
| Pancreas | 4 | 9,372 | 10X Genomics | GSE201230 | GEO |
| Intestine | 2 | 7,641 | 10X Genomics | E-MTAB-9363 | ArrayExpress |
| Intestine | 10 | 54,547 | 10X Genomics | E-MTAB-9489 | ArrayExpress |
| Skin | 7 | 63,020 | 10X Genomics | E-MTAB-7407 | ArrayExpress |
| Eye | 4 | 7,902 | 10X Genomics | GSE142526 | GEO |

### Supplementary Table S2. Summary of GWAS summary statistics on three COVID-19 outcomes used in the current study

| Phenotypes | Year | Data ID | Population | Sample size | Number of SNPs | Resources |
|---|---|---|---|---|---|---|
| Very severe COVID-19 | 2022 | A2_ALL_leave_23andme (Round 7) | Primary European | 1,163,698 | 11,732,503 | https://www.covid19hg.org/results/r7/ |
| Hospitalized COVID-19 | 2022 | B2_ALL_leave_23andme (Round 7) | Primary European | 2,401,372 | 12,030,868 | https://www.covid19hg.org/results/r7/ |
| Susceptible COVID-19 | 2022 | C2_ALL_leave_23andme (Round 7) | Primary European | 2,942,817 | 14,335,927 | https://www.covid19hg.org/results/r7/ |
| Anorexia nervosa | 2017 | ieu-a-1186 | European | 14,477 | 10,641,224 | https://gwas.mrcieu.ac.uk/datasets/ieu-a-1186/ |
| Autism spectrum disorder | 2017 | ieu-a-1185 | European | 46,351 | 9,112,386 | https://gwas.mrcieu.ac.uk/datasets/ieu-a-1185/ |
| Bipolar disorder | 2019 | ieu-b-41 | European | 51,710 | 13,413,244 | https://gwas.mrcieu.ac.uk/datasets/ieu-b-41/ |
| Attention deficit hyperactivity disorder | 2017 | ieu-a-1183 | European | 55,374 | 8,047,420 | https://gwas.mrcieu.ac.uk/datasets/ieu-a-1183/ |
| Major Depression Disease | 2018 | ukb-b-12064 | European | 462,933 | 9,851,867 | https://gwas.mrcieu.ac.uk/datasets/ukb-b-12064/ |
| Schizophrenia | 2020 | PGC | European | 161,405 | 7,585,078 | https://www.medrxiv.org/content/10.1101/2020.09.12.20192922v1.full.pdf |
| Insomnia | 2019 | NA | European | 386,533 | 1,145,847 | https://ctg.cncr.nl/software/summary_statistics |
| Multiple sclerosis | 2019 | ieu-b-18 | European | 115,803 | 6,304,359 | https://gwas.mrcieu.ac.uk/datasets/ieu-b-18/ |
| Migraine | 2018 | ukb-b-16868 | European | 462,933 | 9,851,867 | https://gwas.mrcieu.ac.uk/datasets/ukb-b-16868/ |
| Ischemic stroke | 2018 | ebi-a-GCST006908 | European | 440,328 | 8,296,492 | https://gwas.mrcieu.ac.uk/datasets/ebi-a-GCST006908/ |
| Epilepsy | 2018 | ieu-b-8 | European | 44,889 | 4,880,492 | https://gwas.mrcieu.ac.uk/datasets/ieu-b-8/ |
| Focal epilepsy | 2018 | ieu-b-10 | European | 39,348 | 4,862,782 | https://gwas.mrcieu.ac.uk/datasets/ieu-b-10/ |
| Generalized epilepsy | 2018 | ieu-b-9 | European | 33,446 | 4,867,068 | https://gwas.mrcieu.ac.uk/datasets/ieu-b-9/ |
| Juvenile myoclonic epilepsy | 2018 | ieu-b-17 | European | 30,858 | 4,983,225 | https://gwas.mrcieu.ac.uk/datasets/ieu-b-17/ |
| Alzheimer's disease | 2019 | ieu-b-2 | European | 63,926 | 10,528,610 | https://gwas.mrcieu.ac.uk/datasets/ieu-b-2/ |
| Parkinson's disease | 2019 | ieu-b-7 | European | 482,730 | 17,891,936 | https://gwas.mrcieu.ac.uk/datasets/ieu-b-7/ |
| Snoring | 2019 | NA | European | 359,916 | 1,146,184 | https://ctg.cncr.nl/software/summary_statistics |
| Daytime Napping | 2019 | NA | European | 386,577 | 1,160,017 | https://ctg.cncr.nl/software/summary_statistics |

| | | | | | | |
|---|---|---|---|---|---|---|
| Happiness | 2018 | ukb-b-4062 | European | 152,348 | 9,851,867 | https://gwas.mrcieu.ac.uk/datasets/ukb-b-4062/ |
| Cigarettes smoked per day | 2019 | ieu-b-142 | European | 249,752 | 12,003,613 | https://gwas.mrcieu.ac.uk/datasets/ieu-b-142/ |
| Extraversion | 2012 | ieu-a-115 | European | 17,375 | 2,305,823 | https://gwas.mrcieu.ac.uk/files/ieu-a-115/ieu-a-115.vcf.gz |
| Subjective well-being | 2016 | ebi-a-GCST003766 | European | 298,420 | 2,058,391 | https://gwas.mrcieu.ac.uk/datasets/ebi-a-GCST003766/ |
| Conscientiousness | 2010 | ebi-a-GCST006326 | European | 17,375 | 2,300,083 | https://gwas.mrcieu.ac.uk/files/ebi-a-GCST006326/ebi-a-GCST006326.vcf.gz |
| Openness to experience | 2012 | ieu-a-117 | European | 17,375 | 2,305,641 | https://gwas.mrcieu.ac.uk/datasets/ieu-a-117/ |
| Neuroticism | 2017 | ebi-a-GCST005232 | European | 329,821 | 18,436,568 | https://gwas.mrcieu.ac.uk/datasets/ebi-a-GCST005232/ |
| Cognitive Performance | 2018 | ebi-a-GCST006572 | European | 257,841 | 10,066,414 | https://gwas.mrcieu.ac.uk/datasets/ebi-a-GCST006572/ |
| Smoking cesstion | 2019 | bbj-a-81 | European | 76,047 | 5,961,480 | https://gwas.mrcieu.ac.uk/datasets/bbj-a-81/ |
| Age of smoking initiation | 2019 | ieu-b-24 | European | 341,427 | 11,894,779 | https://gwas.mrcieu.ac.uk/datasets/ieu-b-24/ |
| Hypertension | 2018 | ukb-b-14057 | European | 462,933 | 9,851,867 | https://gwas.mrcieu.ac.uk/datasets/ukb-b-14057/ |
| Systolic blood pressure | 2019 | bbj-a-46 | European | 757,601 | 7,088,083 | https://gwas.mrcieu.ac.uk/datasets/bbj-a-46/ |
| Pulse pressure | 2019 | bbj-a-46 | East Asian | 136,249 | 6,108,953 | https://gwas.mrcieu.ac.uk/datasets/bbj-a-46/ |
| High blood pressure | 2018 | ukb-b-14177 | European | 124,227 | 9,851,867 | https://gwas.mrcieu.ac.uk/datasets/ukb-b-14177/ |
| Diastolic blood pressure | 2018 | ieu-b-39 | European | 757,601 | 7,160,619 | https://gwas.mrcieu.ac.uk/datasets/ieu-b-39/ |
| Diastolic blood pressure | 2019 | bbj-a-17 | East Asian | 136,615 | 6,108,953 | https://gwas.mrcieu.ac.uk/datasets/bbj-a-17/ |
| Diastolic blood pressure | 2019 | ebi-a-GCST008029 | Hispanic | 21,549 | 31,859,670 | https://gwas.mrcieu.ac.uk/datasets/ebi-a-GCST008029/ |
| Coronary artery disease | 2017 | ebi-a-GCST005195 | European | 547,261 | 7,934,254 | https://gwas.mrcieu.ac.uk/datasets/ebi-a-GCST005195/ |
| Type II diabetes | 2018 | ebi-a-GCST006867 | European | 655,666 | 5,030,727 | https://gwas.mrcieu.ac.uk/datasets/ebi-a-GCST006867/ |
| Ulcerative Colitis | 2017 | ebi-a-GCST004133 | Mixed | 45,975 | 9,474,559 | https://gwas.mrcieu.ac.uk/datasets/ebi-a-GCST004133/ |
| Psoriasis | 2012 | ebi-a-GCST005527 | European | 33,394 | 138,661 | https://gwas.mrcieu.ac.uk/datasets/ebi-a-GCST005527/ |
| Coeliac disease | 2010 | ebi-a-GCST000612 | European | 15,283 | 528,969 | https://gwas.mrcieu.ac.uk/datasets/ebi-a-GCST000612/ |
| Primary biliary cholangitis | 2015 | ebi-a-GCST003129 | European | 13,239 | 1,124,241 | https://gwas.mrcieu.ac.uk/datasets/ebi-a-GCST003129/ |
| Rheumatoid arthritis | 2014 | ieu-a-833 | Mixed | 80,799 | 9,739,304 | https://gwas.mrcieu.ac.uk/datasets/ieu-a-833/ |
| Inflammatory bowel disease | 2017 | ebi-a-GCST004131 | Mixed | 59,957 | 9,619,016 | https://gwas.mrcieu.ac.uk/datasets/ebi-a-GCST004131/ |
| Systemic lupus erythematosus | 2015 | ebi-a-GCST003156 | European | 14,267 | 7,071,163 | https://gwas.mrcieu.ac.uk/datasets/ebi-a-GCST003156/ |
| Waist Circumference | 2015 | ieu-a-60 | European | 224,459 | 2,566,630 | https://gwas.mrcieu.ac.uk/datasets/ieu-a-60/ |
| Total cholesterol | 2013 | ieu-a-301 | European | 187,365 | 2,446,982 | https://gwas.mrcieu.ac.uk/datasets/ieu-a-301/ |
| Chronic kidney disease | 2016 | ebi-a-GCST003374 | European | 117,165 | 2,179,497 | https://gwas.mrcieu.ac.uk/datasets/ebi-a-GCST003374/ |
| Body fat percentage | 2018 | ukb-b-8909 | European | 454,633 | 9,851,867 | https://gwas.mrcieu.ac.uk/datasets/ukb-b-8909/ |

| | | | | | | |
|---|---|---|---|---|---|---|
| Triglyceride level | 2017 | ebi-a-GCST005073 | European | 9,745 | 23,163,055 | https://gwas.mrcieu.ac.uk/datasets/ebi-a-GCST005073/ |
| High-density lipoprotein (HDL) cholesterol | 2013 | ieu-a-299 | European | 187,167 | 2,447,442 | https://gwas.mrcieu.ac.uk/datasets/ieu-a-299/ |
| Low-density lipoprotein (LDL) cholesterol | 2013 | ieu-a-300 | European | 173,082 | 2,437,752 | https://gwas.mrcieu.ac.uk/datasets/ieu-a-300/ |
| Tuberculosis | 2018 | ukb-b-15622 | European | 462,933 | 9,851,867 | https://gwas.mrcieu.ac.uk/datasets/ukb-b-15622/ |
| Adult asthma | 2018 | ukb-b-18113 | European | 462,933 | 9,851,867 | https://gwas.mrcieu.ac.uk/datasets/ukb-b-18113/ |
| Small cell lung cancer | 2022 | finngen_R8_C3_SCLC_EXALLC | European | 260,160 | 20,166,402 | https://www.finngen.fi/en/access_results |
| Pulmonary embolism | 2018 | ukb-b-16048 | European | 462,933 | 9,851,867 | https://gwas.mrcieu.ac.uk/datasets/ukb-b-16048/ |
| Pulmonary artery | 2018 | ukb-b-18616 | European | 463,010 | 9,851,867 | https://gwas.mrcieu.ac.uk/datasets/ukb-b-18616/ |
| Non-small cell lung cancer, squamous | 2022 | finngen_R8_C3_NSCLC_SQUAM | European | 260,793 | 20,166,432 | https://www.finngen.fi/en/access_results |
| Non-small cell lung cancer, adenocarcinoma | 2022 | finngen_R8_C3_NSCLC_ADENO | European | 260,820 | 20,166,443 | https://www.finngen.fi/en/access_results |
| Non-small cell lung cancer | 2022 | finngen_R8_C3_LUNG_NONSMALL | European | 263,448 | 20,166,592 | https://www.finngen.fi/en/access_results |
| Lung volume | 2019 | ebi-a-GCST90016668 | European | 32,860 | 9,275,407 | https://gwas.mrcieu.ac.uk/datasets/ebi-a-GCST90016668/ |
| Lung function (FVC) | 2019 | ebi-a-GCST007429 | European | 321,047 | 19,676,344 | https://gwas.mrcieu.ac.uk/datasets/ebi-a-GCST007429/ |
| Influenza without pneumonia | 2021 | finn-b-INFLUENZA | European | 291,090 | 16,380,370 | https://gwas.mrcieu.ac.uk/datasets/finn-b-INFLUENZA/ |
| Influenza with pneumonia | 2021 | finn-b-J10_INFLUPNEU | European | 342,499 | 16,380,466 | https://gwas.mrcieu.ac.uk/datasets/finn-b-J10_INFLUPNEU/ |
| All influenza | 2021 | finn-b-J10_INFLUENZA | European | 193,130 | 16,380,378 | https://gwas.mrcieu.ac.uk/datasets/finn-b-J10_INFLUENZA/ |
| Emphysema/chronic bronchitis | 2018 | ukb-b-7280 | European | 462,933 | 9,851,867 | https://gwas.mrcieu.ac.uk/datasets/ukb-b-7280/ |
| Chronic obstructive pulmonary disease | 2018 | ukb-b-16751 | European | 463,010 | 9,851,867 | https://gwas.mrcieu.ac.uk/datasets/ukb-b-16751/ |

### Supplementary Table S3. The percent of significant cell types and significant number of datasets for three COVID-19 outcomes using scPagwas

| Human organoids | Cell types | Very Severe COVID-19 | | Hospitalized COVID-19 | | Susceptible Covid-19 | |
|---|---|---|---|---|---|---|---|
| | | Significant Percent | Significant datasets | Significant Percent | Significant datasets | Significant Percent | Significant datasets |
| Brain | Endothelial Cell | 54.55% | 12 | 22.73% | 5 | 54.55% | 12 |
| Brain | Microglia | 40.00% | 2 | 40.00% | 2 | 60.00% | 3 |
| Brain | Radial Glia | 34.78% | 8 | 4.35% | 1 | 4.35% | 1 |
| Brain | Astrocytic | 26.09% | 6 | 4.35% | 1 | 0.00% | 0 |
| Brain | IPC | 26.09% | 6 | 65.22% | 15 | 26.09% | 6 |
| Brain | Excitatory Neuron | 25.00% | 6 | 8.33% | 2 | 20.83% | 5 |
| Brain | Oligodendrocyte | 20.00% | 1 | 0.00% | 1 | 0.00% | 1 |
| Brain | Inhibitory Neuron | 8.33% | 2 | 12.50% | 3 | 66.67% | 16 |
| Brain | NPC | 0.00% | 0 | 13.64% | 3 | 9.09% | 2 |
| Brain | OPC | 0.00% | 0 | 0.00% | 0 | 100.00% | 1 |
| Lung | MSC | 50.00% | 1 | 50.00% | 1 | 50.00% | 1 |

| | | | | | | | |
|---|---|---|---|---|---|---|---|
| Intestine | Membranous Cell | 63.64% | 7 | 45.45% | 5 | 54.55% | 6 |
| Intestine | Enterocyte | 50.00% | 8 | 31.25% | 5 | 43.75% | 7 |
| Intestine | Tuft | 50.00% | 6 | 50.00% | 6 | 16.67% | 2 |
| Intestine | Stem Cell | 17.65% | 3 | 52.94% | 9 | 41.18% | 7 |
| Intestine | Goblet | 14.29% | 1 | 14.29% | 1 | 0.00% | 0 |
| Intestine | Enteroendocrine | 7.69% | 1 | 15.38% | 2 | 23.08% | 3 |
| Kidney | NPC | 70.00% | 7 | 50.00% | 5 | 50.00% | 5 |
| Kidney | DN | 55.56% | 5 | 0.00% | 0 | 11.11% | 1 |
| Eye | Horizontal | 75.00% | 3 | 25.00% | 1 | 25.00% | 1 |
| Eye | Rod | 75.00% | 3 | 25.00% | 1 | 25.00% | 1 |
| Eye | RPC | 50.00% | 2 | 50.00% | 2 | 25.00% | 1 |
| Eye | RPE | 50.00% | 1 | 0.00% | 0 | 0.00% | 0 |
| Eye | Astrocytic | 33.33% | 1 | 0.00% | 0 | 0.00% | 0 |
| Eye | Muller Glia | 25.00% | 1 | 25.00% | 1 | 25.00% | 1 |
| Eye | Cone | 25.00% | 1 | 25.00% | 1 | 25.00% | 1 |
| Eye | Bipolar Cell | 0.00% | 0 | 33.33% | 1 | 33.33% | 1 |
| Eye | RGC | 0.00% | 0 | 25.00% | 1 | 50.00% | 2 |
| Eye | Amacrine Cell | 0.00% | 0 | 0.00% | 0 | 25.00% | 1 |
| Eye | PP | 0.00% | 0 | 0.00% | 0 | 25.00% | 1 |
| Liver | Stellate | 100.00% | 6 | 0.00% | 0 | 0.00% | 0 |
| Liver | Cholangiocyte | 27.27% | 3 | 9.09% | 1 | 0.00% | 0 |
| Liver | Hepatocyte | 20.00% | 2 | 70.00% | 7 | 100.00% | 10 |
| Pancreas | Endothelial Cell | 100.00% | 1 | 100.00% | 1 | 100.00% | 1 |
| Pancreas | Alpha | 100.00% | 1 | 0.00% | 0 | 100.00% | 1 |
| Pancreas | Ductal Cell | 100.00% | 1 | 0.00% | 0 | 100.00% | 1 |
| Pancreas | Proliferating Cell | 0.00% | 0 | 100.00% | 1 | 0.00% | 0 |
| Heart | Endothelial Cell | 100.00% | 1 | 100.00% | 1 | 100.00% | 1 |
| Heart | Pluripotent Cell | 100.00% | 1 | 0.00% | 0 | 100.00% | 1 |

**Supplementary Table S4. The 243 significant genes associated with very severe COVID-19 identified by S-MultiXcan analysis based on 49 GTEx tissues**

| Gene ID | Gene name | P value | FDR |
|---|---|---|---|
| ENSG00000163817.15 | *SLC6A20* | 7.16E-227 | 1.60E-222 |
| ENSG00000173585.15 | *CCR9* | 6.50E-220 | 7.26E-216 |
| ENSG00000172215.5 | *CXCR6* | 2.46E-118 | 1.83E-114 |
| ENSG00000173578.7 | *XCR1* | 1.81E-112 | 1.01E-108 |
| ENSG00000163818.16 | *LZTFL1* | 1.64E-101 | 7.33E-98 |
| ENSG00000183625.14 | *CCR3* | 7.31E-62 | 2.72E-58 |
| ENSG00000142002.16 | *DPP9* | 5.70E-51 | 1.82E-47 |
| ENSG00000163823.3 | *CCR1* | 2.85E-50 | 7.96E-47 |
| ENSG00000159110.19 | *IFNAR2* | 2.05E-47 | 5.09E-44 |
| ENSG00000160791.13 | *CCR5* | 9.97E-47 | 2.23E-43 |
| ENSG00000121807.5 | *CCR2* | 8.23E-45 | 1.67E-41 |
| ENSG00000243646.9 | *IL10RB* | 4.16E-42 | 7.74E-39 |
| ENSG00000163820.14 | *FYCO1* | 2.62E-41 | 4.50E-38 |
| ENSG00000121797.9 | *CCRL2* | 7.73E-35 | 1.23E-31 |
| ENSG00000173171.14 | *MTX1* | 1.52E-27 | 2.26E-24 |

| | | | |
|---|---|---|---|
| ENSG00000185499.16 | *MUC1* | 5.20E-25 | 7.26E-22 |
| ENSG00000116539.12 | *ASH1L* | 4.31E-22 | 5.66E-19 |
| ENSG00000163462.17 | *TRIM46* | 1.68E-19 | 2.08E-16 |
| ENSG00000137166.14 | *FOXP4* | 2.39E-17 | 2.79E-14 |
| ENSG00000273102.1 | *AP000569.9* | 2.50E-17 | 2.79E-14 |
| ENSG00000283646.1 | *LINC02009* | 1.41E-15 | 1.50E-12 |
| ENSG00000105397.13 | *TYK2* | 1.92E-15 | 1.95E-12 |
| ENSG00000169241.17 | *SLC50A1* | 1.78E-14 | 1.73E-11 |
| ENSG00000241837.6 | *ATP5O* | 2.45E-14 | 2.28E-11 |
| ENSG00000012223.12 | *LTF* | 6.51E-14 | 5.82E-11 |
| ENSG00000204542.2 | *C6orf15* | 2.56E-13 | 2.20E-10 |
| ENSG00000175164.13 | *ABO* | 4.92E-13 | 4.07E-10 |
| ENSG00000143537.13 | *ADAM15* | 5.20E-13 | 4.15E-10 |
| ENSG00000185361.8 | *TNFAIP8L1* | 1.00E-12 | 7.70E-10 |
| ENSG00000243364.7 | *EFNA4* | 5.09E-12 | 3.79E-09 |
| ENSG00000121691.4 | *CAT* | 9.25E-12 | 6.67E-09 |
| ENSG00000116580.18 | *GON4L* | 2.16E-11 | 1.51E-08 |
| ENSG00000243927.5 | *MRPS6* | 3.52E-11 | 2.38E-08 |
| ENSG00000204536.13 | *CCHCR1* | 3.86E-11 | 2.54E-08 |
| ENSG00000267198.1 | *RP11-798G7.6* | 9.93E-11 | 6.34E-08 |
| ENSG00000065989.15 | *PDE4A* | 2.09E-10 | 1.30E-07 |
| ENSG00000225190.10 | *PLEKHM1* | 2.29E-10 | 1.38E-07 |
| ENSG00000131400.7 | *NAPSA* | 2.36E-10 | 1.39E-07 |
| ENSG00000197536.11 | *C5orf56* | 5.25E-10 | 3.01E-07 |
| ENSG00000204525.16 | *HLA-C* | 6.13E-10 | 3.42E-07 |
| ENSG00000176920.11 | *FUT2* | 9.04E-10 | 4.93E-07 |
| ENSG00000204538.3 | *PSORS1C2* | 1.07E-09 | 5.69E-07 |
| ENSG00000135374.9 | *ELF5* | 1.30E-09 | 6.75E-07 |
| ENSG00000159314.11 | *ARHGAP27* | 1.36E-09 | 6.90E-07 |
| ENSG00000133661.15 | *SFTPD* | 1.94E-09 | 9.63E-07 |
| ENSG00000176715.15 | *ACSF3* | 2.03E-09 | 9.86E-07 |
| ENSG00000232300.1 | *FAM215B* | 2.12E-09 | 1.01E-06 |
| ENSG00000137310.11 | *TCF19* | 2.17E-09 | 1.01E-06 |
| ENSG00000267673.6 | *FDX2* | 2.37E-09 | 1.08E-06 |
| ENSG00000068650.18 | *ATP11A* | 2.60E-09 | 1.16E-06 |
| ENSG00000163463.11 | *KRTCAP2* | 3.02E-09 | 1.32E-06 |
| ENSG00000105376.4 | *ICAM5* | 4.01E-09 | 1.72E-06 |
| ENSG00000089127.12 | *OAS1* | 4.10E-09 | 1.73E-06 |
| ENSG00000220201.7 | *ZGLP1* | 5.36E-09 | 2.22E-06 |
| ENSG00000168743.12 | *NPNT* | 5.92E-09 | 2.40E-06 |
| ENSG00000112787.12 | *FBRSL1* | 7.15E-09 | 2.85E-06 |
| ENSG00000108379.9 | *WNT3* | 7.59E-09 | 2.97E-06 |
| ENSG00000105538.9 | *RASIP1* | 1.11E-08 | 4.28E-06 |
| ENSG00000121073.13 | *SLC35B1* | 1.21E-08 | 4.58E-06 |
| ENSG00000163354.14 | *DCST2* | 1.41E-08 | 5.25E-06 |
| ENSG00000176909.11 | *MAMSTR* | 1.60E-08 | 5.86E-06 |

| | | | |
|---|---|---|---|
| ENSG00000142233.11 | *NTN5* | 1.63E-08 | 5.87E-06 |
| ENSG00000206503.12 | *HLA-A* | 1.68E-08 | 5.96E-06 |
| ENSG00000167680.15 | *SEMA6B* | 3.46E-08 | 1.21E-05 |
| ENSG00000185303.16 | *SFTPA2* | 4.29E-08 | 1.47E-05 |
| ENSG00000226676.1 | *RP11-589B3.6* | 4.88E-08 | 1.65E-05 |
| ENSG00000211456.10 | *SACM1L* | 7.54E-08 | 2.51E-05 |
| ENSG00000111331.12 | *OAS3* | 1.24E-07 | 4.07E-05 |
| ENSG00000076662.9 | *ICAM3* | 1.41E-07 | 4.56E-05 |
| ENSG00000182264.8 | *IZUMO1* | 1.55E-07 | 4.95E-05 |
| ENSG00000244733.5 | *RP11-506M13.3* | 1.73E-07 | 5.44E-05 |
| ENSG00000228570.7 | *NUTM2E* | 1.76E-07 | 5.46E-05 |
| ENSG00000231485.1 | *RP4-535B20.1* | 1.88E-07 | 5.75E-05 |
| ENSG00000163825.3 | *RTP3* | 2.08E-07 | 6.28E-05 |
| ENSG00000104388.14 | *RAB2A* | 2.78E-07 | 8.28E-05 |
| ENSG00000204540.10 | *PSORS1C1* | 3.12E-07 | 9.17E-05 |
| ENSG00000090339.8 | *ICAM1* | 4.35E-07 | 1.26E-04 |
| ENSG00000161847.13 | *RAVER1* | 5.26E-07 | 1.51E-04 |
| ENSG00000272076.1 | *RP11-11C20.3* | 5.46E-07 | 1.54E-04 |
| ENSG00000251396.6 | *LINC01301* | 5.82E-07 | 1.63E-04 |
| ENSG00000174951.10 | *FUT1* | 6.25E-07 | 1.72E-04 |
| ENSG00000166508.17 | *MCM7* | 7.28E-07 | 1.98E-04 |
| ENSG00000179335.18 | *CLK3* | 9.45E-07 | 2.54E-04 |
| ENSG00000198563.13 | *DDX39B* | 1.19E-06 | 3.16E-04 |
| ENSG00000204619.7 | *PPP1R11* | 1.26E-06 | 3.31E-04 |
| ENSG00000204539.3 | *CDSN* | 1.33E-06 | 3.45E-04 |
| ENSG00000164398.12 | *ACSL6* | 1.44E-06 | 3.70E-04 |
| ENSG00000096696.13 | *DSP* | 1.51E-06 | 3.83E-04 |
| ENSG00000237541.3 | *HLA-DQA2* | 1.54E-06 | 3.87E-04 |
| ENSG00000142166.12 | *IFNAR1* | 1.60E-06 | 3.97E-04 |
| ENSG00000074842.7 | *MYDGF* | 1.86E-06 | 4.54E-04 |
| ENSG00000272447.1 | *RP11-182L21.6* | 1.87E-06 | 4.54E-04 |
| ENSG00000148730.6 | *EIF4EBP2* | 1.98E-06 | 4.76E-04 |
| ENSG00000204287.13 | *HLA-DRA* | 2.22E-06 | 5.28E-04 |
| ENSG00000238083.7 | *LRRC37A2* | 2.28E-06 | 5.33E-04 |
| ENSG00000259803.6 | *SLC22A31* | 2.29E-06 | 5.33E-04 |
| ENSG00000106261.16 | *ZKSCAN1* | 2.40E-06 | 5.53E-04 |
| ENSG00000272666.1 | *CTA-384D8.35* | 3.17E-06 | 7.23E-04 |
| ENSG00000170100.13 | *ZNF778* | 3.34E-06 | 7.54E-04 |
| ENSG00000127946.16 | *HIP1* | 3.52E-06 | 7.86E-04 |
| ENSG00000105401.8 | *CDC37* | 3.99E-06 | 8.82E-04 |
| ENSG00000129910.7 | *CDH15* | 4.69E-06 | 1.03E-03 |
| ENSG00000108344.14 | *PSMD3* | 4.86E-06 | 1.05E-03 |
| ENSG00000230746.1 | *AC006007.1* | 4.92E-06 | 1.06E-03 |
| ENSG00000131398.13 | *KCNC3* | 6.25E-06 | 1.32E-03 |
| ENSG00000204642.13 | *HLA-F* | 6.25E-06 | 1.32E-03 |
| ENSG00000204516.9 | *MICB* | 7.39E-06 | 1.54E-03 |

| | | | |
|---|---|---|---|
| ENSG00000186827.10 | *TNFRSF4* | 7.61E-06 | 1.57E-03 |
| ENSG00000196683.10 | *TOMM7* | 8.33E-06 | 1.71E-03 |
| ENSG00000130733.10 | *YIPF2* | 8.69E-06 | 1.76E-03 |
| ENSG00000203684.5 | *IBA57-AS1* | 1.08E-05 | 2.15E-03 |
| ENSG00000144802.11 | *NFKBIZ* | 1.08E-05 | 2.15E-03 |
| ENSG00000072682.18 | *P4HA2* | 1.13E-05 | 2.21E-03 |
| ENSG00000115520.8 | *COQ10B* | 1.13E-05 | 2.21E-03 |
| ENSG00000160505.15 | *NLRP4* | 1.36E-05 | 2.64E-03 |
| ENSG00000213420.7 | *GPC2* | 1.39E-05 | 2.68E-03 |
| ENSG00000188157.14 | *AGRN* | 1.42E-05 | 2.69E-03 |
| ENSG00000168090.9 | *COPS6* | 1.42E-05 | 2.69E-03 |
| ENSG00000176444.18 | *CLK2* | 1.82E-05 | 3.42E-03 |
| ENSG00000160862.12 | *AZGP1* | 2.15E-05 | 4.00E-03 |
| ENSG00000170608.2 | *FOXA3* | 2.24E-05 | 4.14E-03 |
| ENSG00000143630.9 | *HCN3* | 2.30E-05 | 4.21E-03 |
| ENSG00000162434.11 | *JAK1* | 2.40E-05 | 4.36E-03 |
| ENSG00000038210.12 | *PI4K2B* | 2.45E-05 | 4.41E-03 |
| ENSG00000116521.10 | *SCAMP3* | 2.50E-05 | 4.42E-03 |
| ENSG00000180422.3 | *LINC00304* | 2.51E-05 | 4.42E-03 |
| ENSG00000169242.11 | *EFNA1* | 2.52E-05 | 4.42E-03 |
| ENSG00000160685.13 | *ZBTB7B* | 2.54E-05 | 4.42E-03 |
| ENSG00000151498.11 | *ACAD8* | 2.55E-05 | 4.42E-03 |
| ENSG00000242574.8 | *HLA-DMB* | 2.60E-05 | 4.47E-03 |
| ENSG00000132972.18 | *RNF17* | 2.82E-05 | 4.81E-03 |
| ENSG00000204574.12 | *ABCF1* | 2.88E-05 | 4.87E-03 |
| ENSG00000142444.6 | *TIMM29* | 3.10E-05 | 5.21E-03 |
| ENSG00000205015.1 | *LINC02138* | 3.33E-05 | 5.55E-03 |
| ENSG00000185829.17 | *ARL17A* | 3.43E-05 | 5.68E-03 |
| ENSG00000204580.13 | *DDR1* | 3.49E-05 | 5.73E-03 |
| ENSG00000143622.10 | *RIT1* | 3.69E-05 | 6.02E-03 |
| ENSG00000188199.10 | *NUTM2B* | 3.93E-05 | 6.36E-03 |
| ENSG00000139725.7 | *RHOF* | 4.38E-05 | 7.04E-03 |
| ENSG00000132718.8 | *SYT11* | 4.61E-05 | 7.36E-03 |
| ENSG00000166997.7 | *CNPY4* | 4.78E-05 | 7.57E-03 |
| ENSG00000151914.19 | *DST* | 4.97E-05 | 7.82E-03 |
| ENSG00000166016.5 | *ABTB2* | 5.05E-05 | 7.89E-03 |
| ENSG00000162613.16 | *FUBP1* | 5.10E-05 | 7.91E-03 |
| ENSG00000163344.5 | *PMVK* | 5.35E-05 | 8.24E-03 |
| ENSG00000126214.21 | *KLC1* | 5.61E-05 | 8.58E-03 |
| ENSG00000185294.6 | *SPPL2C* | 5.70E-05 | 8.62E-03 |
| ENSG00000163346.16 | *PBXIP1* | 5.71E-05 | 8.62E-03 |
| ENSG00000108342.12 | *CSF3* | 5.78E-05 | 8.67E-03 |
| ENSG00000247626.4 | *MARS2* | 6.25E-05 | 9.28E-03 |
| ENSG00000078967.12 | *UBE2D4* | 6.27E-05 | 9.28E-03 |
| ENSG00000160752.14 | *FDPS* | 6.32E-05 | 9.29E-03 |
| ENSG00000256053.7 | *APOPT1* | 6.54E-05 | 9.55E-03 |

| | | | |
|---|---|---|---|
| ENSG00000226979.8 | *LTA* | 6.60E-05 | 9.57E-03 |
| ENSG00000239521.7 | *GATS* | 6.76E-05 | 9.74E-03 |
| ENSG00000117682.16 | *DHDDS* | 6.88E-05 | 9.77E-03 |
| ENSG00000171840.11 | *NINJ2* | 6.89E-05 | 9.77E-03 |
| ENSG00000008838.19 | *MED24* | 6.91E-05 | 9.77E-03 |
| ENSG00000204356.13 | *NELFE* | 7.86E-05 | 1.10E-02 |
| ENSG00000263893.2 | *CTD-3010D24.3* | 7.94E-05 | 1.11E-02 |
| ENSG00000162616.8 | *DNAJB4* | 8.40E-05 | 1.16E-02 |
| ENSG00000087266.15 | *SH3BP2* | 8.47E-05 | 1.16E-02 |
| ENSG00000118200.14 | *CAMSAP2* | 8.53E-05 | 1.16E-02 |
| ENSG00000168228.14 | *ZCCHC4* | 8.54E-05 | 1.16E-02 |
| ENSG00000165424.6 | *ZCCHC24* | 8.93E-05 | 1.21E-02 |
| ENSG00000021461.16 | *CYP3A43* | 9.26E-05 | 1.25E-02 |
| ENSG00000213722.8 | *DDAH2* | 9.84E-05 | 1.32E-02 |
| ENSG00000225180.7 | *AATK-AS1* | 1.00E-04 | 1.32E-02 |
| ENSG00000204610.12 | *TRIM15* | 1.00E-04 | 1.32E-02 |
| ENSG00000053900.10 | *ANAPC4* | 1.01E-04 | 1.32E-02 |
| ENSG00000143590.13 | *EFNA3* | 1.01E-04 | 1.32E-02 |
| ENSG00000143443.9 | *C1orf56* | 1.07E-04 | 1.39E-02 |
| ENSG00000197093.10 | *GAL3ST4* | 1.10E-04 | 1.42E-02 |
| ENSG00000163746.11 | *PLSCR2* | 1.19E-04 | 1.52E-02 |
| ENSG00000241370.5 | *RPP21* | 1.28E-04 | 1.64E-02 |
| ENSG00000119596.17 | *YLPM1* | 1.31E-04 | 1.66E-02 |
| ENSG00000204290.10 | *BTNL2* | 1.33E-04 | 1.67E-02 |
| ENSG00000163374.19 | *YY1AP1* | 1.33E-04 | 1.67E-02 |
| ENSG00000137478.14 | *FCHSD2* | 1.34E-04 | 1.67E-02 |
| ENSG00000228862.3 | *RP11-91I20.3* | 1.42E-04 | 1.77E-02 |
| ENSG00000122483.17 | *CCDC18* | 1.51E-04 | 1.86E-02 |
| ENSG00000213676.10 | *ATF6B* | 1.53E-04 | 1.87E-02 |
| ENSG00000162437.14 | *RAVER2* | 1.56E-04 | 1.91E-02 |
| ENSG00000217128.11 | *FNIP1* | 1.60E-04 | 1.94E-02 |
| ENSG00000188186.10 | *LAMTOR4* | 1.65E-04 | 1.98E-02 |
| ENSG00000197208.5 | *SLC22A4* | 1.65E-04 | 1.98E-02 |
| ENSG00000267011.5 | *CTB-50L17.16* | 1.66E-04 | 1.98E-02 |
| ENSG00000080819.6 | *CPOX* | 1.79E-04 | 2.13E-02 |
| ENSG00000198685.3 | *LINC01565* | 1.82E-04 | 2.15E-02 |
| ENSG00000163467.11 | *TSACC* | 1.95E-04 | 2.29E-02 |
| ENSG00000145495.15 | *6-Mar* | 1.97E-04 | 2.30E-02 |
| ENSG00000179361.17 | *ARID3B* | 2.07E-04 | 2.40E-02 |
| ENSG00000129354.11 | *AP1M2* | 2.11E-04 | 2.43E-02 |
| ENSG00000160767.20 | *FAM189B* | 2.11E-04 | 2.43E-02 |
| ENSG00000103355.13 | *PRSS33* | 2.21E-04 | 2.53E-02 |
| ENSG00000256875.2 | *RP11-503G7.2* | 2.34E-04 | 2.67E-02 |
| ENSG00000185507.19 | *IRF7* | 2.40E-04 | 2.73E-02 |
| ENSG00000243649.8 | *CFB* | 2.48E-04 | 2.80E-02 |
| ENSG00000241186.9 | *TDGF1* | 2.50E-04 | 2.81E-02 |

| | | | |
|---|---|---|---|
| ENSG00000120907.17 | *ADRA1A* | 2.51E-04 | 2.81E-02 |
| ENSG00000204174.7 | *NPY4R* | 2.54E-04 | 2.82E-02 |
| ENSG00000170791.17 | *CHCHD7* | 2.70E-04 | 2.98E-02 |
| ENSG00000115541.10 | *HSPE1* | 2.81E-04 | 3.09E-02 |
| ENSG00000198743.6 | *SLC5A3* | 2.97E-04 | 3.23E-02 |
| ENSG00000165841.9 | *CYP2C19* | 2.97E-04 | 3.23E-02 |
| ENSG00000117505.12 | *DR1* | 2.98E-04 | 3.23E-02 |
| ENSG00000172340.14 | *SUCLG2* | 3.00E-04 | 3.24E-02 |
| ENSG00000152382.5 | *TADA1* | 3.06E-04 | 3.29E-02 |
| ENSG00000110711.9 | *AIP* | 3.17E-04 | 3.39E-02 |
| ENSG00000166526.16 | *ZNF3* | 3.24E-04 | 3.43E-02 |
| ENSG00000146278.10 | *PNRC1* | 3.24E-04 | 3.43E-02 |
| ENSG00000087253.12 | *LPCAT2* | 3.31E-04 | 3.48E-02 |
| ENSG00000168010.10 | *ATG16L2* | 3.34E-04 | 3.50E-02 |
| ENSG00000150540.13 | *HNMT* | 3.37E-04 | 3.52E-02 |
| ENSG00000163902.11 | *RPN1* | 3.52E-04 | 3.65E-02 |
| ENSG00000132793.11 | *LPIN3* | 3.55E-04 | 3.65E-02 |
| ENSG00000120235.4 | *IFNA6* | 3.55E-04 | 3.65E-02 |
| ENSG00000236699.8 | *ARHGEF38* | 3.63E-04 | 3.72E-02 |
| ENSG00000198835.3 | *GJC2* | 3.70E-04 | 3.77E-02 |
| ENSG00000132676.15 | *DAP3* | 3.80E-04 | 3.86E-02 |
| ENSG00000105516.10 | *DBP* | 3.96E-04 | 4.00E-02 |
| ENSG00000183682.7 | *BMP8A* | 4.00E-04 | 4.03E-02 |
| ENSG00000105559.11 | *PLEKHA4* | 4.08E-04 | 4.09E-02 |
| ENSG00000182168.14 | *UNC5C* | 4.11E-04 | 4.10E-02 |
| ENSG00000186868.15 | *MAPT* | 4.21E-04 | 4.17E-02 |
| ENSG00000227507.2 | *LTB* | 4.23E-04 | 4.18E-02 |
| ENSG00000204438.10 | *GPANK1* | 4.37E-04 | 4.29E-02 |
| ENSG00000204531.17 | *POU5F1* | 4.38E-04 | 4.29E-02 |
| ENSG00000120071.13 | *KANSL1* | 4.40E-04 | 4.29E-02 |
| ENSG00000132680.10 | *KIAA0907* | 4.52E-04 | 4.37E-02 |
| ENSG00000167914.11 | *GSDMA* | 4.52E-04 | 4.37E-02 |
| ENSG00000197102.10 | *DYNC1H1* | 4.58E-04 | 4.41E-02 |
| ENSG00000160785.13 | *SLC25A44* | 4.61E-04 | 4.42E-02 |
| ENSG00000166589.12 | *CDH16* | 4.78E-04 | 4.56E-02 |
| ENSG00000204396.10 | *VWA7* | 5.00E-04 | 4.75E-02 |
| ENSG00000204252.13 | *HLA-DOA* | 5.12E-04 | 4.85E-02 |
| ENSG00000162944.10 | *RFTN2* | 5.16E-04 | 4.86E-02 |
| ENSG00000130164.13 | *LDLR* | 5.19E-04 | 4.88E-02 |
| ENSG00000135144.7 | *DTX1* | 5.25E-04 | 4.90E-02 |
| ENSG00000177103.13 | *DSCAML1* | 5.27E-04 | 4.90E-02 |
| ENSG00000188501.11 | *LCTL* | 5.30E-04 | 4.91E-02 |
| ENSG00000104804.7 | *TULP2* | 5.34E-04 | 4.93E-02 |
| ENSG00000169567.11 | *HINT1* | 5.38E-04 | 4.95E-02 |

**Supplementary Table S5. The 277 significant genes associated with hospitalized COVID-19 identified by S-MultiXcan analysis based on 49 GTEx tissues**

| Gene ID | Gene name | P value | FDR |
|---|---|---|---|
| ENSG00000173585.15 | *CCR9* | 3.04E-229 | 6.79E-225 |
| ENSG00000163817.15 | *SLC6A20* | 2.30E-217 | 2.57E-213 |
| ENSG00000172215.5 | *CXCR6* | 7.53E-114 | 5.61E-110 |
| ENSG00000163818.16 | *LZTFL1* | 5.65E-100 | 3.16E-96 |
| ENSG00000173578.7 | *XCR1* | 1.53E-86 | 6.84E-83 |
| ENSG00000183625.14 | *CCR3* | 6.93E-57 | 2.58E-53 |
| ENSG00000163820.14 | *FYCO1* | 2.54E-53 | 8.11E-50 |
| ENSG00000159110.19 | *IFNAR2* | 1.67E-50 | 4.66E-47 |
| ENSG00000163823.3 | *CCR1* | 5.25E-46 | 1.30E-42 |
| ENSG00000142002.16 | *DPP9* | 1.11E-41 | 2.48E-38 |
| ENSG00000243646.9 | *IL10RB* | 4.82E-41 | 9.79E-38 |
| ENSG00000160791.13 | *CCR5* | 7.76E-39 | 1.44E-35 |
| ENSG00000121807.5 | *CCR2* | 5.43E-38 | 9.33E-35 |
| ENSG00000121797.9 | *CCRL2* | 5.12E-35 | 8.17E-32 |
| ENSG00000137166.14 | *FOXP4* | 5.15E-26 | 7.67E-23 |
| ENSG00000185499.16 | *MUC1* | 2.21E-24 | 3.09E-21 |
| ENSG00000175164.13 | *ABO* | 9.66E-23 | 1.27E-19 |
| ENSG00000173171.14 | *MTX1* | 4.60E-22 | 5.71E-19 |
| ENSG00000273102.1 | *AP000569.9* | 5.59E-21 | 6.57E-18 |
| ENSG00000241837.6 | *ATP5O* | 4.95E-18 | 5.53E-15 |
| ENSG00000116539.12 | *ASH1L* | 3.14E-17 | 3.34E-14 |
| ENSG00000131400.7 | *NAPSA* | 4.33E-17 | 4.40E-14 |
| ENSG00000243927.5 | *MRPS6* | 2.59E-16 | 2.52E-13 |
| ENSG00000283646.1 | *LINC02009* | 4.35E-16 | 4.05E-13 |
| ENSG00000135374.9 | *ELF5* | 1.81E-14 | 1.62E-11 |
| ENSG00000121691.4 | *CAT* | 3.20E-14 | 2.75E-11 |
| ENSG00000012223.12 | *LTF* | 5.44E-14 | 4.50E-11 |
| ENSG00000144802.11 | *NFKBIZ* | 6.81E-13 | 5.43E-10 |
| ENSG00000068650.18 | *ATP11A* | 2.53E-12 | 1.95E-09 |
| ENSG00000163462.17 | *TRIM46* | 8.52E-12 | 6.34E-09 |
| ENSG00000143537.13 | *ADAM15* | 9.88E-12 | 7.12E-09 |
| ENSG00000267198.1 | *RP11-798G7.6* | 1.60E-11 | 1.12E-08 |
| ENSG00000169241.17 | *SLC50A1* | 2.12E-11 | 1.44E-08 |
| ENSG00000112787.12 | *FBRSL1* | 2.24E-11 | 1.47E-08 |
| ENSG00000204542.2 | *C6orf15* | 6.91E-11 | 4.41E-08 |
| ENSG00000163463.11 | *KRTCAP2* | 7.93E-11 | 4.92E-08 |
| ENSG00000111331.12 | *OAS3* | 9.33E-11 | 5.63E-08 |
| ENSG00000144815.15 | *NXPE3* | 1.06E-10 | 6.23E-08 |
| ENSG00000267673.6 | *FDX2* | 1.21E-10 | 6.93E-08 |
| ENSG00000204536.13 | *CCHCR1* | 1.38E-10 | 7.71E-08 |
| ENSG00000133661.15 | *SFTPD* | 3.46E-10 | 1.89E-07 |
| ENSG00000220201.7 | *ZGLP1* | 3.84E-10 | 2.04E-07 |
| ENSG00000137310.11 | *TCF19* | 4.83E-10 | 2.51E-07 |

| | | | |
|---|---|---|---|
| ENSG00000159314.11 | *ARHGAP27* | 6.74E-10 | 3.42E-07 |
| ENSG00000121073.13 | *SLC35B1* | 7.18E-10 | 3.56E-07 |
| ENSG00000204525.16 | *HLA-C* | 9.51E-10 | 4.58E-07 |
| ENSG00000197536.11 | *C5orf56* | 9.66E-10 | 4.58E-07 |
| ENSG00000198563.13 | *DDX39B* | 9.84E-10 | 4.58E-07 |
| ENSG00000105397.13 | *TYK2* | 1.41E-09 | 6.34E-07 |
| ENSG00000176920.11 | *FUT2* | 1.42E-09 | 6.34E-07 |
| ENSG00000225190.10 | *PLEKHM1* | 1.67E-09 | 7.31E-07 |
| ENSG00000232300.1 | *FAM215B* | 1.71E-09 | 7.35E-07 |
| ENSG00000065989.15 | *PDE4A* | 2.66E-09 | 1.12E-06 |
| ENSG00000161847.13 | *RAVER1* | 2.81E-09 | 1.16E-06 |
| ENSG00000182504.10 | *CEP97* | 2.92E-09 | 1.19E-06 |
| ENSG00000076662.9 | *ICAM3* | 4.54E-09 | 1.81E-06 |
| ENSG00000168228.14 | *ZCCHC4* | 4.68E-09 | 1.82E-06 |
| ENSG00000131398.13 | *KCNC3* | 4.73E-09 | 1.82E-06 |
| ENSG00000066422.4 | *ZBTB11* | 5.48E-09 | 2.07E-06 |
| ENSG00000185361.8 | *TNFAIP8L1* | 6.77E-09 | 2.52E-06 |
| ENSG00000108379.9 | *WNT3* | 9.15E-09 | 3.35E-06 |
| ENSG00000038210.12 | *PI4K2B* | 2.11E-08 | 7.60E-06 |
| ENSG00000204538.3 | *PSORS1C2* | 3.28E-08 | 1.16E-05 |
| ENSG00000185303.16 | *SFTPA2* | 4.27E-08 | 1.49E-05 |
| ENSG00000129910.7 | *CDH15* | 4.59E-08 | 1.58E-05 |
| ENSG00000251396.6 | *LINC01301* | 4.93E-08 | 1.67E-05 |
| ENSG00000104388.14 | *RAB2A* | 5.91E-08 | 1.97E-05 |
| ENSG00000089127.12 | *OAS1* | 9.67E-08 | 3.18E-05 |
| ENSG00000203684.5 | *IBA57-AS1* | 1.05E-07 | 3.40E-05 |
| ENSG00000133884.9 | *DPF2* | 1.33E-07 | 4.24E-05 |
| ENSG00000163344.5 | *PMVK* | 1.63E-07 | 5.13E-05 |
| ENSG00000103653.16 | *CSK* | 1.86E-07 | 5.77E-05 |
| ENSG00000204516.9 | *MICB* | 1.94E-07 | 5.94E-05 |
| ENSG00000226676.1 | *RP11-589B3.6* | 2.69E-07 | 8.12E-05 |
| ENSG00000185829.17 | *ARL17A* | 2.76E-07 | 8.22E-05 |
| ENSG00000163825.3 | *RTP3* | 3.72E-07 | 1.09E-04 |
| ENSG00000243364.7 | *EFNA4* | 4.83E-07 | 1.40E-04 |
| ENSG00000168743.12 | *NPNT* | 5.17E-07 | 1.48E-04 |
| ENSG00000211456.10 | *SACM1L* | 5.31E-07 | 1.50E-04 |
| ENSG00000197208.5 | *SLC22A4* | 6.01E-07 | 1.68E-04 |
| ENSG00000176715.15 | *ACSF3* | 7.10E-07 | 1.96E-04 |
| ENSG00000115520.8 | *COQ10B* | 7.76E-07 | 2.11E-04 |
| ENSG00000226979.8 | *LTA* | 8.00E-07 | 2.15E-04 |
| ENSG00000238083.7 | *LRRC37A2* | 8.62E-07 | 2.29E-04 |
| ENSG00000142233.11 | *NTN5* | 1.07E-06 | 2.81E-04 |
| ENSG00000166526.16 | *ZNF3* | 1.09E-06 | 2.83E-04 |
| ENSG00000170100.13 | *ZNF778* | 1.21E-06 | 3.11E-04 |
| ENSG00000198743.6 | *SLC5A3* | 1.32E-06 | 3.35E-04 |
| ENSG00000096088.16 | *PGC* | 1.46E-06 | 3.66E-04 |

| | | | |
|---|---|---|---|
| ENSG00000170608.2 | *FOXA3* | 1.72E-06 | 4.22E-04 |
| ENSG00000105376.4 | *ICAM5* | 1.72E-06 | 4.22E-04 |
| ENSG00000135144.7 | *DTX1* | 1.84E-06 | 4.47E-04 |
| ENSG00000053900.10 | *ANAPC4* | 1.99E-06 | 4.78E-04 |
| ENSG00000259803.6 | *SLC22A31* | 2.06E-06 | 4.90E-04 |
| ENSG00000106261.16 | *ZKSCAN1* | 2.26E-06 | 5.26E-04 |
| ENSG00000074842.7 | *MYDGF* | 2.26E-06 | 5.26E-04 |
| ENSG00000112578.9 | *BYSL* | 3.12E-06 | 7.19E-04 |
| ENSG00000166508.17 | *MCM7* | 3.18E-06 | 7.25E-04 |
| ENSG00000164398.12 | *ACSL6* | 4.66E-06 | 1.05E-03 |
| ENSG00000090339.8 | *ICAM1* | 4.70E-06 | 1.05E-03 |
| ENSG00000272447.1 | *RP11-182L21.6* | 4.90E-06 | 1.08E-03 |
| ENSG00000204287.13 | *HLA-DRA* | 5.00E-06 | 1.10E-03 |
| ENSG00000162613.16 | *FUBP1* | 5.13E-06 | 1.10E-03 |
| ENSG00000087266.15 | *SH3BP2* | 5.14E-06 | 1.10E-03 |
| ENSG00000221843.4 | *C2orf16* | 5.63E-06 | 1.20E-03 |
| ENSG00000116580.18 | *GON4L* | 5.81E-06 | 1.22E-03 |
| ENSG00000081154.11 | *PCNP* | 6.17E-06 | 1.29E-03 |
| ENSG00000138468.15 | *SENP7* | 6.32E-06 | 1.31E-03 |
| ENSG00000239521.7 | *GATS* | 7.30E-06 | 1.49E-03 |
| ENSG00000160752.14 | *FDPS* | 7.32E-06 | 1.49E-03 |
| ENSG00000231485.1 | *RP4-535B20.1* | 7.79E-06 | 1.57E-03 |
| ENSG00000140465.13 | *CYP1A1* | 8.19E-06 | 1.63E-03 |
| ENSG00000223865.10 | *HLA-DPB1* | 1.03E-05 | 2.04E-03 |
| ENSG00000204531.17 | *POU5F1* | 1.09E-05 | 2.14E-03 |
| ENSG00000269387.1 | *RP11-298J23.8* | 1.23E-05 | 2.39E-03 |
| ENSG00000162616.8 | *DNAJB4* | 1.36E-05 | 2.62E-03 |
| ENSG00000137218.10 | *FRS3* | 1.38E-05 | 2.63E-03 |
| ENSG00000204540.10 | *PSORS1C1* | 1.39E-05 | 2.63E-03 |
| ENSG00000213676.10 | *ATF6B* | 1.45E-05 | 2.70E-03 |
| ENSG00000108344.14 | *PSMD3* | 1.45E-05 | 2.70E-03 |
| ENSG00000143774.16 | *GUK1* | 1.58E-05 | 2.92E-03 |
| ENSG00000163354.14 | *DCST2* | 1.63E-05 | 2.98E-03 |
| ENSG00000124593.15 | *RP11-298J23.10* | 1.80E-05 | 3.27E-03 |
| ENSG00000213420.7 | *GPC2* | 2.03E-05 | 3.66E-03 |
| ENSG00000277186.1 | *RP13-554M15.8* | 2.10E-05 | 3.75E-03 |
| ENSG00000196683.10 | *TOMM7* | 2.23E-05 | 3.95E-03 |
| ENSG00000204610.12 | *TRIM15* | 2.70E-05 | 4.75E-03 |
| ENSG00000204290.10 | *BTNL2* | 2.97E-05 | 5.18E-03 |
| ENSG00000180422.3 | *LINC00304* | 3.06E-05 | 5.30E-03 |
| ENSG00000168090.9 | *COPS6* | 3.11E-05 | 5.34E-03 |
| ENSG00000227507.2 | *LTB* | 3.44E-05 | 5.87E-03 |
| ENSG00000147655.10 | *RSPO2* | 3.59E-05 | 6.08E-03 |
| ENSG00000122852.14 | *SFTPA1* | 3.68E-05 | 6.18E-03 |
| ENSG00000244733.5 | *RP11-506M13.3* | 3.90E-05 | 6.50E-03 |
| ENSG00000160685.13 | *ZBTB7B* | 4.10E-05 | 6.78E-03 |

| | | | |
|---|---|---|---|
| ENSG00000142687.17 | *KIAA0319L* | 4.19E-05 | 6.85E-03 |
| ENSG00000227059.6 | *ANHX* | 4.20E-05 | 6.85E-03 |
| ENSG00000278505.4 | *C17orf78* | 4.24E-05 | 6.86E-03 |
| ENSG00000143793.12 | *C1orf35* | 4.32E-05 | 6.94E-03 |
| ENSG00000105538.9 | *RASIP1* | 4.51E-05 | 7.20E-03 |
| ENSG00000167183.2 | *PRR15L* | 4.65E-05 | 7.36E-03 |
| ENSG00000142166.12 | *IFNAR1* | 4.71E-05 | 7.36E-03 |
| ENSG00000204344.14 | *STK19* | 4.72E-05 | 7.36E-03 |
| ENSG00000125967.16 | *NECAB3* | 4.77E-05 | 7.36E-03 |
| ENSG00000120235.4 | *IFNA6* | 4.78E-05 | 7.36E-03 |
| ENSG00000204642.13 | *HLA-F* | 5.27E-05 | 8.06E-03 |
| ENSG00000278224.5 | *PRICKLE4* | 5.55E-05 | 8.43E-03 |
| ENSG00000175130.6 | *MARCKSL1* | 6.52E-05 | 9.84E-03 |
| ENSG00000228570.7 | *NUTM2E* | 7.04E-05 | 1.06E-02 |
| ENSG00000266010.1 | *GATA6-AS1* | 7.34E-05 | 1.09E-02 |
| ENSG00000168487.17 | *BMP1* | 7.38E-05 | 1.09E-02 |
| ENSG00000234745.10 | *HLA-B* | 7.45E-05 | 1.09E-02 |
| ENSG00000120071.13 | *KANSL1* | 7.69E-05 | 1.12E-02 |
| ENSG00000152582.13 | *SPEF2* | 7.91E-05 | 1.15E-02 |
| ENSG00000166016.5 | *ABTB2* | 8.15E-05 | 1.17E-02 |
| ENSG00000204356.13 | *NELFE* | 8.15E-05 | 1.17E-02 |
| ENSG00000166529.14 | *ZSCAN21* | 8.23E-05 | 1.17E-02 |
| ENSG00000188199.10 | *NUTM2B* | 8.69E-05 | 1.23E-02 |
| ENSG00000156587.15 | *UBE2L6* | 8.72E-05 | 1.23E-02 |
| ENSG00000249931.4 | *GOLGA8K* | 8.92E-05 | 1.25E-02 |
| ENSG00000116819.6 | *TFAP2E* | 9.23E-05 | 1.28E-02 |
| ENSG00000148737.16 | *TCF7L2* | 9.52E-05 | 1.31E-02 |
| ENSG00000125755.18 | *SYMPK* | 9.69E-05 | 1.33E-02 |
| ENSG00000164463.12 | *CREBRF* | 1.04E-04 | 1.42E-02 |
| ENSG00000125459.14 | *MSTO1* | 1.07E-04 | 1.45E-02 |
| ENSG00000122359.17 | *ANXA11* | 1.07E-04 | 1.45E-02 |
| ENSG00000008838.19 | *MED24* | 1.10E-04 | 1.47E-02 |
| ENSG00000121067.17 | *SPOP* | 1.14E-04 | 1.51E-02 |
| ENSG00000272076.1 | *RP11-11C20.3* | 1.24E-04 | 1.64E-02 |
| ENSG00000228696.8 | *ARL17B* | 1.28E-04 | 1.67E-02 |
| ENSG00000204387.12 | *C6orf48* | 1.28E-04 | 1.67E-02 |
| ENSG00000111335.12 | *OAS2* | 1.33E-04 | 1.72E-02 |
| ENSG00000186868.15 | *MAPT* | 1.35E-04 | 1.75E-02 |
| ENSG00000034713.7 | *GABARAPL2* | 1.37E-04 | 1.76E-02 |
| ENSG00000142973.13 | *CYP4B1* | 1.40E-04 | 1.79E-02 |
| ENSG00000108342.12 | *CSF3* | 1.42E-04 | 1.80E-02 |
| ENSG00000179151.11 | *EDC3* | 1.45E-04 | 1.82E-02 |
| ENSG00000243649.8 | *CFB* | 1.48E-04 | 1.86E-02 |
| ENSG00000105401.8 | *CDC37* | 1.52E-04 | 1.89E-02 |
| ENSG00000141027.20 | *NCOR1* | 1.53E-04 | 1.89E-02 |
| ENSG00000028528.14 | *SNX1* | 1.53E-04 | 1.89E-02 |

| | | | |
|---|---|---|---|
| ENSG00000124721.17 | *DNAH8* | 1.56E-04 | 1.91E-02 |
| ENSG00000137767.13 | *SQRDL* | 1.59E-04 | 1.94E-02 |
| ENSG00000267505.1 | *CTC-296K1.3* | 1.64E-04 | 1.99E-02 |
| ENSG00000159147.17 | *DONSON* | 1.66E-04 | 2.00E-02 |
| ENSG00000186075.12 | *ZPBP2* | 1.66E-04 | 2.00E-02 |
| ENSG00000163346.16 | *PBXIP1* | 1.67E-04 | 2.00E-02 |
| ENSG00000204539.3 | *CDSN* | 1.68E-04 | 2.00E-02 |
| ENSG00000020129.15 | *NCDN* | 1.74E-04 | 2.06E-02 |
| ENSG00000181873.12 | *IBA57* | 1.77E-04 | 2.08E-02 |
| ENSG00000179218.13 | *CALR* | 1.78E-04 | 2.08E-02 |
| ENSG00000126091.19 | *ST3GAL3* | 1.80E-04 | 2.10E-02 |
| ENSG00000164031.16 | *DNAJB14* | 1.81E-04 | 2.10E-02 |
| ENSG00000125743.10 | *SNRPD2* | 1.87E-04 | 2.16E-02 |
| ENSG00000176909.11 | *MAMSTR* | 1.89E-04 | 2.16E-02 |
| ENSG00000163357.10 | *DCST1* | 1.96E-04 | 2.23E-02 |
| ENSG00000172057.9 | *ORMDL3* | 1.98E-04 | 2.23E-02 |
| ENSG00000166997.7 | *CNPY4* | 1.98E-04 | 2.23E-02 |
| ENSG00000162434.11 | *JAK1* | 2.06E-04 | 2.31E-02 |
| ENSG00000126214.21 | *KLC1* | 2.13E-04 | 2.38E-02 |
| ENSG00000204520.12 | *MICA* | 2.14E-04 | 2.38E-02 |
| ENSG00000230174.1 | *LINC01149* | 2.27E-04 | 2.51E-02 |
| ENSG00000106290.14 | *TAF6* | 2.29E-04 | 2.51E-02 |
| ENSG00000204438.10 | *GPANK1* | 2.29E-04 | 2.51E-02 |
| ENSG00000227456.7 | *LINC00310* | 2.34E-04 | 2.54E-02 |
| ENSG00000075539.13 | *FRYL* | 2.35E-04 | 2.54E-02 |
| ENSG00000101412.12 | *E2F1* | 2.35E-04 | 2.54E-02 |
| ENSG00000204314.10 | *PRRT1* | 2.39E-04 | 2.56E-02 |
| ENSG00000237523.1 | *LINC00857* | 2.43E-04 | 2.60E-02 |
| ENSG00000214941.7 | *ZSWIM7* | 2.47E-04 | 2.63E-02 |
| ENSG00000197093.10 | *GAL3ST4* | 2.55E-04 | 2.69E-02 |
| ENSG00000171103.10 | *TRMT61B* | 2.55E-04 | 2.69E-02 |
| ENSG00000198835.3 | *GJC2* | 2.57E-04 | 2.69E-02 |
| ENSG00000126067.11 | *PSMB2* | 2.59E-04 | 2.69E-02 |
| ENSG00000122420.9 | *PTGFR* | 2.60E-04 | 2.69E-02 |
| ENSG00000111237.18 | *VPS29* | 2.60E-04 | 2.69E-02 |
| ENSG00000204256.12 | *BRD2* | 2.65E-04 | 2.73E-02 |
| ENSG00000143590.13 | *EFNA3* | 2.67E-04 | 2.74E-02 |
| ENSG00000247626.4 | *MARS2* | 2.72E-04 | 2.77E-02 |
| ENSG00000272666.1 | *CTA-384D8.35* | 2.92E-04 | 2.96E-02 |
| ENSG00000248161.5 | *RP11-499E18.1* | 2.93E-04 | 2.96E-02 |
| ENSG00000167635.11 | *ZNF146* | 2.95E-04 | 2.97E-02 |
| ENSG00000113734.17 | *BNIP1* | 2.98E-04 | 2.98E-02 |
| ENSG00000178257.3 | *PRM3* | 3.08E-04 | 3.08E-02 |
| ENSG00000166797.10 | *FAM96A* | 3.11E-04 | 3.09E-02 |
| ENSG00000126215.13 | *XRCC3* | 3.29E-04 | 3.25E-02 |
| ENSG00000215009.5 | *ACSM4* | 3.35E-04 | 3.30E-02 |

| | | | |
|---|---|---|---|
| ENSG00000131435.12 | *PDLIM4* | 3.38E-04 | 3.31E-02 |
| ENSG00000188186.10 | *LAMTOR4* | 3.41E-04 | 3.32E-02 |
| ENSG00000168476.11 | *REEP4* | 3.49E-04 | 3.39E-02 |
| ENSG00000231389.7 | *HLA-DPA1* | 3.56E-04 | 3.45E-02 |
| ENSG00000221838.9 | *AP4M1* | 3.63E-04 | 3.49E-02 |
| ENSG00000206503.12 | *HLA-A* | 3.81E-04 | 3.65E-02 |
| ENSG00000187987.9 | *ZSCAN23* | 3.90E-04 | 3.72E-02 |
| ENSG00000198040.10 | *ZNF84* | 3.94E-04 | 3.72E-02 |
| ENSG00000140506.16 | *LMAN1L* | 3.94E-04 | 3.72E-02 |
| ENSG00000228672.3 | *PROB1* | 3.95E-04 | 3.72E-02 |
| ENSG00000259772.6 | *RP11-16E12.2* | 3.96E-04 | 3.72E-02 |
| ENSG00000073605.18 | *GSDMB* | 4.05E-04 | 3.78E-02 |
| ENSG00000153179.12 | *RASSF3* | 4.13E-04 | 3.84E-02 |
| ENSG00000112706.11 | *IMPG1* | 4.25E-04 | 3.91E-02 |
| ENSG00000109390.11 | *NDUFC1* | 4.27E-04 | 3.91E-02 |
| ENSG00000021461.16 | *CYP3A43* | 4.27E-04 | 3.91E-02 |
| ENSG00000225930.3 | *DKFZP434L187* | 4.29E-04 | 3.91E-02 |
| ENSG00000122591.11 | *FAM126A* | 4.29E-04 | 3.91E-02 |
| ENSG00000213578.5 | *CPLX3* | 4.31E-04 | 3.91E-02 |
| ENSG00000161405.16 | *IKZF3* | 4.40E-04 | 3.98E-02 |
| ENSG00000187626.8 | *ZKSCAN4* | 4.56E-04 | 4.10E-02 |
| ENSG00000171695.10 | *LKAAEAR1* | 4.57E-04 | 4.10E-02 |
| ENSG00000167083.6 | *GNGT2* | 4.62E-04 | 4.13E-02 |
| ENSG00000163171.7 | *CDC42EP3* | 4.68E-04 | 4.15E-02 |
| ENSG00000115541.10 | *HSPE1* | 4.71E-04 | 4.15E-02 |
| ENSG00000173418.11 | *NAA20* | 4.71E-04 | 4.15E-02 |
| ENSG00000110079.16 | *MS4A4A* | 4.74E-04 | 4.15E-02 |
| ENSG00000154358.20 | *OBSCN* | 4.76E-04 | 4.15E-02 |
| ENSG00000162913.9 | *C1orf145* | 4.76E-04 | 4.15E-02 |
| ENSG00000164663.14 | *USP49* | 4.79E-04 | 4.16E-02 |
| ENSG00000167914.11 | *GSDMA* | 4.83E-04 | 4.18E-02 |
| ENSG00000120451.10 | *SNX19* | 4.87E-04 | 4.19E-02 |
| ENSG00000155034.18 | *FBXL18* | 4.87E-04 | 4.19E-02 |
| ENSG00000169242.11 | *EFNA1* | 4.94E-04 | 4.23E-02 |
| ENSG00000177202.2 | *SPACA4* | 5.01E-04 | 4.27E-02 |
| ENSG00000160505.15 | *NLRP4* | 5.04E-04 | 4.28E-02 |
| ENSG00000164134.12 | *NAA15* | 5.07E-04 | 4.29E-02 |
| ENSG00000132153.14 | *DHX30* | 5.10E-04 | 4.29E-02 |
| ENSG00000168300.13 | *PCMTD1* | 5.11E-04 | 4.29E-02 |
| ENSG00000100601.9 | *ALKBH1* | 5.19E-04 | 4.34E-02 |
| ENSG00000072682.18 | *P4HA2* | 5.34E-04 | 4.45E-02 |
| ENSG00000163374.19 | *YY1AP1* | 5.35E-04 | 4.45E-02 |
| ENSG00000078018.19 | *MAP2* | 5.54E-04 | 4.58E-02 |
| ENSG00000066923.17 | *STAG3* | 5.57E-04 | 4.59E-02 |
| ENSG00000137265.14 | *IRF4* | 5.61E-04 | 4.60E-02 |
| ENSG00000105559.11 | *PLEKHA4* | 5.67E-04 | 4.64E-02 |

| | | | |
|---|---|---|---|
| ENSG00000057294.14 | *PKP2* | 5.69E-04 | 4.64E-02 |
| ENSG00000079819.18 | *EPB41L2* | 5.80E-04 | 4.72E-02 |
| ENSG00000110711.9 | *AIP* | 5.87E-04 | 4.75E-02 |
| ENSG00000204410.14 | *MSH5* | 5.94E-04 | 4.79E-02 |

**Supplementary Table S6. The 158 significant genes associated with susceptible COVID-19 identified by S-MultiXcan analysis based on 49 GTEx tissues**

| Gene | Gene name | P value | FDR |
|---|---|---|---|
| ENSG00000163817.15 | *SLC6A20* | 1.92E-171 | 4.29E-167 |
| ENSG00000175164.13 | *ABO* | 2.37E-82 | 2.65E-78 |
| ENSG00000173585.15 | *CCR9* | 5.27E-76 | 3.92E-72 |
| ENSG00000172215.5 | *CXCR6* | 2.02E-36 | 1.13E-32 |
| ENSG00000163818.16 | *LZTFL1* | 1.59E-35 | 7.10E-32 |
| ENSG00000173578.7 | *XCR1* | 8.96E-30 | 3.34E-26 |
| ENSG00000144802.11 | *NFKBIZ* | 2.22E-29 | 7.08E-26 |
| ENSG00000144815.15 | *NXPE3* | 6.68E-28 | 1.87E-24 |
| ENSG00000182504.10 | *CEP97* | 5.21E-27 | 1.29E-23 |
| ENSG00000066422.4 | *ZBTB11* | 7.65E-25 | 1.71E-21 |
| ENSG00000142002.16 | *DPP9* | 2.27E-23 | 4.61E-20 |
| ENSG00000163820.14 | *FYCO1* | 4.83E-20 | 8.99E-17 |
| ENSG00000160325.14 | *CACFD1* | 5.89E-20 | 1.01E-16 |
| ENSG00000104804.7 | *TULP2* | 5.44E-17 | 8.68E-14 |
| ENSG00000081154.11 | *PCNP* | 1.16E-16 | 1.73E-13 |
| ENSG00000138468.15 | *SENP7* | 3.46E-16 | 4.83E-13 |
| ENSG00000183625.14 | *CCR3* | 5.44E-16 | 7.15E-13 |
| ENSG00000163823.3 | *CCR1* | 7.72E-16 | 9.58E-13 |
| ENSG00000159110.19 | *IFNAR2* | 9.43E-16 | 1.11E-12 |
| ENSG00000176920.11 | *FUT2* | 2.50E-15 | 2.79E-12 |
| ENSG00000148297.15 | *MED22* | 1.03E-14 | 1.10E-11 |
| ENSG00000243646.9 | *IL10RB* | 1.31E-13 | 1.33E-10 |
| ENSG00000160326.13 | *SLC2A6* | 4.51E-13 | 4.38E-10 |
| ENSG00000137166.14 | *FOXP4* | 6.10E-13 | 5.68E-10 |
| ENSG00000105559.11 | *PLEKHA4* | 7.47E-13 | 6.67E-10 |
| ENSG00000142233.11 | *NTN5* | 1.15E-12 | 9.88E-10 |
| ENSG00000160323.18 | *ADAMTS13* | 3.38E-12 | 2.80E-09 |
| ENSG00000087074.7 | *PPP1R15A* | 1.63E-11 | 1.30E-08 |
| ENSG00000105538.9 | *RASIP1* | 3.09E-11 | 2.38E-08 |
| ENSG00000104805.15 | *NUCB1* | 4.39E-11 | 3.27E-08 |
| ENSG00000176909.11 | *MAMSTR* | 4.55E-11 | 3.28E-08 |
| ENSG00000160791.13 | *CCR5* | 7.64E-11 | 5.33E-08 |
| ENSG00000185499.16 | *MUC1* | 8.53E-11 | 5.77E-08 |
| ENSG00000121807.5 | *CCR2* | 9.53E-11 | 6.26E-08 |
| ENSG00000174951.10 | *FUT1* | 2.93E-10 | 1.87E-07 |
| ENSG00000133661.15 | *SFTPD* | 9.84E-10 | 6.11E-07 |
| ENSG00000182264.8 | *IZUMO1* | 1.13E-09 | 6.82E-07 |

| | | | |
|---|---|---|---|
| ENSG00000148300.11 | *REXO4* | 1.92E-09 | 1.13E-06 |
| ENSG00000163462.17 | *TRIM46* | 6.17E-09 | 3.53E-06 |
| ENSG00000163463.11 | *KRTCAP2* | 1.51E-08 | 8.43E-06 |
| ENSG00000121797.9 | *CCRL2* | 1.83E-08 | 9.97E-06 |
| ENSG00000087076.8 | *HSD17B14* | 1.90E-08 | 1.01E-05 |
| ENSG00000108342.12 | *CSF3* | 2.46E-08 | 1.28E-05 |
| ENSG00000148303.16 | *RPL7A* | 2.55E-08 | 1.29E-05 |
| ENSG00000131400.7 | *NAPSA* | 3.19E-08 | 1.58E-05 |
| ENSG00000204536.13 | *CCHCR1* | 3.53E-08 | 1.71E-05 |
| ENSG00000111331.12 | *OAS3* | 4.69E-08 | 2.23E-05 |
| ENSG00000169231.13 | *THBS3* | 5.75E-08 | 2.68E-05 |
| ENSG00000173171.14 | *MTX1* | 6.21E-08 | 2.83E-05 |
| ENSG00000008838.19 | *MED24* | 1.16E-07 | 5.18E-05 |
| ENSG00000167914.11 | *GSDMA* | 1.45E-07 | 6.35E-05 |
| ENSG00000108344.14 | *PSMD3* | 1.65E-07 | 7.09E-05 |
| ENSG00000267673.6 | *FDX2* | 1.71E-07 | 7.21E-05 |
| ENSG00000204525.16 | *HLA-C* | 2.34E-07 | 9.68E-05 |
| ENSG00000273102.1 | *AP000569.9* | 2.41E-07 | 9.79E-05 |
| ENSG00000265799.1 | *RP11-387H17.6* | 2.47E-07 | 9.85E-05 |
| ENSG00000231389.7 | *HLA-DPA1* | 3.07E-07 | 1.20E-04 |
| ENSG00000233577.6 | *RP3-462D8.2* | 3.56E-07 | 1.37E-04 |
| ENSG00000116604.17 | *MEF2D* | 3.76E-07 | 1.42E-04 |
| ENSG00000114391.12 | *RPL24* | 5.53E-07 | 2.06E-04 |
| ENSG00000177628.15 | *GBA* | 6.65E-07 | 2.44E-04 |
| ENSG00000241837.6 | *ATP5O* | 7.14E-07 | 2.57E-04 |
| ENSG00000081148.11 | *IMPG2* | 8.01E-07 | 2.80E-04 |
| ENSG00000204538.3 | *PSORS1C2* | 8.02E-07 | 2.80E-04 |
| ENSG00000105516.10 | *DBP* | 8.83E-07 | 3.03E-04 |
| ENSG00000223865.10 | *HLA-DPB1* | 1.78E-06 | 6.02E-04 |
| ENSG00000177202.2 | *SPACA4* | 2.07E-06 | 6.90E-04 |
| ENSG00000089127.12 | *OAS1* | 2.11E-06 | 6.93E-04 |
| ENSG00000106327.12 | *TFR2* | 2.19E-06 | 7.09E-04 |
| ENSG00000160785.13 | *SLC25A44* | 2.29E-06 | 7.31E-04 |
| ENSG00000160783.19 | *PMF1* | 2.49E-06 | 7.83E-04 |
| ENSG00000106330.11 | *MOSPD3* | 2.80E-06 | 8.69E-04 |
| ENSG00000077080.9 | *ACTL6B* | 3.26E-06 | 9.98E-04 |
| ENSG00000243927.5 | *MRPS6* | 3.42E-06 | 1.03E-03 |
| ENSG00000185361.8 | *TNFAIP8L1* | 3.48E-06 | 1.04E-03 |
| ENSG00000169242.11 | *EFNA1* | 3.93E-06 | 1.16E-03 |
| ENSG00000153922.10 | *CHD1* | 4.09E-06 | 1.19E-03 |
| ENSG00000106333.12 | *PCOLCE* | 4.47E-06 | 1.28E-03 |
| ENSG00000063180.8 | *CA11* | 5.05E-06 | 1.43E-03 |
| ENSG00000143590.13 | *EFNA3* | 5.41E-06 | 1.51E-03 |
| ENSG00000215375.6 | *MYL5* | 5.97E-06 | 1.65E-03 |
| ENSG00000131355.14 | *ADGRE3* | 6.06E-06 | 1.65E-03 |
| ENSG00000166250.11 | *CLMP* | 6.16E-06 | 1.66E-03 |

| | | | |
|---|---|---|---|
| ENSG00000121691.4 | *CAT* | 7.14E-06 | 1.90E-03 |
| ENSG00000146839.18 | *ZAN* | 7.54E-06 | 1.98E-03 |
| ENSG00000105552.14 | *BCAT2* | 7.74E-06 | 2.01E-03 |
| ENSG00000137310.11 | *TCF19* | 1.12E-05 | 2.88E-03 |
| ENSG00000169241.17 | *SLC50A1* | 1.29E-05 | 3.27E-03 |
| ENSG00000143537.13 | *ADAM15* | 1.36E-05 | 3.41E-03 |
| ENSG00000204540.10 | *PSORS1C1* | 1.95E-05 | 4.84E-03 |
| ENSG00000148291.9 | *SURF2* | 2.36E-05 | 5.79E-03 |
| ENSG00000170498.8 | *KISS1* | 2.87E-05 | 6.97E-03 |
| ENSG00000143627.17 | *PKLR* | 3.00E-05 | 7.21E-03 |
| ENSG00000105523.3 | *FAM83E* | 3.04E-05 | 7.22E-03 |
| ENSG00000012223.12 | *LTF* | 3.32E-05 | 7.81E-03 |
| ENSG00000241186.9 | *TDGF1* | 3.50E-05 | 8.14E-03 |
| ENSG00000131398.13 | *KCNC3* | 4.64E-05 | 1.07E-02 |
| ENSG00000112787.12 | *FBRSL1* | 4.84E-05 | 1.10E-02 |
| ENSG00000125503.12 | *PPP1R12C* | 6.21E-05 | 1.40E-02 |
| ENSG00000101443.17 | *WFDC2* | 7.09E-05 | 1.57E-02 |
| ENSG00000106018.13 | *VIPR2* | 7.11E-05 | 1.57E-02 |
| ENSG00000166508.17 | *MCM7* | 7.15E-05 | 1.57E-02 |
| ENSG00000242252.1 | *BGLAP* | 7.54E-05 | 1.64E-02 |
| ENSG00000108370.16 | *RGS9* | 7.87E-05 | 1.69E-02 |
| ENSG00000146830.9 | *GIGYF1* | 8.74E-05 | 1.86E-02 |
| ENSG00000109111.14 | *SUPT6H* | 9.62E-05 | 2.03E-02 |
| ENSG00000277186.1 | *RP13-554M15.8* | 1.06E-04 | 2.22E-02 |
| ENSG00000106952.7 | *TNFSF8* | 1.08E-04 | 2.23E-02 |
| ENSG00000204248.10 | *COL11A2* | 1.10E-04 | 2.26E-02 |
| ENSG00000105610.4 | *KLF1* | 1.26E-04 | 2.55E-02 |
| ENSG00000161847.13 | *RAVER1* | 1.27E-04 | 2.55E-02 |
| ENSG00000272892.1 | *RP11-57G10.8* | 1.30E-04 | 2.59E-02 |
| ENSG00000167491.17 | *GATAD2A* | 1.33E-04 | 2.60E-02 |
| ENSG00000164683.16 | *HEY1* | 1.33E-04 | 2.60E-02 |
| ENSG00000243364.7 | *EFNA4* | 1.36E-04 | 2.64E-02 |
| ENSG00000164398.12 | *ACSL6* | 1.41E-04 | 2.70E-02 |
| ENSG00000076662.9 | *ICAM3* | 1.41E-04 | 2.70E-02 |
| ENSG00000126351.12 | *THRA* | 1.42E-04 | 2.70E-02 |
| ENSG00000113140.10 | *SPARC* | 1.46E-04 | 2.72E-02 |
| ENSG00000103148.15 | *NPRL3* | 1.47E-04 | 2.72E-02 |
| ENSG00000131037.14 | *EPS8L1* | 1.47E-04 | 2.72E-02 |
| ENSG00000238012.1 | *AC114752.1* | 1.50E-04 | 2.74E-02 |
| ENSG00000229891.1 | *LINC01315* | 1.51E-04 | 2.74E-02 |
| ENSG00000176715.15 | *ACSF3* | 1.52E-04 | 2.74E-02 |
| ENSG00000074842.7 | *MYDGF* | 1.56E-04 | 2.78E-02 |
| ENSG00000278463.1 | *HIST1H2AB* | 1.57E-04 | 2.78E-02 |
| ENSG00000140262.17 | *TCF12* | 1.62E-04 | 2.85E-02 |
| ENSG00000130427.2 | *EPO* | 1.63E-04 | 2.85E-02 |
| ENSG00000235568.6 | *NFAM1* | 1.68E-04 | 2.92E-02 |

| | | | |
|---|---|---|---|
| ENSG00000172057.9 | *ORMDL3* | 1.75E-04 | 3.00E-02 |
| ENSG00000197343.10 | *ZNF655* | 1.76E-04 | 3.01E-02 |
| ENSG00000086570.12 | *FAT2* | 1.80E-04 | 3.04E-02 |
| ENSG00000104388.14 | *RAB2A* | 1.83E-04 | 3.07E-02 |
| ENSG00000172354.9 | *GNB2* | 1.90E-04 | 3.17E-02 |
| ENSG00000034713.7 | *GABARAPL2* | 1.94E-04 | 3.22E-02 |
| ENSG00000131323.14 | *TRAF3* | 1.98E-04 | 3.25E-02 |
| ENSG00000146463.11 | *ZMYM4* | 2.01E-04 | 3.27E-02 |
| ENSG00000171885.13 | *AQP4* | 2.06E-04 | 3.33E-02 |
| ENSG00000135374.9 | *ELF5* | 2.11E-04 | 3.38E-02 |
| ENSG00000134917.9 | *ADAMTS8* | 2.12E-04 | 3.38E-02 |
| ENSG00000132676.15 | *DAP3* | 2.25E-04 | 3.54E-02 |
| ENSG00000220201.7 | *ZGLP1* | 2.25E-04 | 3.54E-02 |
| ENSG00000167123.18 | *CERCAM* | 2.44E-04 | 3.80E-02 |
| ENSG00000106261.16 | *ZKSCAN1* | 2.49E-04 | 3.86E-02 |
| ENSG00000226846.1 | *LINC00348* | 2.63E-04 | 4.05E-02 |
| ENSG00000226676.1 | *RP11-589B3.6* | 2.66E-04 | 4.06E-02 |
| ENSG00000151461.19 | *UPF2* | 2.69E-04 | 4.08E-02 |
| ENSG00000276410.3 | *HIST1H2BB* | 2.79E-04 | 4.20E-02 |
| ENSG00000251396.6 | *LINC01301* | 2.80E-04 | 4.20E-02 |
| ENSG00000105607.12 | *GCDH* | 2.93E-04 | 4.37E-02 |
| ENSG00000124126.13 | *PREX1* | 3.10E-04 | 4.58E-02 |
| ENSG00000180305.4 | *WFDC10A* | 3.15E-04 | 4.63E-02 |
| ENSG00000186075.12 | *ZPBP2* | 3.26E-04 | 4.75E-02 |
| ENSG00000204542.2 | *C6orf15* | 3.37E-04 | 4.85E-02 |
| ENSG00000197935.6 | *ZNF311* | 3.37E-04 | 4.85E-02 |
| ENSG00000267100.1 | *ILF3-AS1* | 3.40E-04 | 4.87E-02 |
| ENSG00000185303.16 | *SFTPA2* | 3.43E-04 | 4.87E-02 |
| ENSG00000184203.7 | *PPP1R2* | 3.48E-04 | 4.93E-02 |

**Supplementary Table S7. The 67 common risk genes associated with three COVID-19 outcomes identified by S-MultiXcan analysis based on 49 GTEx tissues**

| **Gene name** | **Susceptible COVID-19 (P-value)** | **Susceptible COVID-19 (FDR)** | **Hospitalized COVID-19 (P-value)** | **Hospitalized COVID-19 (FDR)** | **Very severe COVID-19 (P-value)** | **Very severe COVID-19 (FDR)** |
|---|---|---|---|---|---|---|
| *ABO* | 2.37E-82 | 2.65E-78 | 9.66E-23 | 1.27E-19 | 4.92E-13 | 4.07E-10 |
| *ACSF3* | 1.52E-04 | 2.74E-02 | 7.10E-07 | 1.96E-04 | 2.03E-09 | 9.86E-07 |
| *ACSL6* | 1.41E-04 | 2.70E-02 | 4.66E-06 | 1.05E-03 | 1.44E-06 | 3.70E-04 |
| *ADAM15* | 1.36E-05 | 3.41E-03 | 9.88E-12 | 7.12E-09 | 5.20E-13 | 4.15E-10 |
| *AP000569.9* | 2.41E-07 | 9.79E-05 | 5.59E-21 | 6.57E-18 | 2.50E-17 | 2.79E-14 |
| *ATP5O* | 7.14E-07 | 2.57E-04 | 4.95E-18 | 5.53E-15 | 2.45E-14 | 2.28E-11 |
| *C6orf15* | 3.37E-04 | 4.85E-02 | 6.91E-11 | 4.41E-08 | 2.56E-13 | 2.20E-10 |
| *CAT* | 7.14E-06 | 1.90E-03 | 3.20E-14 | 2.75E-11 | 9.25E-12 | 6.67E-09 |
| *CCHCR1* | 3.53E-08 | 1.71E-05 | 1.38E-10 | 7.71E-08 | 3.86E-11 | 2.54E-08 |

| | | | | | | |
|---|---|---|---|---|---|---|
| *CCR1* | 7.72E-16 | 9.58E-13 | 5.25E-46 | 1.30E-42 | 2.85E-50 | 7.96E-47 |
| *CCR2* | 9.53E-11 | 6.26E-08 | 5.43E-38 | 9.33E-35 | 8.23E-45 | 1.67E-41 |
| *CCR3* | 5.44E-16 | 7.15E-13 | 6.93E-57 | 2.58E-53 | 7.31E-62 | 2.72E-58 |
| *CCR5* | 7.64E-11 | 5.33E-08 | 7.76E-39 | 1.44E-35 | 9.97E-47 | 2.23E-43 |
| *CCR9* | 5.27E-76 | 3.92E-72 | 3.04E-229 | 6.79E-225 | 6.50E-220 | 7.26E-216 |
| *CCRL2* | 1.83E-08 | 9.97E-06 | 5.12E-35 | 8.17E-32 | 7.73E-35 | 1.23E-31 |
| *CSF3* | 2.46E-08 | 1.28E-05 | 1.42E-04 | 1.80E-02 | 5.78E-05 | 8.67E-03 |
| *CXCR6* | 2.02E-36 | 1.13E-32 | 7.53E-114 | 5.61E-110 | 2.46E-118 | 1.83E-114 |
| *DPP9* | 2.27E-23 | 4.61E-20 | 1.11E-41 | 2.48E-38 | 5.70E-51 | 1.82E-47 |
| *EFNA1* | 3.93E-06 | 1.16E-03 | 4.94E-04 | 4.23E-02 | 2.52E-05 | 4.42E-03 |
| *EFNA3* | 5.41E-06 | 1.51E-03 | 2.67E-04 | 2.74E-02 | 1.01E-04 | 1.32E-02 |
| *EFNA4* | 1.36E-04 | 2.64E-02 | 4.83E-07 | 1.40E-04 | 5.09E-12 | 3.79E-09 |
| *ELF5* | 2.11E-04 | 3.38E-02 | 1.81E-14 | 1.62E-11 | 1.30E-09 | 6.75E-07 |
| *FBRSL1* | 4.84E-05 | 1.10E-02 | 2.24E-11 | 1.47E-08 | 7.15E-09 | 2.85E-06 |
| *FDX2* | 1.71E-07 | 7.21E-05 | 1.21E-10 | 6.93E-08 | 2.37E-09 | 1.08E-06 |
| *FOXP4* | 6.10E-13 | 5.68E-10 | 5.15E-26 | 7.67E-23 | 2.39E-17 | 2.79E-14 |
| *FUT2* | 2.50E-15 | 2.79E-12 | 1.42E-09 | 6.34E-07 | 9.04E-10 | 4.93E-07 |
| *FYCO1* | 4.83E-20 | 8.99E-17 | 2.54E-53 | 8.11E-50 | 2.62E-41 | 4.50E-38 |
| *GSDMA* | 1.45E-07 | 6.35E-05 | 4.83E-04 | 4.18E-02 | 4.52E-04 | 4.37E-02 |
| *HLA-C* | 2.34E-07 | 9.68E-05 | 9.51E-10 | 4.58E-07 | 6.13E-10 | 3.42E-07 |
| *ICAM3* | 1.41E-04 | 2.70E-02 | 4.54E-09 | 1.81E-06 | 1.41E-07 | 4.56E-05 |
| *IFNAR2* | 9.43E-16 | 1.11E-12 | 1.67E-50 | 4.66E-47 | 2.05E-47 | 5.09E-44 |
| *IL10RB* | 1.31E-13 | 1.33E-10 | 4.82E-41 | 9.79E-38 | 4.16E-42 | 7.74E-39 |
| *KCNC3* | 4.64E-05 | 1.07E-02 | 4.73E-09 | 1.82E-06 | 6.25E-06 | 1.32E-03 |
| *KRTCAP2* | 1.51E-08 | 8.43E-06 | 7.93E-11 | 4.92E-08 | 3.02E-09 | 1.32E-06 |
| *LINC01301* | 2.80E-04 | 4.20E-02 | 4.93E-08 | 1.67E-05 | 5.82E-07 | 1.63E-04 |
| *LTF* | 3.32E-05 | 7.81E-03 | 5.44E-14 | 4.50E-11 | 6.51E-14 | 5.82E-11 |
| *LZTFL1* | 1.59E-35 | 7.10E-32 | 5.65E-100 | 3.16E-96 | 1.64E-101 | 7.33E-98 |
| *MAMSTR* | 4.55E-11 | 3.28E-08 | 1.89E-04 | 2.16E-02 | 1.60E-08 | 5.86E-06 |
| *MCM7* | 7.15E-05 | 1.57E-02 | 3.18E-06 | 7.25E-04 | 7.28E-07 | 1.98E-04 |
| *MED24* | 1.16E-07 | 5.18E-05 | 1.10E-04 | 1.47E-02 | 6.91E-05 | 9.77E-03 |
| *MRPS6* | 3.42E-06 | 1.03E-03 | 2.59E-16 | 2.52E-13 | 3.52E-11 | 2.38E-08 |
| *MTX1* | 6.21E-08 | 2.83E-05 | 4.60E-22 | 5.71E-19 | 1.52E-27 | 2.26E-24 |
| *MUC1* | 8.53E-11 | 5.77E-08 | 2.21E-24 | 3.09E-21 | 5.20E-25 | 7.26E-22 |
| *MYDGF* | 1.56E-04 | 2.78E-02 | 2.26E-06 | 5.26E-04 | 1.86E-06 | 4.54E-04 |
| *NAPSA* | 3.19E-08 | 1.58E-05 | 4.33E-17 | 4.40E-14 | 2.36E-10 | 1.39E-07 |
| *NFKBIZ* | 2.22E-29 | 7.08E-26 | 6.81E-13 | 5.43E-10 | 1.08E-05 | 2.15E-03 |
| *NTN5* | 1.15E-12 | 9.88E-10 | 1.07E-06 | 2.81E-04 | 1.63E-08 | 5.87E-06 |
| *OAS1* | 2.11E-06 | 6.93E-04 | 9.67E-08 | 3.18E-05 | 4.10E-09 | 1.73E-06 |
| *OAS3* | 4.69E-08 | 2.23E-05 | 9.33E-11 | 5.63E-08 | 1.24E-07 | 4.07E-05 |
| *PLEKHA4* | 7.47E-13 | 6.67E-10 | 5.67E-04 | 4.64E-02 | 4.08E-04 | 4.09E-02 |
| *PSMD3* | 1.65E-07 | 7.09E-05 | 1.45E-05 | 2.70E-03 | 4.86E-06 | 1.05E-03 |
| *PSORS1C1* | 1.95E-05 | 4.84E-03 | 1.39E-05 | 2.63E-03 | 3.12E-07 | 9.17E-05 |
| *PSORS1C2* | 8.02E-07 | 2.80E-04 | 3.28E-08 | 1.16E-05 | 1.07E-09 | 5.69E-07 |
| *RAB2A* | 1.83E-04 | 3.07E-02 | 5.91E-08 | 1.97E-05 | 2.78E-07 | 8.28E-05 |
| *RASIP1* | 3.09E-11 | 2.38E-08 | 4.51E-05 | 7.20E-03 | 1.11E-08 | 4.28E-06 |

| | | | | | | |
|---|---|---|---|---|---|---|
| *RAVER1* | 1.27E-04 | 2.55E-02 | 2.81E-09 | 1.16E-06 | 5.26E-07 | 1.51E-04 |
| *RP11-589B3.6* | 2.66E-04 | 4.06E-02 | 2.69E-07 | 8.12E-05 | 4.88E-08 | 1.65E-05 |
| *SFTPA2* | 3.43E-04 | 4.87E-02 | 4.27E-08 | 1.49E-05 | 4.29E-08 | 1.47E-05 |
| *SFTPD* | 9.84E-10 | 6.11E-07 | 3.46E-10 | 1.89E-07 | 1.94E-09 | 9.63E-07 |
| *SLC50A1* | 1.29E-05 | 3.27E-03 | 2.12E-11 | 1.44E-08 | 1.78E-14 | 1.73E-11 |
| *SLC6A20* | 1.92E-171 | 4.29E-167 | 2.30E-217 | 2.57E-213 | 7.16E-227 | 1.60E-222 |
| *TCF19* | 1.12E-05 | 2.88E-03 | 4.83E-10 | 2.51E-07 | 2.17E-09 | 1.01E-06 |
| *TNFAIP8L1* | 3.48E-06 | 1.04E-03 | 6.77E-09 | 2.52E-06 | 1.00E-12 | 7.70E-10 |
| *TRIM46* | 6.17E-09 | 3.53E-06 | 8.52E-12 | 6.34E-09 | 1.68E-19 | 2.08E-16 |
| *XCR1* | 8.96E-30 | 3.34E-26 | 1.53E-86 | 6.84E-83 | 1.81E-112 | 1.01E-108 |
| *ZGLP1* | 2.25E-04 | 3.54E-02 | 3.84E-10 | 2.04E-07 | 5.36E-09 | 2.22E-06 |
| *ZKSCAN1* | 2.49E-04 | 3.86E-02 | 2.26E-06 | 5.26E-04 | 2.40E-06 | 5.53E-04 |

**Supplementary Table S8. The S-MultiXcan-identified 438 risk genes validated by S-PrediXcan analysis based on GTEx lung tissue**

| Gene Name | Susceptible COVID-19 (P-value) | Susceptible COVID-19 (FDR) | Hospitalized COVID-19 (P-value) | Hospitalized COVID-19 (FDR) | Very severe COVID-19 (P-value) | Very severe COVID-19 (FDR) |
|---|---|---|---|---|---|---|
| *CCR9* | NA | NA | NA | NA | NA | NA |
| *SLC6A20* | 2.21E-17 | 8.06E-14 | 8.23E-08 | 6.67E-05 | 9.32E-08 | 7.77E-05 |
| *CXCR6* | 1.10E-16 | 3.21E-13 | 1.67E-83 | 2.44E-79 | 1.13E-87 | 1.65E-83 |
| *XCR1* | 4.61E-02 | 5.81E-01 | 3.06E-01 | 8.56E-01 | 7.59E-02 | 6.42E-01 |
| *LZTFL1* | 5.40E-02 | 6.05E-01 | 4.32E-01 | 9.02E-01 | 7.16E-01 | 9.68E-01 |
| *ABO* | 3.36E-52 | 4.90E-48 | 1.58E-14 | 2.88E-11 | 3.33E-05 | 1.24E-02 |
| *CCR3* | 5.47E-03 | 3.38E-01 | 1.94E-10 | 2.57E-07 | 8.87E-11 | 1.26E-07 |
| *FYCO1* | 2.72E-02 | 5.16E-01 | 2.39E-19 | 8.71E-16 | 8.27E-22 | 4.02E-18 |
| *DPP9* | 9.08E-02 | 6.99E-01 | 3.54E-01 | 8.76E-01 | 2.95E-02 | 5.29E-01 |
| *IFNAR2* | 1.16E-03 | 1.75E-01 | 1.93E-06 | 1.08E-03 | 1.94E-06 | 1.09E-03 |
| *CCR1* | 4.69E-01 | 9.25E-01 | 7.51E-03 | 2.98E-01 | 3.09E-04 | 6.09E-02 |
| *CCR5* | 2.00E-17 | 8.06E-14 | 2.66E-43 | 1.94E-39 | 3.69E-50 | 2.69E-46 |
| *CCR2* | 3.12E-04 | 6.81E-02 | 3.55E-02 | 5.22E-01 | 6.00E-02 | 6.26E-01 |
| *IL10RB* | 4.25E-09 | 5.63E-06 | 2.30E-18 | 6.68E-15 | 1.32E-18 | 3.85E-15 |
| *CCRL2* | 3.93E-02 | 5.58E-01 | 1.57E-04 | 3.42E-02 | 3.79E-05 | 1.38E-02 |
| *NFKBIZ* | NA | NA | NA | NA | NA | NA |
| *NXPE3* | 1.89E-12 | 3.06E-09 | 1.56E-03 | 1.45E-01 | 4.26E-03 | 2.82E-01 |
| *MTX1* | 4.72E-08 | 4.05E-05 | 8.75E-01 | 9.88E-01 | 6.63E-04 | 1.04E-01 |
| *CEP97* | 9.42E-28 | 6.87E-24 | 1.25E-11 | 1.82E-08 | 5.66E-05 | 1.79E-02 |
| *FOXP4* | 2.17E-15 | 4.52E-12 | 5.44E-30 | 2.64E-26 | 1.67E-19 | 6.09E-16 |
| *MUC1* | 7.36E-05 | 2.16E-02 | 6.18E-03 | 2.73E-01 | 1.60E-09 | 1.94E-06 |
| *ZBTB11* | 1.46E-08 | 1.64E-05 | 5.78E-04 | 8.43E-02 | 3.51E-02 | 5.42E-01 |
| *ASH1L* | NA | NA | NA | NA | NA | NA |
| *AP000569.9* | NA | NA | NA | NA | NA | NA |
| *CACFD1* | 2.05E-04 | 5.06E-02 | 7.67E-01 | 9.75E-01 | 9.60E-01 | 9.96E-01 |
| *TRIM46* | NA | NA | NA | NA | NA | NA |
| *ATP5O* | 3.09E-05 | 1.10E-02 | 2.75E-18 | 6.68E-15 | 4.07E-13 | 9.89E-10 |

| | | | | | | |
|---|---|---|---|---|---|---|
| *NAPSA* | 8.14E-01 | 9.86E-01 | 5.30E-02 | 5.69E-01 | 8.50E-01 | 9.87E-01 |
| *TULP2* | 1.18E-13 | 2.15E-10 | 3.48E-04 | 5.76E-02 | 1.03E-03 | 1.40E-01 |
| *PCNP* | 2.13E-06 | 1.00E-03 | 6.77E-02 | 6.13E-01 | 2.15E-01 | 7.94E-01 |
| *MRPS6* | 1.40E-01 | 7.50E-01 | 9.26E-02 | 6.58E-01 | 5.49E-01 | 9.34E-01 |
| *SENP7* | 1.45E-06 | 7.56E-04 | 7.25E-03 | 2.95E-01 | 4.40E-01 | 8.98E-01 |
| *LINC02009* | NA | NA | NA | NA | NA | NA |
| *TYK2* | 3.70E-03 | 2.94E-01 | 8.75E-05 | 2.16E-02 | 2.80E-08 | 2.92E-05 |
| *FUT2* | 1.35E-16 | 3.28E-13 | 3.79E-10 | 4.25E-07 | 9.53E-11 | 1.26E-07 |
| *MED22* | 2.60E-04 | 6.02E-02 | 1.80E-02 | 4.21E-01 | 1.74E-02 | 4.65E-01 |
| *SLC50A1* | 2.07E-08 | 2.16E-05 | 7.27E-02 | 6.24E-01 | 8.21E-02 | 6.57E-01 |
| *ELF5* | 1.12E-05 | 4.46E-03 | 2.83E-15 | 5.90E-12 | 5.87E-11 | 9.51E-08 |
| *CAT* | 2.75E-03 | 2.57E-01 | 5.36E-05 | 1.56E-02 | 5.38E-05 | 1.74E-02 |
| *LTF* | 7.80E-01 | 9.82E-01 | 8.97E-01 | 9.89E-01 | 3.43E-01 | 8.69E-01 |
| *C6orf15* | NA | NA | NA | NA | NA | NA |
| *SLC2A6* | NA | NA | NA | NA | NA | NA |
| *ADAM15* | 3.78E-01 | 8.95E-01 | 1.83E-04 | 3.87E-02 | 8.13E-08 | 7.41E-05 |
| *PLEKHA4* | 1.57E-07 | 1.09E-04 | 1.69E-02 | 4.10E-01 | 2.64E-02 | 5.23E-01 |
| *TNFAIP8L1* | 5.69E-04 | 1.04E-01 | 4.44E-03 | 2.25E-01 | 4.11E-02 | 5.65E-01 |
| *NTN5* | 1.13E-05 | 4.46E-03 | 2.20E-07 | 1.53E-04 | 6.31E-06 | 2.97E-03 |
| *ATP11A* | 4.23E-01 | 9.09E-01 | 5.65E-01 | 9.36E-01 | 2.08E-01 | 7.89E-01 |
| *ADAMTS13* | 3.60E-02 | 5.45E-01 | 2.53E-01 | 8.26E-01 | 5.18E-01 | 9.22E-01 |
| *EFNA4* | NA | NA | NA | NA | NA | NA |
| *RP11-798G7.6* | NA | NA | NA | NA | NA | NA |
| *PPP1R15A* | 2.76E-08 | 2.68E-05 | 9.51E-03 | 3.24E-01 | 1.83E-02 | 4.71E-01 |
| *GON4L* | NA | NA | NA | NA | NA | NA |
| *FBRSL1* | 9.01E-01 | 9.93E-01 | 7.38E-02 | 6.26E-01 | 2.63E-02 | 5.23E-01 |
| *RASIP1* | 4.63E-09 | 5.63E-06 | 6.31E-04 | 8.85E-02 | 6.33E-08 | 6.15E-05 |
| *CCHCR1* | 4.84E-05 | 1.60E-02 | 1.90E-07 | 1.39E-04 | 1.39E-04 | 3.55E-02 |
| *NUCB1* | 1.22E-09 | 1.78E-06 | 7.73E-03 | 3.02E-01 | 1.03E-02 | 3.88E-01 |
| *MAMSTR* | 1.25E-01 | 7.37E-01 | 1.24E-01 | 7.05E-01 | 8.01E-03 | 3.70E-01 |
| *KRTCAP2* | NA | NA | NA | NA | NA | NA |
| *OAS3* | 2.10E-01 | 8.20E-01 | 1.70E-02 | 4.11E-01 | 7.16E-02 | 6.37E-01 |
| *FDX2* | 1.47E-02 | 4.37E-01 | 1.57E-01 | 7.45E-01 | 2.92E-02 | 5.29E-01 |
| *PDE4A* | 7.52E-01 | 9.77E-01 | 4.66E-01 | 9.13E-01 | 9.56E-01 | 9.95E-01 |
| *PLEKHM1* | NA | NA | NA | NA | NA | NA |
| *FUT1* | 1.21E-02 | 4.22E-01 | 2.55E-01 | 8.30E-01 | 5.75E-02 | 6.15E-01 |
| *SFTPD* | 3.38E-03 | 2.90E-01 | 1.30E-02 | 3.62E-01 | 5.25E-01 | 9.24E-01 |
| *ZGLP1* | 4.78E-03 | 3.27E-01 | 1.58E-03 | 1.45E-01 | 2.85E-02 | 5.28E-01 |
| *TCF19* | 9.72E-07 | 5.25E-04 | 9.44E-08 | 7.25E-05 | 3.90E-07 | 2.47E-04 |
| *C5orf56* | 8.38E-02 | 6.82E-01 | 2.09E-01 | 7.97E-01 | 1.35E-01 | 7.27E-01 |
| *HLA-C* | 7.54E-04 | 1.22E-01 | 2.29E-05 | 7.95E-03 | 6.04E-05 | 1.81E-02 |
| *ARHGAP27* | 2.69E-03 | 2.57E-01 | 2.26E-10 | 2.75E-07 | 3.57E-11 | 6.51E-08 |
| *SLC35B1* | 5.67E-01 | 9.48E-01 | 8.28E-01 | 9.85E-01 | 6.12E-01 | 9.46E-01 |
| *DDX39B* | 7.36E-01 | 9.75E-01 | 1.27E-02 | 3.60E-01 | 3.50E-01 | 8.71E-01 |
| *PSORS1C2* | 3.97E-06 | 1.81E-03 | 4.99E-05 | 1.55E-02 | 7.33E-06 | 3.33E-03 |
| *IZUMO1* | 2.38E-03 | 2.50E-01 | 3.00E-01 | 8.51E-01 | 2.41E-02 | 5.06E-01 |

| | | | | | | |
|---|---|---|---|---|---|---|
| *FAM215B* | 3.11E-01 | 8.72E-01 | 5.45E-05 | 1.56E-02 | 1.30E-02 | 4.17E-01 |
| *REXO4* | 5.24E-03 | 3.35E-01 | 7.37E-01 | 9.72E-01 | 3.59E-01 | 8.75E-01 |
| *ACSF3* | 5.40E-01 | 9.43E-01 | 3.25E-02 | 5.11E-01 | 2.84E-04 | 5.73E-02 |
| *RAVER1* | 5.66E-05 | 1.76E-02 | 2.66E-05 | 9.02E-03 | 5.66E-04 | 9.86E-02 |
| *ICAM5* | 1.59E-01 | 7.79E-01 | 1.56E-01 | 7.42E-01 | 3.14E-03 | 2.41E-01 |
| *OAS1* | 3.07E-07 | 2.04E-04 | 8.22E-09 | 7.49E-06 | 4.75E-06 | 2.39E-03 |
| *ICAM3* | 1.11E-01 | 7.22E-01 | 3.81E-02 | 5.29E-01 | 2.37E-02 | 5.02E-01 |
| *ZCCHC4* | 1.10E-05 | 4.46E-03 | 2.27E-08 | 1.95E-05 | 6.09E-05 | 1.81E-02 |
| *KCNC3* | 1.39E-02 | 4.36E-01 | 8.91E-03 | 3.18E-01 | 2.79E-01 | 8.43E-01 |
| *NPNT* | 4.86E-01 | 9.32E-01 | 1.31E-02 | 3.63E-01 | 1.15E-02 | 4.00E-01 |
| *WNT3* | 1.09E-02 | 4.10E-01 | 1.81E-12 | 2.93E-09 | 1.25E-12 | 2.60E-09 |
| *DCST2* | 4.45E-03 | 3.16E-01 | 1.58E-01 | 7.46E-01 | 4.74E-01 | 9.09E-01 |
| *HLA-A* | 6.16E-02 | 6.27E-01 | 8.76E-01 | 9.88E-01 | 1.79E-02 | 4.67E-01 |
| *HSD17B14* | 1.28E-07 | 9.63E-05 | 1.58E-03 | 1.45E-01 | 2.10E-03 | 1.96E-01 |
| *PI4K2B* | 1.32E-01 | 7.40E-01 | 1.12E-01 | 6.90E-01 | 5.87E-03 | 3.27E-01 |
| *CSF3* | 7.39E-08 | 5.99E-05 | 9.81E-06 | 4.21E-03 | 3.17E-06 | 1.65E-03 |
| *RPL7A* | 5.43E-01 | 9.44E-01 | 4.86E-01 | 9.19E-01 | 8.02E-01 | 9.80E-01 |
| *SEMA6B* | 1.94E-01 | 8.15E-01 | 2.93E-01 | 8.51E-01 | 3.50E-01 | 8.71E-01 |
| *SFTPA2* | 1.29E-04 | 3.42E-02 | 4.32E-07 | 2.86E-04 | 9.60E-08 | 7.77E-05 |
| *CDH15* | 2.19E-02 | 4.98E-01 | 6.56E-06 | 3.09E-03 | 6.45E-04 | 1.03E-01 |
| *RP11-589B3.6* | 2.00E-02 | 4.80E-01 | 2.40E-03 | 1.82E-01 | 1.07E-06 | 6.50E-04 |
| *LINC01301* | 2.69E-05 | 9.81E-03 | 4.16E-09 | 4.04E-06 | 1.09E-07 | 7.95E-05 |
| *THBS3* | 4.15E-07 | 2.53E-04 | 5.91E-01 | 9.42E-01 | 9.03E-03 | 3.78E-01 |
| *RAB2A* | 2.69E-05 | 9.81E-03 | 4.16E-09 | 4.04E-06 | 1.09E-07 | 7.95E-05 |
| *SACM1L* | 8.29E-01 | 9.86E-01 | 5.70E-01 | 9.38E-01 | 5.25E-02 | 6.04E-01 |
| *IBA57-AS1* | NA | NA | NA | NA | NA | NA |
| *MED24* | 8.86E-02 | 6.96E-01 | 2.17E-01 | 8.02E-01 | 3.63E-01 | 8.75E-01 |
| *DPF2* | 2.53E-01 | 8.50E-01 | 4.97E-04 | 7.54E-02 | 7.27E-01 | 9.70E-01 |
| *GSDMA* | 1.32E-07 | 9.63E-05 | 6.28E-06 | 3.05E-03 | 5.70E-06 | 2.77E-03 |
| *PMVK* | 3.47E-02 | 5.39E-01 | 3.96E-01 | 8.92E-01 | 5.82E-02 | 6.16E-01 |
| *PSMD3* | 7.33E-01 | 9.75E-01 | 3.60E-01 | 8.77E-01 | 8.42E-01 | 9.85E-01 |
| *RP11-506M13.3* | NA | NA | NA | NA | NA | NA |
| *NUTM2E* | 9.64E-01 | 9.98E-01 | 1.30E-01 | 7.10E-01 | 7.74E-04 | 1.18E-01 |
| *CSK* | 1.73E-01 | 7.98E-01 | 3.25E-01 | 8.64E-01 | 5.60E-01 | 9.36E-01 |
| *RP4-535B20.1* | NA | NA | NA | NA | NA | NA |
| *MICB* | 1.23E-01 | 7.37E-01 | 4.24E-01 | 8.98E-01 | 1.96E-01 | 7.83E-01 |
| *RTP3* | NA | NA | NA | NA | NA | NA |
| *RP11-387H17.6* | 1.73E-06 | 8.66E-04 | 3.51E-03 | 2.11E-01 | 7.06E-03 | 3.49E-01 |
| *ARL17A* | 1.01E-01 | 7.15E-01 | 4.54E-02 | 5.55E-01 | 1.10E-01 | 6.96E-01 |
| *HLA-DPA1* | 3.98E-01 | 9.00E-01 | 8.19E-01 | 9.82E-01 | 7.82E-01 | 9.78E-01 |
| *PSORS1C1* | 4.59E-02 | 5.81E-01 | 4.48E-01 | 9.08E-01 | 4.10E-01 | 8.89E-01 |
| *RP3-462D8.2* | NA | NA | NA | NA | NA | NA |
| *MEF2D* | NA | NA | NA | NA | NA | NA |

| | | | | | | |
|---|---|---|---|---|---|---|
| *ICAM1* | NA | NA | NA | NA | NA | NA |
| *RP11-11C20.3* | 3.29E-01 | 8.81E-01 | 2.82E-02 | 4.90E-01 | 1.09E-04 | 2.90E-02 |
| *RPL24* | 5.96E-07 | 3.40E-04 | 1.32E-04 | 2.96E-02 | 1.45E-03 | 1.69E-01 |
| *SLC22A4* | 8.96E-01 | 9.93E-01 | 4.65E-01 | 9.13E-01 | 8.97E-01 | 9.93E-01 |
| *GBA* | 1.28E-01 | 7.37E-01 | 2.14E-01 | 8.01E-01 | 4.98E-01 | 9.17E-01 |
| *MCM7* | 5.69E-01 | 9.49E-01 | 5.16E-01 | 9.26E-01 | 2.32E-01 | 8.07E-01 |
| *COQ10B* | 7.72E-01 | 9.81E-01 | 4.52E-03 | 2.25E-01 | 1.50E-01 | 7.49E-01 |
| *LTA* | NA | NA | NA | NA | NA | NA |
| *IMPG2* | 2.06E-01 | 8.18E-01 | 4.68E-01 | 9.14E-01 | 4.08E-01 | 8.88E-01 |
| *LRRC37A2* | 2.56E-01 | 8.53E-01 | 2.06E-05 | 7.51E-03 | 3.14E-03 | 2.41E-01 |
| *DBP* | NA | NA | NA | NA | NA | NA |
| *CLK3* | 5.33E-01 | 9.41E-01 | 7.08E-02 | 6.23E-01 | 7.29E-01 | 9.71E-01 |
| *ZNF3* | 9.16E-02 | 7.02E-01 | 9.15E-01 | 9.91E-01 | 6.86E-01 | 9.60E-01 |
| *ZNF778* | 1.82E-02 | 4.60E-01 | 6.47E-07 | 3.93E-04 | 7.95E-05 | 2.19E-02 |
| *PPP1R11* | 7.72E-02 | 6.74E-01 | 8.88E-03 | 3.18E-01 | 6.01E-05 | 1.81E-02 |
| *SLC5A3* | 6.92E-01 | 9.70E-01 | 7.57E-02 | 6.28E-01 | 1.79E-01 | 7.70E-01 |
| *CDSN* | NA | NA | NA | NA | NA | NA |
| *ACSL6* | 3.69E-03 | 2.94E-01 | 7.39E-05 | 2.00E-02 | 1.12E-02 | 3.99E-01 |
| *PGC* | 1.74E-01 | 7.98E-01 | 9.77E-02 | 6.63E-01 | 1.18E-01 | 7.05E-01 |
| *DSP* | 4.95E-02 | 5.92E-01 | 1.93E-03 | 1.63E-01 | 1.47E-07 | 1.02E-04 |
| *HLA-DQA2* | 8.05E-02 | 6.80E-01 | 2.79E-03 | 1.94E-01 | 1.83E-04 | 4.23E-02 |
| *IFNAR1* | 1.22E-01 | 7.37E-01 | 3.12E-05 | 1.01E-02 | 8.91E-06 | 3.82E-03 |
| *FOXA3* | 5.61E-01 | 9.48E-01 | 3.14E-03 | 2.00E-01 | 4.87E-02 | 5.92E-01 |
| *HLA-DPB1* | 8.22E-01 | 9.86E-01 | 3.13E-02 | 5.05E-01 | 1.84E-01 | 7.73E-01 |
| *DTX1* | 2.04E-01 | 8.18E-01 | 1.17E-01 | 6.97E-01 | 1.04E-02 | 3.90E-01 |
| *MYDGF* | 6.15E-02 | 6.27E-01 | 2.51E-01 | 8.26E-01 | 4.05E-01 | 8.87E-01 |
| *RP11-182L21.6* | 1.99E-01 | 8.15E-01 | 3.77E-02 | 5.29E-01 | 5.58E-01 | 9.36E-01 |
| *EIF4EBP2* | 1.82E-01 | 8.04E-01 | 9.02E-01 | 9.90E-01 | 6.58E-02 | 6.37E-01 |
| *ANAPC4* | 2.80E-02 | 5.16E-01 | 8.04E-05 | 2.02E-02 | 7.44E-04 | 1.14E-01 |
| *SLC22A31* | 4.84E-02 | 5.87E-01 | 2.32E-03 | 1.78E-01 | 3.38E-02 | 5.39E-01 |
| *SPACA4* | NA | NA | NA | NA | NA | NA |
| *TFR2* | 4.16E-07 | 2.53E-04 | 6.52E-04 | 8.89E-02 | 2.28E-03 | 2.07E-01 |
| *HLA-DRA* | 2.54E-02 | 5.10E-01 | 1.08E-01 | 6.83E-01 | 1.04E-01 | 6.86E-01 |
| *ZKSCAN1* | 4.25E-01 | 9.09E-01 | 3.82E-01 | 8.86E-01 | 6.71E-02 | 6.37E-01 |
| *SLC25A44* | 4.25E-06 | 1.88E-03 | 1.70E-03 | 1.49E-01 | 6.14E-04 | 1.02E-01 |
| *PMF1* | 5.69E-06 | 2.44E-03 | 1.46E-02 | 3.83E-01 | 6.69E-04 | 1.04E-01 |
| *MOSPD3* | 6.06E-07 | 3.40E-04 | 7.54E-04 | 9.77E-02 | 2.49E-03 | 2.15E-01 |
| *BYSL* | 7.89E-02 | 6.79E-01 | 2.74E-02 | 4.86E-01 | 6.16E-01 | 9.46E-01 |
| *CTA-384D8.35* | 4.72E-01 | 9.26E-01 | 8.38E-02 | 6.45E-01 | 8.15E-01 | 9.81E-01 |
| *ACTL6B* | NA | NA | NA | NA | NA | NA |
| *HIP1* | 2.75E-01 | 8.64E-01 | 4.68E-03 | 2.27E-01 | 2.13E-04 | 4.70E-02 |
| *EFNA1* | 4.51E-08 | 4.05E-05 | 4.61E-01 | 9.13E-01 | 1.15E-02 | 4.00E-01 |
| *CDC37* | NA | NA | NA | NA | NA | NA |
| *CHD1* | NA | NA | NA | NA | NA | NA |

| | | | | | | |
|---|---|---|---|---|---|---|
| *PCOLCE* | 7.17E-01 | 9.73E-01 | 5.86E-02 | 5.87E-01 | 2.25E-01 | 8.04E-01 |
| *AC006007.1* | NA | NA | NA | NA | NA | NA |
| *CA11* | 4.91E-01 | 9.33E-01 | 3.31E-01 | 8.66E-01 | 4.96E-01 | 9.17E-01 |
| *FUBP1* | 3.13E-03 | 2.78E-01 | 5.33E-07 | 3.38E-04 | 2.44E-06 | 1.32E-03 |
| *SH3BP2* | 6.04E-02 | 6.23E-01 | 1.13E-02 | 3.43E-01 | 1.49E-01 | 7.47E-01 |
| *EFNA3* | NA | NA | NA | NA | NA | NA |
| *C2orf16* | 1.89E-02 | 4.65E-01 | 8.79E-07 | 5.13E-04 | 1.11E-03 | 1.44E-01 |
| *MYL5* | 8.14E-01 | 9.86E-01 | 1.32E-01 | 7.12E-01 | 2.44E-01 | 8.18E-01 |
| *ADGRE3* | 1.99E-01 | 8.15E-01 | 4.16E-01 | 8.98E-01 | 5.84E-01 | 9.41E-01 |
| *CLMP* | 1.28E-02 | 4.25E-01 | 2.02E-03 | 1.67E-01 | 9.59E-04 | 1.34E-01 |
| *HLA-F* | 1.61E-01 | 7.82E-01 | 2.51E-01 | 8.26E-01 | 4.93E-02 | 5.92E-01 |
| *GATS* | NA | NA | NA | NA | NA | NA |
| *FDPS* | 4.65E-01 | 9.23E-01 | 7.69E-01 | 9.75E-01 | 1.92E-02 | 4.77E-01 |
| *ZAN* | NA | NA | NA | NA | NA | NA |
| *TNFRSF4* | 3.25E-01 | 8.79E-01 | 4.01E-01 | 8.94E-01 | 2.51E-01 | 8.23E-01 |
| *BCAT2* | 1.78E-06 | 8.66E-04 | 3.73E-02 | 5.28E-01 | 1.37E-01 | 7.31E-01 |
| *CYP1A1* | 6.41E-04 | 1.08E-01 | 1.10E-05 | 4.46E-03 | 5.25E-02 | 6.04E-01 |
| *TOMM7* | 2.63E-01 | 8.59E-01 | 8.30E-04 | 1.01E-01 | 7.04E-05 | 2.05E-02 |
| *YIPF2* | 4.93E-01 | 9.33E-01 | 6.75E-01 | 9.59E-01 | 2.83E-02 | 5.27E-01 |
| *POU5F1* | 1.76E-01 | 7.98E-01 | 8.27E-01 | 9.85E-01 | 4.97E-02 | 5.92E-01 |
| *P4HA2* | 4.43E-01 | 9.15E-01 | 2.26E-01 | 8.09E-01 | 6.27E-01 | 9.48E-01 |
| *RP11-298J23.8* | NA | NA | NA | NA | NA | NA |
| *DNAJB4* | 1.42E-01 | 7.54E-01 | 6.64E-01 | 9.57E-01 | 2.91E-01 | 8.47E-01 |
| *NLRP4* | NA | NA | NA | NA | NA | NA |
| *FRS3* | 6.88E-02 | 6.55E-01 | 8.67E-01 | 9.88E-01 | 6.44E-01 | 9.51E-01 |
| *GPC2* | NA | NA | NA | NA | NA | NA |
| *AGRN* | 3.18E-01 | 8.75E-01 | 6.21E-01 | 9.46E-01 | 6.18E-01 | 9.46E-01 |
| *COPS6* | 2.35E-01 | 8.37E-01 | 4.42E-01 | 9.05E-01 | 5.95E-02 | 6.22E-01 |
| *ATF6B* | 3.96E-02 | 5.60E-01 | 1.45E-05 | 5.72E-03 | 4.03E-05 | 1.43E-02 |
| *GUK1* | 2.31E-01 | 8.34E-01 | 2.16E-02 | 4.50E-01 | 1.03E-02 | 3.88E-01 |
| *RP11-298J23.10* | 4.66E-01 | 9.23E-01 | 7.02E-01 | 9.66E-01 | 7.71E-01 | 9.77E-01 |
| *CLK2* | 8.23E-01 | 9.86E-01 | 6.25E-03 | 2.73E-01 | 1.69E-05 | 6.66E-03 |
| *RP13-554M15.8* | 1.59E-01 | 7.79E-01 | 3.01E-02 | 4.99E-01 | 4.47E-02 | 5.81E-01 |
| *AZGP1* | NA | NA | NA | NA | NA | NA |
| *HCN3* | 7.96E-01 | 9.86E-01 | 4.73E-03 | 2.27E-01 | 2.30E-05 | 8.82E-03 |
| *SURF2* | 3.77E-01 | 8.95E-01 | 9.68E-01 | 9.96E-01 | 6.54E-01 | 9.51E-01 |
| *JAK1* | 7.98E-03 | 3.82E-01 | 4.34E-06 | 2.18E-03 | 3.02E-07 | 2.00E-04 |
| *SCAMP3* | NA | NA | NA | NA | NA | NA |
| *LINC00304* | 5.47E-01 | 9.45E-01 | 1.38E-03 | 1.36E-01 | 2.45E-04 | 5.26E-02 |
| *ZBTB7B* | NA | NA | NA | NA | NA | NA |
| *ACAD8* | 7.26E-01 | 9.73E-01 | 1.01E-01 | 6.69E-01 | 3.76E-02 | 5.52E-01 |
| *HLA-DMB* | 8.09E-02 | 6.81E-01 | 9.18E-03 | 3.21E-01 | 4.87E-03 | 3.01E-01 |
| *TRIM15* | 5.84E-01 | 9.51E-01 | 3.42E-01 | 8.71E-01 | 3.13E-02 | 5.30E-01 |
| *RNF17* | NA | NA | NA | NA | NA | NA |

| | | | | | | |
|---|---|---|---|---|---|---|
| *KISS1* | 2.66E-01 | 8.61E-01 | 5.95E-02 | 5.89E-01 | 1.21E-02 | 4.03E-01 |
| *ABCF1* | 3.45E-01 | 8.88E-01 | 6.75E-02 | 6.13E-01 | 8.51E-02 | 6.61E-01 |
| *BTNL2* | 7.64E-01 | 9.80E-01 | 7.04E-01 | 9.66E-01 | 8.43E-01 | 9.86E-01 |
| *PKLR* | NA | NA | NA | NA | NA | NA |
| *FAM83E* | 3.79E-04 | 7.67E-02 | 1.43E-02 | 3.78E-01 | 2.25E-02 | 5.00E-01 |
| *TIMM29* | NA | NA | NA | NA | NA | NA |
| *LINC02138* | NA | NA | NA | NA | NA | NA |
| *LTB* | NA | NA | NA | NA | NA | NA |
| *DDR1* | 5.30E-02 | 6.04E-01 | 3.37E-01 | 8.69E-01 | 2.86E-01 | 8.45E-01 |
| *TDGF1* | 2.82E-01 | 8.66E-01 | 3.51E-02 | 5.22E-01 | 8.97E-03 | 3.78E-01 |
| *RSPO2* | 8.60E-01 | 9.89E-01 | 8.87E-01 | 9.88E-01 | 2.35E-01 | 8.11E-01 |
| *SFTPA1* | 1.17E-03 | 1.75E-01 | 3.07E-06 | 1.66E-03 | 6.70E-09 | 7.51E-06 |
| *RIT1* | 2.10E-01 | 8.20E-01 | 6.88E-01 | 9.62E-01 | 9.89E-01 | 9.98E-01 |
| *NUTM2B* | 1.97E-01 | 8.15E-01 | 3.47E-02 | 5.22E-01 | 2.15E-02 | 4.90E-01 |
| *KIAA0319L* | 9.89E-03 | 3.97E-01 | 2.12E-05 | 7.54E-03 | 6.35E-04 | 1.03E-01 |
| *ANHX* | NA | NA | NA | NA | NA | NA |
| *C17orf78* | NA | NA | NA | NA | NA | NA |
| *C1orf35* | NA | NA | NA | NA | NA | NA |
| *RHOF* | NA | NA | NA | NA | NA | NA |
| *SYT11* | 6.74E-01 | 9.66E-01 | 1.98E-01 | 7.89E-01 | 4.67E-05 | 1.61E-02 |
| *PRR15L* | 8.38E-04 | 1.31E-01 | 2.17E-03 | 1.72E-01 | 5.11E-03 | 3.08E-01 |
| *STK19* | 2.85E-01 | 8.68E-01 | 1.64E-03 | 1.47E-01 | 7.32E-05 | 2.09E-02 |
| *NECAB3* | 8.47E-01 | 9.89E-01 | 3.36E-01 | 8.68E-01 | 2.38E-01 | 8.13E-01 |
| *CNPY4* | 1.06E-01 | 7.20E-01 | 5.61E-03 | 2.56E-01 | 9.41E-03 | 3.79E-01 |
| *IFNA6* | NA | NA | NA | NA | NA | NA |
| *DST* | NA | NA | NA | NA | NA | NA |
| *ABTB2* | 2.01E-02 | 4.81E-01 | 1.34E-04 | 2.96E-02 | 2.08E-04 | 4.66E-02 |
| *PRICKLE4* | 4.12E-01 | 9.05E-01 | 1.46E-01 | 7.30E-01 | 1.93E-01 | 7.80E-01 |
| *KLC1* | 7.18E-01 | 9.73E-01 | 7.40E-01 | 9.73E-01 | 9.15E-01 | 9.93E-01 |
| *SPPL2C* | NA | NA | NA | NA | NA | NA |
| *PBXIP1* | NA | NA | NA | NA | NA | NA |
| *PPP1R12C* | NA | NA | NA | NA | NA | NA |
| *MARS2* | NA | NA | NA | NA | NA | NA |
| *UBE2D4* | 9.54E-01 | 9.97E-01 | 2.93E-01 | 8.51E-01 | 2.99E-02 | 5.30E-01 |
| *MARCKSL1* | NA | NA | NA | NA | NA | NA |
| *APOPT1* | 7.64E-03 | 3.76E-01 | 8.13E-06 | 3.71E-03 | 1.40E-06 | 8.16E-04 |
| *DHDDS* | 5.69E-04 | 1.04E-01 | 5.10E-03 | 2.39E-01 | 1.58E-02 | 4.53E-01 |
| *NINJ2* | NA | NA | NA | NA | NA | NA |
| *WFDC2* | NA | NA | NA | NA | NA | NA |
| *VIPR2* | 1.17E-01 | 7.27E-01 | 9.23E-01 | 9.92E-01 | 9.41E-01 | 9.95E-01 |
| *GATA6-AS1* | NA | NA | NA | NA | NA | NA |
| *BMP1* | 4.28E-02 | 5.68E-01 | 4.14E-05 | 1.31E-02 | 2.75E-03 | 2.30E-01 |
| *HLA-B* | 6.65E-01 | 9.66E-01 | 6.55E-01 | 9.55E-01 | 2.90E-01 | 8.46E-01 |
| *BGLAP* | 5.22E-01 | 9.40E-01 | 9.63E-01 | 9.95E-01 | 8.97E-01 | 9.93E-01 |
| *KANSL1* | NA | NA | NA | NA | NA | NA |
| *NELFE* | 4.60E-01 | 9.20E-01 | 6.75E-01 | 9.59E-01 | 5.38E-01 | 9.29E-01 |

| | | | | | | |
|---|---|---|---|---|---|---|
| *RGS9* | NA | NA | NA | NA | NA | NA |
| *SPEF2* | 6.02E-01 | 9.53E-01 | 5.47E-01 | 9.32E-01 | 2.40E-01 | 8.14E-01 |
| *CTD-3010D24.3* | NA | NA | NA | NA | NA | NA |
| *ZSCAN21* | 9.36E-02 | 7.06E-01 | 1.45E-03 | 1.39E-01 | 9.73E-02 | 6.74E-01 |
| *CAMSAP2* | 4.12E-01 | 9.05E-01 | 4.19E-01 | 8.98E-01 | 2.65E-01 | 8.34E-01 |
| *UBE2L6* | 3.67E-01 | 8.92E-01 | 1.38E-01 | 7.20E-01 | 2.09E-01 | 7.89E-01 |
| *GIGYF1* | 6.17E-03 | 3.48E-01 | 8.63E-01 | 9.88E-01 | 7.39E-02 | 6.39E-01 |
| *GOLGA8K* | NA | NA | NA | NA | NA | NA |
| *ZCCHC24* | 4.37E-01 | 9.12E-01 | 7.24E-01 | 9.71E-01 | 5.41E-01 | 9.31E-01 |
| *TFAP2E* | 1.15E-01 | 7.27E-01 | 6.44E-04 | 8.89E-02 | 2.47E-03 | 2.15E-01 |
| *CYP3A43* | 7.53E-01 | 9.77E-01 | 8.47E-01 | 9.87E-01 | 2.29E-01 | 8.06E-01 |
| *TCF7L2* | 5.95E-01 | 9.53E-01 | 8.31E-01 | 9.85E-01 | 1.48E-01 | 7.47E-01 |
| *SUPT6H* | NA | NA | NA | NA | NA | NA |
| *SYMPK* | 8.10E-01 | 9.86E-01 | 8.03E-01 | 9.80E-01 | 8.88E-01 | 9.91E-01 |
| *DDAH2* | 6.20E-01 | 9.55E-01 | 3.00E-01 | 8.51E-01 | 8.89E-02 | 6.66E-01 |
| *AATK-AS1* | NA | NA | NA | NA | NA | NA |
| *CREBRF* | NA | NA | NA | NA | NA | NA |
| *C1orf56* | 3.14E-01 | 8.72E-01 | 8.66E-02 | 6.51E-01 | 8.06E-03 | 3.70E-01 |
| *MSTO1* | NA | NA | NA | NA | NA | NA |
| *ANXA11* | 5.18E-01 | 9.40E-01 | 6.95E-01 | 9.66E-01 | 5.89E-01 | 9.41E-01 |
| *TNFSF8* | NA | NA | NA | NA | NA | NA |
| *GAL3ST4* | 2.98E-01 | 8.69E-01 | 1.09E-01 | 6.84E-01 | 2.57E-02 | 5.20E-01 |
| *COL11A2* | 4.30E-01 | 9.10E-01 | 8.87E-01 | 9.88E-01 | 6.09E-01 | 9.45E-01 |
| *SPOP* | 2.11E-01 | 8.22E-01 | 1.52E-01 | 7.35E-01 | 4.05E-01 | 8.87E-01 |
| *PLSCR2* | 8.53E-01 | 9.89E-01 | 2.28E-01 | 8.10E-01 | 1.13E-01 | 7.00E-01 |
| *KLF1* | 8.24E-03 | 3.85E-01 | 2.48E-01 | 8.25E-01 | 7.99E-01 | 9.80E-01 |
| *ARL17B* | 3.11E-01 | 8.72E-01 | 5.45E-05 | 1.56E-02 | 1.30E-02 | 4.17E-01 |
| *C6orf48* | 3.13E-01 | 8.72E-01 | 2.33E-01 | 8.13E-01 | 7.35E-01 | 9.72E-01 |
| *RPP21* | 2.09E-01 | 8.19E-01 | 8.25E-01 | 9.84E-01 | 4.58E-01 | 9.05E-01 |
| *RP11-57G10.8* | NA | NA | NA | NA | NA | NA |
| *YLPM1* | 6.58E-01 | 9.65E-01 | 6.37E-03 | 2.77E-01 | 7.54E-06 | 3.33E-03 |
| *GATAD2A* | 3.97E-01 | 9.00E-01 | 6.71E-01 | 9.58E-01 | 5.47E-01 | 9.33E-01 |
| *OAS2* | 4.13E-01 | 9.05E-01 | 1.35E-02 | 3.67E-01 | 5.85E-02 | 6.17E-01 |
| *HEY1* | NA | NA | NA | NA | NA | NA |
| *YY1AP1* | 8.77E-01 | 9.92E-01 | 1.33E-01 | 7.13E-01 | 1.32E-03 | 1.56E-01 |
| *FCHSD2* | 1.98E-01 | 8.15E-01 | 8.77E-01 | 9.88E-01 | 3.73E-01 | 8.76E-01 |
| *MAPT* | NA | NA | NA | NA | NA | NA |
| *GABARAPL2* | 3.65E-01 | 8.92E-01 | 2.35E-03 | 1.79E-01 | 1.61E-01 | 7.59E-01 |
| *CYP4B1* | 1.58E-01 | 7.78E-01 | 3.50E-01 | 8.75E-01 | 3.19E-01 | 8.60E-01 |
| *RP11-91I20.3* | NA | NA | NA | NA | NA | NA |
| *THRA* | 7.23E-04 | 1.19E-01 | 2.20E-03 | 1.72E-01 | 2.43E-03 | 2.14E-01 |
| *EDC3* | NA | NA | NA | NA | NA | NA |
| *SPARC* | 9.11E-02 | 7.00E-01 | 2.79E-01 | 8.46E-01 | 9.54E-01 | 9.95E-01 |
| *NPRL3* | 1.84E-01 | 8.05E-01 | 1.64E-01 | 7.56E-01 | 6.83E-01 | 9.59E-01 |

| | | | | | | |
|---|---|---|---|---|---|---|
| *EPS8L1* | 1.39E-03 | 1.89E-01 | 4.10E-02 | 5.44E-01 | 7.37E-01 | 9.72E-01 |
| *CFB* | NA | NA | NA | NA | NA | NA |
| *AC114752.1* | NA | NA | NA | NA | NA | NA |
| *CCDC18* | 1.82E-01 | 8.04E-01 | 2.59E-01 | 8.32E-01 | 1.90E-02 | 4.76E-01 |
| *LINC01315* | 1.11E-02 | 4.12E-01 | 9.02E-01 | 9.90E-01 | 6.75E-01 | 9.56E-01 |
| *NCOR1* | 2.43E-02 | 5.08E-01 | 5.63E-04 | 8.38E-02 | 2.29E-03 | 2.07E-01 |
| *SNX1* | 5.98E-01 | 9.53E-01 | 7.77E-02 | 6.32E-01 | 9.26E-02 | 6.71E-01 |
| *DNAH8* | NA | NA | NA | NA | NA | NA |
| *RAVER2* | 6.55E-01 | 9.64E-01 | 2.16E-01 | 8.02E-01 | 9.64E-01 | 9.96E-01 |
| *HIST1H2AB* | NA | NA | NA | NA | NA | NA |
| *SQRDL* | 8.61E-02 | 6.88E-01 | 1.98E-04 | 4.12E-02 | 4.01E-02 | 5.62E-01 |
| *FNIP1* | NA | NA | NA | NA | NA | NA |
| *TCF12* | 1.43E-04 | 3.71E-02 | 1.84E-01 | 7.72E-01 | 2.51E-01 | 8.23E-01 |
| *EPO* | NA | NA | NA | NA | NA | NA |
| *CTC-296K1.3* | 2.92E-02 | 5.17E-01 | 8.52E-02 | 6.49E-01 | 1.86E-01 | 7.75E-01 |
| *LAMTOR4* | 2.74E-01 | 8.64E-01 | 3.75E-01 | 8.82E-01 | 8.08E-01 | 9.80E-01 |
| *CTB-50L17.16* | NA | NA | NA | NA | NA | NA |
| *DONSON* | 4.82E-01 | 9.30E-01 | 1.24E-02 | 3.57E-01 | 4.33E-01 | 8.96E-01 |
| *ZPBP2* | NA | NA | NA | NA | NA | NA |
| *NFAM1* | NA | NA | NA | NA | NA | NA |
| *NCDN* | 6.41E-02 | 6.37E-01 | 7.33E-01 | 9.71E-01 | 8.23E-01 | 9.81E-01 |
| *ORMDL3* | 5.47E-05 | 1.73E-02 | 2.03E-05 | 7.51E-03 | 1.92E-03 | 1.94E-01 |
| *ZNF655* | 7.85E-01 | 9.83E-01 | 9.33E-02 | 6.59E-01 | 3.44E-01 | 8.70E-01 |
| *IBA57* | 5.78E-02 | 6.19E-01 | 7.90E-04 | 9.92E-02 | 3.05E-02 | 5.30E-01 |
| *CALR* | 7.78E-01 | 9.82E-01 | 7.45E-01 | 9.74E-01 | 8.99E-01 | 9.93E-01 |
| *CPOX* | 3.88E-03 | 2.98E-01 | 1.49E-01 | 7.34E-01 | 6.01E-02 | 6.26E-01 |
| *FAT2* | 1.27E-01 | 7.37E-01 | 3.15E-02 | 5.06E-01 | 6.35E-02 | 6.36E-01 |
| *ST3GAL3* | 9.63E-01 | 9.98E-01 | 2.41E-02 | 4.66E-01 | 1.71E-02 | 4.62E-01 |
| *DNAJB14* | 9.05E-01 | 9.94E-01 | 4.08E-01 | 8.97E-01 | 8.88E-01 | 9.91E-01 |
| *LINC01565* | 6.70E-01 | 9.66E-01 | 5.63E-03 | 2.56E-01 | 1.13E-01 | 7.00E-01 |
| *SNRPD2* | 1.65E-01 | 7.89E-01 | 7.15E-02 | 6.24E-01 | 5.82E-01 | 9.41E-01 |
| *GNB2* | 1.53E-02 | 4.38E-01 | 2.60E-01 | 8.34E-01 | 2.41E-01 | 8.14E-01 |
| *TSACC* | 1.84E-01 | 8.04E-01 | 8.55E-01 | 9.87E-01 | 9.83E-03 | 3.85E-01 |
| *DCST1* | NA | NA | NA | NA | NA | NA |
| *6-Mar* | NA | NA | NA | NA | NA | NA |
| *TRAF3* | 2.05E-03 | 2.34E-01 | 1.47E-01 | 7.31E-01 | 1.10E-02 | 3.97E-01 |
| *ZMYM4* | NA | NA | NA | NA | NA | NA |
| *AQP4* | 7.96E-01 | 9.86E-01 | 4.12E-01 | 8.97E-01 | 4.32E-02 | 5.78E-01 |
| *ARID3B* | NA | NA | NA | NA | NA | NA |
| *AP1M2* | 9.85E-02 | 7.15E-01 | 9.49E-01 | 9.95E-01 | 8.13E-01 | 9.80E-01 |
| *FAM189B* | 7.15E-01 | 9.73E-01 | 6.00E-01 | 9.43E-01 | 1.94E-01 | 7.81E-01 |
| *ADAMTS8* | 6.39E-01 | 9.59E-01 | 6.02E-01 | 9.43E-01 | 7.20E-01 | 9.69E-01 |
| *MICA* | 8.65E-01 | 9.90E-01 | 1.28E-02 | 3.60E-01 | 3.11E-02 | 5.30E-01 |
| *PRSS33* | NA | NA | NA | NA | NA | NA |
| *DAP3* | 8.77E-01 | 9.92E-01 | 1.33E-01 | 7.13E-01 | 1.32E-03 | 1.56E-01 |

| | | | | | | |
|---|---|---|---|---|---|---|
| *LINC01149* | NA | NA | NA | NA | NA | NA |
| *TAF6* | 3.94E-01 | 9.00E-01 | 1.02E-01 | 6.69E-01 | 7.68E-01 | 9.77E-01 |
| *GPANK1* | 1.96E-01 | 8.15E-01 | 2.00E-01 | 7.93E-01 | 3.55E-02 | 5.42E-01 |
| *LINC00310* | 4.64E-01 | 9.21E-01 | 2.65E-02 | 4.81E-01 | 7.95E-02 | 6.53E-01 |
| *RP11-503G7.2* | NA | NA | NA | NA | NA | NA |
| *FRYL* | 3.13E-02 | 5.27E-01 | 3.43E-04 | 5.76E-02 | 8.70E-03 | 3.77E-01 |
| *E2F1* | 7.04E-02 | 6.59E-01 | 4.63E-02 | 5.56E-01 | 4.11E-01 | 8.89E-01 |
| *PRRT1* | 1.03E-01 | 7.16E-01 | 1.28E-02 | 3.60E-01 | 2.92E-02 | 5.29E-01 |
| *IRF7* | 1.55E-02 | 4.38E-01 | 2.17E-02 | 4.50E-01 | 8.51E-03 | 3.76E-01 |
| *LINC00857* | NA | NA | NA | NA | NA | NA |
| *CERCAM* | 7.76E-03 | 3.80E-01 | 5.44E-01 | 9.31E-01 | 5.82E-01 | 9.41E-01 |
| *ZSWIM7* | 1.27E-02 | 4.24E-01 | 5.89E-05 | 1.65E-02 | 2.51E-04 | 5.31E-02 |
| *ADRA1A* | 1.45E-01 | 7.58E-01 | 3.04E-01 | 8.56E-01 | 4.68E-01 | 9.08E-01 |
| *NPY4R* | NA | NA | NA | NA | NA | NA |
| *TRMT61B* | 3.04E-03 | 2.76E-01 | 7.86E-05 | 2.02E-02 | 9.96E-02 | 6.79E-01 |
| *GJC2* | 2.73E-01 | 8.63E-01 | 2.61E-02 | 4.79E-01 | 1.97E-03 | 1.94E-01 |
| *PSMB2* | 2.34E-01 | 8.36E-01 | 3.21E-03 | 2.02E-01 | 5.71E-02 | 6.14E-01 |
| *PTGFR* | 9.06E-01 | 9.94E-01 | 8.78E-01 | 9.88E-01 | 8.91E-01 | 9.92E-01 |
| *VPS29* | 5.69E-01 | 9.49E-01 | 6.13E-01 | 9.43E-01 | 2.91E-01 | 8.46E-01 |
| *LINC00348* | NA | NA | NA | NA | NA | NA |
| *BRD2* | 5.89E-02 | 6.21E-01 | 4.76E-01 | 9.17E-01 | 6.64E-01 | 9.54E-01 |
| *UPF2* | 1.08E-01 | 7.20E-01 | 9.55E-01 | 9.95E-01 | 8.49E-02 | 6.61E-01 |
| *CHCHD7* | 8.30E-02 | 6.82E-01 | 7.51E-02 | 6.26E-01 | 1.23E-03 | 1.52E-01 |
| *HIST1H2BB* | NA | NA | NA | NA | NA | NA |
| *HSPE1* | 9.47E-01 | 9.97E-01 | 1.31E-01 | 7.11E-01 | 3.52E-03 | 2.52E-01 |
| *RP11-499E18.1* | 5.49E-01 | 9.45E-01 | 1.34E-01 | 7.14E-01 | 3.81E-01 | 8.79E-01 |
| *GCDH* | 7.57E-01 | 9.78E-01 | 2.15E-01 | 8.01E-01 | 4.77E-01 | 9.10E-01 |
| *ZNF146* | 6.74E-01 | 9.66E-01 | 5.23E-02 | 5.67E-01 | 9.69E-04 | 1.34E-01 |
| *CYP2C19* | 5.19E-01 | 9.40E-01 | 3.52E-01 | 8.75E-01 | 4.89E-02 | 5.92E-01 |
| *BNIP1* | 2.23E-01 | 8.30E-01 | 3.93E-02 | 5.38E-01 | 8.37E-01 | 9.84E-01 |
| *DR1* | 2.60E-03 | 2.57E-01 | 4.04E-03 | 2.25E-01 | 3.21E-04 | 6.23E-02 |
| *SUCLG2* | 3.47E-01 | 8.89E-01 | 3.85E-01 | 8.88E-01 | 8.69E-01 | 9.89E-01 |
| *TADA1* | 5.63E-01 | 9.48E-01 | 7.24E-02 | 6.24E-01 | 1.99E-01 | 7.84E-01 |
| *PRM3* | NA | NA | NA | NA | NA | NA |
| *PREX1* | 2.46E-01 | 8.47E-01 | 4.60E-01 | 9.13E-01 | 8.30E-01 | 9.83E-01 |
| *FAM96A* | NA | NA | NA | NA | NA | NA |
| *WFDC10A* | NA | NA | NA | NA | NA | NA |
| *AIP* | 2.95E-02 | 5.17E-01 | 4.93E-04 | 7.54E-02 | 1.83E-04 | 4.23E-02 |
| *PNRC1* | 4.25E-01 | 9.09E-01 | 2.96E-01 | 8.51E-01 | 1.15E-02 | 4.00E-01 |
| *XRCC3* | 2.81E-02 | 5.16E-01 | 3.55E-04 | 5.81E-02 | 2.15E-03 | 1.98E-01 |
| *LPCAT2* | NA | NA | NA | NA | NA | NA |
| *ATG16L2* | 1.67E-01 | 7.92E-01 | 3.65E-03 | 2.16E-01 | 1.57E-05 | 6.54E-03 |
| *ACSM4* | 7.42E-01 | 9.76E-01 | 2.25E-01 | 8.08E-01 | 8.55E-02 | 6.62E-01 |
| *ZNF311* | 5.24E-02 | 6.03E-01 | 5.44E-01 | 9.31E-01 | 6.43E-01 | 9.51E-01 |
| *HNMT* | 1.20E-01 | 7.34E-01 | 1.79E-01 | 7.68E-01 | 3.84E-01 | 8.80E-01 |

| | | | | | | |
|---|---|---|---|---|---|---|
| *PDLIM4* | 7.95E-01 | 9.86E-01 | 9.35E-01 | 9.93E-01 | 7.51E-01 | 9.75E-01 |
| *ILF3-AS1* | 3.68E-02 | 5.48E-01 | 4.88E-01 | 9.19E-01 | 4.23E-01 | 8.92E-01 |
| *PPP1R2* | 4.01E-01 | 9.00E-01 | 9.50E-01 | 9.95E-01 | 8.63E-01 | 9.88E-01 |
| *REEP4* | 4.47E-01 | 9.17E-01 | 6.98E-01 | 9.66E-01 | 9.97E-01 | 9.99E-01 |
| *RPN1* | 8.92E-01 | 9.93E-01 | 9.43E-02 | 6.59E-01 | 3.64E-01 | 8.75E-01 |
| *LPIN3* | 9.74E-03 | 3.95E-01 | 4.13E-02 | 5.45E-01 | 5.01E-02 | 5.95E-01 |
| *ARHGEF38* | NA | NA | NA | NA | NA | NA |
| *AP4M1* | 2.52E-02 | 5.10E-01 | 8.54E-04 | 1.03E-01 | 3.41E-02 | 5.40E-01 |
| *ZSCAN23* | 6.21E-01 | 9.55E-01 | 1.02E-02 | 3.32E-01 | 3.89E-02 | 5.58E-01 |
| *ZNF84* | 1.57E-01 | 7.75E-01 | 8.96E-02 | 6.54E-01 | 7.44E-02 | 6.39E-01 |
| *LMAN1L* | NA | NA | NA | NA | NA | NA |
| *PROB1* | 3.71E-03 | 2.94E-01 | 2.16E-02 | 4.50E-01 | 1.86E-03 | 1.94E-01 |
| *RP11-16E12.2* | 3.40E-01 | 8.86E-01 | 5.69E-02 | 5.80E-01 | 6.58E-02 | 6.37E-01 |
| *BMP8A* | 5.02E-01 | 9.35E-01 | 2.78E-01 | 8.46E-01 | 1.00E+00 | 1.00E+00 |
| *GSDMB* | 1.81E-04 | 4.55E-02 | 4.28E-06 | 2.18E-03 | 8.35E-04 | 1.23E-01 |
| *UNC5C* | NA | NA | NA | NA | NA | NA |
| *RASSF3* | 7.05E-01 | 9.70E-01 | 1.82E-01 | 7.70E-01 | 3.23E-01 | 8.62E-01 |
| *IMPG1* | 7.84E-01 | 9.83E-01 | 9.65E-01 | 9.96E-01 | 6.13E-01 | 9.46E-01 |
| *NDUFC1* | 6.86E-01 | 9.70E-01 | 4.73E-01 | 9.15E-01 | 8.64E-01 | 9.88E-01 |
| *DKFZP434L187* | NA | NA | NA | NA | NA | NA |
| *FAM126A* | 1.66E-01 | 7.90E-01 | 7.61E-01 | 9.75E-01 | 6.36E-01 | 9.50E-01 |
| *CPLX3* | 3.11E-01 | 8.72E-01 | 2.43E-04 | 4.82E-02 | 7.23E-03 | 3.50E-01 |
| *IKZF3* | 6.26E-04 | 1.08E-01 | 1.24E-04 | 2.88E-02 | 1.99E-03 | 1.94E-01 |
| *KIAA0907* | 3.10E-01 | 8.72E-01 | 7.86E-01 | 9.77E-01 | 2.03E-03 | 1.94E-01 |
| *ZKSCAN4* | 3.64E-02 | 5.47E-01 | 4.34E-03 | 2.25E-01 | 1.07E-01 | 6.90E-01 |
| *LKAAEAR1* | 1.65E-01 | 7.89E-01 | 1.38E-03 | 1.36E-01 | 2.03E-01 | 7.87E-01 |
| *DYNC1H1* | 5.52E-02 | 6.10E-01 | 1.17E-01 | 6.97E-01 | 4.52E-02 | 5.81E-01 |
| *GNGT2* | 9.03E-02 | 6.98E-01 | 2.11E-03 | 1.71E-01 | 3.78E-02 | 5.53E-01 |
| *CDC42EP3* | NA | NA | NA | NA | NA | NA |
| *NAA20* | NA | NA | NA | NA | NA | NA |
| *MS4A4A* | 4.08E-01 | 9.04E-01 | 1.16E-02 | 3.47E-01 | 6.47E-02 | 6.37E-01 |
| *OBSCN* | NA | NA | NA | NA | NA | NA |
| *C1orf145* | NA | NA | NA | NA | NA | NA |
| *CDH16* | NA | NA | NA | NA | NA | NA |
| *USP49* | 1.36E-02 | 4.33E-01 | 2.09E-04 | 4.24E-02 | 3.32E-01 | 8.64E-01 |
| *SNX19* | 1.14E-02 | 4.17E-01 | 7.74E-04 | 9.81E-02 | 2.43E-02 | 5.08E-01 |
| *FBXL18* | 1.80E-01 | 8.01E-01 | 1.88E-01 | 7.75E-01 | 9.29E-01 | 9.95E-01 |
| *VWA7* | 7.28E-01 | 9.73E-01 | 3.31E-02 | 5.14E-01 | 3.14E-02 | 5.30E-01 |
| *NAA15* | 7.16E-01 | 9.73E-01 | 2.46E-03 | 1.82E-01 | 6.69E-02 | 6.37E-01 |
| *DHX30* | NA | NA | NA | NA | NA | NA |
| *PCMTD1* | 7.90E-01 | 9.84E-01 | 1.40E-02 | 3.74E-01 | 5.51E-03 | 3.25E-01 |
| *HLA-DOA* | 4.25E-01 | 9.09E-01 | 4.73E-01 | 9.15E-01 | 7.79E-01 | 9.78E-01 |
| *RFTN2* | NA | NA | NA | NA | NA | NA |
| *ALKBH1* | 4.89E-01 | 9.32E-01 | 1.88E-02 | 4.27E-01 | 6.08E-01 | 9.45E-01 |
| *LDLR* | NA | NA | NA | NA | NA | NA |

| | | | | | | |
|---|---|---|---|---|---|---|
| *DSCAML1* | 7.97E-01 | 9.86E-01 | 3.49E-01 | 8.74E-01 | 1.65E-01 | 7.60E-01 |
| *LCTL* | 9.66E-01 | 9.98E-01 | 6.64E-02 | 6.10E-01 | 1.84E-02 | 4.71E-01 |
| *HINT1* | 1.02E-01 | 7.15E-01 | 1.88E-01 | 7.76E-01 | 1.92E-02 | 4.77E-01 |
| *MAP2* | 2.00E-02 | 4.80E-01 | 3.18E-01 | 8.60E-01 | 4.88E-01 | 9.14E-01 |
| *STAG3* | 8.31E-01 | 9.86E-01 | 9.67E-01 | 9.96E-01 | 3.33E-01 | 8.66E-01 |
| *IRF4* | 3.56E-02 | 5.43E-01 | 3.15E-04 | 5.51E-02 | 5.89E-02 | 6.19E-01 |
| *PKP2* | 2.78E-01 | 8.64E-01 | 5.17E-01 | 9.26E-01 | 1.96E-01 | 7.83E-01 |
| *EPB41L2* | 6.37E-01 | 9.58E-01 | 8.85E-01 | 9.88E-01 | 2.39E-01 | 8.13E-01 |
| *MSH5* | NA | NA | NA | NA | NA | NA |

Note: NA represents not applicable.

**Supplementary Table S9. The S-MultiXcan-identified 438 risk genes validated by S-PrediXcan analysis based on the GTEx whole blood**

| Gene Name | Susceptible COVID-19 (P-value) | Susceptible COVID-19 (FDR) | Hospitalized COVID-19 (P-value) | Hospitalized COVID-19 (FDR) | Very severe COVID-19 (P-value) | Very severe COVID-19 (FDR) |
|---|---|---|---|---|---|---|
| *CCR9* | 5.72E-71 | 6.99E-67 | 9.15E-216 | 1.12E-211 | 5.26E-217 | 6.43E-213 |
| *SLC6A20* | NA | NA | NA | NA | NA | NA |
| *CXCR6* | NA | NA | NA | NA | NA | NA |
| *XCR1* | 1.98E-04 | 7.14E-02 | 9.90E-09 | 2.02E-05 | 4.80E-08 | 5.86E-05 |
| *LZTFL1* | 5.19E-03 | 3.50E-01 | 2.45E-02 | 4.57E-01 | 1.34E-02 | 4.36E-01 |
| *ABO* | 3.32E-16 | 2.03E-12 | 5.61E-05 | 1.52E-02 | 6.07E-04 | 1.03E-01 |
| *CCR3* | 1.45E-01 | 7.54E-01 | 4.99E-02 | 5.85E-01 | 2.58E-02 | 5.11E-01 |
| *FYCO1* | 3.42E-01 | 8.86E-01 | 5.35E-05 | 1.49E-02 | 7.88E-04 | 1.14E-01 |
| *DPP9* | 1.49E-05 | 9.59E-03 | 4.29E-07 | 4.77E-04 | 1.80E-09 | 3.66E-06 |
| *IFNAR2* | 7.05E-02 | 6.45E-01 | 6.61E-05 | 1.72E-02 | 2.66E-03 | 2.29E-01 |
| *CCR1* | 2.92E-03 | 2.75E-01 | 1.84E-01 | 7.85E-01 | 2.74E-01 | 8.44E-01 |
| *CCR5* | 6.78E-02 | 6.39E-01 | 1.59E-01 | 7.60E-01 | 1.66E-01 | 7.83E-01 |
| *CCR2* | 1.19E-04 | 5.21E-02 | 3.47E-01 | 8.77E-01 | 6.40E-01 | 9.55E-01 |
| *IL10RB* | 6.71E-01 | 9.61E-01 | 4.23E-02 | 5.43E-01 | 9.89E-04 | 1.28E-01 |
| *CCRL2* | NA | NA | NA | NA | NA | NA |
| *NFKBIZ* | NA | NA | NA | NA | NA | NA |
| *NXPE3* | 6.49E-14 | 2.64E-10 | 7.25E-04 | 8.44E-02 | 7.95E-04 | 1.14E-01 |
| *MTX1* | 4.66E-08 | 5.77E-05 | 4.32E-02 | 5.48E-01 | 7.42E-01 | 9.70E-01 |
| *CEP97* | 4.09E-01 | 9.11E-01 | 3.52E-01 | 8.80E-01 | 5.87E-01 | 9.49E-01 |
| *FOXP4* | NA | NA | NA | NA | NA | NA |
| *MUC1* | 5.96E-01 | 9.57E-01 | 7.00E-10 | 2.14E-06 | 8.86E-15 | 5.41E-11 |
| *ZBTB11* | NA | NA | NA | NA | NA | NA |
| *ASH1L* | NA | NA | NA | NA | NA | NA |
| *AP000569.9* | NA | NA | NA | NA | NA | NA |
| *CACFD1* | 6.70E-01 | 9.61E-01 | 7.06E-01 | 9.68E-01 | 8.77E-01 | 9.84E-01 |
| *TRIM46* | NA | NA | NA | NA | NA | NA |
| *ATP5O* | 4.25E-01 | 9.15E-01 | 2.91E-06 | 1.87E-03 | 3.56E-04 | 7.38E-02 |
| *NAPSA* | 1.06E-08 | 2.16E-05 | 2.17E-18 | 1.33E-14 | 7.17E-11 | 2.19E-07 |
| *TULP2* | NA | NA | NA | NA | NA | NA |
| *PCNP* | 2.19E-03 | 2.46E-01 | 2.51E-03 | 1.62E-01 | 2.87E-01 | 8.48E-01 |

| | | | | | | |
|---|---|---|---|---|---|---|
| *MRPS6* | 8.40E-01 | 9.84E-01 | 6.28E-01 | 9.49E-01 | 8.80E-01 | 9.84E-01 |
| *SENP7* | 2.62E-10 | 8.01E-07 | 5.24E-04 | 6.97E-02 | 1.63E-01 | 7.83E-01 |
| *LINC02009* | 2.94E-02 | 5.31E-01 | 2.62E-03 | 1.67E-01 | 1.46E-02 | 4.43E-01 |
| *TYK2* | 2.76E-04 | 8.65E-02 | 2.11E-08 | 3.68E-05 | 2.79E-12 | 1.14E-08 |
| *FUT2* | NA | NA | NA | NA | NA | NA |
| *MED22* | 5.35E-01 | 9.46E-01 | 4.51E-01 | 9.17E-01 | 2.97E-01 | 8.59E-01 |
| *SLC50A1* | 8.99E-02 | 6.79E-01 | 8.75E-01 | 9.88E-01 | 3.40E-01 | 8.73E-01 |
| *ELF5* | NA | NA | NA | NA | NA | NA |
| *CAT* | 3.95E-02 | 5.64E-01 | 3.90E-03 | 2.10E-01 | 4.40E-03 | 2.76E-01 |
| *LTF* | NA | NA | NA | NA | NA | NA |
| *C6orf15* | NA | NA | NA | NA | NA | NA |
| *SLC2A6* | 1.68E-08 | 2.93E-05 | 4.03E-03 | 2.13E-01 | 3.39E-02 | 5.45E-01 |
| *ADAM15* | 4.81E-04 | 1.20E-01 | 1.47E-04 | 2.85E-02 | 4.95E-03 | 2.88E-01 |
| *PLEKHA4* | 3.18E-01 | 8.76E-01 | 2.41E-01 | 8.21E-01 | 2.06E-01 | 8.13E-01 |
| *TNFAIP8L1* | 8.25E-03 | 4.09E-01 | 3.07E-02 | 4.95E-01 | 1.00E-01 | 7.08E-01 |
| *NTN5* | 2.05E-05 | 1.21E-02 | 5.06E-03 | 2.34E-01 | 2.29E-03 | 2.16E-01 |
| *ATP11A* | 4.78E-03 | 3.43E-01 | 1.32E-06 | 1.01E-03 | 2.24E-04 | 5.17E-02 |
| *ADAMTS13* | 2.51E-02 | 5.24E-01 | 9.73E-03 | 3.23E-01 | 6.63E-01 | 9.59E-01 |
| *EFNA4* | NA | NA | NA | NA | NA | NA |
| *RP11-798G7.6* | NA | NA | NA | NA | NA | NA |
| *PPP1R15A* | 2.36E-01 | 8.31E-01 | 1.69E-01 | 7.70E-01 | 1.87E-01 | 8.03E-01 |
| *GON4L* | NA | NA | NA | NA | NA | NA |
| *FBRSL1* | 9.72E-03 | 4.23E-01 | 1.91E-03 | 1.44E-01 | 2.84E-01 | 8.47E-01 |
| *RASIP1* | 4.63E-09 | 1.13E-05 | 6.31E-04 | 7.64E-02 | 6.33E-08 | 7.03E-05 |
| *CCHCR1* | 1.80E-06 | 1.69E-03 | 3.58E-07 | 4.38E-04 | 8.89E-05 | 2.78E-02 |
| *NUCB1* | 2.91E-04 | 8.88E-02 | 3.30E-01 | 8.70E-01 | 8.92E-02 | 6.97E-01 |
| *MAMSTR* | 2.24E-08 | 3.42E-05 | 8.89E-05 | 2.03E-02 | 2.49E-08 | 3.38E-05 |
| *KRTCAP2* | NA | NA | NA | NA | NA | NA |
| *OAS3* | 1.36E-05 | 9.24E-03 | 2.67E-09 | 6.53E-06 | 4.03E-06 | 2.46E-03 |
| *FDX2* | 3.86E-01 | 9.05E-01 | 1.94E-01 | 7.94E-01 | 6.50E-01 | 9.57E-01 |
| *PDE4A* | 9.00E-01 | 9.90E-01 | 3.68E-01 | 8.88E-01 | 7.94E-01 | 9.74E-01 |
| *PLEKHM1* | NA | NA | NA | NA | NA | NA |
| *FUT1* | NA | NA | NA | NA | NA | NA |
| *SFTPD* | 1.82E-01 | 7.92E-01 | 1.46E-04 | 2.85E-02 | 3.75E-07 | 2.86E-04 |
| *ZGLP1* | 4.11E-03 | 3.22E-01 | 4.38E-05 | 1.43E-02 | 6.89E-04 | 1.09E-01 |
| *TCF19* | 3.08E-06 | 2.45E-03 | 2.42E-07 | 3.70E-04 | 2.88E-05 | 1.13E-02 |
| *C5orf56* | 3.21E-03 | 2.84E-01 | 9.75E-04 | 9.54E-02 | 7.71E-03 | 3.59E-01 |
| *HLA-C* | 4.11E-06 | 2.96E-03 | 2.79E-07 | 3.79E-04 | 4.40E-09 | 6.72E-06 |
| *ARHGAP27* | 1.77E-02 | 4.97E-01 | 5.60E-06 | 3.11E-03 | 4.52E-02 | 5.99E-01 |
| *SLC35B1* | 5.67E-01 | 9.54E-01 | 8.28E-01 | 9.84E-01 | 6.12E-01 | 9.52E-01 |
| *DDX39B* | 4.67E-01 | 9.30E-01 | 2.63E-04 | 4.40E-02 | 3.41E-02 | 5.45E-01 |
| *PSORS1C2* | NA | NA | NA | NA | NA | NA |
| *IZUMO1* | NA | NA | NA | NA | NA | NA |
| *FAM215B* | NA | NA | NA | NA | NA | NA |

| | | | | | | |
|---|---|---|---|---|---|---|
| *REXO4* | 4.47E-02 | 5.82E-01 | 9.06E-01 | 9.91E-01 | 2.78E-01 | 8.46E-01 |
| *ACSF3* | 6.22E-03 | 3.70E-01 | 7.70E-01 | 9.77E-01 | 3.88E-01 | 8.83E-01 |
| *RAVER1* | 1.29E-02 | 4.48E-01 | 4.83E-06 | 2.95E-03 | 1.61E-05 | 7.87E-03 |
| *ICAM5* | 3.91E-02 | 5.63E-01 | 6.14E-03 | 2.62E-01 | 1.25E-01 | 7.41E-01 |
| *OAS1* | 1.64E-04 | 6.29E-02 | 4.43E-05 | 1.43E-02 | 2.32E-03 | 2.16E-01 |
| *ICAM3* | 9.40E-01 | 9.94E-01 | 4.90E-01 | 9.27E-01 | 3.55E-01 | 8.74E-01 |
| *ZCCHC4* | 3.17E-04 | 9.04E-02 | 1.23E-04 | 2.53E-02 | 1.25E-02 | 4.23E-01 |
| *KCNC3* | NA | NA | NA | NA | NA | NA |
| *NPNT* | 1.48E-01 | 7.56E-01 | 5.12E-03 | 2.34E-01 | 5.71E-03 | 3.14E-01 |
| *WNT3* | 9.49E-03 | 4.21E-01 | 2.89E-10 | 1.18E-06 | 1.23E-10 | 3.01E-07 |
| *DCST2* | 2.36E-02 | 5.16E-01 | 5.32E-03 | 2.36E-01 | 5.44E-03 | 3.06E-01 |
| *HLA-A* | 6.92E-01 | 9.65E-01 | 2.94E-02 | 4.85E-01 | 3.41E-02 | 5.45E-01 |
| *HSD17B14* | NA | NA | NA | NA | NA | NA |
| *PI4K2B* | 2.41E-02 | 5.19E-01 | 4.36E-05 | 1.43E-02 | 1.29E-02 | 4.27E-01 |
| *CSF3* | NA | NA | NA | NA | NA | NA |
| *RPL7A* | 2.74E-02 | 5.31E-01 | 1.04E-01 | 6.92E-01 | 7.83E-02 | 6.81E-01 |
| *SEMA6B* | 1.85E-01 | 7.96E-01 | 2.72E-01 | 8.40E-01 | 2.10E-01 | 8.14E-01 |
| *SFTPA2* | 4.57E-01 | 9.27E-01 | 2.65E-02 | 4.70E-01 | 1.51E-01 | 7.74E-01 |
| *CDH15* | NA | NA | NA | NA | NA | NA |
| *RP11-589B3.6* | NA | NA | NA | NA | NA | NA |
| *LINC01301* | NA | NA | NA | NA | NA | NA |
| *THBS3* | 4.72E-08 | 5.77E-05 | 8.75E-01 | 9.88E-01 | 6.63E-04 | 1.07E-01 |
| *RAB2A* | 3.48E-03 | 2.99E-01 | 4.46E-04 | 6.34E-02 | 8.38E-03 | 3.68E-01 |
| *SACM1L* | 2.63E-02 | 5.29E-01 | 4.80E-01 | 9.25E-01 | 3.62E-01 | 8.74E-01 |
| *IBA57-AS1* | NA | NA | NA | NA | NA | NA |
| *MED24* | 7.21E-05 | 3.53E-02 | 1.59E-04 | 3.03E-02 | 6.28E-05 | 2.07E-02 |
| *DPF2* | 2.53E-01 | 8.44E-01 | 4.97E-04 | 6.82E-02 | 7.27E-01 | 9.67E-01 |
| *GSDMA* | 8.71E-02 | 6.76E-01 | 6.64E-01 | 9.59E-01 | 9.18E-01 | 9.90E-01 |
| *PMVK* | 5.03E-02 | 5.97E-01 | 7.15E-01 | 9.69E-01 | 1.98E-01 | 8.06E-01 |
| *PSMD3* | 7.11E-01 | 9.66E-01 | 3.84E-01 | 8.92E-01 | 8.38E-01 | 9.79E-01 |
| *RP11-506M13.3* | 2.85E-02 | 5.31E-01 | 1.63E-04 | 3.06E-02 | 9.15E-04 | 1.22E-01 |
| *NUTM2E* | NA | NA | NA | NA | NA | NA |
| *CSK* | 3.59E-04 | 9.66E-02 | 4.67E-05 | 1.43E-02 | 1.11E-01 | 7.19E-01 |
| *RP4-535B20.1* | NA | NA | NA | NA | NA | NA |
| *MICB* | 1.08E-01 | 7.11E-01 | 2.00E-03 | 1.47E-01 | 2.19E-02 | 5.01E-01 |
| *RTP3* | NA | NA | NA | NA | NA | NA |
| *RP11-387H17.6* | NA | NA | NA | NA | NA | NA |
| *ARL17A* | NA | NA | NA | NA | NA | NA |
| *HLA-DPA1* | 4.54E-02 | 5.85E-01 | 3.37E-01 | 8.73E-01 | 4.05E-01 | 8.89E-01 |
| *PSORS1C1* | NA | NA | NA | NA | NA | NA |
| *RP3-462D8.2* | NA | NA | NA | NA | NA | NA |
| *MEF2D* | 1.80E-02 | 4.97E-01 | 2.87E-01 | 8.44E-01 | 7.63E-01 | 9.73E-01 |
| *ICAM1* | 1.58E-02 | 4.80E-01 | 1.49E-05 | 6.75E-03 | 2.99E-04 | 6.30E-02 |

| | | | | | | |
|---|---|---|---|---|---|---|
| *RP11-11C20.3* | NA | NA | NA | NA | NA | NA |
| *RPL24* | 2.55E-07 | 2.60E-04 | 1.23E-04 | 2.53E-02 | 1.67E-03 | 1.85E-01 |
| *SLC22A4* | 3.02E-01 | 8.73E-01 | 3.17E-01 | 8.59E-01 | 4.08E-01 | 8.90E-01 |
| *GBA* | NA | NA | NA | NA | NA | NA |
| *MCM7* | 2.88E-01 | 8.66E-01 | 7.75E-01 | 9.77E-01 | 8.41E-02 | 6.92E-01 |
| *COQ10B* | NA | NA | NA | NA | NA | NA |
| *LTA* | 3.42E-01 | 8.86E-01 | 2.99E-03 | 1.84E-01 | 4.83E-02 | 6.02E-01 |
| *IMPG2* | 3.37E-01 | 8.83E-01 | 9.21E-02 | 6.70E-01 | 3.16E-02 | 5.39E-01 |
| *LRRC37A2* | NA | NA | NA | NA | NA | NA |
| *DBP* | NA | NA | NA | NA | NA | NA |
| *CLK3* | 2.48E-01 | 8.41E-01 | 1.15E-02 | 3.41E-01 | 1.71E-03 | 1.88E-01 |
| *ZNF3* | 3.94E-01 | 9.06E-01 | 1.59E-03 | 1.31E-01 | 1.52E-02 | 4.49E-01 |
| *ZNF778* | 4.04E-01 | 9.08E-01 | 2.10E-01 | 8.06E-01 | 9.71E-01 | 9.96E-01 |
| *PPP1R11* | 1.76E-01 | 7.83E-01 | 1.97E-01 | 7.97E-01 | 2.65E-02 | 5.11E-01 |
| *SLC5A3* | NA | NA | NA | NA | NA | NA |
| *CDSN* | NA | NA | NA | NA | NA | NA |
| *ACSL6* | 2.44E-05 | 1.32E-02 | 8.14E-07 | 7.11E-04 | 3.40E-06 | 2.31E-03 |
| *PGC* | NA | NA | NA | NA | NA | NA |
| *DSP* | NA | NA | NA | NA | NA | NA |
| *HLA-DQA2* | 4.88E-03 | 3.46E-01 | 7.29E-05 | 1.78E-02 | 1.26E-07 | 1.18E-04 |
| *IFNAR1* | 1.35E-01 | 7.43E-01 | 3.98E-04 | 5.90E-02 | 1.28E-04 | 3.46E-02 |
| *FOXA3* | NA | NA | NA | NA | NA | NA |
| *HLA-DPB1* | 1.72E-02 | 4.93E-01 | 7.42E-01 | 9.75E-01 | 2.75E-01 | 8.44E-01 |
| *DTX1* | 7.13E-02 | 6.46E-01 | 1.28E-01 | 7.31E-01 | 4.28E-01 | 8.99E-01 |
| *MYDGF* | 2.07E-05 | 1.21E-02 | 2.64E-06 | 1.79E-03 | 2.66E-07 | 2.32E-04 |
| *RP11-182L21.6* | 3.21E-01 | 8.77E-01 | 1.88E-01 | 7.91E-01 | 1.93E-04 | 4.71E-02 |
| *EIF4EBP2* | 6.45E-02 | 6.30E-01 | 4.05E-01 | 9.01E-01 | 5.36E-01 | 9.36E-01 |
| *ANAPC4* | 2.04E-03 | 2.42E-01 | 1.25E-06 | 1.01E-03 | 1.22E-04 | 3.39E-02 |
| *SLC22A31* | 1.79E-01 | 7.88E-01 | 1.77E-05 | 7.46E-03 | 2.92E-04 | 6.25E-02 |
| *SPACA4* | NA | NA | NA | NA | NA | NA |
| *TFR2* | 5.27E-02 | 6.02E-01 | 7.25E-01 | 9.71E-01 | 6.26E-01 | 9.53E-01 |
| *HLA-DRA* | NA | NA | NA | NA | NA | NA |
| *ZKSCAN1* | 4.25E-01 | 9.15E-01 | 3.82E-01 | 8.90E-01 | 6.71E-02 | 6.58E-01 |
| *SLC25A44* | NA | NA | NA | NA | NA | NA |
| *PMF1* | 3.20E-06 | 2.45E-03 | 1.64E-02 | 3.94E-01 | 2.01E-04 | 4.71E-02 |
| *MOSPD3* | 1.90E-07 | 2.11E-04 | 9.21E-04 | 9.54E-02 | 2.34E-03 | 2.16E-01 |
| *BYSL* | 5.81E-01 | 9.57E-01 | 5.16E-01 | 9.36E-01 | 7.31E-01 | 9.68E-01 |
| *CTA-384D8.35* | 5.36E-01 | 9.46E-01 | 3.12E-01 | 8.57E-01 | 1.65E-02 | 4.61E-01 |
| *ACTL6B* | NA | NA | NA | NA | NA | NA |
| *HIP1* | 4.40E-02 | 5.79E-01 | 1.76E-05 | 7.46E-03 | 1.09E-07 | 1.11E-04 |
| *EFNA1* | 7.36E-01 | 9.69E-01 | 1.14E-05 | 5.36E-03 | 2.47E-09 | 4.31E-06 |
| *CDC37* | NA | NA | NA | NA | NA | NA |
| *CHD1* | 1.36E-01 | 7.43E-01 | 1.60E-01 | 7.61E-01 | 1.86E-01 | 8.03E-01 |
| *PCOLCE* | 7.17E-01 | 9.66E-01 | 5.86E-02 | 5.99E-01 | 2.25E-01 | 8.20E-01 |

| | | | | | | |
|---|---|---|---|---|---|---|
| *AC006007.1* | NA | NA | NA | NA | NA | NA |
| *CA11* | 2.10E-02 | 5.12E-01 | 2.77E-01 | 8.42E-01 | 1.96E-01 | 8.06E-01 |
| *FUBP1* | 2.59E-03 | 2.63E-01 | 5.16E-07 | 4.85E-04 | 2.03E-06 | 1.46E-03 |
| *SH3BP2* | 1.46E-01 | 7.54E-01 | 1.33E-01 | 7.34E-01 | 2.17E-01 | 8.19E-01 |
| *EFNA3* | 7.01E-01 | 9.65E-01 | 7.01E-02 | 6.29E-01 | 9.39E-01 | 9.92E-01 |
| *C2orf16* | NA | NA | NA | NA | NA | NA |
| *MYL5* | 3.11E-01 | 8.75E-01 | 3.13E-02 | 4.97E-01 | 2.11E-01 | 8.14E-01 |
| *ADGRE3* | 1.01E-02 | 4.24E-01 | 3.82E-01 | 8.90E-01 | 5.60E-01 | 9.42E-01 |
| *CLMP* | 2.19E-01 | 8.17E-01 | 3.44E-01 | 8.75E-01 | 9.40E-02 | 7.03E-01 |
| *HLA-F* | 2.05E-01 | 8.07E-01 | 1.92E-01 | 7.92E-01 | 5.11E-02 | 6.13E-01 |
| *GATS* | 6.77E-01 | 9.61E-01 | 8.38E-01 | 9.85E-01 | 3.45E-02 | 5.47E-01 |
| *FDPS* | 6.44E-01 | 9.61E-01 | 4.87E-01 | 9.27E-01 | 1.21E-02 | 4.21E-01 |
| *ZAN* | NA | NA | NA | NA | NA | NA |
| *TNFRSF4* | 2.43E-01 | 8.38E-01 | 2.59E-01 | 8.34E-01 | 1.75E-01 | 7.91E-01 |
| *BCAT2* | 2.27E-03 | 2.46E-01 | 7.29E-01 | 9.71E-01 | 6.74E-01 | 9.62E-01 |
| *CYP1A1* | 7.15E-01 | 9.66E-01 | 1.32E-01 | 7.34E-01 | 7.39E-01 | 9.70E-01 |
| *TOMM7* | 2.37E-01 | 8.31E-01 | 9.73E-02 | 6.76E-01 | 1.00E-02 | 4.00E-01 |
| *YIPF2* | 4.93E-01 | 9.37E-01 | 6.75E-01 | 9.62E-01 | 2.83E-02 | 5.20E-01 |
| *POU5F1* | 6.84E-01 | 9.64E-01 | 5.18E-01 | 9.36E-01 | 5.24E-01 | 9.32E-01 |
| *P4HA2* | 8.23E-01 | 9.81E-01 | 4.89E-01 | 9.27E-01 | 5.18E-01 | 9.31E-01 |
| *RP11-298J23.8* | NA | NA | NA | NA | NA | NA |
| *DNAJB4* | NA | NA | NA | NA | NA | NA |
| *NLRP4* | NA | NA | NA | NA | NA | NA |
| *FRS3* | 8.52E-02 | 6.74E-01 | 1.50E-03 | 1.26E-01 | 7.52E-02 | 6.79E-01 |
| *GPC2* | 1.35E-03 | 2.09E-01 | 4.72E-05 | 1.43E-02 | 2.22E-02 | 5.02E-01 |
| *AGRN* | 2.61E-02 | 5.29E-01 | 6.21E-01 | 9.49E-01 | 6.18E-01 | 9.52E-01 |
| *COPS6* | 6.71E-01 | 9.61E-01 | 7.72E-01 | 9.77E-01 | 4.70E-02 | 5.99E-01 |
| *ATF6B* | 5.29E-04 | 1.29E-01 | 8.98E-05 | 2.03E-02 | 5.60E-04 | 9.63E-02 |
| *GUK1* | 6.04E-01 | 9.57E-01 | 4.19E-01 | 9.09E-01 | 8.51E-01 | 9.81E-01 |
| *RP11-298J23.10* | 4.24E-01 | 9.14E-01 | 1.48E-01 | 7.47E-01 | 1.89E-01 | 8.03E-01 |
| *CLK2* | NA | NA | NA | NA | NA | NA |
| *RP13-554M15.8* | NA | NA | NA | NA | NA | NA |
| *AZGP1* | NA | NA | NA | NA | NA | NA |
| *HCN3* | 7.96E-01 | 9.79E-01 | 4.73E-03 | 2.28E-01 | 2.30E-05 | 1.02E-02 |
| *SURF2* | 2.70E-01 | 8.54E-01 | 2.20E-01 | 8.09E-01 | 2.33E-01 | 8.21E-01 |
| *JAK1* | 7.38E-01 | 9.70E-01 | 7.52E-01 | 9.75E-01 | 7.93E-01 | 9.74E-01 |
| *SCAMP3* | NA | NA | NA | NA | NA | NA |
| *LINC00304* | 4.29E-01 | 9.16E-01 | 1.08E-04 | 2.32E-02 | 1.96E-05 | 9.21E-03 |
| *ZBTB7B* | 1.00E-01 | 6.98E-01 | 9.07E-01 | 9.91E-01 | 5.64E-01 | 9.42E-01 |
| *ACAD8* | NA | NA | NA | NA | NA | NA |
| *HLA-DMB* | 8.87E-01 | 9.89E-01 | 6.19E-02 | 6.09E-01 | 6.60E-04 | 1.07E-01 |
| *TRIM15* | 9.76E-01 | 9.98E-01 | 2.16E-02 | 4.48E-01 | 1.34E-02 | 4.36E-01 |
| *RNF17* | NA | NA | NA | NA | NA | NA |
| *KISS1* | NA | NA | NA | NA | NA | NA |

| | | | | | | |
|---|---|---|---|---|---|---|
| *ABCF1* | NA | NA | NA | NA | NA | NA |
| *BTNL2* | NA | NA | NA | NA | NA | NA |
| *PKLR* | NA | NA | NA | NA | NA | NA |
| *FAM83E* | NA | NA | NA | NA | NA | NA |
| *TIMM29* | 6.18E-01 | 9.59E-01 | 8.52E-01 | 9.87E-01 | 7.84E-01 | 9.74E-01 |
| *LINC02138* | NA | NA | NA | NA | NA | NA |
| *LTB* | 2.34E-02 | 5.16E-01 | 1.07E-03 | 1.01E-01 | 5.21E-02 | 6.18E-01 |
| *DDR1* | 9.33E-01 | 9.94E-01 | 3.94E-01 | 8.94E-01 | 7.57E-01 | 9.73E-01 |
| *TDGF1* | NA | NA | NA | NA | NA | NA |
| *RSPO2* | NA | NA | NA | NA | NA | NA |
| *SFTPA1* | 9.37E-01 | 9.94E-01 | 6.86E-01 | 9.65E-01 | 2.67E-01 | 8.41E-01 |
| *RIT1* | 6.38E-01 | 9.61E-01 | 6.02E-01 | 9.48E-01 | 4.46E-05 | 1.65E-02 |
| *NUTM2B* | 1.62E-01 | 7.72E-01 | 2.26E-02 | 4.52E-01 | 2.61E-02 | 5.11E-01 |
| *KIAA0319L* | 9.89E-03 | 4.24E-01 | 2.12E-05 | 8.36E-03 | 6.35E-04 | 1.05E-01 |
| *ANHX* | NA | NA | NA | NA | NA | NA |
| *C17orf78* | NA | NA | NA | NA | NA | NA |
| *C1orf35* | 4.82E-01 | 9.34E-01 | 3.38E-01 | 8.73E-01 | 2.14E-01 | 8.15E-01 |
| *RHOF* | 6.05E-01 | 9.57E-01 | 7.30E-02 | 6.35E-01 | 5.27E-02 | 6.19E-01 |
| *SYT11* | 6.74E-01 | 9.61E-01 | 1.98E-01 | 7.97E-01 | 4.67E-05 | 1.68E-02 |
| *PRR15L* | 2.66E-01 | 8.53E-01 | 4.63E-02 | 5.69E-01 | 1.07E-01 | 7.16E-01 |
| *STK19* | 3.62E-01 | 8.96E-01 | 1.40E-01 | 7.37E-01 | 2.40E-03 | 2.16E-01 |
| *NECAB3* | 8.47E-01 | 9.85E-01 | 3.36E-01 | 8.72E-01 | 2.38E-01 | 8.21E-01 |
| *CNPY4* | 5.01E-03 | 3.46E-01 | 2.62E-04 | 4.40E-02 | 7.53E-06 | 4.00E-03 |
| *IFNA6* | NA | NA | NA | NA | NA | NA |
| *DST* | 6.74E-01 | 9.61E-01 | 5.80E-01 | 9.41E-01 | 9.70E-01 | 9.96E-01 |
| *ABTB2* | 6.59E-04 | 1.47E-01 | 5.10E-07 | 4.85E-04 | 3.44E-07 | 2.80E-04 |
| *PRICKLE4* | 4.12E-01 | 9.11E-01 | 1.46E-01 | 7.44E-01 | 1.93E-01 | 8.05E-01 |
| *KLC1* | 9.27E-02 | 6.84E-01 | 5.47E-03 | 2.38E-01 | 1.96E-02 | 4.80E-01 |
| *SPPL2C* | NA | NA | NA | NA | NA | NA |
| *PBXIP1* | 1.48E-01 | 7.56E-01 | 8.13E-01 | 9.82E-01 | 4.08E-01 | 8.90E-01 |
| *PPP1R12C* | 2.49E-05 | 1.32E-02 | 2.05E-01 | 8.03E-01 | 8.12E-01 | 9.76E-01 |
| *MARS2* | NA | NA | NA | NA | NA | NA |
| *UBE2D4* | 7.68E-01 | 9.76E-01 | 3.00E-01 | 8.49E-01 | 1.96E-03 | 2.06E-01 |
| *MARCKSL1* | NA | NA | NA | NA | NA | NA |
| *APOPT1* | 2.64E-02 | 5.29E-01 | 3.96E-05 | 1.38E-02 | 4.90E-06 | 2.72E-03 |
| *DHDDS* | 5.69E-04 | 1.31E-01 | 5.10E-03 | 2.34E-01 | 1.58E-02 | 4.53E-01 |
| *NINJ2* | 3.22E-01 | 8.77E-01 | 1.21E-01 | 7.29E-01 | 1.26E-03 | 1.54E-01 |
| *WFDC2* | 2.48E-01 | 8.41E-01 | 4.52E-01 | 9.17E-01 | 8.35E-01 | 9.79E-01 |
| *VIPR2* | NA | NA | NA | NA | NA | NA |
| *GATA6-AS1* | NA | NA | NA | NA | NA | NA |
| *BMP1* | 2.61E-01 | 8.50E-01 | 9.57E-02 | 6.75E-01 | 3.33E-01 | 8.71E-01 |
| *HLA-B* | 3.06E-02 | 5.33E-01 | 1.91E-04 | 3.55E-02 | 1.13E-02 | 4.12E-01 |
| *BGLAP* | 1.64E-03 | 2.19E-01 | 1.33E-01 | 7.34E-01 | 2.04E-02 | 4.89E-01 |
| *KANSL1* | NA | NA | NA | NA | NA | NA |
| *NELFE* | 4.07E-01 | 9.09E-01 | 7.69E-01 | 9.77E-01 | 3.90E-01 | 8.85E-01 |

| | | | | | | |
|---|---|---|---|---|---|---|
| *RGS9* | 2.56E-01 | 8.46E-01 | 7.30E-01 | 9.71E-01 | 1.99E-01 | 8.09E-01 |
| *SPEF2* | NA | NA | NA | NA | NA | NA |
| *CTD-3010D24.3* | NA | NA | NA | NA | NA | NA |
| *ZSCAN21* | 2.23E-03 | 2.46E-01 | 1.97E-06 | 1.42E-03 | 1.21E-04 | 3.39E-02 |
| *CAMSAP2* | 6.54E-01 | 9.61E-01 | 1.45E-01 | 7.44E-01 | 5.39E-02 | 6.22E-01 |
| *UBE2L6* | 9.25E-01 | 9.94E-01 | 4.17E-01 | 9.07E-01 | 6.14E-01 | 9.52E-01 |
| *GIGYF1* | 5.70E-04 | 1.31E-01 | 5.15E-01 | 9.36E-01 | 4.35E-02 | 5.93E-01 |
| *GOLGA8K* | NA | NA | NA | NA | NA | NA |
| *ZCCHC24* | 3.10E-01 | 8.75E-01 | 4.02E-01 | 8.99E-01 | 9.45E-01 | 9.92E-01 |
| *TFAP2E* | 9.01E-02 | 6.79E-01 | 4.46E-01 | 9.17E-01 | 8.67E-01 | 9.82E-01 |
| *CYP3A43* | NA | NA | NA | NA | NA | NA |
| *TCF7L2* | 1.85E-01 | 7.96E-01 | 7.89E-01 | 9.79E-01 | 4.31E-01 | 9.00E-01 |
| *SUPT6H* | 2.31E-02 | 5.15E-01 | 1.86E-01 | 7.87E-01 | 8.35E-02 | 6.92E-01 |
| *SYMPK* | 3.87E-03 | 3.13E-01 | 1.31E-02 | 3.58E-01 | 5.97E-03 | 3.19E-01 |
| *DDAH2* | 1.32E-01 | 7.39E-01 | 3.31E-02 | 5.05E-01 | 2.03E-02 | 4.89E-01 |
| *AATK-AS1* | NA | NA | NA | NA | NA | NA |
| *CREBRF* | NA | NA | NA | NA | NA | NA |
| *C1orf56* | 7.06E-02 | 6.45E-01 | 5.14E-03 | 2.34E-01 | 4.26E-05 | 1.63E-02 |
| *MSTO1* | NA | NA | NA | NA | NA | NA |
| *ANXA11* | 2.36E-01 | 8.31E-01 | 2.51E-01 | 8.27E-01 | 5.43E-01 | 9.39E-01 |
| *TNFSF8* | 7.70E-01 | 9.76E-01 | 8.46E-01 | 9.86E-01 | 3.34E-01 | 8.72E-01 |
| *GAL3ST4* | 9.07E-01 | 9.91E-01 | 7.32E-01 | 9.72E-01 | 2.35E-01 | 8.21E-01 |
| *COL11A2* | 6.12E-01 | 9.58E-01 | 1.66E-01 | 7.68E-01 | 1.17E-01 | 7.26E-01 |
| *SPOP* | 5.84E-01 | 9.57E-01 | 4.33E-01 | 9.13E-01 | 5.05E-01 | 9.27E-01 |
| *PLSCR2* | 1.66E-01 | 7.77E-01 | 7.75E-04 | 8.61E-02 | 1.88E-04 | 4.71E-02 |
| *KLF1* | 2.28E-01 | 8.24E-01 | 3.13E-02 | 4.97E-01 | 1.67E-01 | 7.83E-01 |
| *ARL17B* | 9.11E-01 | 9.91E-01 | NA | NA | NA | NA |
| *C6orf48* | 3.57E-01 | 8.92E-01 | 2.76E-01 | 8.42E-01 | 7.73E-01 | 9.73E-01 |
| *RPP21* | 1.39E-01 | 7.47E-01 | 4.74E-02 | 5.74E-01 | 2.63E-02 | 5.11E-01 |
| *RP11-57G10.8* | 1.34E-01 | 7.40E-01 | 7.86E-01 | 9.78E-01 | 8.32E-01 | 9.78E-01 |
| *YLPM1* | NA | NA | NA | NA | NA | NA |
| *GATAD2A* | 3.71E-01 | 9.00E-01 | 7.38E-01 | 9.73E-01 | 5.23E-01 | 9.31E-01 |
| *OAS2* | NA | NA | NA | NA | NA | NA |
| *HEY1* | 1.72E-02 | 4.93E-01 | 5.61E-01 | 9.39E-01 | 2.41E-01 | 8.23E-01 |
| *YY1AP1* | 1.34E-02 | 4.52E-01 | 5.27E-01 | 9.37E-01 | 1.34E-01 | 7.55E-01 |
| *FCHSD2* | 1.91E-01 | 8.02E-01 | 9.87E-01 | 9.99E-01 | 5.11E-01 | 9.30E-01 |
| *MAPT* | NA | NA | NA | NA | NA | NA |
| *GABARAPL2* | 5.69E-01 | 9.55E-01 | 2.75E-01 | 8.42E-01 | 9.19E-01 | 9.90E-01 |
| *CYP4B1* | NA | NA | NA | NA | NA | NA |
| *RP11-91I20.3* | NA | NA | NA | NA | NA | NA |
| *THRA* | 9.73E-01 | 9.98E-01 | 8.77E-01 | 9.88E-01 | 6.19E-01 | 9.52E-01 |
| *EDC3* | NA | NA | NA | NA | NA | NA |
| *SPARC* | NA | NA | NA | NA | NA | NA |
| *NPRL3* | 1.74E-01 | 7.80E-01 | 6.88E-01 | 9.65E-01 | 2.33E-01 | 8.21E-01 |

| | | | | | | |
|---|---|---|---|---|---|---|
| *EPS8L1* | 1.46E-03 | 2.17E-01 | 7.04E-02 | 6.29E-01 | 6.71E-01 | 9.61E-01 |
| *CFB* | NA | NA | NA | NA | NA | NA |
| *AC114752.1* | NA | NA | NA | NA | NA | NA |
| *CCDC18* | 2.11E-01 | 8.12E-01 | 8.02E-01 | 9.81E-01 | 1.41E-01 | 7.63E-01 |
| *LINC01315* | NA | NA | NA | NA | NA | NA |
| *NCOR1* | 1.10E-03 | 1.92E-01 | 1.03E-05 | 5.25E-03 | 9.81E-05 | 2.85E-02 |
| *SNX1* | 6.54E-01 | 9.61E-01 | 8.65E-02 | 6.60E-01 | 9.28E-02 | 7.03E-01 |
| *DNAH8* | NA | NA | NA | NA | NA | NA |
| *RAVER2* | 4.49E-01 | 9.24E-01 | 3.07E-01 | 8.55E-01 | 5.07E-01 | 9.29E-01 |
| *HIST1H2AB* | NA | NA | NA | NA | NA | NA |
| *SQRDL* | 1.47E-01 | 7.56E-01 | 5.49E-04 | 7.18E-02 | 1.66E-01 | 7.83E-01 |
| *FNIP1* | 4.56E-02 | 5.85E-01 | 2.84E-02 | 4.78E-01 | 3.80E-01 | 8.79E-01 |
| *TCF12* | 1.43E-04 | 5.81E-02 | 1.84E-01 | 7.85E-01 | 2.51E-01 | 8.26E-01 |
| *EPO* | NA | NA | NA | NA | NA | NA |
| *CTC-296K1.3* | NA | NA | NA | NA | NA | NA |
| *LAMTOR4* | 7.71E-01 | 9.76E-01 | 5.65E-01 | 9.39E-01 | 3.11E-02 | 5.37E-01 |
| *CTB-50L17.16* | NA | NA | NA | NA | NA | NA |
| *DONSON* | 2.16E-01 | 8.13E-01 | 2.26E-04 | 4.00E-02 | 8.00E-03 | 3.65E-01 |
| *ZPBP2* | NA | NA | NA | NA | NA | NA |
| *NFAM1* | NA | NA | NA | NA | NA | NA |
| *NCDN* | 4.61E-02 | 5.87E-01 | 9.18E-01 | 9.91E-01 | 9.74E-01 | 9.97E-01 |
| *ORMDL3* | 1.65E-04 | 6.29E-02 | 4.90E-05 | 1.43E-02 | 1.04E-02 | 4.05E-01 |
| *ZNF655* | 6.39E-01 | 9.61E-01 | 3.99E-01 | 8.97E-01 | 8.22E-01 | 9.78E-01 |
| *IBA57* | 6.78E-02 | 6.39E-01 | 3.36E-05 | 1.27E-02 | 8.09E-02 | 6.87E-01 |
| *CALR* | 4.46E-01 | 9.22E-01 | 9.16E-01 | 9.91E-01 | 8.44E-01 | 9.79E-01 |
| *CPOX* | 8.36E-01 | 9.84E-01 | 2.32E-01 | 8.17E-01 | 5.69E-01 | 9.44E-01 |
| *FAT2* | 5.02E-02 | 5.97E-01 | 9.14E-01 | 9.91E-01 | 7.61E-01 | 9.73E-01 |
| *ST3GAL3* | 9.39E-01 | 9.94E-01 | 1.02E-02 | 3.28E-01 | 1.02E-02 | 4.02E-01 |
| *DNAJB14* | 6.24E-01 | 9.59E-01 | 2.78E-01 | 8.42E-01 | 7.42E-01 | 9.70E-01 |
| *LINC01565* | NA | NA | NA | NA | NA | NA |
| *SNRPD2* | 4.05E-03 | 3.22E-01 | 1.09E-05 | 5.33E-03 | 2.33E-05 | 1.02E-02 |
| *GNB2* | 4.22E-02 | 5.78E-01 | 1.05E-01 | 6.93E-01 | 8.94E-02 | 6.97E-01 |
| *TSACC* | 2.30E-01 | 8.26E-01 | 4.80E-02 | 5.79E-01 | 1.79E-01 | 7.97E-01 |
| *DCST1* | 2.36E-02 | 5.16E-01 | 5.32E-03 | 2.36E-01 | 5.44E-03 | 3.06E-01 |
| *6-Mar* | 2.60E-01 | 8.50E-01 | 8.01E-01 | 9.81E-01 | 3.67E-01 | 8.74E-01 |
| *TRAF3* | 4.29E-03 | 3.22E-01 | 1.07E-01 | 6.96E-01 | 4.10E-02 | 5.80E-01 |
| *ZMYM4* | 2.15E-04 | 7.31E-02 | 1.27E-03 | 1.13E-01 | 4.02E-03 | 2.72E-01 |
| *AQP4* | NA | NA | NA | NA | NA | NA |
| *ARID3B* | 9.22E-02 | 6.84E-01 | 2.29E-03 | 1.54E-01 | 3.75E-06 | 2.41E-03 |
| *AP1M2* | 4.62E-01 | 9.30E-01 | 5.08E-01 | 9.34E-01 | 4.23E-01 | 8.97E-01 |
| *FAM189B* | 3.70E-01 | 8.99E-01 | 1.32E-01 | 7.34E-01 | 6.49E-03 | 3.32E-01 |
| *ADAMTS8* | NA | NA | NA | NA | NA | NA |
| *MICA* | 9.35E-02 | 6.84E-01 | 6.89E-05 | 1.75E-02 | 9.76E-05 | 2.85E-02 |
| *PRSS33* | NA | NA | NA | NA | NA | NA |

| | | | | | | |
|---|---|---|---|---|---|---|
| *DAP3* | 8.78E-01 | 9.87E-01 | 1.30E-01 | 7.34E-01 | 2.69E-03 | 2.30E-01 |
| *LINC01149* | NA | NA | NA | NA | NA | NA |
| *TAF6* | 5.81E-02 | 6.12E-01 | 6.87E-04 | 8.07E-02 | 5.37E-02 | 6.22E-01 |
| *GPANK1* | 2.91E-02 | 5.31E-01 | 8.68E-02 | 6.61E-01 | 6.72E-03 | 3.39E-01 |
| *LINC00310* | 6.18E-01 | 9.59E-01 | 3.93E-01 | 8.94E-01 | 6.08E-01 | 9.52E-01 |
| *RP11-503G7.2* | NA | NA | NA | NA | NA | NA |
| *FRYL* | NA | NA | NA | NA | NA | NA |
| *E2F1* | NA | NA | NA | NA | NA | NA |
| *PRRT1* | 2.66E-01 | 8.53E-01 | 2.61E-02 | 4.68E-01 | 2.68E-02 | 5.11E-01 |
| *IRF7* | 1.68E-01 | 7.77E-01 | 3.86E-01 | 8.93E-01 | 5.00E-01 | 9.26E-01 |
| *LINC00857* | NA | NA | NA | NA | NA | NA |
| *CERCAM* | NA | NA | NA | NA | NA | NA |
| *ZSWIM7* | NA | NA | NA | NA | NA | NA |
| *ADRA1A* | NA | NA | NA | NA | NA | NA |
| *NPY4R* | NA | NA | NA | NA | NA | NA |
| *TRMT61B* | 3.04E-03 | 2.80E-01 | 7.86E-05 | 1.85E-02 | 9.96E-02 | 7.07E-01 |
| *GJC2* | NA | NA | NA | NA | NA | NA |
| *PSMB2* | 3.26E-01 | 8.79E-01 | 4.12E-02 | 5.38E-01 | 2.60E-01 | 8.34E-01 |
| *PTGFR* | 6.98E-01 | 9.65E-01 | 2.56E-01 | 8.31E-01 | 4.30E-01 | 8.99E-01 |
| *VPS29* | 5.58E-01 | 9.52E-01 | 9.24E-02 | 6.71E-01 | 1.07E-01 | 7.16E-01 |
| *LINC00348* | NA | NA | NA | NA | NA | NA |
| *BRD2* | 3.05E-03 | 2.80E-01 | 1.04E-02 | 3.29E-01 | 8.73E-01 | 9.83E-01 |
| *UPF2* | NA | NA | NA | NA | NA | NA |
| *CHCHD7* | 1.30E-01 | 7.35E-01 | 1.61E-02 | 3.91E-01 | 1.47E-02 | 4.44E-01 |
| *HIST1H2BB* | NA | NA | NA | NA | NA | NA |
| *HSPE1* | 9.47E-01 | 9.96E-01 | 1.31E-01 | 7.34E-01 | 3.52E-03 | 2.57E-01 |
| *RP11-499E18.1* | NA | NA | NA | NA | NA | NA |
| *GCDH* | 1.41E-01 | 7.50E-01 | 8.09E-01 | 9.82E-01 | 9.38E-01 | 9.92E-01 |
| *ZNF146* | 4.20E-01 | 9.14E-01 | 2.51E-01 | 8.27E-01 | 6.14E-01 | 9.52E-01 |
| *CYP2C19* | NA | NA | NA | NA | NA | NA |
| *BNIP1* | 4.88E-01 | 9.37E-01 | 7.15E-02 | 6.33E-01 | 6.54E-01 | 9.57E-01 |
| *DR1* | 7.12E-02 | 6.46E-01 | 2.25E-01 | 8.11E-01 | 9.76E-01 | 9.97E-01 |
| *SUCLG2* | 8.37E-01 | 9.84E-01 | 9.91E-01 | 9.99E-01 | 3.57E-01 | 8.74E-01 |
| *TADA1* | 8.16E-01 | 9.81E-01 | 6.93E-01 | 9.67E-01 | 5.08E-01 | 9.29E-01 |
| *PRM3* | NA | NA | NA | NA | NA | NA |
| *PREX1* | 9.92E-01 | 9.99E-01 | 7.54E-01 | 9.75E-01 | 7.30E-01 | 9.67E-01 |
| *FAM96A* | NA | NA | NA | NA | NA | NA |
| *WFDC10A* | NA | NA | NA | NA | NA | NA |
| *AIP* | NA | NA | NA | NA | NA | NA |
| *PNRC1* | 2.89E-01 | 8.66E-01 | 5.26E-02 | 5.93E-01 | 2.52E-03 | 2.20E-01 |
| *XRCC3* | 1.92E-02 | 4.98E-01 | 4.38E-04 | 6.30E-02 | 7.98E-04 | 1.14E-01 |
| *LPCAT2* | 5.90E-01 | 9.57E-01 | 5.74E-01 | 9.40E-01 | 4.30E-01 | 8.99E-01 |
| *ATG16L2* | NA | NA | NA | NA | NA | NA |
| *ACSM4* | NA | NA | NA | NA | NA | NA |

| | | | | | | |
|---|---|---|---|---|---|---|
| *ZNF311* | NA | NA | NA | NA | NA | NA |
| *HNMT* | 4.25E-02 | 5.78E-01 | 2.20E-02 | 4.51E-01 | 1.57E-01 | 7.81E-01 |
| *PDLIM4* | 7.95E-01 | 9.79E-01 | 9.35E-01 | 9.92E-01 | 7.51E-01 | 9.72E-01 |
| *ILF3-AS1* | 7.35E-03 | 3.89E-01 | 9.33E-01 | 9.92E-01 | 9.53E-01 | 9.94E-01 |
| *PPP1R2* | 1.97E-01 | 8.05E-01 | 5.24E-01 | 9.36E-01 | 4.11E-01 | 8.92E-01 |
| *REEP4* | 1.94E-01 | 8.03E-01 | 5.65E-01 | 9.39E-01 | 7.88E-01 | 9.74E-01 |
| *RPN1* | 9.30E-01 | 9.94E-01 | 6.12E-01 | 9.48E-01 | 1.69E-01 | 7.85E-01 |
| *LPIN3* | NA | NA | NA | NA | NA | NA |
| *ARHGEF38* | NA | NA | NA | NA | NA | NA |
| *AP4M1* | 1.94E-02 | 5.02E-01 | 5.22E-04 | 6.97E-02 | 2.19E-02 | 5.01E-01 |
| *ZSCAN23* | NA | NA | NA | NA | NA | NA |
| *ZNF84* | 3.52E-01 | 8.92E-01 | 2.27E-02 | 4.52E-01 | 2.93E-02 | 5.27E-01 |
| *LMAN1L* | 2.27E-01 | 8.24E-01 | 5.79E-01 | 9.41E-01 | 6.76E-01 | 9.62E-01 |
| *PROB1* | 7.17E-04 | 1.53E-01 | 6.90E-02 | 6.26E-01 | 1.82E-02 | 4.72E-01 |
| *RP11-16E12.2* | 3.40E-01 | 8.86E-01 | 5.69E-02 | 5.95E-01 | 6.58E-02 | 6.56E-01 |
| *BMP8A* | 4.17E-02 | 5.78E-01 | 2.51E-01 | 8.27E-01 | 1.40E-02 | 4.42E-01 |
| *GSDMB* | 1.38E-03 | 2.11E-01 | 3.43E-05 | 1.27E-02 | 6.14E-03 | 3.25E-01 |
| *UNC5C* | NA | NA | NA | NA | NA | NA |
| *RASSF3* | 2.73E-01 | 8.56E-01 | 7.73E-04 | 8.61E-02 | 3.66E-01 | 8.74E-01 |
| *IMPG1* | NA | NA | NA | NA | NA | NA |
| *NDUFC1* | 6.95E-01 | 9.65E-01 | 7.10E-01 | 9.68E-01 | 8.85E-01 | 9.84E-01 |
| *DKFZP434L187* | NA | NA | NA | NA | NA | NA |
| *FAM126A* | 7.44E-01 | 9.71E-01 | 1.91E-01 | 7.92E-01 | 3.41E-01 | 8.73E-01 |
| *CPLX3* | 7.64E-02 | 6.54E-01 | 1.00E-01 | 6.83E-01 | 9.50E-01 | 9.93E-01 |
| *IKZF3* | 6.26E-04 | 1.42E-01 | 1.24E-04 | 2.53E-02 | 1.99E-03 | 2.06E-01 |
| *KIAA0907* | 3.10E-01 | 8.75E-01 | 7.86E-01 | 9.78E-01 | 2.03E-03 | 2.07E-01 |
| *ZKSCAN4* | 3.18E-01 | 8.76E-01 | 5.80E-02 | 5.99E-01 | 3.18E-01 | 8.65E-01 |
| *LKAAEAR1* | 1.29E-01 | 7.35E-01 | 4.84E-04 | 6.72E-02 | 1.83E-01 | 8.01E-01 |
| *DYNC1H1* | NA | NA | NA | NA | NA | NA |
| *GNGT2* | 2.93E-02 | 5.31E-01 | 9.79E-02 | 6.76E-01 | 7.03E-01 | 9.65E-01 |
| *CDC42EP3* | 3.86E-01 | 9.05E-01 | 4.54E-01 | 9.18E-01 | 1.90E-01 | 8.03E-01 |
| *NAA20* | 2.71E-01 | 8.55E-01 | 3.27E-01 | 8.68E-01 | 3.10E-01 | 8.62E-01 |
| *MS4A4A* | 3.44E-01 | 8.86E-01 | 4.55E-01 | 9.18E-01 | 1.71E-01 | 7.87E-01 |
| *OBSCN* | NA | NA | NA | NA | NA | NA |
| *C1orf145* | 3.32E-02 | 5.46E-01 | 7.48E-05 | 1.79E-02 | 1.83E-02 | 4.73E-01 |
| *CDH16* | NA | NA | NA | NA | NA | NA |
| *USP49* | 4.36E-02 | 5.78E-01 | 6.65E-03 | 2.70E-01 | 7.65E-01 | 9.73E-01 |
| *SNX19* | 1.70E-01 | 7.77E-01 | 1.96E-02 | 4.24E-01 | 2.09E-01 | 8.14E-01 |
| *FBXL18* | 3.42E-01 | 8.86E-01 | 5.04E-01 | 9.32E-01 | 9.97E-01 | 9.99E-01 |
| *VWA7* | 1.87E-01 | 7.99E-01 | 7.73E-02 | 6.46E-01 | 6.65E-02 | 6.57E-01 |
| *NAA15* | 7.16E-01 | 9.66E-01 | 2.46E-03 | 1.62E-01 | 6.69E-02 | 6.57E-01 |
| *DHX30* | NA | NA | NA | NA | NA | NA |
| *PCMTD1* | 8.23E-01 | 9.81E-01 | 1.01E-01 | 6.85E-01 | 8.62E-02 | 6.94E-01 |
| *HLA-DOA* | 1.62E-01 | 7.71E-01 | 3.66E-03 | 2.05E-01 | 7.66E-02 | 6.79E-01 |
| *RFTN2* | 5.73E-01 | 9.56E-01 | 7.51E-01 | 9.75E-01 | 6.76E-02 | 6.60E-01 |

| | | | | | | |
|---|---|---|---|---|---|---|
| *ALKBH1* | NA | NA | NA | NA | NA | NA |
| *LDLR* | 3.46E-01 | 8.88E-01 | 6.90E-01 | 9.66E-01 | 8.61E-01 | 9.82E-01 |
| *DSCAML1* | NA | NA | NA | NA | NA | NA |
| *LCTL* | NA | NA | NA | NA | NA | NA |
| *HINT1* | 9.51E-01 | 9.96E-01 | 2.80E-01 | 8.43E-01 | 3.00E-02 | 5.30E-01 |
| *MAP2* | 8.91E-03 | 4.15E-01 | 7.15E-02 | 6.33E-01 | 7.06E-01 | 9.65E-01 |
| *STAG3* | 2.81E-03 | 2.70E-01 | 9.35E-05 | 2.08E-02 | 2.42E-03 | 2.16E-01 |
| *IRF4* | 3.56E-02 | 5.55E-01 | 3.15E-04 | 5.00E-02 | 5.89E-02 | 6.37E-01 |
| *PKP2* | 1.83E-01 | 7.93E-01 | 6.11E-01 | 9.48E-01 | 5.52E-01 | 9.39E-01 |
| *EPB41L2* | 8.49E-01 | 9.85E-01 | 3.38E-03 | 1.96E-01 | 3.18E-01 | 8.65E-01 |
| *MSH5* | 7.33E-01 | 9.69E-01 | 4.22E-02 | 5.43E-01 | 8.36E-01 | 9.79E-01 |

Note: NA represents not applicable.

**Supplementary Table S10. The S-MultiXcan-identified 438 risk genes validated by MAGMA-based gene-level association analysis**

| Gene Name | Susceptible COVID-19 (P-value) | Susceptible COVID-19 (FDR) | Hospitalized COVID-19 (P-value) | Hospitalized COVID-19 (FDR) | Very severe COVID-19 (P-value) | Very severe COVID-19 (FDR) |
|---|---|---|---|---|---|---|
| *CCR9* | 2.18E-35 | 7.18E-32 | 8.56E-57 | 5.63E-53 | 3.60E-60 | 1.78E-56 |
| *SLC6A20* | 1.10E-96 | 2.17E-92 | 1.03E-74 | 1.02E-70 | 1.09E-75 | 1.08E-71 |
| *CXCR6* | 1.84E-30 | 3.64E-27 | 4.00E-44 | 1.13E-40 | 5.71E-46 | 1.41E-42 |
| *XCR1* | 3.14E-26 | 4.78E-23 | 3.68E-54 | 1.45E-50 | 1.60E-55 | 6.33E-52 |
| *LZTFL1* | 8.49E-64 | 8.39E-60 | 2.68E-75 | 5.29E-71 | 7.98E-78 | 1.58E-73 |
| *ABO* | 4.14E-47 | 2.73E-43 | 2.33E-13 | 1.92E-10 | 3.91E-09 | 1.00E-06 |
| *CCR3* | 1.66E-18 | 1.93E-15 | 2.19E-43 | 5.40E-40 | 2.12E-49 | 5.99E-46 |
| *FYCO1* | 7.75E-37 | 3.06E-33 | 2.90E-56 | 1.43E-52 | 1.38E-61 | 9.09E-58 |
| *DPP9* | 5.92E-16 | 6.50E-13 | 2.33E-23 | 3.54E-20 | 2.16E-25 | 3.28E-22 |
| *IFNAR2* | 5.74E-19 | 7.09E-16 | 1.28E-51 | 4.21E-48 | 6.94E-51 | 2.29E-47 |
| *CCR1* | 2.91E-13 | 1.69E-10 | 5.03E-39 | 1.10E-35 | 7.10E-40 | 1.56E-36 |
| *CCR5* | 4.41E-11 | 1.82E-08 | 3.41E-27 | 5.61E-24 | 1.55E-30 | 2.79E-27 |
| *CCR2* | 5.88E-13 | 3.19E-10 | 7.70E-32 | 1.52E-28 | 6.48E-36 | 1.28E-32 |
| *IL10RB* | 4.85E-10 | 1.59E-07 | 1.01E-27 | 1.81E-24 | 2.13E-28 | 3.51E-25 |
| *CCRL2* | 1.19E-05 | 1.67E-03 | 4.53E-16 | 5.59E-13 | 8.40E-18 | 1.11E-14 |
| *NFKBIZ* | 2.03E-22 | 2.87E-19 | 1.86E-07 | 3.06E-05 | 5.98E-03 | 1.77E-01 |
| *NXPE3* | 7.59E-40 | 3.75E-36 | 7.51E-14 | 6.74E-11 | 2.17E-06 | 3.02E-04 |
| *MTX1* | 7.16E-12 | 3.45E-09 | 7.38E-10 | 1.80E-07 | 1.70E-08 | 3.82E-06 |
| *CEP97* | 5.77E-33 | 1.63E-29 | 1.19E-12 | 7.12E-10 | 2.66E-05 | 2.68E-03 |
| *FOXP4* | 1.02E-03 | 5.84E-02 | 2.71E-06 | 3.30E-04 | 4.95E-05 | 4.44E-03 |
| *MUC1* | 7.93E-14 | 5.60E-11 | 6.84E-14 | 6.43E-11 | 5.17E-14 | 6.01E-11 |
| *ZBTB11* | 6.69E-30 | 1.20E-26 | 1.97E-12 | 1.11E-09 | 1.41E-05 | 1.53E-03 |
| *ASH1L* | 2.50E-04 | 2.10E-02 | 4.04E-07 | 6.13E-05 | 1.41E-08 | 3.20E-06 |
| *AP000569.9* | NA | NA | NA | NA | NA | NA |
| *CACFD1* | 1.90E-19 | 2.50E-16 | 4.23E-03 | 1.06E-01 | 1.91E-02 | 3.49E-01 |
| *TRIM46* | 1.43E-13 | 9.75E-11 | 4.30E-12 | 2.02E-09 | 3.30E-12 | 2.17E-09 |
| *ATP5O* | 7.96E-07 | 1.64E-04 | 3.65E-14 | 3.60E-11 | 3.14E-12 | 2.17E-09 |
| *NAPSA* | 3.18E-09 | 9.53E-07 | 2.99E-14 | 3.11E-11 | 2.69E-10 | 8.78E-08 |

| | | | | | | |
|---|---|---|---|---|---|---|
| *TULP2* | 3.05E-14 | 2.41E-11 | 1.34E-04 | 8.16E-03 | 1.92E-04 | 1.35E-02 |
| *PCNP* | 2.40E-14 | 1.98E-11 | 1.11E-06 | 1.55E-04 | 2.41E-02 | 3.92E-01 |
| *MRPS6* | 7.48E-02 | 5.62E-01 | 7.90E-04 | 3.26E-02 | 5.88E-02 | 5.71E-01 |
| *SENP7* | 1.41E-10 | 5.36E-08 | 1.86E-03 | 5.98E-02 | 3.96E-01 | 8.85E-01 |
| *LINC02009* | NA | NA | NA | NA | NA | NA |
| *TYK2* | 3.12E-03 | 1.24E-01 | 7.69E-10 | 1.85E-07 | 1.53E-10 | 5.60E-08 |
| *FUT2* | 2.73E-13 | 1.64E-10 | 1.35E-08 | 2.67E-06 | 1.02E-09 | 2.84E-07 |
| *MED22* | 4.91E-10 | 1.59E-07 | 5.35E-02 | 4.52E-01 | 8.22E-02 | 6.35E-01 |
| *SLC50A1* | 7.57E-12 | 3.52E-09 | 1.44E-06 | 1.95E-04 | 2.24E-07 | 4.10E-05 |
| *ELF5* | 2.16E-08 | 5.93E-06 | 5.95E-17 | 7.83E-14 | 3.86E-15 | 4.77E-12 |
| *CAT* | 1.15E-06 | 2.25E-04 | 3.24E-12 | 1.64E-09 | 9.74E-11 | 3.85E-08 |
| *LTF* | 1.75E-06 | 3.30E-04 | 1.69E-22 | 2.38E-19 | 1.90E-22 | 2.68E-19 |
| *C6orf15* | 1.47E-04 | 1.41E-02 | 1.16E-07 | 1.99E-05 | 1.72E-08 | 3.82E-06 |
| *SLC2A6* | 2.24E-14 | 1.98E-11 | 3.17E-02 | 3.54E-01 | 9.70E-02 | 6.72E-01 |
| *ADAM15* | 8.30E-08 | 2.03E-05 | 1.26E-12 | 7.32E-10 | 3.20E-12 | 2.17E-09 |
| *PLEKHA4* | 6.52E-16 | 6.78E-13 | 1.09E-05 | 1.09E-03 | 1.25E-04 | 9.41E-03 |
| *TNFAIP8L1* | 7.76E-08 | 1.94E-05 | 5.52E-09 | 1.15E-06 | 9.58E-10 | 2.74E-07 |
| *NTN5* | 5.52E-06 | 8.40E-04 | 7.42E-03 | 1.54E-01 | 2.74E-03 | 1.03E-01 |
| *ATP11A* | 1.44E-01 | 6.73E-01 | 9.50E-07 | 1.37E-04 | 3.51E-07 | 6.09E-05 |
| *ADAMTS13* | 4.44E-15 | 4.18E-12 | 4.68E-02 | 4.28E-01 | 3.47E-02 | 4.60E-01 |
| *EFNA4* | 1.98E-08 | 5.51E-06 | 6.07E-13 | 4.13E-10 | 5.70E-13 | 4.90E-10 |
| *RP11-798G7.6* | NA | NA | NA | NA | NA | NA |
| *PPP1R15A* | 6.74E-14 | 4.94E-11 | 6.68E-05 | 4.74E-03 | 9.11E-05 | 7.26E-03 |
| *GON4L* | 6.47E-06 | 9.62E-04 | 9.23E-03 | 1.77E-01 | 2.56E-05 | 2.62E-03 |
| *FBRSL1* | 7.87E-08 | 1.94E-05 | 4.32E-13 | 3.16E-10 | 8.88E-12 | 5.16E-09 |
| *RASIP1* | 1.15E-15 | 1.14E-12 | 3.88E-09 | 8.61E-07 | 2.10E-11 | 1.05E-08 |
| *CCHCR1* | 4.48E-08 | 1.18E-05 | 1.52E-11 | 6.25E-09 | 2.28E-11 | 1.05E-08 |
| *NUCB1* | 2.40E-11 | 1.03E-08 | 2.93E-03 | 8.14E-02 | 8.39E-03 | 2.17E-01 |
| *MAMSTR* | 2.39E-14 | 1.98E-11 | 5.04E-10 | 1.29E-07 | 2.26E-11 | 1.05E-08 |
| *KRTCAP2* | 5.97E-13 | 3.19E-10 | 2.82E-09 | 6.33E-07 | 1.06E-09 | 2.91E-07 |
| *OAS3* | 7.66E-12 | 3.52E-09 | 1.93E-14 | 2.13E-11 | 4.72E-10 | 1.48E-07 |
| *FDX2* | NA | NA | NA | NA | NA | NA |
| *PDE4A* | 1.56E-02 | 3.10E-01 | 9.30E-06 | 9.41E-04 | 1.34E-07 | 2.55E-05 |
| *PLEKHM1* | 6.15E-03 | 1.87E-01 | 2.40E-10 | 6.77E-08 | 2.71E-10 | 8.78E-08 |
| *FUT1* | 2.01E-13 | 1.24E-10 | 4.38E-06 | 4.91E-04 | 8.08E-10 | 2.46E-07 |
| *SFTPD* | 1.29E-07 | 3.04E-05 | 4.75E-10 | 1.25E-07 | 9.74E-10 | 2.75E-07 |
| *ZGLP1* | 5.35E-04 | 3.75E-02 | 1.04E-10 | 3.31E-08 | 2.07E-12 | 1.57E-09 |
| *TCF19* | 2.34E-08 | 6.34E-06 | 4.04E-11 | 1.48E-08 | 7.47E-12 | 4.47E-09 |
| *C5orf56* | NA | NA | NA | NA | NA | NA |
| *HLA-C* | 7.26E-04 | 4.61E-02 | 1.44E-05 | 1.34E-03 | 2.48E-07 | 4.47E-05 |
| *ARHGAP27* | 4.70E-04 | 3.39E-02 | 9.96E-13 | 6.14E-10 | 8.86E-11 | 3.57E-08 |
| *SLC35B1* | 1.78E-02 | 3.25E-01 | 1.69E-07 | 2.80E-05 | 2.93E-05 | 2.91E-03 |
| *DDX39B* | 1.54E-03 | 7.54E-02 | 6.30E-11 | 2.22E-08 | 2.41E-09 | 6.44E-07 |
| *PSORS1C2* | 1.12E-08 | 3.16E-06 | 6.34E-13 | 4.17E-10 | 3.42E-12 | 2.18E-09 |
| *IZUMO1* | 1.85E-13 | 1.18E-10 | 1.45E-06 | 1.95E-04 | 2.12E-10 | 7.17E-08 |
| *FAM215B* | NA | NA | NA | NA | NA | NA |

| | | | | | | |
|---|---|---|---|---|---|---|
| *REXO4* | 1.77E-07 | 4.02E-05 | 3.08E-01 | 8.12E-01 | 1.51E-01 | 7.58E-01 |
| *ACSF3* | 3.74E-02 | 4.39E-01 | 3.11E-08 | 5.74E-06 | 1.33E-10 | 4.96E-08 |
| *RAVER1* | 1.42E-04 | 1.38E-02 | 2.46E-11 | 9.34E-09 | 1.28E-11 | 7.23E-09 |
| *ICAM5* | 1.08E-04 | 1.09E-02 | 1.06E-11 | 4.55E-09 | 1.22E-13 | 1.27E-10 |
| *OAS1* | 1.31E-11 | 5.89E-09 | 1.94E-14 | 2.13E-11 | 1.72E-10 | 5.96E-08 |
| *ICAM3* | 5.26E-04 | 3.70E-02 | 1.09E-10 | 3.42E-08 | 1.71E-10 | 5.96E-08 |
| *ZCCHC4* | 5.26E-04 | 3.70E-02 | 5.40E-10 | 1.37E-07 | 3.44E-05 | 3.33E-03 |
| *KCNC3* | 4.81E-08 | 1.25E-05 | 3.13E-07 | 4.94E-05 | 1.07E-04 | 8.35E-03 |
| *NPNT* | 4.57E-01 | 8.58E-01 | 9.09E-03 | 1.77E-01 | 1.59E-03 | 7.23E-02 |
| *WNT3* | 9.06E-02 | 5.97E-01 | 3.32E-13 | 2.62E-10 | 2.42E-13 | 2.28E-10 |
| *DCST2* | 1.39E-03 | 7.03E-02 | 2.13E-08 | 4.00E-06 | 1.30E-08 | 3.02E-06 |
| *HLA-A* | 3.08E-01 | 8.07E-01 | 5.87E-06 | 6.37E-04 | 4.10E-08 | 8.53E-06 |
| *HSD17B14* | 1.89E-11 | 8.30E-09 | 3.68E-04 | 1.79E-02 | 3.75E-03 | 1.27E-01 |
| *PI4K2B* | 2.73E-04 | 2.22E-02 | 1.94E-07 | 3.17E-05 | 5.34E-04 | 3.15E-02 |
| *CSF3* | 2.43E-10 | 8.74E-08 | 1.30E-05 | 1.25E-03 | 2.35E-06 | 3.25E-04 |
| *RPL7A* | 5.54E-10 | 1.74E-07 | 4.28E-02 | 4.11E-01 | 7.70E-02 | 6.20E-01 |
| *SEMA6B* | 1.41E-01 | 6.69E-01 | 3.70E-02 | 3.84E-01 | 3.39E-05 | 3.32E-03 |
| *SFTPA2* | 1.21E-06 | 2.35E-04 | 6.76E-10 | 1.67E-07 | 8.20E-10 | 2.46E-07 |
| *CDH15* | 2.52E-02 | 3.78E-01 | 2.45E-10 | 6.81E-08 | 4.24E-11 | 1.75E-08 |
| *RP11-589B3.6* | NA | NA | NA | NA | NA | NA |
| *LINC01301* | NA | NA | NA | NA | NA | NA |
| *THBS3* | 5.04E-13 | 2.85E-10 | 4.14E-12 | 1.99E-09 | 1.45E-12 | 1.15E-09 |
| *RAB2A* | 1.34E-05 | 1.80E-03 | 2.40E-11 | 9.29E-09 | 2.15E-08 | 4.72E-06 |
| *SACM1L* | 3.15E-02 | 4.14E-01 | 4.87E-10 | 1.26E-07 | 1.05E-08 | 2.50E-06 |
| *IBA57-AS1* | NA | NA | NA | NA | NA | NA |
| *MED24* | 1.23E-10 | 4.77E-08 | 3.31E-05 | 2.69E-03 | 4.26E-06 | 5.50E-04 |
| *DPF2* | 3.30E-01 | 8.18E-01 | 4.02E-02 | 3.98E-01 | 4.45E-01 | 8.94E-01 |
| *GSDMA* | 2.31E-10 | 8.46E-08 | 1.18E-06 | 1.63E-04 | 9.75E-07 | 1.48E-04 |
| *PMVK* | 2.39E-04 | 2.03E-02 | 5.77E-05 | 4.30E-03 | 4.96E-05 | 4.44E-03 |
| *PSMD3* | 2.83E-10 | 9.65E-08 | 1.46E-06 | 1.95E-04 | 8.15E-07 | 1.27E-04 |
| *RP11-506M13.3* | NA | NA | NA | NA | NA | NA |
| *NUTM2E* | NA | NA | NA | NA | NA | NA |
| *CSK* | 9.65E-03 | 2.40E-01 | 1.36E-05 | 1.28E-03 | 2.30E-02 | 3.84E-01 |
| *RP4-535B20.1* | NA | NA | NA | NA | NA | NA |
| *MICB* | 3.44E-03 | 1.31E-01 | 4.34E-09 | 9.31E-07 | 1.97E-07 | 3.64E-05 |
| *RTP3* | 5.37E-03 | 1.71E-01 | 1.42E-11 | 5.96E-09 | 1.43E-11 | 7.85E-09 |
| *RP11-387H17.6* | NA | NA | NA | NA | NA | NA |
| *ARL17A* | 3.11E-01 | 8.09E-01 | 2.73E-04 | 1.43E-02 | 9.11E-02 | 6.55E-01 |
| *HLA-DPA1* | 8.36E-09 | 2.40E-06 | 4.70E-06 | 5.21E-04 | 8.94E-03 | 2.24E-01 |
| *PSORS1C1* | 2.67E-07 | 5.87E-05 | 3.96E-12 | 1.95E-09 | 8.03E-13 | 6.61E-10 |
| *RP3-462D8.2* | NA | NA | NA | NA | NA | NA |
| *MEF2D* | 6.70E-04 | 4.36E-02 | 4.39E-01 | 8.57E-01 | 8.53E-01 | 9.12E-01 |
| *ICAM1* | 7.39E-04 | 4.65E-02 | 4.17E-10 | 1.13E-07 | 5.46E-13 | 4.90E-10 |

| | | | | | | |
|---|---|---|---|---|---|---|
| *RP11-11C20.3* | NA | NA | NA | NA | NA | NA |
| *RPL24* | 7.03E-29 | 1.16E-25 | 8.99E-12 | 3.94E-09 | 9.04E-05 | 7.23E-03 |
| *SLC22A4* | 1.67E-02 | 3.16E-01 | 2.29E-06 | 2.84E-04 | 4.82E-04 | 2.96E-02 |
| *GBA* | 7.16E-04 | 4.60E-02 | 5.20E-03 | 1.21E-01 | 1.10E-04 | 8.49E-03 |
| *MCM7* | 7.51E-03 | 2.10E-01 | 3.40E-05 | 2.75E-03 | 4.54E-05 | 4.19E-03 |
| *COQ10B* | 4.98E-01 | 8.63E-01 | 2.01E-06 | 2.59E-04 | 1.53E-04 | 1.12E-02 |
| *LTA* | 4.65E-03 | 1.59E-01 | 1.74E-09 | 3.95E-07 | 4.59E-07 | 7.75E-05 |
| *IMPG2* | 1.16E-09 | 3.58E-07 | 3.27E-03 | 8.88E-02 | 3.83E-01 | 8.81E-01 |
| *LRRC37A2* | 2.40E-01 | 7.71E-01 | 2.73E-04 | 1.43E-02 | 8.56E-02 | 6.46E-01 |
| *DBP* | 3.98E-06 | 6.56E-04 | 2.41E-02 | 3.09E-01 | 1.42E-02 | 2.92E-01 |
| *CLK3* | 2.94E-01 | 7.99E-01 | 4.96E-03 | 1.18E-01 | 2.31E-05 | 2.43E-03 |
| *ZNF3* | 1.98E-03 | 8.92E-02 | 9.78E-07 | 1.40E-04 | 2.96E-06 | 3.95E-04 |
| *ZNF778* | 1.20E-01 | 6.42E-01 | 2.45E-05 | 2.11E-03 | 5.11E-05 | 4.55E-03 |
| *PPP1R11* | 1.89E-01 | 7.27E-01 | 4.87E-02 | 4.33E-01 | 1.32E-02 | 2.79E-01 |
| *SLC5A3* | 4.31E-02 | 4.63E-01 | 5.07E-04 | 2.33E-02 | 2.84E-02 | 4.21E-01 |
| *CDSN* | 1.32E-05 | 1.79E-03 | 3.98E-09 | 8.63E-07 | 4.88E-10 | 1.51E-07 |
| *ACSL6* | 6.74E-05 | 7.44E-03 | 1.09E-06 | 1.55E-04 | 6.27E-05 | 5.48E-03 |
| *PGC* | 2.52E-02 | 3.78E-01 | 1.52E-03 | 5.14E-02 | 1.51E-01 | 7.58E-01 |
| *DSP* | 5.70E-01 | 8.73E-01 | 4.34E-01 | 8.56E-01 | 5.44E-03 | 1.65E-01 |
| *HLA-DQA2* | 7.06E-01 | 8.84E-01 | 6.52E-01 | 8.88E-01 | 1.66E-01 | 7.78E-01 |
| *IFNAR1* | 5.15E-02 | 4.96E-01 | 1.10E-04 | 7.05E-03 | 4.03E-06 | 5.28E-04 |
| *FOXA3* | 4.18E-01 | 8.50E-01 | 6.64E-04 | 2.87E-02 | 7.14E-03 | 1.98E-01 |
| *HLA-DPB1* | 7.57E-09 | 2.20E-06 | 3.88E-06 | 4.45E-04 | 3.53E-03 | 1.23E-01 |
| *DTX1* | 4.79E-02 | 4.83E-01 | 4.29E-02 | 4.12E-01 | 1.62E-02 | 3.18E-01 |
| *MYDGF* | 9.92E-07 | 2.00E-04 | 1.48E-08 | 2.88E-06 | 9.04E-09 | 2.18E-06 |
| *RP11-182L21.6* | NA | NA | NA | NA | NA | NA |
| *EIF4EBP2* | 3.15E-01 | 8.11E-01 | 3.34E-01 | 8.21E-01 | 9.31E-02 | 6.59E-01 |
| *ANAPC4* | 7.74E-04 | 4.84E-02 | 3.93E-09 | 8.62E-07 | 1.48E-05 | 1.60E-03 |
| *SLC22A31* | 3.24E-02 | 4.17E-01 | 1.64E-09 | 3.76E-07 | 2.89E-08 | 6.14E-06 |
| *SPACA4* | 2.63E-05 | 3.27E-03 | 7.44E-02 | 5.22E-01 | 5.47E-02 | 5.56E-01 |
| *TFR2* | 5.16E-10 | 1.65E-07 | 5.57E-04 | 2.50E-02 | 7.09E-04 | 3.93E-02 |
| *HLA-DRA* | 1.51E-01 | 6.81E-01 | 6.68E-03 | 1.44E-01 | 8.15E-03 | 2.12E-01 |
| *ZKSCAN1* | 1.83E-04 | 1.66E-02 | 3.29E-07 | 5.15E-05 | 2.29E-08 | 4.92E-06 |
| *SLC25A44* | 2.93E-05 | 3.62E-03 | 2.93E-03 | 8.14E-02 | 4.37E-04 | 2.72E-02 |
| *PMF1* | 3.57E-05 | 4.25E-03 | 5.14E-03 | 1.20E-01 | 4.33E-04 | 2.71E-02 |
| *MOSPD3* | 2.17E-10 | 8.09E-08 | 2.54E-04 | 1.35E-02 | 7.98E-04 | 4.24E-02 |
| *BYSL* | 1.14E-02 | 2.63E-01 | 7.36E-06 | 7.73E-04 | 1.08E-01 | 6.92E-01 |
| *CTA-384D8.35* | NA | NA | NA | NA | NA | NA |
| *ACTL6B* | 2.26E-09 | 6.87E-07 | 3.78E-03 | 9.82E-02 | 2.58E-03 | 1.00E-01 |
| *HIP1* | 4.06E-01 | 8.46E-01 | 3.92E-02 | 3.94E-01 | 1.31E-04 | 9.83E-03 |
| *EFNA1* | 4.82E-11 | 1.94E-08 | 4.84E-08 | 8.53E-06 | 1.32E-08 | 3.03E-06 |
| *CDC37* | 3.26E-02 | 4.17E-01 | 1.46E-05 | 1.35E-03 | 5.03E-06 | 6.25E-04 |
| *CHD1* | 6.01E-02 | 5.24E-01 | 1.38E-01 | 6.46E-01 | 2.42E-01 | 8.36E-01 |
| *PCOLCE* | 2.49E-10 | 8.79E-08 | 2.39E-04 | 1.29E-02 | 8.42E-04 | 4.43E-02 |

| | | | | | | |
|---|---|---|---|---|---|---|
| *AC006007.1* | NA | NA | NA | NA | NA | NA |
| *CA11* | 1.03E-06 | 2.06E-04 | 8.54E-03 | 1.68E-01 | 5.09E-03 | 1.58E-01 |
| *FUBP1* | 2.01E-01 | 7.39E-01 | 3.04E-01 | 8.10E-01 | 2.27E-01 | 8.28E-01 |
| *SH3BP2* | 2.47E-02 | 3.74E-01 | 1.12E-02 | 2.02E-01 | 1.02E-01 | 6.80E-01 |
| *EFNA3* | 2.84E-11 | 1.19E-08 | 1.68E-11 | 6.75E-09 | 1.29E-10 | 4.90E-08 |
| *C2orf16* | 6.97E-01 | 8.83E-01 | 3.16E-01 | 8.16E-01 | 7.46E-01 | 8.99E-01 |
| *MYL5* | 1.66E-01 | 7.01E-01 | 2.13E-01 | 7.42E-01 | 5.87E-01 | 8.95E-01 |
| *ADGRE3* | 1.07E-01 | 6.29E-01 | 1.05E-01 | 5.90E-01 | 4.30E-02 | 5.05E-01 |
| *CLMP* | 2.51E-01 | 7.79E-01 | 2.88E-01 | 8.00E-01 | 2.56E-02 | 4.01E-01 |
| *HLA-F* | 1.96E-02 | 3.40E-01 | 1.50E-02 | 2.37E-01 | 3.05E-04 | 1.99E-02 |
| *GATS* | 8.34E-04 | 5.08E-02 | 1.66E-06 | 2.18E-04 | 1.52E-03 | 7.01E-02 |
| *FDPS* | 1.85E-01 | 7.22E-01 | 1.07E-03 | 3.97E-02 | 4.58E-06 | 5.80E-04 |
| *ZAN* | 5.40E-06 | 8.28E-04 | 7.04E-03 | 1.49E-01 | 1.83E-01 | 7.96E-01 |
| *TNFRSF4* | 3.83E-01 | 8.41E-01 | 2.41E-01 | 7.67E-01 | 2.62E-01 | 8.48E-01 |
| *BCAT2* | 1.46E-04 | 1.41E-02 | 4.82E-02 | 4.33E-01 | 8.33E-02 | 6.39E-01 |
| *CYP1A1* | 1.42E-01 | 6.69E-01 | 1.64E-03 | 5.44E-02 | 3.19E-04 | 2.05E-02 |
| *TOMM7* | 3.87E-01 | 8.42E-01 | 3.54E-06 | 4.20E-04 | 1.32E-07 | 2.53E-05 |
| *YIPF2* | 2.18E-01 | 7.53E-01 | 1.43E-04 | 8.55E-03 | 5.83E-08 | 1.18E-05 |
| *POU5F1* | 3.84E-08 | 1.03E-05 | 3.08E-11 | 1.15E-08 | 1.26E-10 | 4.88E-08 |
| *P4HA2* | 3.96E-03 | 1.43E-01 | 3.86E-06 | 4.45E-04 | 1.20E-02 | 2.65E-01 |
| *RP11-298J23.8* | NA | NA | NA | NA | NA | NA |
| *DNAJB4* | 2.08E-01 | 7.45E-01 | 2.66E-01 | 7.88E-01 | 1.73E-01 | 7.89E-01 |
| *NLRP4* | 2.82E-01 | 7.94E-01 | 6.27E-01 | 8.84E-01 | 4.37E-01 | 8.92E-01 |
| *FRS3* | 9.09E-04 | 5.35E-02 | 1.10E-05 | 1.10E-03 | 3.62E-02 | 4.69E-01 |
| *GPC2* | 1.22E-02 | 2.72E-01 | 9.09E-05 | 6.17E-03 | 3.84E-03 | 1.29E-01 |
| *AGRN* | 6.68E-02 | 5.39E-01 | 5.13E-02 | 4.42E-01 | 2.02E-02 | 3.61E-01 |
| *COPS6* | 2.86E-03 | 1.16E-01 | 3.80E-06 | 4.41E-04 | 8.52E-06 | 9.91E-04 |
| *ATF6B* | 2.66E-01 | 7.88E-01 | 4.37E-05 | 3.41E-03 | 5.17E-04 | 3.11E-02 |
| *GUK1* | 1.39E-01 | 6.66E-01 | 6.49E-05 | 4.68E-03 | 3.44E-04 | 2.19E-02 |
| *RP11-298J23.10* | NA | NA | NA | NA | NA | NA |
| *CLK2* | 1.82E-01 | 7.21E-01 | 1.23E-02 | 2.11E-01 | 8.74E-06 | 1.01E-03 |
| *RP13-554M15.8* | NA | NA | NA | NA | NA | NA |
| *AZGP1* | 1.58E-03 | 7.65E-02 | 2.92E-03 | 8.14E-02 | 6.82E-07 | 1.11E-04 |
| *HCN3* | 1.45E-01 | 6.76E-01 | 5.62E-03 | 1.28E-01 | 9.42E-06 | 1.08E-03 |
| *SURF2* | 2.81E-10 | 9.65E-08 | 2.68E-02 | 3.30E-01 | 4.89E-02 | 5.31E-01 |
| *JAK1* | 5.62E-02 | 5.09E-01 | 1.26E-07 | 2.13E-05 | 4.91E-09 | 1.23E-06 |
| *SCAMP3* | 1.06E-01 | 6.28E-01 | 7.95E-03 | 1.61E-01 | 1.93E-05 | 2.06E-03 |
| *LINC00304* | NA | NA | NA | NA | NA | NA |
| *ZBTB7B* | 1.79E-03 | 8.25E-02 | 4.49E-06 | 5.01E-04 | 7.25E-06 | 8.69E-04 |
| *ACAD8* | 5.92E-01 | 8.75E-01 | 2.98E-01 | 8.06E-01 | 3.61E-02 | 4.68E-01 |
| *HLA-DMB* | 2.07E-01 | 7.44E-01 | 6.94E-03 | 1.47E-01 | 6.82E-05 | 5.84E-03 |
| *TRIM15* | 4.43E-01 | 8.57E-01 | 8.22E-02 | 5.43E-01 | 6.26E-03 | 1.80E-01 |
| *RNF17* | 3.42E-01 | 8.24E-01 | 5.46E-01 | 8.76E-01 | 4.26E-02 | 5.03E-01 |
| *KISS1* | 3.07E-01 | 8.07E-01 | 3.13E-02 | 3.53E-01 | 3.04E-01 | 8.63E-01 |

| | | | | | | |
|---|---|---|---|---|---|---|
| *ABCF1* | 7.28E-01 | 8.86E-01 | 8.28E-01 | 9.07E-01 | 7.08E-01 | 8.96E-01 |
| *BTNL2* | 3.50E-01 | 8.28E-01 | 2.82E-03 | 7.98E-02 | 6.48E-03 | 1.85E-01 |
| *PKLR* | 3.10E-01 | 8.09E-01 | 3.66E-03 | 9.63E-02 | 8.05E-06 | 9.47E-04 |
| *FAM83E* | 1.04E-04 | 1.06E-02 | 9.49E-02 | 5.71E-01 | 8.75E-02 | 6.51E-01 |
| *TIMM29* | NA | NA | NA | NA | NA | NA |
| *LINC02138* | NA | NA | NA | NA | NA | NA |
| *LTB* | 5.81E-03 | 1.81E-01 | 1.62E-08 | 3.10E-06 | 3.18E-06 | 4.22E-04 |
| *DDR1* | 3.75E-01 | 8.36E-01 | 5.91E-01 | 8.81E-01 | 3.91E-01 | 8.85E-01 |
| *TDGF1* | 3.14E-03 | 1.24E-01 | 1.40E-02 | 2.28E-01 | 2.12E-03 | 8.91E-02 |
| *RSPO2* | 4.94E-02 | 4.90E-01 | 9.07E-05 | 6.17E-03 | 2.91E-02 | 4.25E-01 |
| *SFTPA1* | 6.37E-05 | 7.16E-03 | 5.20E-08 | 9.08E-06 | 3.43E-09 | 8.92E-07 |
| *RIT1* | 1.13E-01 | 6.37E-01 | 9.55E-02 | 5.72E-01 | 4.78E-05 | 4.35E-03 |
| *NUTM2B* | 1.80E-02 | 3.25E-01 | 1.11E-05 | 1.10E-03 | 6.96E-07 | 1.13E-04 |
| *KIAA0319L* | 4.85E-02 | 4.86E-01 | 6.65E-05 | 4.74E-03 | 2.42E-03 | 9.62E-02 |
| *ANHX* | 5.06E-02 | 4.94E-01 | 2.11E-03 | 6.54E-02 | 8.25E-02 | 6.37E-01 |
| *C17orf78* | 5.75E-01 | 8.73E-01 | 2.34E-02 | 3.05E-01 | 1.57E-01 | 7.67E-01 |
| *C1orf35* | 2.97E-01 | 8.01E-01 | 9.98E-06 | 1.01E-03 | 7.59E-07 | 1.20E-04 |
| *RHOF* | 4.81E-01 | 8.60E-01 | 8.09E-02 | 5.41E-01 | 1.74E-01 | 7.91E-01 |
| *SYT11* | 1.14E-05 | 1.61E-03 | 4.74E-03 | 1.15E-01 | 7.75E-06 | 9.17E-04 |
| *PRR15L* | 2.11E-02 | 3.49E-01 | 6.16E-03 | 1.38E-01 | 3.36E-01 | 8.70E-01 |
| *STK19* | 4.36E-02 | 4.64E-01 | 1.92E-04 | 1.07E-02 | 4.44E-03 | 1.42E-01 |
| *NECAB3* | 3.40E-02 | 4.24E-01 | 5.13E-05 | 3.88E-03 | 3.21E-03 | 1.16E-01 |
| *CNPY4* | 1.85E-02 | 3.30E-01 | 4.03E-05 | 3.21E-03 | 2.53E-05 | 2.60E-03 |
| *IFNA6* | 3.23E-04 | 2.53E-02 | 9.59E-05 | 6.31E-03 | 1.18E-04 | 9.02E-03 |
| *DST* | 7.96E-01 | 8.99E-01 | 9.23E-02 | 5.67E-01 | 1.49E-01 | 7.54E-01 |
| *ABTB2* | 4.34E-02 | 4.63E-01 | 1.05E-04 | 6.80E-03 | 4.17E-06 | 5.42E-04 |
| *PRICKLE4* | 1.53E-03 | 7.52E-02 | 1.19E-05 | 1.17E-03 | 2.75E-02 | 4.17E-01 |
| *KLC1* | 2.42E-02 | 3.71E-01 | 2.70E-04 | 1.42E-02 | 6.94E-03 | 1.94E-01 |
| *SPPL2C* | 4.70E-04 | 3.39E-02 | 3.00E-12 | 1.56E-09 | 1.66E-11 | 8.87E-09 |
| *PBXIP1* | 1.05E-06 | 2.08E-04 | 8.91E-07 | 1.29E-04 | 1.37E-06 | 2.04E-04 |
| *PPP1R12C* | 1.23E-05 | 1.70E-03 | 1.64E-01 | 6.80E-01 | 3.80E-01 | 8.81E-01 |
| *MARS2* | 3.56E-01 | 8.30E-01 | 1.56E-04 | 9.14E-03 | 1.19E-04 | 9.04E-03 |
| *UBE2D4* | 7.48E-01 | 8.90E-01 | 7.65E-01 | 8.98E-01 | 3.13E-01 | 8.66E-01 |
| *MARCKSL1* | 6.31E-01 | 8.79E-01 | 1.64E-01 | 6.80E-01 | 9.31E-01 | 9.45E-01 |
| *APOPT1* | 2.98E-02 | 4.04E-01 | 3.76E-05 | 3.01E-03 | 7.52E-05 | 6.27E-03 |
| *DHDDS* | 9.67E-04 | 5.58E-02 | 3.79E-03 | 9.83E-02 | 4.51E-04 | 2.80E-02 |
| *NINJ2* | 1.07E-01 | 6.29E-01 | 1.33E-01 | 6.40E-01 | 2.37E-02 | 3.89E-01 |
| *WFDC2* | 2.52E-02 | 3.78E-01 | 2.58E-01 | 7.82E-01 | 8.08E-01 | 9.03E-01 |
| *VIPR2* | 4.44E-01 | 8.57E-01 | 8.45E-01 | 9.11E-01 | 8.39E-01 | 9.09E-01 |
| *GATA6-AS1* | NA | NA | NA | NA | NA | NA |
| *BMP1* | 1.04E-02 | 2.50E-01 | 6.11E-07 | 8.93E-05 | 2.90E-04 | 1.92E-02 |
| *HLA-B* | 6.22E-03 | 1.88E-01 | 1.11E-06 | 1.55E-04 | 6.36E-06 | 7.71E-04 |
| *BGLAP* | 6.42E-05 | 7.17E-03 | 3.42E-03 | 9.17E-02 | 6.21E-04 | 3.59E-02 |
| *KANSL1* | 4.53E-03 | 1.56E-01 | 1.59E-10 | 4.68E-08 | 3.96E-09 | 1.00E-06 |
| *NELFE* | 9.98E-02 | 6.16E-01 | 8.56E-04 | 3.44E-02 | 8.19E-03 | 2.13E-01 |
| *RGS9* | 1.19E-01 | 6.42E-01 | 5.35E-01 | 8.75E-01 | 7.84E-01 | 9.00E-01 |

| | | | | | | |
|---|---|---|---|---|---|---|
| *SPEF2* | 3.46E-02 | 4.25E-01 | 1.95E-03 | 6.19E-02 | 1.60E-01 | 7.73E-01 |
| *CTD-3010D24.3* | NA | NA | NA | NA | NA | NA |
| *ZSCAN21* | 5.01E-04 | 3.56E-02 | 3.63E-08 | 6.64E-06 | 4.03E-07 | 6.87E-05 |
| *CAMSAP2* | 1.80E-01 | 7.17E-01 | 4.91E-02 | 4.34E-01 | 2.95E-03 | 1.08E-01 |
| *UBE2L6* | 3.03E-02 | 4.06E-01 | 1.75E-01 | 6.97E-01 | 5.67E-01 | 8.95E-01 |
| *GIGYF1* | 2.10E-06 | 3.77E-04 | 3.09E-02 | 3.52E-01 | 6.06E-02 | 5.76E-01 |
| *GOLGA8K* | NA | NA | NA | NA | NA | NA |
| *ZCCHC24* | 7.11E-02 | 5.51E-01 | 3.97E-02 | 3.95E-01 | 2.68E-03 | 1.02E-01 |
| *TFAP2E* | 3.91E-02 | 4.46E-01 | 4.84E-03 | 1.16E-01 | 7.54E-02 | 6.18E-01 |
| *CYP3A43* | 4.77E-01 | 8.59E-01 | 3.27E-01 | 8.19E-01 | 1.11E-01 | 6.97E-01 |
| *TCF7L2* | 3.13E-01 | 8.10E-01 | 2.84E-02 | 3.40E-01 | 5.28E-01 | 8.95E-01 |
| *SUPT6H* | 3.78E-01 | 8.38E-01 | 5.46E-01 | 8.76E-01 | 7.70E-01 | 8.99E-01 |
| *SYMPK* | 4.97E-01 | 8.63E-01 | 1.32E-03 | 4.66E-02 | 1.27E-02 | 2.73E-01 |
| *DDAH2* | 3.64E-01 | 8.33E-01 | 5.91E-02 | 4.73E-01 | 3.47E-03 | 1.22E-01 |
| *AATK-AS1* | NA | NA | NA | NA | NA | NA |
| *CREBRF* | 1.94E-01 | 7.31E-01 | 2.83E-03 | 7.99E-02 | 6.47E-01 | 8.95E-01 |
| *C1orf56* | 2.22E-01 | 7.56E-01 | 2.19E-01 | 7.48E-01 | 1.41E-01 | 7.44E-01 |
| *MSTO1* | 2.15E-06 | 3.83E-04 | 3.12E-07 | 4.94E-05 | 7.31E-07 | 1.17E-04 |
| *ANXA11* | 2.64E-02 | 3.84E-01 | 1.71E-03 | 5.62E-02 | 5.36E-02 | 5.52E-01 |
| *TNFSF8* | 2.02E-03 | 9.01E-02 | 1.34E-02 | 2.22E-01 | 2.16E-02 | 3.75E-01 |
| *GAL3ST4* | 1.08E-03 | 6.04E-02 | 2.09E-06 | 2.64E-04 | 8.41E-05 | 6.79E-03 |
| *COL11A2* | 1.20E-03 | 6.49E-02 | 1.74E-02 | 2.58E-01 | 4.37E-02 | 5.08E-01 |
| *SPOP* | 5.00E-02 | 4.91E-01 | 1.57E-04 | 9.16E-03 | 1.58E-01 | 7.70E-01 |
| *PLSCR2* | 2.39E-01 | 7.70E-01 | 3.83E-03 | 9.87E-02 | 2.62E-05 | 2.67E-03 |
| *KLF1* | 3.57E-04 | 2.72E-02 | 1.54E-01 | 6.66E-01 | 5.98E-01 | 8.95E-01 |
| *ARL17B* | 7.27E-02 | 5.55E-01 | 2.18E-08 | 4.06E-06 | 2.17E-06 | 3.02E-04 |
| *C6orf48* | 7.71E-02 | 5.68E-01 | 4.05E-03 | 1.02E-01 | 1.94E-03 | 8.31E-02 |
| *RPP21* | 6.28E-01 | 8.79E-01 | 4.40E-04 | 2.08E-02 | 2.02E-03 | 8.60E-02 |
| *RP11-57G10.8* | NA | NA | NA | NA | NA | NA |
| *YLPM1* | 2.03E-01 | 7.43E-01 | 1.48E-02 | 2.35E-01 | 4.78E-05 | 4.35E-03 |
| *GATAD2A* | 3.33E-01 | 8.19E-01 | 3.95E-01 | 8.44E-01 | 9.65E-01 | 9.68E-01 |
| *OAS2* | 1.06E-07 | 2.56E-05 | 6.72E-11 | 2.33E-08 | 3.18E-07 | 5.58E-05 |
| *HEY1* | 5.47E-02 | 5.08E-01 | 6.58E-01 | 8.88E-01 | 7.05E-01 | 8.96E-01 |
| *YY1AP1* | 7.24E-04 | 4.61E-02 | 1.22E-05 | 1.19E-03 | 2.49E-07 | 4.47E-05 |
| *FCHSD2* | 6.48E-01 | 8.79E-01 | 5.49E-02 | 4.57E-01 | 9.72E-03 | 2.35E-01 |
| *MAPT* | 2.26E-04 | 1.95E-02 | 5.83E-12 | 2.62E-09 | 2.19E-11 | 1.05E-08 |
| *GABARAPL2* | 2.46E-02 | 3.74E-01 | 6.27E-05 | 4.58E-03 | 1.06E-02 | 2.46E-01 |
| *CYP4B1* | 1.80E-02 | 3.25E-01 | 7.27E-06 | 7.72E-04 | 6.76E-02 | 5.99E-01 |
| *RP11-91I20.3* | NA | NA | NA | NA | NA | NA |
| *THRA* | 1.81E-06 | 3.38E-04 | 4.82E-03 | 1.16E-01 | 7.48E-03 | 2.02E-01 |
| *EDC3* | 2.15E-01 | 7.51E-01 | 2.62E-03 | 7.54E-02 | 6.86E-05 | 5.84E-03 |
| *SPARC* | 6.92E-02 | 5.46E-01 | 3.35E-01 | 8.21E-01 | 1.38E-01 | 7.41E-01 |
| *NPRL3* | 6.13E-01 | 8.78E-01 | 9.46E-02 | 5.71E-01 | 1.44E-01 | 7.48E-01 |
| *EPS8L1* | 6.28E-06 | 9.41E-04 | 3.11E-02 | 3.53E-01 | 5.62E-01 | 8.95E-01 |

| | | | | | | |
|---|---|---|---|---|---|---|
| *CFB* | 1.06E-01 | 6.27E-01 | 5.74E-04 | 2.55E-02 | 3.74E-03 | 1.27E-01 |
| *AC114752.1* | NA | NA | NA | NA | NA | NA |
| *CCDC18* | 3.67E-03 | 1.37E-01 | 4.70E-03 | 1.15E-01 | 2.48E-06 | 3.38E-04 |
| *LINC01315* | NA | NA | NA | NA | NA | NA |
| *NCOR1* | 2.22E-02 | 3.57E-01 | 1.63E-05 | 1.49E-03 | 4.10E-05 | 3.88E-03 |
| *SNX1* | 4.22E-01 | 8.51E-01 | 4.46E-05 | 3.44E-03 | 1.06E-02 | 2.46E-01 |
| *DNAH8* | 3.71E-01 | 8.36E-01 | 4.94E-04 | 2.29E-02 | 3.73E-01 | 8.80E-01 |
| *RAVER2* | 7.09E-01 | 8.84E-01 | 2.50E-02 | 3.17E-01 | 2.36E-03 | 9.50E-02 |
| *HIST1H2AB* | 2.00E-04 | 1.78E-02 | 1.39E-02 | 2.27E-01 | 9.24E-01 | 9.42E-01 |
| *SQRDL* | 2.13E-01 | 7.49E-01 | 1.98E-03 | 6.28E-02 | 7.03E-02 | 6.10E-01 |
| *FNIP1* | 1.76E-03 | 8.22E-02 | 1.03E-04 | 6.66E-03 | 1.12E-03 | 5.47E-02 |
| *TCF12* | 1.95E-05 | 2.49E-03 | 7.15E-01 | 8.93E-01 | 4.99E-01 | 8.95E-01 |
| *EPO* | 1.64E-04 | 1.51E-02 | 5.41E-02 | 4.55E-01 | 1.87E-01 | 7.98E-01 |
| *CTC-296K1.3* | NA | NA | NA | NA | NA | NA |
| *LAMTOR4* | 2.34E-03 | 9.93E-02 | 8.17E-06 | 8.41E-04 | 2.96E-05 | 2.93E-03 |
| *CTB-50L17.16* | NA | NA | NA | NA | NA | NA |
| *DONSON* | 1.89E-01 | 7.27E-01 | 7.54E-06 | 7.88E-04 | 1.53E-02 | 3.06E-01 |
| *ZPBP2* | 1.68E-03 | 7.96E-02 | 3.09E-04 | 1.56E-02 | 5.80E-03 | 1.73E-01 |
| *NFAM1* | 3.58E-07 | 7.61E-05 | 9.12E-03 | 1.77E-01 | 1.83E-03 | 8.01E-02 |
| *NCDN* | 1.86E-02 | 3.32E-01 | 5.45E-05 | 4.08E-03 | 5.59E-03 | 1.68E-01 |
| *ORMDL3* | 2.28E-07 | 5.12E-05 | 1.22E-07 | 2.08E-05 | 3.85E-05 | 3.69E-03 |
| *ZNF655* | 1.79E-01 | 7.17E-01 | 9.98E-02 | 5.82E-01 | 4.84E-01 | 8.95E-01 |
| *IBA57* | 3.22E-02 | 4.16E-01 | 3.75E-07 | 5.78E-05 | 6.84E-04 | 3.84E-02 |
| *CALR* | 1.27E-01 | 6.50E-01 | 6.42E-02 | 4.89E-01 | 3.92E-01 | 8.85E-01 |
| *CPOX* | 3.07E-02 | 4.09E-01 | 2.73E-01 | 7.92E-01 | 4.63E-02 | 5.20E-01 |
| *FAT2* | 1.36E-01 | 6.61E-01 | 1.05E-01 | 5.90E-01 | 6.81E-01 | 8.96E-01 |
| *ST3GAL3* | 9.24E-01 | 9.41E-01 | 2.16E-02 | 2.91E-01 | 8.83E-03 | 2.23E-01 |
| *DNAJB14* | 7.46E-01 | 8.90E-01 | 6.16E-02 | 4.81E-01 | 1.17E-01 | 7.08E-01 |
| *LINC01565* | NA | NA | NA | NA | NA | NA |
| *SNRPD2* | 3.48E-02 | 4.27E-01 | 7.60E-06 | 7.90E-04 | 2.55E-03 | 9.95E-02 |
| *GNB2* | 2.46E-07 | 5.46E-05 | 2.45E-02 | 3.13E-01 | 1.83E-02 | 3.42E-01 |
| *TSACC* | 5.98E-03 | 1.85E-01 | 2.08E-03 | 6.47E-02 | 3.74E-03 | 1.27E-01 |
| *DCST1* | 4.27E-06 | 6.92E-04 | 4.91E-11 | 1.76E-08 | 3.05E-10 | 9.72E-08 |
| *6-Mar* | 5.08E-01 | 8.63E-01 | 4.78E-01 | 8.67E-01 | 2.65E-01 | 8.48E-01 |
| *TRAF3* | 1.85E-06 | 3.42E-04 | 4.74E-02 | 4.30E-01 | 8.87E-03 | 2.23E-01 |
| *ZMYM4* | 1.30E-01 | 6.53E-01 | 2.06E-03 | 6.45E-02 | 7.54E-03 | 2.03E-01 |
| *AQP4* | 1.89E-01 | 7.27E-01 | 2.52E-01 | 7.78E-01 | 2.86E-01 | 8.58E-01 |
| *ARID3B* | 3.09E-01 | 8.07E-01 | 1.01E-02 | 1.88E-01 | 1.07E-05 | 1.20E-03 |
| *AP1M2* | 3.72E-01 | 8.36E-01 | 8.62E-02 | 5.52E-01 | 6.02E-02 | 5.75E-01 |
| *FAM189B* | 2.01E-02 | 3.45E-01 | 9.96E-03 | 1.86E-01 | 1.44E-04 | 1.07E-02 |
| *ADAMTS8* | 7.77E-01 | 8.94E-01 | 7.61E-01 | 8.98E-01 | 9.26E-01 | 9.43E-01 |
| *MICA* | 8.58E-02 | 5.86E-01 | 7.32E-06 | 7.73E-04 | 4.43E-05 | 4.11E-03 |
| *PRSS33* | 8.54E-02 | 5.86E-01 | 5.50E-01 | 8.76E-01 | 2.61E-01 | 8.46E-01 |
| *DAP3* | 1.24E-05 | 1.70E-03 | 2.91E-04 | 1.49E-02 | 8.05E-07 | 1.26E-04 |
| *LINC01149* | NA | NA | NA | NA | NA | NA |

| | | | | | | |
|---|---|---|---|---|---|---|
| *TAF6* | 3.23E-02 | 4.17E-01 | 4.64E-04 | 2.18E-02 | 1.09E-04 | 8.42E-03 |
| *GPANK1* | 6.27E-01 | 8.79E-01 | 4.74E-03 | 1.15E-01 | 1.73E-04 | 1.26E-02 |
| *LINC00310* | NA | NA | NA | NA | NA | NA |
| *RP11-503G7.2* | NA | NA | NA | NA | NA | NA |
| *FRYL* | 4.86E-02 | 4.86E-01 | 4.98E-04 | 2.30E-02 | 5.77E-01 | 8.95E-01 |
| *E2F1* | 3.85E-02 | 4.43E-01 | 1.45E-05 | 1.34E-03 | 1.38E-03 | 6.50E-02 |
| *PRRT1* | 2.61E-01 | 7.87E-01 | 4.30E-05 | 3.40E-03 | 7.33E-04 | 4.00E-02 |
| *IRF7* | 1.96E-02 | 3.40E-01 | 4.29E-01 | 8.55E-01 | 8.88E-03 | 2.23E-01 |
| *LINC00857* | NA | NA | NA | NA | NA | NA |
| *CERCAM* | 4.90E-03 | 1.62E-01 | 6.03E-02 | 4.77E-01 | 3.65E-01 | 8.77E-01 |
| *ZSWIM7* | 7.55E-02 | 5.64E-01 | 6.14E-05 | 4.51E-03 | 6.91E-04 | 3.86E-02 |
| *ADRA1A* | 1.93E-01 | 7.30E-01 | 2.05E-01 | 7.35E-01 | 5.36E-03 | 1.63E-01 |
| *NPY4R* | NA | NA | NA | NA | 3.19E-01 | 8.66E-01 |
| *TRMT61B* | 8.03E-02 | 5.73E-01 | 5.18E-04 | 2.36E-02 | 2.55E-01 | 8.43E-01 |
| *GJC2* | 1.42E-01 | 6.69E-01 | 9.35E-05 | 6.28E-03 | 8.93E-04 | 4.60E-02 |
| *PSMB2* | 8.61E-02 | 5.86E-01 | 2.61E-02 | 3.24E-01 | 2.28E-01 | 8.29E-01 |
| *PTGFR* | 5.96E-01 | 8.76E-01 | 7.00E-02 | 5.10E-01 | 7.14E-01 | 8.96E-01 |
| *VPS29* | 1.02E-01 | 6.22E-01 | 5.42E-04 | 2.46E-02 | 3.67E-01 | 8.78E-01 |
| *LINC00348* | NA | NA | NA | NA | NA | NA |
| *BRD2* | 6.66E-02 | 5.38E-01 | 1.03E-03 | 3.85E-02 | 2.37E-04 | 1.63E-02 |
| *UPF2* | 5.99E-02 | 5.23E-01 | 8.15E-01 | 9.04E-01 | 7.06E-01 | 8.96E-01 |
| *CHCHD7* | 6.42E-01 | 8.79E-01 | 3.45E-01 | 8.28E-01 | 2.31E-01 | 8.30E-01 |
| *HIST1H2BB* | 2.70E-04 | 2.21E-02 | 2.48E-02 | 3.15E-01 | 9.61E-01 | 9.65E-01 |
| *HSPE1* | 3.07E-01 | 8.06E-01 | 6.40E-05 | 4.63E-03 | 5.36E-04 | 3.15E-02 |
| *RP11-499E18.1* | NA | NA | NA | NA | NA | NA |
| *GCDH* | 7.77E-03 | 2.14E-01 | 5.99E-02 | 4.76E-01 | 4.58E-01 | 8.94E-01 |
| *ZNF146* | 4.40E-01 | 8.57E-01 | 3.90E-04 | 1.88E-02 | 1.83E-02 | 3.42E-01 |
| *CYP2C19* | 5.89E-01 | 8.75E-01 | 4.85E-01 | 8.69E-01 | 6.32E-02 | 5.84E-01 |
| *BNIP1* | 9.47E-02 | 6.05E-01 | 7.57E-04 | 3.16E-02 | 7.15E-01 | 8.97E-01 |
| *DR1* | 6.37E-04 | 4.21E-02 | 3.23E-03 | 8.79E-02 | 2.07E-05 | 2.20E-03 |
| *SUCLG2* | 2.68E-01 | 7.89E-01 | 2.35E-01 | 7.63E-01 | 2.77E-01 | 8.52E-01 |
| *TADA1* | 4.76E-01 | 8.59E-01 | 1.00E-01 | 5.82E-01 | 1.17E-01 | 7.08E-01 |
| *PRM3* | 7.52E-02 | 5.63E-01 | 2.09E-03 | 6.48E-02 | 2.59E-02 | 4.03E-01 |
| *PREX1* | 1.53E-06 | 2.94E-04 | 5.79E-03 | 1.30E-01 | 5.92E-01 | 8.95E-01 |
| *FAM96A* | 4.54E-01 | 8.58E-01 | 9.60E-05 | 6.31E-03 | 5.69E-02 | 5.65E-01 |
| *WFDC10A* | 1.14E-03 | 6.25E-02 | 2.90E-01 | 8.02E-01 | 7.55E-01 | 8.99E-01 |
| *AIP* | 3.62E-02 | 4.34E-01 | 8.15E-02 | 5.42E-01 | 4.53E-02 | 5.17E-01 |
| *PNRC1* | 7.10E-01 | 8.84E-01 | 4.91E-01 | 8.72E-01 | 1.61E-02 | 3.16E-01 |
| *XRCC3* | 3.65E-03 | 1.37E-01 | 2.92E-05 | 2.43E-03 | 1.87E-03 | 8.16E-02 |
| *LPCAT2* | 2.67E-01 | 7.89E-01 | 2.85E-01 | 8.00E-01 | 8.91E-02 | 6.55E-01 |
| *ATG16L2* | 1.27E-01 | 6.50E-01 | 2.60E-03 | 7.54E-02 | 7.13E-05 | 6.05E-03 |
| *ACSM4* | 4.30E-01 | 8.53E-01 | 1.17E-02 | 2.06E-01 | 3.12E-01 | 8.65E-01 |
| *ZNF311* | 6.34E-02 | 5.30E-01 | 5.01E-02 | 4.38E-01 | 2.26E-01 | 8.28E-01 |
| *HNMT* | 7.56E-02 | 5.65E-01 | 4.24E-01 | 8.54E-01 | 1.61E-01 | 7.73E-01 |
| *PDLIM4* | 2.94E-02 | 4.02E-01 | 1.04E-04 | 6.72E-03 | 2.28E-03 | 9.26E-02 |

| | | | | | | |
|---|---|---|---|---|---|---|
| *ILF3-AS1* | NA | NA | NA | NA | NA | NA |
| *PPP1R2* | 1.08E-01 | 6.31E-01 | 4.27E-01 | 8.55E-01 | 1.73E-01 | 7.89E-01 |
| *REEP4* | 1.85E-01 | 7.22E-01 | 8.38E-03 | 1.67E-01 | 6.57E-01 | 8.96E-01 |
| *RPN1* | 6.30E-01 | 8.79E-01 | 1.26E-01 | 6.27E-01 | 1.46E-01 | 7.51E-01 |
| *LPIN3* | 9.06E-03 | 2.34E-01 | 8.05E-02 | 5.40E-01 | 4.03E-03 | 1.32E-01 |
| *ARHGEF38* | 1.54E-01 | 6.86E-01 | 1.44E-06 | 1.95E-04 | 1.49E-07 | 2.81E-05 |
| *AP4M1* | 1.25E-02 | 2.74E-01 | 1.40E-04 | 8.47E-03 | 6.60E-05 | 5.71E-03 |
| *ZSCAN23* | 1.59E-02 | 3.11E-01 | 3.68E-03 | 9.66E-02 | 2.20E-02 | 3.77E-01 |
| *ZNF84* | NA | NA | NA | NA | NA | NA |
| *LMAN1L* | 2.12E-02 | 3.50E-01 | 1.17E-04 | 7.33E-03 | 9.87E-02 | 6.75E-01 |
| *PROB1* | 8.66E-04 | 5.19E-02 | 1.80E-01 | 7.04E-01 | 3.99E-03 | 1.32E-01 |
| *RP11-16E12.2* | NA | NA | NA | NA | NA | NA |
| *BMP8A* | 8.91E-03 | 2.33E-01 | 1.92E-04 | 1.07E-02 | 5.35E-04 | 3.15E-02 |
| *GSDMB* | 1.92E-06 | 3.48E-04 | 4.81E-07 | 7.14E-05 | 8.21E-05 | 6.66E-03 |
| *UNC5C* | 6.69E-01 | 8.81E-01 | 8.16E-01 | 9.04E-01 | 8.38E-01 | 9.09E-01 |
| *RASSF3* | 4.85E-01 | 8.61E-01 | 8.45E-03 | 1.67E-01 | 1.13E-01 | 7.01E-01 |
| *IMPG1* | 4.21E-01 | 8.50E-01 | 6.78E-01 | 8.88E-01 | 7.18E-01 | 8.97E-01 |
| *NDUFC1* | 8.79E-01 | 9.21E-01 | 3.28E-03 | 8.89E-02 | 1.76E-01 | 7.92E-01 |
| *DKFZP434L187* | NA | NA | NA | NA | NA | NA |
| *FAM126A* | 9.12E-03 | 2.34E-01 | 1.35E-02 | 2.23E-01 | 2.74E-02 | 4.17E-01 |
| *CPLX3* | 2.14E-02 | 3.52E-01 | 3.17E-04 | 1.59E-02 | 1.28E-01 | 7.26E-01 |
| *IKZF3* | 7.13E-02 | 5.52E-01 | 3.05E-02 | 3.50E-01 | 8.42E-02 | 6.43E-01 |
| *KIAA0907* | 2.08E-01 | 7.45E-01 | 1.09E-01 | 5.99E-01 | 6.86E-05 | 5.84E-03 |
| *ZKSCAN4* | 5.41E-04 | 3.76E-02 | 5.63E-04 | 2.51E-02 | 1.20E-02 | 2.65E-01 |
| *LKAAEAR1* | 3.26E-01 | 8.17E-01 | 2.60E-03 | 7.54E-02 | 5.10E-01 | 8.95E-01 |
| *DYNC1H1* | 7.36E-04 | 4.65E-02 | 3.28E-02 | 3.61E-01 | 1.49E-01 | 7.55E-01 |
| *GNGT2* | 6.45E-01 | 8.79E-01 | 1.14E-01 | 6.09E-01 | 3.74E-01 | 8.80E-01 |
| *CDC42EP3* | 5.84E-01 | 8.74E-01 | 4.61E-01 | 8.64E-01 | 5.92E-01 | 8.95E-01 |
| *NAA20* | 1.67E-01 | 7.03E-01 | 7.20E-02 | 5.16E-01 | 5.52E-01 | 8.95E-01 |
| *MS4A4A* | 5.24E-01 | 8.67E-01 | 6.26E-01 | 8.84E-01 | 4.16E-01 | 8.89E-01 |
| *OBSCN* | 2.72E-02 | 3.87E-01 | 1.33E-05 | 1.27E-03 | 6.37E-03 | 1.82E-01 |
| *C1orf145* | NA | NA | NA | NA | NA | NA |
| *CDH16* | 3.44E-01 | 8.25E-01 | 1.24E-01 | 6.22E-01 | 1.26E-03 | 6.02E-02 |
| *USP49* | 1.36E-02 | 2.87E-01 | 9.58E-05 | 6.31E-03 | 6.20E-02 | 5.82E-01 |
| *SNX19* | 1.86E-04 | 1.68E-02 | 1.33E-04 | 8.14E-03 | 8.77E-03 | 2.23E-01 |
| *FBXL18* | 9.07E-01 | 9.34E-01 | 3.28E-04 | 1.63E-02 | 1.80E-01 | 7.94E-01 |
| *VWA7* | 2.84E-01 | 7.95E-01 | 2.88E-02 | 3.42E-01 | 7.36E-03 | 2.01E-01 |
| *NAA15* | 8.58E-01 | 9.14E-01 | 2.45E-03 | 7.23E-02 | 1.43E-01 | 7.46E-01 |
| *DHX30* | 1.53E-02 | 3.05E-01 | 1.37E-04 | 8.33E-03 | 2.81E-01 | 8.56E-01 |
| *PCMTD1* | 5.07E-01 | 8.63E-01 | 8.78E-04 | 3.50E-02 | 6.73E-04 | 3.82E-02 |
| *HLA-DOA* | 7.68E-03 | 2.13E-01 | 6.61E-04 | 2.86E-02 | 2.56E-04 | 1.73E-02 |
| *RFTN2* | 4.43E-01 | 8.57E-01 | 1.89E-05 | 1.67E-03 | 2.34E-05 | 2.43E-03 |
| *ALKBH1* | 5.49E-01 | 8.69E-01 | 8.86E-03 | 1.73E-01 | 1.64E-01 | 7.77E-01 |
| *LDLR* | 7.73E-01 | 8.94E-01 | 1.55E-01 | 6.68E-01 | 7.17E-02 | 6.14E-01 |
| *DSCAML1* | 3.76E-01 | 8.37E-01 | 5.52E-02 | 4.58E-01 | 3.57E-01 | 8.77E-01 |

| | | | | | | |
|---|---|---|---|---|---|---|
| *LCTL* | 6.90E-01 | 8.82E-01 | 1.64E-01 | 6.80E-01 | 1.05E-02 | 2.46E-01 |
| *HINT1* | 7.14E-01 | 8.84E-01 | 4.08E-02 | 4.01E-01 | 2.17E-04 | 1.50E-02 |
| *MAP2* | 7.17E-02 | 5.53E-01 | 5.75E-03 | 1.30E-01 | 4.84E-02 | 5.28E-01 |
| *STAG3* | 3.27E-02 | 4.17E-01 | 9.97E-04 | 3.78E-02 | 1.61E-02 | 3.16E-01 |
| *IRF4* | 5.48E-02 | 5.08E-01 | 2.69E-01 | 7.88E-01 | 3.97E-01 | 8.86E-01 |
| *PKP2* | 1.86E-01 | 7.25E-01 | 4.58E-01 | 8.63E-01 | 8.73E-01 | 9.19E-01 |
| *EPB41L2* | 3.86E-02 | 4.44E-01 | 8.54E-04 | 3.44E-02 | 1.37E-02 | 2.85E-01 |
| *MSH5* | 3.53E-01 | 8.28E-01 | 2.86E-02 | 3.41E-01 | 2.58E-03 | 1.00E-01 |

Note: NA represents not applicable.

**Supplementary Table S11. The 23 common pathways significantly enriched by three S-MultiXcan gene sets**

| **Pathway ID** | **Pathway name** | **Susceptible COVID-19 (P-value)** | **Susceptible COVID-19 (FDR)** | **Hospitalized COVID-19 (P-value)** | **Hospitalized COVID-19 (FDR)** | **Very severe COVID-19 (P-value)** | **Very severe COVID-19 (FDR)** |
|---|---|---|---|---|---|---|---|
| hsa05169 | Epstein-Barr virus infection | 3.48E-04 | 2.83E-02 | 8.00E-11 | 2.61E-08 | 1.45E-09 | 4.71E-07 |
| hsa05168 | Herpes simplex infection | 1.12E-03 | 7.33E-02 | 1.37E-09 | 1.49E-07 | 3.35E-09 | 5.46E-07 |
| hsa05164 | Influenza A | 1.69E-02 | 2.76E-01 | 1.90E-07 | 1.24E-05 | 8.98E-09 | 9.76E-07 |
| hsa04940 | Type I diabetes mellitus | 6.25E-03 | 2.04E-01 | 1.72E-07 | 1.24E-05 | 9.41E-08 | 7.67E-06 |
| hsa05167 | Kaposi sarcoma-associated herpesvirus infection | 3.16E-05 | 3.90E-03 | 1.81E-10 | 2.94E-08 | 2.03E-07 | 1.32E-05 |
| hsa05320 | Autoimmune thyroid disease | 1.11E-02 | 2.58E-01 | 9.29E-07 | 5.05E-05 | 5.14E-07 | 2.79E-05 |
| hsa05330 | Allograft rejection | 4.41E-03 | 1.80E-01 | 1.13E-06 | 5.28E-05 | 6.70E-07 | 3.12E-05 |
| hsa05332 | Graft-versus-host disease | 5.46E-03 | 1.98E-01 | 1.95E-06 | 7.82E-05 | 1.15E-06 | 4.35E-05 |
| hsa05416 | Viral myocarditis | 1.49E-02 | 2.76E-01 | 2.16E-06 | 7.82E-05 | 1.20E-06 | 4.35E-05 |
| hsa04060 | Cytokine-cytokine receptor interaction | 7.59E-06 | 2.48E-03 | 1.04E-04 | 2.11E-03 | 2.11E-06 | 6.89E-05 |
| hsa05145 | Toxoplasmosis | 1.67E-02 | 2.76E-01 | 1.32E-03 | 2.14E-02 | 2.09E-05 | 6.18E-04 |
| hsa04514 | Cell adhesion molecules (CAMs) | 3.75E-02 | 4.71E-01 | 5.02E-05 | 1.09E-03 | 2.57E-05 | 6.97E-04 |
| hsa04145 | Phagosome | 2.09E-03 | 9.72E-02 | 1.40E-05 | 3.81E-04 | 4.09E-05 | 9.72E-04 |
| hsa05160 | Hepatitis C | 2.78E-02 | 4.09E-01 | 6.97E-04 | 1.33E-02 | 7.24E-05 | 1.57E-03 |
| hsa04612 | Antigen processing and presentation | 3.00E-02 | 4.09E-01 | 1.64E-05 | 4.12E-04 | 8.25E-05 | 1.68E-03 |
| hsa04672 | Intestinal immune network for IgA production | 8.98E-03 | 2.44E-01 | 5.84E-03 | 7.61E-02 | 5.25E-04 | 8.56E-03 |
| hsa05310 | Asthma | 3.01E-02 | 4.09E-01 | 1.06E-02 | 1.12E-01 | 7.97E-04 | 1.24E-02 |
| hsa05163 | Human cytomegalovirus infection | 1.38E-02 | 2.76E-01 | 5.21E-06 | 1.55E-04 | 9.97E-04 | 1.48E-02 |
| hsa04621 | NOD-like receptor signaling pathway | 1.58E-02 | 2.76E-01 | 8.35E-04 | 1.43E-02 | 2.10E-03 | 2.74E-02 |
| hsa00601 | Glycosphingolipid biosynthesis | 1.64E-03 | 8.89E-02 | 7.19E-03 | 8.69E-02 | 5.81E-03 | 6.53E-02 |
| hsa05203 | Viral carcinogenesis | 3.14E-02 | 4.09E-01 | 9.85E-03 | 1.12E-01 | 6.28E-03 | 6.82E-02 |
| hsa04062 | Chemokine signaling pathway | 3.59E-05 | 3.90E-03 | 6.90E-03 | 8.65E-02 | 1.50E-02 | 1.40E-01 |
| hsa04151 | PI3K-Akt signaling pathway | 1.19E-02 | 2.58E-01 | 1.56E-02 | 1.50E-01 | 2.32E-02 | 2.04E-01 |

**Supplementary Table S12. LDSC analysis identifies the genetic correlations between three COVID-19 phenotypes and 66 complex diseases or traits**

| Phenotypes | Susceptible COVID-19 | | Hospitalized COVID-19 | | Very severe COVID-19 | |
|---|---|---|---|---|---|---|
| | $r_g$ | P value | $r_g$ | P value | $r_g$ | P value |
| Anorexia nervosa | 0.010 | 8.97E-01 | -0.129 | 4.29E-02 | -0.131 | 4.05E-02 |
| Autism spectrum disorder | -0.063 | 2.32E-01 | -0.062 | 1.81E-01 | -0.062 | 1.94E-01 |
| Bipolar disorder | -0.063 | 1.40E-01 | -0.039 | 2.27E-01 | -0.060 | 9.97E-02 |
| Attention deficit hyperactivity disorder | 0.224 | 3.12E-07 | 0.248 | 3.12E-08 | 0.181 | 1.11E-05 |
| Major Depression Disease | 0.088 | 1.07E-01 | 0.097 | 5.05E-02 | 0.070 | 1.46E-01 |
| Schizophrenia | 0.043 | 1.32E-01 | -0.012 | 6.17E-01 | 0.004 | 8.86E-01 |
| Insomnia | -0.185 | 5.93E-06 | -0.181 | 7.12E-05 | -0.102 | 2.75E-02 |
| Multiple sclerosis | 0.074 | 1.35E-01 | 0.136 | 8.84E-03 | 0.106 | 5.73E-02 |
| Migraine | -0.049 | 3.74E-01 | 0.019 | 7.03E-01 | 0.016 | 7.56E-01 |
| Ischemic stroke | 0.082 | 1.82E-01 | 0.206 | 9.73E-05 | 0.189 | 1.23E-03 |
| Epilepsy | -0.059 | 5.01E-01 | -0.046 | 5.24E-01 | -0.108 | 1.39E-01 |
| Focal epilepsy | -0.092 | 5.56E-01 | -0.044 | 7.29E-01 | -0.104 | 4.14E-01 |
| Generalized epilepsy | -0.040 | 5.75E-01 | -0.054 | 3.48E-01 | -0.120 | 4.01E-02 |
| Juvenile myoclonic epilepsy | -0.079 | 3.32E-01 | -0.081 | 2.56E-01 | -0.123 | 9.90E-02 |
| Alzheimer’s disease | 0.219 | 4.90E-02 | 0.067 | 3.96E-01 | 0.052 | 5.47E-01 |
| Parkinson’s disease | -0.022 | 7.06E-01 | -0.031 | 5.97E-01 | -0.032 | 5.94E-01 |
| Snoring | 0.110 | 1.85E-03 | 0.131 | 5.87E-05 | 0.140 | 6.08E-05 |
| Daytime Napping | 0.132 | 1.47E-02 | 0.158 | 5.38E-03 | 0.153 | 1.53E-02 |
| Happiness | -0.094 | 8.72E-02 | -0.005 | 9.09E-01 | 0.006 | 8.98E-01 |
| Cigarettes smoked per day | 0.131 | 1.05E-03 | 0.200 | 7.56E-09 | 0.177 | 2.31E-06 |
| Extraversion | -0.094 | 8.72E-02 | -0.005 | 9.09E-01 | 0.006 | 8.98E-01 |
| Subjective well-being | -0.014 | 7.84E-01 | 0.018 | 7.19E-01 | 0.020 | 7.37E-01 |
| Conscientiousness | 0.116 | 3.46E-01 | 0.040 | 7.03E-01 | -0.003 | 9.81E-01 |
| Openness to experience | -0.162 | 1.27E-01 | -0.113 | 2.23E-01 | -0.045 | 6.20E-01 |
| Neuroticism | 0.022 | 5.06E-01 | 0.078 | 1.09E-02 | 0.084 | 1.70E-02 |
| Cognitive Performance | -0.298 | 4.96E-18 | -0.216 | 1.23E-11 | -0.138 | 3.47E-05 |
| Smoking cesstion | -0.104 | 2.07E-01 | -0.016 | 8.25E-01 | -0.050 | 5.20E-01 |
| Age of smoking initiation | 0.074 | 1.35E-01 | 0.136 | 8.84E-03 | 0.106 | 5.73E-02 |
| Hypertension | 0.153 | 7.86E-06 | 0.187 | 4.74E-09 | 0.155 | 1.12E-05 |
| Systolic blood pressure | -0.049 | 9.17E-02 | 0.005 | 8.45E-01 | -0.006 | 8.02E-01 |
| Pulse pressure | 0.154 | 1.33E-02 | 0.084 | 1.39E-01 | 0.086 | 1.33E-01 |
| High blood pressure | 0.148 | 1.19E-05 | 0.186 | 1.56E-08 | 0.154 | 1.86E-05 |

| | | | | | | |
|---|---|---|---|---|---|---|
| Diastolic blood pressure (East Asian) | 0.091 | 1.40E-01 | 0.037 | 4.85E-01 | 0.087 | 1.34E-01 |
| Diastolic blood pressure (European) | -0.038 | 2.22E-01 | 0.007 | 7.66E-01 | -0.006 | 8.21E-01 |
| Diastolic blood pressure (Hispanic) | 0.047 | 5.86E-01 | 0.048 | 4.91E-01 | 0.031 | 6.61E-01 |
| Coronary artery disease | 0.152 | 2.83E-04 | 0.205 | 9.12E-07 | 0.125 | 4.21E-03 |
| Type II diabetes | 0.174 | 6.20E-06 | 0.282 | 3.24E-09 | 0.241 | 3.93E-06 |
| Ulcerative Colitis | -0.072 | 1.72E-01 | 0.004 | 9.31E-01 | -0.013 | 7.95E-01 |
| Psoriasis | 0.091 | 3.31E-01 | 0.045 | 5.50E-01 | -0.047 | 5.54E-01 |
| Coeliac disease | 0.007 | 9.43E-01 | 0.015 | 8.70E-01 | -0.054 | 6.20E-01 |
| Primary biliary cholangitis | 0.024 | 7.44E-01 | 0.131 | 7.82E-02 | 0.121 | 1.34E-01 |
| Rheumatoid arthritis | 0.144 | 2.68E-03 | 0.134 | 1.49E-03 | 0.088 | 6.42E-02 |
| Inflammatory bowel disease | -0.071 | 1.14E-01 | -0.002 | 9.59E-01 | -0.035 | 3.55E-01 |
| Systemic lupus erythematosus | -0.020 | 7.82E-01 | 0.079 | 2.14E-01 | 0.094 | 1.44E-01 |
| Waist Circumference | 0.260 | 5.73E-10 | 0.330 | 7.64E-10 | 0.324 | 5.56E-07 |
| Total cholesterol | -0.106 | 3.94E-02 | -0.107 | 1.58E-02 | -0.083 | 8.94E-02 |
| Chronic kidney disease | 0.096 | 4.03E-01 | -0.004 | 9.64E-01 | -0.022 | 8.21E-01 |
| Body fat percentage | 0.281 | 1.49E-18 | 0.362 | 1.31E-15 | 0.338 | 2.77E-09 |
| Triglyceride level | 0.014 | 8.74E-01 | 0.164 | 3.27E-02 | 0.142 | 9.41E-02 |
| High-density lipoprotein (HDL) cholesterol | -0.159 | 2.42E-06 | -0.196 | 8.91E-09 | -0.178 | 1.77E-05 |
| Low-density lipoprotein (LDL) cholesterol | -0.039 | 2.99E-01 | -0.019 | 5.96E-01 | -0.021 | 5.79E-01 |
| Tuberculosis | 0.429 | 3.50E-01 | 0.013 | 9.57E-01 | 0.267 | 4.13E-01 |
| Adult asthma | 0.080 | 8.54E-02 | -0.015 | 7.05E-01 | -0.041 | 3.20E-01 |
| Small cell lung cancer | 0.024 | 9.12E-01 | 0.119 | 4.99E-01 | 0.051 | 7.69E-01 |
| Pulmonary embolism | 0.248 | 1.70E-02 | 0.222 | 3.16E-02 | 0.221 | 2.51E-02 |
| Pulmonary artery | 0.218 | 1.36E-01 | 0.398 | 3.16E-02 | 0.417 | 7.17E-03 |
| Non-small cell lung cancer, squamous | 0.091 | 6.09E-01 | 0.306 | 3.16E-02 | 0.075 | 6.04E-01 |
| Non-small cell lung cancer, adenocarcinoma | 0.077 | 5.85E-01 | 0.255 | 3.16E-02 | 0.229 | 9.65E-02 |
| Non-small cell lung cancer | 0.066 | 4.80E-01 | 0.191 | 3.16E-02 | 0.174 | 6.32E-02 |
| Lung volume | -0.058 | 6.04E-02 | -0.066 | 6.22E-05 | -0.037 | 2.43E-01 |
| Lung function (FVC) | 0.059 | 5.77E-02 | 0.180 | 6.22E-05 | 0.211 | 1.42E-07 |
| Influenza without pneumonia | 0.220 | 2.27E-01 | 0.173 | 2.06E-01 | 0.079 | 5.43E-01 |
| Influenza with pneumonia | 0.180 | 1.56E-02 | 0.254 | 1.08E-02 | 0.220 | 6.94E-04 |
| All influenza | 0.196 | 1.32E-01 | 0.225 | 1.08E-02 | 0.106 | 2.85E-01 |

| | | | | | | |
|---|---|---|---|---|---|---|
| Emphysema/chronic bronchitis | 0.149 | 5.38E-02 | 0.169 | 1.08E-02 | 0.164 | 1.98E-02 |
| Chronic obstructive pulmonary disease | 0.229 | 1.00E-02 | 0.193 | 1.11E-02 | 0.146 | 8.15E-02 |

### Supplementary Table S13. Significant pathways enriched by up-regulated genes in MSC positive cells

| Description | Gene size | Enrichment ratio | P Value | FDR |
|---|---|---|---|---|
| Parathyroid hormone synthesis, secretion and action | 106 | 2.13 | 3.03E-03 | 2.47E-02 |
| Herpes simplex infection | 185 | 1.83 | 2.73E-03 | 2.28E-02 |
| Influenza A | 171 | 1.90 | 2.07E-03 | 1.79E-02 |
| IL-17 signaling pathway | 93 | 2.28 | 2.09E-03 | 1.79E-02 |
| Relaxin signaling pathway | 130 | 2.06 | 1.90E-03 | 1.75E-02 |
| Viral carcinogenesis | 201 | 1.83 | 1.93E-03 | 1.75E-02 |
| Fluid shear stress and atherosclerosis | 139 | 2.03 | 1.77E-03 | 1.69E-02 |
| Ferroptosis | 40 | 3.18 | 1.57E-03 | 1.56E-02 |
| Melanoma | 72 | 2.55 | 1.43E-03 | 1.46E-02 |
| Transcriptional misregulation in cancer | 186 | 1.90 | 1.36E-03 | 1.43E-02 |
| Th17 cell differentiation | 107 | 2.24 | 1.29E-03 | 1.40E-02 |
| Regulation of actin cytoskeleton | 213 | 1.86 | 1.02E-03 | 1.15E-02 |
| Renal cell carcinoma | 69 | 2.66 | 9.50E-04 | 1.11E-02 |
| Cellular senescence | 160 | 2.03 | 8.36E-04 | 1.01E-02 |
| TNF signaling pathway | 110 | 2.31 | 6.54E-04 | 8.20E-03 |
| Hippo signaling pathway | 154 | 2.11 | 4.84E-04 | 6.31E-03 |
| Prostate cancer | 97 | 2.47 | 4.05E-04 | 5.94E-03 |
| MAPK signaling pathway | 295 | 1.77 | 4.28E-04 | 5.94E-03 |
| Adherens junction | 72 | 2.75 | 4.37E-04 | 5.94E-03 |
| Human T-cell leukemia virus 1 infection | 255 | 1.88 | 2.33E-04 | 3.77E-03 |
| Human cytomegalovirus infection | 225 | 1.95 | 2.43E-04 | 3.77E-03 |
| EGFR tyrosine kinase inhibitor resistance | 79 | 2.86 | 1.05E-04 | 1.97E-03 |
| Amoebiasis | 96 | 2.65 | 1.14E-04 | 1.97E-03 |
| Malaria | 49 | 3.46 | 1.15E-04 | 1.97E-03 |
| JAK-STAT signaling pathway | 162 | 2.27 | 6.27E-05 | 1.32E-03 |
| Hepatitis B | 144 | 2.35 | 6.47E-05 | 1.32E-03 |
| FoxO signaling pathway | 132 | 2.46 | 4.48E-05 | 1.04E-03 |
| Osteoclast differentiation | 128 | 2.54 | 2.70E-05 | 6.76E-04 |
| TGF-beta signaling pathway | 84 | 3.03 | 1.76E-05 | 4.77E-04 |
| Signaling pathways regulating pluripotency of stem cells | 139 | 2.64 | 3.78E-06 | 1.17E-04 |
| Proteoglycans in cancer | 201 | 2.32 | 3.96E-06 | 1.17E-04 |
| ECM-receptor interaction | 82 | 3.27 | 3.03E-06 | 1.10E-04 |
| AGE-RAGE signaling pathway in diabetic complications | 99 | 3.14 | 1.08E-06 | 4.40E-05 |
| Protein digestion and absorption | 90 | 3.29 | 8.08E-07 | 3.76E-05 |
| Kaposi sarcoma-associated herpesvirus infection | 186 | 2.51 | 6.63E-07 | 3.60E-05 |
| PI3K-Akt signaling pathway | 354 | 2.15 | 4.07E-08 | 2.65E-06 |
| Focal adhesion | 199 | 2.77 | 3.48E-09 | 7.87E-07 |

| | | | | |
|---|---|---|---|---|
| Human papillomavirus infection | 339 | 2.25 | 8.77E-09 | 7.87E-07 |
| Pathways in cancer | 526 | 1.96 | 9.26E-09 | 7.87E-07 |
| Protein processing in endoplasmic reticulum | 165 | 2.91 | 9.66E-09 | 7.87E-07 |

**Supplementary Table S14. scDrugHunter-identified top-ranked gene-drug interaction pairs in lung MSC positive cells (scDGS > 120)**

| Gene | Drug | Gene specificity rank | Gene correlation rank | Gene-drug interaction rank | SMultiXcan-based gene P value rank | scDGS |
|---|---|---|---|---|---|---|
| *CCR1* | CCX354 | 9.42 | 9.49 | 9.98 | 9.88 | 187.67 |
| *CCR1* | CHEMBL2205805 | 9.42 | 9.49 | 9.91 | 9.88 | 187.03 |
| *CCR1* | BMS-817399 | 9.42 | 9.49 | 9.91 | 9.88 | 187.03 |
| *CCR1* | AZD4818 | 9.42 | 9.49 | 9.70 | 9.88 | 185.02 |
| *CCR1* | TERPYRIDINE | 9.42 | 9.49 | 9.18 | 9.88 | 180.10 |
| *TNFRSF4* | IVUXOLIMAB | 9.36 | 9.54 | 9.86 | 8.31 | 171.71 |
| *TNFRSF4* | VONLEROLIZUMAB | 9.36 | 9.54 | 9.86 | 8.31 | 171.71 |
| *TNFRSF4* | TAVOLIMAB | 9.36 | 9.54 | 9.86 | 8.31 | 171.71 |
| *PDE4A* | DROTAVERINE | 9.13 | 9.38 | 9.22 | 9.21 | 170.59 |
| *PDE4A* | ENPROFYLLINE | 9.13 | 9.38 | 9.11 | 9.21 | 169.61 |
| *PDE4A* | APREMILAST | 9.13 | 9.38 | 8.92 | 9.21 | 167.86 |
| *PDE4A* | ROFLUMILAST | 9.13 | 9.38 | 8.88 | 9.21 | 167.47 |
| *PDE4A* | DYPHYLLINE | 9.13 | 9.38 | 8.77 | 9.21 | 166.43 |
| *PDE4A* | AROFYLLINE | 9.13 | 9.38 | 8.70 | 9.21 | 165.75 |
| *PDE4A* | OGLEMILAST | 9.13 | 9.38 | 8.70 | 9.21 | 165.75 |
| *PDE4A* | CHEMBL1229585 | 9.13 | 9.38 | 8.70 | 9.21 | 165.75 |
| *PDE4A* | LOTAMILAST | 9.13 | 9.38 | 8.70 | 9.21 | 165.75 |
| *PDE4A* | CRISABOROLE | 9.13 | 9.38 | 8.56 | 9.21 | 164.49 |
| *PDE4A* | PICLAMILAST | 9.13 | 9.38 | 8.53 | 9.21 | 164.19 |
| *PDE4A* | OXTRIPHYLLINE | 9.13 | 9.38 | 8.49 | 9.21 | 163.89 |
| *PDE4A* | REVAMILAST | 9.13 | 9.38 | 8.43 | 9.21 | 163.28 |
| *PDE4A* | MK-0873 | 9.13 | 9.38 | 8.43 | 9.21 | 163.28 |
| *PDE4A* | LIRIMILAST | 9.13 | 9.38 | 8.43 | 9.21 | 163.28 |
| *PDE4A* | CHEMBL74078 | 9.13 | 9.38 | 8.43 | 9.21 | 163.28 |
| *PDE4A* | IBUDILAST | 9.13 | 9.38 | 8.24 | 9.21 | 161.56 |
| *PDE4A* | TOFISOPAM | 9.13 | 9.38 | 8.01 | 9.21 | 159.37 |
| *PDE4A* | MILRINONE | 9.13 | 9.38 | 8.01 | 9.21 | 159.37 |
| *PDE4A* | TOFIMILAST | 9.13 | 9.38 | 7.86 | 9.21 | 157.98 |
| *PDE4A* | ROLIPRAM | 9.13 | 9.38 | 7.86 | 9.21 | 157.98 |
| *PDE4A* | DENBUFYLLINE | 9.13 | 9.38 | 7.86 | 9.21 | 157.98 |
| *PDE4A* | HT-0712 | 9.13 | 9.38 | 7.86 | 9.21 | 157.98 |
| *PDE4A* | GSK-356278 | 9.13 | 9.38 | 7.86 | 9.21 | 157.98 |
| *PDE4A* | PROPENTOFYLLINE | 9.13 | 9.38 | 7.86 | 9.21 | 157.98 |
| *PDE4A* | ILOPROST | 9.13 | 9.38 | 7.86 | 9.21 | 157.98 |
| *PDE4A* | THEOPHYLLINE | 9.13 | 9.38 | 7.64 | 9.21 | 156.00 |
| *IFNAR2* | INTERFERON ALFA-N3 | 5.81 | 9.95 | 9.83 | 9.93 | 155.66 |
| *PDE4A* | CDC-801 | 9.13 | 9.38 | 7.48 | 9.21 | 154.47 |

| | | | | | | |
|---|---|---|---|---|---|---|
| *PDE4A* | CDP840 | 9.13 | 9.38 | 7.48 | 9.21 | 154.47 |
| *PDE4A* | TADALAFIL | 9.13 | 9.38 | 7.48 | 9.21 | 154.47 |
| *VIPR2* | SECRETIN | 9.47 | 8.49 | 9.65 | 7.55 | 154.45 |
| *IFNAR2* | INTERFERON ALFA-2B | 5.81 | 9.95 | 9.67 | 9.93 | 154.41 |
| *VIPR2* | VASOACTIVE INTESTINAL PEPTIDE | 9.47 | 8.49 | 9.54 | 7.55 | 153.47 |
| *IFNAR2* | PEGINTERFERON ALFA-2A | 5.81 | 9.95 | 9.00 | 9.93 | 149.12 |
| *IFNAR2* | NYLIDRIN | 5.81 | 9.95 | 9.00 | 9.93 | 149.12 |
| *IFNAR2* | INTERFERON ALFACON-1 | 5.81 | 9.95 | 9.00 | 9.93 | 149.12 |
| *IFNAR2* | PEGINTERFERON BETA-1A | 5.81 | 9.95 | 9.00 | 9.93 | 149.12 |
| *IFNAR2* | INTERFERON BETA-1B | 5.81 | 9.95 | 9.00 | 9.93 | 149.12 |
| *IFNAR2* | ANIFROLUMAB | 5.81 | 9.95 | 9.00 | 9.93 | 149.12 |
| *IFNAR2* | ALBINTERFERON ALFA-2B | 5.81 | 9.95 | 9.00 | 9.93 | 149.12 |
| *IFNAR2* | PEGINTERFERON ALFA-2B | 5.81 | 9.95 | 9.00 | 9.93 | 149.12 |
| *IFNAR2* | INTERFERON BETA-1A | 5.81 | 9.95 | 9.00 | 9.93 | 149.12 |
| *IFNAR2* | INTERFERON ALFA-2A | 5.81 | 9.95 | 9.00 | 9.93 | 149.12 |
| *PDE4A* | FLAVOXATE HYDROCHLORID E | 9.13 | 9.38 | 6.85 | 9.21 | 148.67 |
| *PDE4A* | PENTOXIFYLLINE | 9.13 | 9.38 | 6.80 | 9.21 | 148.18 |
| *PDE4A* | THEOPHYLLINE SODIUM GLYCINATE | 9.13 | 9.38 | 6.65 | 9.21 | 146.81 |
| *PDE4A* | INAMRINONE | 9.13 | 9.38 | 6.65 | 9.21 | 146.81 |
| *IFNAR2* | SIFALIMUMAB | 5.81 | 9.95 | 8.68 | 9.93 | 146.53 |
| *P4HA2* | PROLINE | 8.41 | 8.15 | 9.59 | 8.09 | 146.42 |
| *PDE4A* | AMINOPHYLLINE | 9.13 | 9.38 | 6.60 | 9.21 | 146.37 |
| *PDE4A* | DIPYRIDAMOLE | 9.13 | 9.38 | 6.60 | 9.21 | 146.37 |
| *SLC5A3* | PHLORIZIN | 7.25 | 8.33 | 9.94 | 8.57 | 144.28 |
| *PDE4A* | ETAZOLATE | 9.13 | 9.38 | 6.16 | 9.21 | 142.29 |
| *PDE4A* | AMLEXANOX | 9.13 | 9.38 | 6.06 | 9.21 | 141.39 |
| *SENP7* | CHEMBL378903 | 7.69 | 7.61 | 8.55 | 9.68 | 139.46 |
| *LTA* | PATECLIZUMAB | 6.77 | 8.17 | 9.70 | 8.63 | 136.90 |
| *SENP7* | XANTHOANGELO L B | 7.69 | 7.61 | 8.18 | 9.68 | 136.58 |
| *SENP7* | CHEMBL546865 | 7.69 | 7.61 | 7.99 | 9.68 | 135.16 |
| *SENP7* | CHEMBL584668 | 7.69 | 7.61 | 7.99 | 9.68 | 135.16 |
| *LTA* | BAMINERCEPT | 6.77 | 8.17 | 9.45 | 8.63 | 135.06 |
| *SENP7* | DIHYDROXANTH OHUMOL | 7.69 | 7.61 | 7.89 | 9.68 | 134.39 |
| *SENP7* | CHEMBL546170 | 7.69 | 7.61 | 7.89 | 9.68 | 134.39 |
| *SENP7* | CHEMBL412603 | 7.69 | 7.61 | 7.71 | 9.68 | 132.99 |
| *SENP7* | CHEMBL484663 | 7.69 | 7.61 | 7.71 | 9.68 | 132.99 |
| *TYK2* | TOFACITINIB | 7.89 | 6.94 | 8.39 | 9.54 | 132.87 |

| | | | | | | |
|---|---|---|---|---|---|---|
| *LTA* | ETANERCEPT | 6.77 | 8.17 | 9.13 | 8.63 | 132.68 |
| *TYK2* | BREPOCITINIB | 7.89 | 6.94 | 8.30 | 9.54 | 132.21 |
| *SENP7* | CHEMBL1493528 | 7.69 | 7.61 | 7.55 | 9.68 | 131.79 |
| *SENP7* | CHEMBL602561 | 7.69 | 7.61 | 7.55 | 9.68 | 131.79 |
| *SENP7* | CHEMBL600313 | 7.69 | 7.61 | 7.55 | 9.68 | 131.79 |
| *COL11A2* | COLLAGENASE CLOSTRIDIUM HISTOLYTICUM | 9.44 | 7.23 | 8.45 | 7.30 | 131.33 |
| *HCN3* | ZATEBRADINE | 8.15 | 7.02 | 9.38 | 7.91 | 131.06 |
| *TYK2* | PEFICITINIB | 7.89 | 6.94 | 8.14 | 9.54 | 130.99 |
| *TYK2* | CHEMBL21156 | 7.89 | 6.94 | 8.14 | 9.54 | 130.99 |
| *HCN3* | CILOBRADINE | 8.15 | 7.02 | 9.32 | 7.91 | 130.63 |
| *HCN3* | IVABRADINE | 8.15 | 7.02 | 9.23 | 7.91 | 129.94 |
| *SENP7* | CHEMBL576208 | 7.69 | 7.61 | 7.30 | 9.68 | 129.87 |
| *PLEKHA4* | CHEMBL23552 | 8.13 | 5.35 | 9.89 | 9.35 | 129.67 |
| *SENP7* | IPRIFLAVONE | 7.69 | 7.61 | 7.23 | 9.68 | 129.35 |
| *COL11A2* | OCRIPLASMIN | 9.44 | 7.23 | 8.20 | 7.30 | 129.26 |
| *CLK2* | LORECIVIVINT | 7.77 | 7.80 | 8.54 | 8.01 | 128.75 |
| *SENP7* | CHEMBL1504679 | 7.69 | 7.61 | 7.13 | 9.68 | 128.62 |
| *SENP7* | CHEMBL428789 | 7.69 | 7.61 | 7.03 | 9.68 | 127.86 |
| *LTA* | ABACAVIR | 6.77 | 8.17 | 8.46 | 8.63 | 127.64 |
| *TYK2* | TOFACITINIB CITRATE | 7.89 | 6.94 | 7.64 | 9.54 | 127.33 |
| *TYK2* | OCLACITINIB | 7.89 | 6.94 | 7.64 | 9.54 | 127.33 |
| *TYK2* | SOLCITINIB | 7.89 | 6.94 | 7.64 | 9.54 | 127.33 |
| *TYK2* | BMS-911543 | 7.89 | 6.94 | 7.64 | 9.54 | 127.33 |
| *TYK2* | DELGOCITINIB | 7.89 | 6.94 | 7.64 | 9.54 | 127.33 |
| *TYK2* | NS-018 | 7.89 | 6.94 | 7.64 | 9.54 | 127.33 |
| *TYK2* | FILGOTINIB | 7.89 | 6.94 | 7.64 | 9.54 | 127.33 |
| *CA11* | ZONISAMIDE | 7.86 | 6.21 | 9.63 | 8.44 | 127.12 |
| *SENP7* | CHEMBL299853 | 7.69 | 7.61 | 6.89 | 9.68 | 126.73 |
| *TYK2* | CERDULATINIB | 7.89 | 6.94 | 7.52 | 9.54 | 126.38 |
| *IMPG2* | HYALURONATE SODIUM | 8.58 | 5.32 | 9.57 | 8.62 | 126.34 |
| *SLC22A4* | LEVOCARNITINE | 9.12 | 4.41 | 9.72 | 8.84 | 125.60 |
| *CLK2* | LEUCETTAMINE B | 7.77 | 7.80 | 8.06 | 8.01 | 125.06 |
| *TYK2* | FEDRATINIB | 7.89 | 6.94 | 7.19 | 9.54 | 123.97 |
| *TYK2* | UPADACITINIB | 7.89 | 6.94 | 7.19 | 9.54 | 123.97 |
| *LTA* | CARBAMAZEPINE | 6.77 | 8.17 | 7.79 | 8.63 | 122.63 |
| *TYK2* | BARICITINIB | 7.89 | 6.94 | 6.96 | 9.54 | 122.24 |
| *PTGFR* | TRAVOPROST | 9.34 | 9.14 | 9.98 | 3.20 | 121.78 |
| *SACM1L* | BUPROPION | 8.19 | 4.97 | 9.47 | 8.99 | 121.51 |
| *CLK2* | GATIFLOXACIN | 7.77 | 7.80 | 7.52 | 8.01 | 120.81 |
| *PTGFR* | LATANOPROST | 9.34 | 9.14 | 9.86 | 3.20 | 120.60 |
| *TYK2* | ZOTIRACICLIB | 7.89 | 6.94 | 6.73 | 9.54 | 120.56 |
| *PTGFR* | TAFLUPROST | 9.34 | 9.14 | 9.83 | 3.20 | 120.36 |
| *PTGFR* | BIMATOPROST | 9.34 | 9.14 | 9.82 | 3.20 | 120.25 |

**Supplementary Table S15. Significant pathways enriched by up-regulated genes in intestinal tuft positive cells**

| Description | Gene size | Enrichment ratio | P Value | FDR |
|---|---|---|---|---|
| Ribosome | 153 | 6.02 | <2.2e-16 | 2.20E-16 |
| Relaxin signaling pathway | 130 | 3.22 | 3.04E-06 | 4.23E-04 |
| TNF signaling pathway | 110 | 3.42 | 3.89E-06 | 4.23E-04 |
| Human cytomegalovirus infection | 225 | 2.23 | 1.74E-04 | 1.42E-02 |
| Mitophagy | 65 | 3.54 | 2.23E-04 | 1.45E-02 |
| Protein digestion and absorption | 90 | 3.02 | 3.17E-04 | 1.72E-02 |
| IL-17 signaling pathway | 93 | 2.92 | 4.39E-04 | 2.05E-02 |
| Focal adhesion | 199 | 2.21 | 5.10E-04 | 2.08E-02 |
| Kaposi sarcoma-associated herpesvirus infection | 186 | 2.14 | 1.38E-03 | 5.00E-02 |
| Fluid shear stress and atherosclerosis | 139 | 2.26 | 2.53E-03 | 0.076 |
| AGE-RAGE signaling pathway in diabetic complications | 99 | 2.54 | 2.55E-03 | 0.076 |
| Legionellosis | 55 | 3.04 | 4.28E-03 | 0.111 |
| JAK-STAT signaling pathway | 162 | 2.07 | 4.50E-03 | 0.111 |
| cGMP-PKG signaling pathway | 163 | 2.05 | 4.78E-03 | 0.111 |
| Prostate cancer | 97 | 2.37 | 6.33E-03 | 0.138 |
| Osteoclast differentiation | 128 | 2.12 | 7.96E-03 | 0.156 |
| Cocaine addiction | 49 | 2.99 | 8.15E-03 | 0.156 |
| Human T-cell leukemia virus 1 infection | 255 | 1.72 | 1.03E-02 | 0.180 |
| PI3K-Akt signaling pathway | 354 | 1.60 | 1.05E-02 | 0.180 |
| Vascular smooth muscle contraction | 121 | 2.07 | 1.26E-02 | 0.206 |

**Supplementary Table S16. scDrugHunter-identified top-ranked gene-drug interaction pairs in intestinal tuft positive cells (scDGS > 120)**

| Gene | Drug | Gene specificity rank | Gene correlation rank | Gene-drug interaction rank | SMultiXcan-based gene P value rank | scDGS |
|---|---|---|---|---|---|---|
| *IL10RB* | PEGINTERFERON LAMBDA-1A | 8.33 | 8.53 | 9.94 | 9.86 | 166.83 |
| *ICAM1* | ALICAFORSEN | 7.67 | 9.20 | 9.79 | 8.98 | 158.25 |
| *ICAM1* | BI-505 | 7.67 | 9.20 | 9.69 | 8.98 | 157.46 |
| *ICAM1* | ENLIMOMAB PEGOL | 7.67 | 9.20 | 9.57 | 8.98 | 156.42 |
| *CCR1* | CCX354 | 8.94 | 6.78 | 9.98 | 9.87 | 156.03 |
| *CCR1* | CHEMBL2205805 | 8.94 | 6.78 | 9.91 | 9.87 | 155.53 |
| *CCR1* | BMS-817399 | 8.94 | 6.78 | 9.91 | 9.87 | 155.53 |
| *ICAM1* | LIFITEGRAST | 7.67 | 9.20 | 9.42 | 8.98 | 155.13 |
| *PDE4A* | DROTAVERINE | 8.82 | 7.86 | 9.27 | 9.32 | 155.01 |
| *PDE4A* | ENPROFYLLINE | 8.82 | 7.86 | 9.16 | 9.32 | 154.10 |
| *CCR1* | AZD4818 | 8.94 | 6.78 | 9.71 | 9.87 | 153.98 |

| | | | | | | |
|---|---|---|---|---|---|---|
| *ICAM1* | NATALIZUMAB | 7.67 | 9.20 | 9.13 | 8.98 | 152.67 |
| *ICAM1* | HYALURONAN | 7.67 | 9.20 | 9.13 | 8.98 | 152.67 |
| *PDE4A* | APREMILAST | 8.82 | 7.86 | 8.97 | 9.32 | 152.50 |
| *PDE4A* | ROFLUMILAST | 8.82 | 7.86 | 8.93 | 9.32 | 152.15 |
| *PDE4A* | DYPHYLLINE | 8.82 | 7.86 | 8.85 | 9.32 | 151.45 |
| *PDE4A* | AROFYLLINE | 8.82 | 7.86 | 8.78 | 9.32 | 150.87 |
| *PDE4A* | OGLEMILAST | 8.82 | 7.86 | 8.78 | 9.32 | 150.87 |
| *PDE4A* | CHEMBL1229585 | 8.82 | 7.86 | 8.78 | 9.32 | 150.87 |
| *PDE4A* | LOTAMILAST | 8.82 | 7.86 | 8.78 | 9.32 | 150.87 |
| *TNFRSF4* | IVUXOLIMAB | 8.10 | 8.39 | 9.87 | 8.41 | 150.69 |
| *TNFRSF4* | VONLEROLIZUMAB | 8.10 | 8.39 | 9.87 | 8.41 | 150.69 |
| *TNFRSF4* | TAVOLIMAB | 8.10 | 8.39 | 9.87 | 8.41 | 150.69 |
| *CCR1* | TERPYRIDINE | 8.94 | 6.78 | 9.23 | 9.87 | 150.15 |
| *PDE4A* | CRISABOROLE | 8.82 | 7.86 | 8.64 | 9.32 | 149.72 |
| *PDE4A* | PICLAMILAST | 8.82 | 7.86 | 8.60 | 9.32 | 149.40 |
| *PDE4A* | OXTRIPHYLLINE | 8.82 | 7.86 | 8.57 | 9.32 | 149.17 |
| *PDE4A* | REVAMILAST | 8.82 | 7.86 | 8.51 | 9.32 | 148.67 |
| *PDE4A* | MK-0873 | 8.82 | 7.86 | 8.51 | 9.32 | 148.67 |
| *PDE4A* | LIRIMILAST | 8.82 | 7.86 | 8.51 | 9.32 | 148.67 |
| *PDE4A* | CHEMBL74078 | 8.82 | 7.86 | 8.51 | 9.32 | 148.67 |
| *PDE4A* | IBUDILAST | 8.82 | 7.86 | 8.34 | 9.32 | 147.24 |
| *ICAM1* | NAFAMOSTAT | 7.67 | 9.20 | 8.45 | 8.98 | 146.95 |
| *PDE4A* | TOFISOPAM | 8.82 | 7.86 | 8.11 | 9.32 | 145.34 |
| *PDE4A* | MILRINONE | 8.82 | 7.86 | 8.11 | 9.32 | 145.34 |
| *TYK2* | TOFACITINIB | 7.41 | 8.61 | 8.48 | 9.63 | 145.05 |
| *SENP7* | CHEMBL378903 | 6.69 | 9.06 | 8.63 | 9.78 | 145.01 |
| *TYK2* | BREPOCITINIB | 7.41 | 8.61 | 8.40 | 9.63 | 144.38 |
| *PDE4A* | TOFIMILAST | 8.82 | 7.86 | 7.96 | 9.32 | 144.04 |
| *PDE4A* | ROLIPRAM | 8.82 | 7.86 | 7.96 | 9.32 | 144.04 |
| *PDE4A* | DENBUFYLLINE | 8.82 | 7.86 | 7.96 | 9.32 | 144.04 |
| *PDE4A* | HT-0712 | 8.82 | 7.86 | 7.96 | 9.32 | 144.04 |
| *PDE4A* | GSK-356278 | 8.82 | 7.86 | 7.96 | 9.32 | 144.04 |
| *PDE4A* | PROPENTOFYLLINE | 8.82 | 7.86 | 7.96 | 9.32 | 144.04 |
| *PDE4A* | ILOPROST | 8.82 | 7.86 | 7.96 | 9.32 | 144.04 |
| *P4HA2* | PROLINE | 6.16 | 9.97 | 9.63 | 8.19 | 143.69 |
| *DBP* | ALBUMIN HUMAN | 8.13 | 7.53 | 9.60 | 8.71 | 143.42 |
| *TYK2* | PEFICITINIB | 7.41 | 8.61 | 8.23 | 9.63 | 143.08 |
| *TYK2* | CHEMBL21156 | 7.41 | 8.61 | 8.23 | 9.63 | 143.08 |
| *CLK3* | CHEMBL1236620 | 6.38 | 9.09 | 9.71 | 8.70 | 142.35 |
| *PDE4A* | THEOPHYLLINE | 8.82 | 7.86 | 7.75 | 9.32 | 142.31 |
| *BGLAP* | PHYTONADIONE | 8.25 | 8.34 | 9.54 | 7.62 | 142.27 |
| *ICAM1* | HYALURONATE SODIUM | 7.67 | 9.20 | 7.89 | 8.98 | 142.26 |
| *SENP7* | XANTHOANGELOL B | 6.69 | 9.06 | 8.27 | 9.78 | 142.20 |
| *PLEKHA4* | CHEMBL23552 | 8.02 | 6.60 | 9.90 | 9.47 | 141.46 |
| *PDE4A* | CDC-801 | 8.82 | 7.86 | 7.58 | 9.32 | 140.91 |

| | | | | | | |
|---|---|---|---|---|---|---|
| *PDE4A* | CDP840 | 8.82 | 7.86 | 7.58 | 9.32 | 140.91 |
| *PDE4A* | TADALAFIL | 8.82 | 7.86 | 7.58 | 9.32 | 140.91 |
| *SENP7* | CHEMBL546865 | 6.69 | 9.06 | 8.09 | 9.78 | 140.80 |
| *SENP7* | CHEMBL584668 | 6.69 | 9.06 | 8.09 | 9.78 | 140.80 |
| *SENP7* | DIHYDROXANTHOHU MOL | 6.69 | 9.06 | 7.99 | 9.78 | 139.97 |
| *SENP7* | CHEMBL546170 | 6.69 | 9.06 | 7.99 | 9.78 | 139.97 |
| *CLK3* | CHEMBL261720 | 6.38 | 9.09 | 9.39 | 8.70 | 139.86 |
| *TYK2* | TOFACITINIB CITRATE | 7.41 | 8.61 | 7.75 | 9.63 | 139.20 |
| *TYK2* | OCLACITINIB | 7.41 | 8.61 | 7.75 | 9.63 | 139.20 |
| *TYK2* | SOLCITINIB | 7.41 | 8.61 | 7.75 | 9.63 | 139.20 |
| *TYK2* | BMS-911543 | 7.41 | 8.61 | 7.75 | 9.63 | 139.20 |
| *TYK2* | DELGOCITINIB | 7.41 | 8.61 | 7.75 | 9.63 | 139.20 |
| *TYK2* | NS-018 | 7.41 | 8.61 | 7.75 | 9.63 | 139.20 |
| *TYK2* | FILGOTINIB | 7.41 | 8.61 | 7.75 | 9.63 | 139.20 |
| *CLK3* | LEUCETTAMINE B | 6.38 | 9.09 | 9.29 | 8.70 | 139.10 |
| *SENP7* | CHEMBL412603 | 6.69 | 9.06 | 7.81 | 9.78 | 138.58 |
| *SENP7* | CHEMBL484663 | 6.69 | 9.06 | 7.81 | 9.78 | 138.58 |
| *TYK2* | CERDULATINIB | 7.41 | 8.61 | 7.62 | 9.63 | 138.17 |
| *BGLAP* | PARICALCITOL | 8.25 | 8.34 | 9.00 | 7.62 | 137.81 |
| *SENP7* | CHEMBL1493528 | 6.69 | 9.06 | 7.65 | 9.78 | 137.32 |
| *SENP7* | CHEMBL602561 | 6.69 | 9.06 | 7.65 | 9.78 | 137.32 |
| *SENP7* | CHEMBL600313 | 6.69 | 9.06 | 7.65 | 9.78 | 137.32 |
| *PDE4A* | FLAVOXATE HYDROCHLORIDE | 8.82 | 7.86 | 6.96 | 9.32 | 135.75 |
| *TYK2* | FEDRATINIB | 7.41 | 8.61 | 7.30 | 9.63 | 135.62 |
| *TYK2* | UPADACITINIB | 7.41 | 8.61 | 7.30 | 9.63 | 135.62 |
| *BGLAP* | CHEMBL38397 | 8.25 | 8.34 | 8.72 | 7.62 | 135.50 |
| *SENP7* | CHEMBL576208 | 6.69 | 9.06 | 7.41 | 9.78 | 135.40 |
| *PDE4A* | PENTOXIFYLLINE | 8.82 | 7.86 | 6.91 | 9.32 | 135.30 |
| *CLK3* | HARMINE | 6.38 | 9.09 | 8.80 | 8.70 | 135.24 |
| *SENP7* | IPRIFLAVONE | 6.69 | 9.06 | 7.34 | 9.78 | 134.86 |
| *IFNAR2* | INTERFERON ALFA-N3 | 3.59 | 10.00 | 9.85 | 9.93 | 134.36 |
| *SENP7* | CHEMBL1504679 | 6.69 | 9.06 | 7.24 | 9.78 | 134.10 |
| *PDE4A* | THEOPHYLLINE SODIUM GLYCINATE | 8.82 | 7.86 | 6.76 | 9.32 | 134.04 |
| *PDE4A* | INAMRINONE | 8.82 | 7.86 | 6.76 | 9.32 | 134.04 |
| *TYK2* | BARICITINIB | 7.41 | 8.61 | 7.07 | 9.63 | 133.74 |
| *PDE4A* | AMINOPHYLLINE | 8.82 | 7.86 | 6.71 | 9.32 | 133.64 |
| *PDE4A* | DIPYRIDAMOLE | 8.82 | 7.86 | 6.71 | 9.32 | 133.64 |
| *VIPR2* | SECRETIN | 9.16 | 6.19 | 9.68 | 7.71 | 133.47 |
| *BGLAP* | GALLIUM NITRATE | 8.25 | 8.34 | 8.47 | 7.62 | 133.40 |
| *IFNAR2* | INTERFERON ALFA-2B | 3.59 | 10.00 | 9.70 | 9.93 | 133.34 |
| *SENP7* | CHEMBL428789 | 6.69 | 9.06 | 7.14 | 9.78 | 133.30 |
| *VIPR2* | VASOACTIVE INTESTINAL PEPTIDE | 9.16 | 6.19 | 9.57 | 7.71 | 132.66 |
| *BGLAP* | ISOFLAVONE | 8.25 | 8.34 | 8.35 | 7.62 | 132.43 |

| | | | | | | |
|---|---|---|---|---|---|---|
| *SENP7* | CHEMBL299853 | 6.69 | 9.06 | 7.00 | 9.78 | 132.16 |
| *TYK2* | ZOTIRACICLIB | 7.41 | 8.61 | 6.84 | 9.63 | 131.91 |
| *BGLAP* | CALCITONIN | 8.25 | 8.34 | 8.26 | 7.62 | 131.66 |
| *TYK2* | RUXOLITINIB | 7.41 | 8.61 | 6.71 | 9.63 | 130.88 |
| *TYK2* | ALDESLEUKIN | 7.41 | 8.61 | 6.62 | 9.63 | 130.13 |
| *TYK2* | AT-9283 | 7.41 | 8.61 | 6.62 | 9.63 | 130.13 |
| *PDE4A* | ETAZOLATE | 8.82 | 7.86 | 6.26 | 9.32 | 129.91 |
| *PDE4A* | AMLEXANOX | 8.82 | 7.86 | 6.16 | 9.32 | 129.08 |
| *IFNAR2* | PEGINTERFERON ALFA-2A | 3.59 | 10.00 | 9.05 | 9.93 | 128.95 |
| *IFNAR2* | NYLIDRIN | 3.59 | 10.00 | 9.05 | 9.93 | 128.95 |
| *IFNAR2* | INTERFERON ALFACON-1 | 3.59 | 10.00 | 9.05 | 9.93 | 128.95 |
| *IFNAR2* | PEGINTERFERON BETA-1A | 3.59 | 10.00 | 9.05 | 9.93 | 128.95 |
| *IFNAR2* | INTERFERON BETA-1B | 3.59 | 10.00 | 9.05 | 9.93 | 128.95 |
| *IFNAR2* | ANIFROLUMAB | 3.59 | 10.00 | 9.05 | 9.93 | 128.95 |
| *IFNAR2* | ALBINTERFERON ALFA-2B | 3.59 | 10.00 | 9.05 | 9.93 | 128.95 |
| *IFNAR2* | PEGINTERFERON ALFA-2B | 3.59 | 10.00 | 9.05 | 9.93 | 128.95 |
| *IFNAR2* | INTERFERON BETA-1A | 3.59 | 10.00 | 9.05 | 9.93 | 128.95 |
| *IFNAR2* | INTERFERON ALFA-2A | 3.59 | 10.00 | 9.05 | 9.93 | 128.95 |
| *BGLAP* | CLODRONIC ACID | 8.25 | 8.34 | 7.69 | 7.62 | 126.95 |
| *IFNAR2* | SIFALIMUMAB | 3.59 | 10.00 | 8.76 | 9.93 | 126.93 |
| *BGLAP* | ANDROSTANOLONE | 8.25 | 8.34 | 7.58 | 7.62 | 126.05 |
| *TYK2* | JNJ-7706621 | 7.41 | 8.61 | 6.08 | 9.63 | 125.80 |
| *JAK1* | RUXOLITINIB | 8.54 | 6.35 | 8.92 | 7.90 | 125.23 |
| *BGLAP* | MITOMYCIN | 8.25 | 8.34 | 7.47 | 7.62 | 125.16 |
| *BGLAP* | COUMARIN | 8.25 | 8.34 | 7.47 | 7.62 | 125.16 |
| *JAK1* | ITACITINIB | 8.54 | 6.35 | 8.86 | 7.90 | 124.76 |
| *JAK1* | GLPG-0555 | 8.54 | 6.35 | 8.86 | 7.90 | 124.76 |
| *JAK1* | INCB-047986 | 8.54 | 6.35 | 8.86 | 7.90 | 124.76 |
| *BGLAP* | EXEMESTANE | 8.25 | 8.34 | 7.34 | 7.62 | 124.06 |
| *JAK1* | UPADACITINIB | 8.54 | 6.35 | 8.75 | 7.90 | 123.93 |
| *JAK1* | PEFICITINIB | 8.54 | 6.35 | 8.71 | 7.90 | 123.62 |
| *THRA* | LIOTRIX | 8.72 | 8.15 | 9.89 | 4.75 | 123.51 |
| *BGLAP* | ETIDRONIC ACID | 8.25 | 8.34 | 7.27 | 7.62 | 123.49 |
| *JAK1* | BARICITINIB | 8.54 | 6.35 | 8.65 | 7.90 | 123.22 |
| *JAK1* | FILGOTINIB | 8.54 | 6.35 | 8.65 | 7.90 | 123.22 |
| *JAK1* | SOLCITINIB | 8.54 | 6.35 | 8.65 | 7.90 | 123.22 |
| *BGLAP* | REGRAMOSTIM | 8.25 | 8.34 | 7.21 | 7.62 | 123.01 |
| *JAK1* | ABROCITINIB | 8.54 | 6.35 | 8.62 | 7.90 | 122.95 |
| *BGLAP* | METHIMAZOLE | 8.25 | 8.34 | 7.11 | 7.62 | 122.14 |
| *BGLAP* | PROPRANOLOL | 8.25 | 8.34 | 7.07 | 7.62 | 121.79 |
| *BGLAP* | INFLIXIMAB | 8.25 | 8.34 | 7.03 | 7.62 | 121.47 |
| *PTGFR* | TRAVOPROST | 9.13 | 9.22 | 9.98 | 3.25 | 121.44 |
| *THRA* | DEXTROTHYROXINE | 8.72 | 8.15 | 9.65 | 4.75 | 121.43 |

| | | | | | | |
|---|---|---|---|---|---|---|
| *JAK1* | BREPOCITINIB | 8.54 | 6.35 | 8.40 | 7.90 | 121.30 |
| *JAK1* | MOMELOTINIB | 8.54 | 6.35 | 8.40 | 7.90 | 121.30 |
| *JAK1* | FEDRATINIB | 8.54 | 6.35 | 8.40 | 7.90 | 121.30 |
| *JAK1* | TRICETAMIDE | 8.54 | 6.35 | 8.40 | 7.90 | 121.30 |
| *JAK1* | RUXOLITINIB PHOSPHATE | 8.54 | 6.35 | 8.40 | 7.90 | 121.30 |
| *PTGFR* | LATANOPROST | 9.13 | 9.22 | 9.87 | 3.25 | 120.42 |
| *PTGFR* | TAFLUPROST | 9.13 | 9.22 | 9.84 | 3.25 | 120.17 |
| *JAK1* | CHEMBL21156 | 8.54 | 6.35 | 8.23 | 7.90 | 120.09 |
| *PTGFR* | BIMATOPROST | 9.13 | 9.22 | 9.83 | 3.25 | 120.06 |

**Supplementary Table S17. Significant pathways enriched by up-regulated genes in brain endothelial positive cells**

| Description | Gene size | Enrichment ratio | P Value | FDR |
|---|---|---|---|---|
| Pathways in cancer | 524 | 2.78 | 5.21E-08 | 1.55E-05 |
| Protein digestion and absorption | 90 | 6.37 | 9.53E-08 | 1.55E-05 |
| AGE-RAGE signaling pathway in diabetic complications | 99 | 5.79 | 3.01E-07 | 2.68E-05 |
| PI3K-Akt signaling pathway | 354 | 3.11 | 3.29E-07 | 2.68E-05 |
| Focal adhesion | 199 | 3.99 | 4.93E-07 | 3.21E-05 |
| ECM-receptor interaction | 82 | 5.37 | 1.44E-05 | 7.84E-04 |
| Relaxin signaling pathway | 130 | 4.07 | 3.55E-05 | 1.29E-03 |
| MAPK signaling pathway | 295 | 2.84 | 3.55E-05 | 1.29E-03 |
| TNF signaling pathway | 110 | 4.41 | 3.60E-05 | 1.29E-03 |
| Proteoglycans in cancer | 198 | 3.34 | 3.95E-05 | 1.29E-03 |
| Small cell lung cancer | 92 | 4.31 | 2.22E-04 | 6.58E-03 |
| Fluid shear stress and atherosclerosis | 138 | 3.51 | 2.81E-04 | 7.64E-03 |
| Prostate cancer | 97 | 4.09 | 3.32E-04 | 8.32E-03 |
| Complement and coagulation cascades | 79 | 4.46 | 3.94E-04 | 9.17E-03 |
| MicroRNAs in cancer | 150 | 3.23 | 5.76E-04 | 1.25E-02 |
| Human T-cell leukemia virus 1 infection | 255 | 2.59 | 6.47E-04 | 1.32E-02 |
| Malaria | 49 | 5.40 | 7.77E-04 | 1.49E-02 |
| Amphetamine addiction | 68 | 4.54 | 8.26E-04 | 1.50E-02 |
| Melanoma | 72 | 4.28 | 1.16E-03 | 2.00E-02 |
| Protein processing in endoplasmic reticulum | 165 | 2.94 | 1.27E-03 | 2.07E-02 |
| Pertussis | 76 | 4.06 | 1.60E-03 | 2.49E-02 |
| Human papillomavirus infection | 339 | 2.21 | 1.70E-03 | 2.51E-02 |
| EGFR tyrosine kinase inhibitor resistance | 79 | 3.90 | 2.01E-03 | 2.84E-02 |
| Osteoclast differentiation | 128 | 3.10 | 2.43E-03 | 3.31E-02 |
| Transcriptional misregulation in cancer | 186 | 2.61 | 3.28E-03 | 4.11E-02 |
| Renin secretion | 65 | 4.07 | 3.41E-03 | 4.11E-02 |
| Central carbon metabolism in cancer | 65 | 4.07 | 3.41E-03 | 4.11E-02 |
| Apoptosis | 136 | 2.92 | 3.66E-03 | 4.14E-02 |
| JAK-STAT signaling pathway | 162 | 2.72 | 3.68E-03 | 4.14E-02 |
| Toxoplasmosis | 112 | 3.15 | 3.82E-03 | 4.15E-02 |
| Signaling pathways regulating pluripotency of stem cells | 139 | 2.85 | 4.23E-03 | 4.45E-02 |

| | | | | |
|---|---|---|---|---|
| Human cytomegalovirus infection | 225 | 2.35 | 5.00E-03 | 4.98E-02 |
| IL-17 signaling pathway | 93 | 3.32 | 5.04E-03 | 4.98E-02 |

**Supplementary Table S18. Significant GO-term (biological process) enriched by up-regulated genes in brain endothelial positive cells**

| Description | Gene size | Enrichment ratio | P Value | FDR |
|---|---|---|---|---|
| ossification | 371 | 5.39 | 1.00E-16 | 1.00E-16 |
| angiogenesis | 487 | 4.62 | 1.00E-16 | 1.00E-16 |
| extracellular structure organization | 400 | 6.25 | 1.00E-16 | 1.00E-16 |
| connective tissue development | 265 | 5.47 | 8.75E-14 | 1.86E-11 |
| epithelial cell proliferation | 372 | 4.57 | 1.17E-13 | 1.99E-11 |
| reproductive system development | 428 | 3.97 | 6.31E-12 | 8.50E-10 |
| response to oxygen levels | 337 | 4.45 | 7.00E-12 | 8.50E-10 |
| in utero embryonic development | 345 | 4.35 | 1.27E-11 | 1.29E-09 |
| negative regulation of immune system process | 416 | 3.97 | 1.37E-11 | 1.29E-09 |
| morphogenesis of an epithelium | 480 | 3.65 | 3.37E-11 | 2.87E-09 |
| morphogenesis of a branching structure | 196 | 5.61 | 5.89E-11 | 4.55E-09 |
| negative regulation of cellular component movement | 301 | 4.49 | 7.00E-11 | 4.96E-09 |
| negative regulation of intracellular signal transduction | 495 | 3.54 | 7.90E-11 | 5.17E-09 |
| embryonic organ development | 423 | 3.78 | 9.79E-11 | 5.94E-09 |
| negative regulation of locomotion | 314 | 4.30 | 1.83E-10 | 1.04E-08 |
| respiratory tube development | 172 | 5.82 | 2.38E-10 | 1.26E-08 |
| respiratory system development | 193 | 5.44 | 2.92E-10 | 1.38E-08 |
| positive regulation of cell motility | 493 | 3.45 | 2.92E-10 | 1.38E-08 |
| muscle tissue development | 371 | 3.91 | 3.64E-10 | 1.63E-08 |
| regulation of DNA-templated transcription in response to stress | 78 | 8.98 | 3.89E-10 | 1.65E-08 |
| tissue migration | 283 | 4.42 | 5.06E-10 | 2.05E-08 |
| muscle cell proliferation | 182 | 5.50 | 6.57E-10 | 2.54E-08 |
| gland development | 434 | 3.57 | 8.07E-10 | 2.87E-08 |
| collagen metabolic process | 97 | 7.74 | 8.11E-10 | 2.87E-08 |
| regulation of vasculature development | 313 | 4.16 | 8.53E-10 | 2.90E-08 |
| response to steroid hormone | 388 | 3.74 | 1.04E-09 | 3.39E-08 |
| regulation of hemopoiesis | 389 | 3.73 | 1.10E-09 | 3.46E-08 |
| aging | 303 | 4.13 | 2.10E-09 | 6.38E-08 |
| response to transforming growth factor beta | 238 | 4.62 | 2.48E-09 | 7.26E-08 |
| response to acid chemical | 332 | 3.92 | 2.99E-09 | 8.20E-08 |
| cell-substrate adhesion | 332 | 3.92 | 2.99E-09 | 8.20E-08 |
| response to fibroblast growth factor | 142 | 5.99 | 3.49E-09 | 9.28E-08 |
| regulation of apoptotic signaling pathway | 385 | 3.64 | 3.76E-09 | 9.67E-08 |
| fat cell differentiation | 210 | 4.76 | 8.07E-09 | 2.02E-07 |
| urogenital system development | 326 | 3.84 | 9.32E-09 | 2.19E-07 |
| ERK1 and ERK2 cascade | 326 | 3.84 | 9.32E-09 | 2.19E-07 |
| regulation of cellular response to growth factor stimulus | 256 | 4.30 | 9.58E-09 | 2.19E-07 |
| skeletal system morphogenesis | 234 | 4.49 | 9.78E-09 | 2.19E-07 |

| | | | | |
|---|---|---|---|---|
| transmembrane receptor protein serine/threonine kinase signaling pathway | 328 | 3.81 | 1.05E-08 | 2.30E-07 |
| ameboidal-type cell migration | 381 | 3.55 | 1.26E-08 | 2.67E-07 |
| response to temperature stimulus | 201 | 4.48 | 1.18E-07 | 2.09E-06 |
| gastrulation | 175 | 4.29 | 2.36E-06 | 3.04E-05 |
| cardiocyte differentiation | 141 | 4.61 | 5.02E-06 | 6.10E-05 |
| cell fate commitment | 249 | 3.42 | 1.11E-05 | 1.20E-04 |
| cellular response to external stimulus | 330 | 3.03 | 1.11E-05 | 1.20E-04 |
| negative regulation of proteolysis | 336 | 2.98 | 1.45E-05 | 1.54E-04 |
| leukocyte cell-cell adhesion | 319 | 2.82 | 7.77E-05 | 6.67E-04 |
| negative regulation of transferase activity | 266 | 3.01 | 9.25E-05 | 7.63E-04 |
| coagulation | 326 | 2.76 | 1.02E-04 | 8.27E-04 |
| response to organophosphorus | 138 | 3.99 | 1.03E-04 | 8.27E-04 |
| negative regulation of nervous system development | 297 | 2.86 | 1.03E-04 | 8.27E-04 |
| sensory organ morphogenesis | 250 | 3.00 | 1.57E-04 | 1.19E-03 |
| response to toxic substance | 499 | 2.31 | 1.79E-04 | 1.32E-03 |
| regulation of developmental growth | 314 | 2.71 | 2.01E-04 | 1.44E-03 |
| response to ketone | 189 | 3.18 | 4.26E-04 | 2.70E-03 |
| body morphogenesis | 50 | 6.00 | 4.60E-04 | 2.88E-03 |
| response to interferon-beta | 33 | 7.58 | 4.63E-04 | 2.88E-03 |
| cardiac chamber development | 164 | 3.36 | 4.63E-04 | 2.88E-03 |
| response to tumor necrosis factor | 251 | 2.79 | 5.41E-04 | 3.31E-03 |
| regulation of chemotaxis | 205 | 2.93 | 8.77E-04 | 4.97E-03 |
| stem cell division | 40 | 6.25 | 1.14E-03 | 6.28E-03 |
| negative regulation of defense response | 187 | 2.94 | 1.37E-03 | 7.41E-03 |
| inositol lipid-mediated signaling | 165 | 3.03 | 1.80E-03 | 9.48E-03 |
| protein hydroxylation | 27 | 7.41 | 1.91E-03 | 1.00E-02 |
| coronary vasculature development | 47 | 5.32 | 2.38E-03 | 1.21E-02 |
| STAT cascade | 148 | 3.04 | 2.94E-03 | 1.41E-02 |
| response to endoplasmic reticulum stress | 268 | 2.43 | 2.94E-03 | 1.41E-02 |
| nerve development | 77 | 3.90 | 4.38E-03 | 1.98E-02 |
| protein trimerization | 54 | 4.63 | 4.39E-03 | 1.98E-02 |
| viral life cycle | 285 | 2.28 | 4.94E-03 | 2.16E-02 |
| response to interferon-alpha | 20 | 7.50 | 7.01E-03 | 3.01E-02 |
| regulation of protein catabolic process | 370 | 2.03 | 7.68E-03 | 3.25E-02 |
| hormone-mediated signaling pathway | 237 | 2.32 | 8.27E-03 | 3.45E-02 |
| axon development | 490 | 1.84 | 9.83E-03 | 4.00E-02 |
| receptor metabolic process | 183 | 2.46 | 1.14E-02 | 4.54E-02 |

**Supplementary Table S19. scDrugHunter-identified top-ranked gene-drug interaction pairs in brain endothelial positive cells (scDGS > 120)**

| Gene | Drug | Gene specificity rank | Gene correlation rank | Gene-drug interaction rank | SMultiXcan-based gene P value rank | scDGS |
|---|---|---|---|---|---|---|
| *PLEKHA4* | CHEMBL23552 | 9.40 | 9.41 | 9.88 | 9.27 | 180.17 |
| *LTF* | CHEMBL609880 | 9.51 | 8.58 | 9.90 | 9.31 | 173.66 |
| *LTF* | CHEMBL92636 | 9.51 | 8.58 | 9.90 | 9.31 | 173.66 |
| *ICAM1* | ALICAFORSEN | 9.25 | 9.38 | 9.75 | 8.80 | 172.75 |
| *ICAM1* | BI-505 | 9.25 | 9.38 | 9.64 | 8.80 | 171.76 |

| | | | | | | |
|---|---|---|---|---|---|---|
| *LTF* | PARECOXIB | 9.51 | 8.58 | 9.59 | 9.31 | 170.86 |
| *ICAM1* | ENLIMOMAB PEGOL | 9.25 | 9.38 | 9.50 | 8.80 | 170.47 |
| *LTF* | NITRILOTRIACETIC ACID | 9.51 | 8.58 | 9.53 | 9.31 | 170.35 |
| *P4HA2* | PROLINE | 9.57 | 9.73 | 9.57 | 8.03 | 169.80 |
| *LTF* | TALACTOFERRIN ALFA | 9.51 | 8.58 | 9.41 | 9.31 | 169.29 |
| *LTF* | CHEMBL1230004 | 9.51 | 8.58 | 9.41 | 9.31 | 169.29 |
| *ICAM1* | LIFITEGRAST | 9.25 | 9.38 | 9.32 | 8.80 | 168.76 |
| *LTF* | BACITRACIN | 9.51 | 8.58 | 9.12 | 9.31 | 166.62 |
| *ICAM1* | NATALIZUMAB | 9.25 | 9.38 | 9.02 | 8.80 | 165.96 |
| *ICAM1* | HYALURONAN | 9.25 | 9.38 | 9.02 | 8.80 | 165.96 |
| *PDE4A* | DROTAVERINE | 8.74 | 9.20 | 9.16 | 9.14 | 164.09 |
| *PDE4A* | ENPROFYLLINE | 8.74 | 9.20 | 9.06 | 9.14 | 163.14 |
| *PDE4A* | APREMILAST | 8.74 | 9.20 | 8.86 | 9.14 | 161.39 |
| *PDE4A* | ROFLUMILAST | 8.74 | 9.20 | 8.82 | 9.14 | 161.02 |
| *PDE4A* | DYPHYLLINE | 8.74 | 9.20 | 8.71 | 9.14 | 160.02 |
| *IL10RB* | PEGINTERFERON LAMBDA-1A | 8.57 | 7.72 | 9.93 | 9.68 | 159.75 |
| *LTF* | DODECANOATE | 9.51 | 8.58 | 8.33 | 9.31 | 159.50 |
| *PDE4A* | AROFYLLINE | 8.74 | 9.20 | 8.64 | 9.14 | 159.41 |
| *PDE4A* | OGLEMILAST | 8.74 | 9.20 | 8.64 | 9.14 | 159.41 |
| *PDE4A* | CHEMBL1229585 | 8.74 | 9.20 | 8.64 | 9.14 | 159.41 |
| *PDE4A* | LOTAMILAST | 8.74 | 9.20 | 8.64 | 9.14 | 159.41 |
| *ICAM1* | NAFAMOSTAT | 9.25 | 9.38 | 8.30 | 8.80 | 159.31 |
| *CYP3A43* | GINKGO | 9.97 | 8.82 | 9.57 | 7.36 | 159.04 |
| *CSF3* | BENEGRASTIM | 9.14 | 7.86 | 9.75 | 8.94 | 158.70 |
| *CSF3* | EFLAPEGRASTIM | 9.14 | 7.86 | 9.75 | 8.94 | 158.70 |
| *CCR9* | VERCIRNON | 9.11 | 6.73 | 10.00 | 10.00 | 158.39 |
| *CCR9* | MLN3126 | 9.11 | 6.73 | 9.98 | 10.00 | 158.25 |
| *PDE4A* | CRISABOROLE | 8.74 | 9.20 | 8.50 | 9.14 | 158.14 |
| *LTF* | NIMESULIDE | 9.51 | 8.58 | 8.18 | 9.31 | 158.11 |
| *PDE4A* | PICLAMILAST | 8.74 | 9.20 | 8.46 | 9.14 | 157.80 |
| *PDE4A* | OXTRIPHYLLINE | 8.74 | 9.20 | 8.43 | 9.14 | 157.51 |
| *PDE4A* | REVAMILAST | 8.74 | 9.20 | 8.37 | 9.14 | 156.98 |
| *PDE4A* | MK-0873 | 8.74 | 9.20 | 8.37 | 9.14 | 156.98 |
| *PDE4A* | LIRIMILAST | 8.74 | 9.20 | 8.37 | 9.14 | 156.98 |
| *PDE4A* | CHEMBL74078 | 8.74 | 9.20 | 8.37 | 9.14 | 156.98 |
| *CSF3* | MUPLESTIM | 9.14 | 7.86 | 9.50 | 8.94 | 156.62 |
| *IFNAR2* | INTERFERON ALFA-N3 | 5.79 | 10.00 | 9.82 | 9.92 | 155.81 |
| *PDE4A* | IBUDILAST | 8.74 | 9.20 | 8.20 | 9.14 | 155.47 |
| *CYP3A43* | CHLORZOXAZONE | 9.97 | 8.82 | 9.18 | 7.36 | 155.43 |
| *IFNAR2* | INTERFERON ALFA-2B | 5.79 | 10.00 | 9.65 | 9.92 | 154.46 |
| *ICAM1* | HYALURONATE SODIUM | 9.25 | 9.38 | 7.76 | 8.80 | 154.28 |
| *LTF* | RESERPINE | 9.51 | 8.58 | 7.69 | 9.31 | 153.68 |
| *PDE4A* | TOFISOPAM | 8.74 | 9.20 | 7.98 | 9.14 | 153.46 |

| | | | | | | |
|---|---|---|---|---|---|---|
| *PDE4A* | MILRINONE | 8.74 | 9.20 | 7.98 | 9.14 | 153.46 |
| *CSF3* | TALABOSTAT | 9.14 | 7.86 | 9.12 | 8.94 | 153.37 |
| *CYP3A43* | TICAGRELOR | 9.97 | 8.82 | 8.87 | 7.36 | 152.50 |
| *PDE4A* | TOFIMILAST | 8.74 | 9.20 | 7.83 | 9.14 | 152.14 |
| *PDE4A* | ROLIPRAM | 8.74 | 9.20 | 7.83 | 9.14 | 152.14 |
| *PDE4A* | DENBUFYLLINE | 8.74 | 9.20 | 7.83 | 9.14 | 152.14 |
| *PDE4A* | HT-0712 | 8.74 | 9.20 | 7.83 | 9.14 | 152.14 |
| *PDE4A* | GSK-356278 | 8.74 | 9.20 | 7.83 | 9.14 | 152.14 |
| *PDE4A* | PROPENTOFYLLI NE | 8.74 | 9.20 | 7.83 | 9.14 | 152.14 |
| *PDE4A* | ILOPROST | 8.74 | 9.20 | 7.83 | 9.14 | 152.14 |
| *CYP3A43* | COBICISTAT | 9.97 | 8.82 | 8.70 | 7.36 | 150.89 |
| *PDE4A* | THEOPHYLLINE | 8.74 | 9.20 | 7.63 | 9.14 | 150.31 |
| *IFNAR2* | PEGINTERFERON ALFA-2A | 5.79 | 10.00 | 8.95 | 9.92 | 148.94 |
| *IFNAR2* | NYLIDRIN | 5.79 | 10.00 | 8.95 | 9.92 | 148.94 |
| *IFNAR2* | INTERFERON ALFACON-1 | 5.79 | 10.00 | 8.95 | 9.92 | 148.94 |
| *IFNAR2* | PEGINTERFERON BETA-1A | 5.79 | 10.00 | 8.95 | 9.92 | 148.94 |
| *IFNAR2* | INTERFERON BETA-1B | 5.79 | 10.00 | 8.95 | 9.92 | 148.94 |
| *IFNAR2* | ANIFROLUMAB | 5.79 | 10.00 | 8.95 | 9.92 | 148.94 |
| *IFNAR2* | ALBINTERFERON ALFA-2B | 5.79 | 10.00 | 8.95 | 9.92 | 148.94 |
| *IFNAR2* | PEGINTERFERON ALFA-2B | 5.79 | 10.00 | 8.95 | 9.92 | 148.94 |
| *IFNAR2* | INTERFERON BETA-1A | 5.79 | 10.00 | 8.95 | 9.92 | 148.94 |
| *IFNAR2* | INTERFERON ALFA-2A | 5.79 | 10.00 | 8.95 | 9.92 | 148.94 |
| *PDE4A* | CDC-801 | 8.74 | 9.20 | 7.46 | 9.14 | 148.83 |
| *PDE4A* | CDP840 | 8.74 | 9.20 | 7.46 | 9.14 | 148.83 |
| *PDE4A* | TADALAFIL | 8.74 | 9.20 | 7.46 | 9.14 | 148.83 |
| *IFNAR2* | SIFALIMUMAB | 5.79 | 10.00 | 8.62 | 9.92 | 146.36 |
| *JAK1* | RUXOLITINIB | 8.45 | 9.22 | 8.81 | 7.75 | 146.35 |
| *JAK1* | ITACITINIB | 8.45 | 9.22 | 8.75 | 7.75 | 145.80 |
| *JAK1* | GLPG-0555 | 8.45 | 9.22 | 8.75 | 7.75 | 145.80 |
| *JAK1* | INCB-047986 | 8.45 | 9.22 | 8.75 | 7.75 | 145.80 |
| *VIPR2* | SECRETIN | 8.12 | 8.85 | 9.62 | 7.54 | 145.54 |
| *JAK1* | UPADACITINIB | 8.45 | 9.22 | 8.61 | 7.75 | 144.57 |
| *VIPR2* | VASOACTIVE INTESTINAL PEPTIDE | 8.12 | 8.85 | 9.50 | 7.54 | 144.57 |
| *JAK1* | PEFICITINIB | 8.45 | 9.22 | 8.57 | 7.75 | 144.21 |
| *CYP3A43* | KETOCONAZOLE | 9.97 | 8.82 | 7.99 | 7.36 | 144.21 |
| *PSORS1C 1* | ALLOPURINOL | 9.45 | 7.02 | 8.63 | 8.83 | 143.79 |
| *JAK1* | BARICITINIB | 8.45 | 9.22 | 8.52 | 7.75 | 143.71 |
| *JAK1* | FILGOTINIB | 8.45 | 9.22 | 8.52 | 7.75 | 143.71 |
| *JAK1* | SOLCITINIB | 8.45 | 9.22 | 8.52 | 7.75 | 143.71 |
| *JAK1* | ABROCITINIB | 8.45 | 9.22 | 8.48 | 7.75 | 143.38 |

| | | | | | | |
|---|---|---|---|---|---|---|
| *PDE4A* | FLAVOXATE HYDROCHLORIDE | 8.74 | 9.20 | 6.84 | 9.14 | 143.27 |
| *PDE4A* | PENTOXIFYLLINE | 8.74 | 9.20 | 6.78 | 9.14 | 142.74 |
| *CYP3A43* | MELATONIN | 9.97 | 8.82 | 7.79 | 7.36 | 142.35 |
| *LTA* | PATECLIZUMAB | 7.37 | 8.23 | 9.67 | 8.56 | 142.20 |
| *PDE4A* | THEOPHYLLINE SODIUM GLYCINATE | 8.74 | 9.20 | 6.63 | 9.14 | 141.42 |
| *PDE4A* | INAMRINONE | 8.74 | 9.20 | 6.63 | 9.14 | 141.42 |
| *JAK1* | BREPOCITINIB | 8.45 | 9.22 | 8.25 | 7.75 | 141.39 |
| *JAK1* | MOMELOTINIB | 8.45 | 9.22 | 8.25 | 7.75 | 141.39 |
| *JAK1* | FEDRATINIB | 8.45 | 9.22 | 8.25 | 7.75 | 141.39 |
| *JAK1* | TRICETAMIDE | 8.45 | 9.22 | 8.25 | 7.75 | 141.39 |
| *JAK1* | RUXOLITINIB PHOSPHATE | 8.45 | 9.22 | 8.25 | 7.75 | 141.39 |
| *PDE4A* | AMINOPHYLLINE | 8.74 | 9.20 | 6.59 | 9.14 | 141.00 |
| *PDE4A* | DIPYRIDAMOLE | 8.74 | 9.20 | 6.59 | 9.14 | 141.00 |
| *LTA* | BAMINERCEPT | 7.37 | 8.23 | 9.41 | 8.56 | 140.22 |
| *JAK1* | CHEMBL21156 | 8.45 | 9.22 | 8.10 | 7.75 | 140.06 |
| *JAK1* | TOFACITINIB | 8.45 | 9.22 | 7.93 | 7.75 | 138.50 |
| *JAK1* | XL-019 | 8.45 | 9.22 | 7.93 | 7.75 | 138.50 |
| *JAK1* | AZD-1480 | 8.45 | 9.22 | 7.93 | 7.75 | 138.50 |
| *JAK1* | CERDULATINIB | 8.45 | 9.22 | 7.93 | 7.75 | 138.50 |
| *JAK1* | DECERNOTINIB | 8.45 | 9.22 | 7.93 | 7.75 | 138.50 |
| *CYP3A43* | OLANZAPINE | 9.97 | 8.82 | 7.35 | 7.36 | 138.25 |
| *PSORS1C 1* | ETANERCEPT | 9.45 | 7.02 | 7.93 | 8.83 | 138.04 |
| *LTA* | ETANERCEPT | 7.37 | 8.23 | 9.07 | 8.56 | 137.58 |
| *PDE4A* | ETAZOLATE | 8.74 | 9.20 | 6.15 | 9.14 | 137.06 |
| *TYK2* | TOFACITINIB | 8.13 | 7.24 | 8.34 | 9.46 | 136.81 |
| *PDE4A* | AMLEXANOX | 8.74 | 9.20 | 6.05 | 9.14 | 136.18 |
| *TYK2* | BREPOCITINIB | 8.13 | 7.24 | 8.25 | 9.46 | 136.17 |
| *JAK1* | OCLACITINIB | 8.45 | 9.22 | 7.63 | 7.75 | 135.84 |
| *JAK1* | NS-018 | 8.45 | 9.22 | 7.63 | 7.75 | 135.84 |
| *JAK1* | BMS-911543 | 8.45 | 9.22 | 7.63 | 7.75 | 135.84 |
| *JAK1* | TOFACITINIB CITRATE | 8.45 | 9.22 | 7.63 | 7.75 | 135.84 |
| *JAK1* | DELGOCITINIB | 8.45 | 9.22 | 7.63 | 7.75 | 135.84 |
| *TYK2* | PEFICITINIB | 8.13 | 7.24 | 8.10 | 9.46 | 135.01 |
| *TYK2* | CHEMBL21156 | 8.13 | 7.24 | 8.10 | 9.46 | 135.01 |
| *PSORS1C 1* | CARBOPLATIN | 9.45 | 7.02 | 7.40 | 8.83 | 133.74 |
| *PSORS1C 1* | GEMCITABINE | 9.45 | 7.02 | 7.35 | 8.83 | 133.32 |
| *JAK1* | PACRITINIB | 8.45 | 9.22 | 7.32 | 7.75 | 133.18 |
| *JAK1* | PEMBROLIZUMAB | 8.45 | 9.22 | 7.26 | 7.75 | 132.58 |
| *LTA* | ABACAVIR | 7.37 | 8.23 | 8.40 | 8.56 | 132.28 |
| *TYK2* | TOFACITINIB CITRATE | 8.13 | 7.24 | 7.63 | 9.46 | 131.34 |
| *TYK2* | OCLACITINIB | 8.13 | 7.24 | 7.63 | 9.46 | 131.34 |
| *TYK2* | SOLCITINIB | 8.13 | 7.24 | 7.63 | 9.46 | 131.34 |

| | | | | | | |
|---|---|---|---|---|---|---|
| *TYK2* | BMS-911543 | 8.13 | 7.24 | 7.63 | 9.46 | 131.34 |
| *TYK2* | DELGOCITINIB | 8.13 | 7.24 | 7.63 | 9.46 | 131.34 |
| *TYK2* | NS-018 | 8.13 | 7.24 | 7.63 | 9.46 | 131.34 |
| *TYK2* | FILGOTINIB | 8.13 | 7.24 | 7.63 | 9.46 | 131.34 |
| *CPOX* | GIVOSIRAN | 9.01 | 9.09 | 9.96 | 4.50 | 130.84 |
| *TYK2* | CERDULATINIB | 8.13 | 7.24 | 7.50 | 9.46 | 130.37 |
| *JAK1* | ENZASTAURIN | 8.45 | 9.22 | 6.95 | 7.75 | 129.85 |
| *TYK2* | FEDRATINIB | 8.13 | 7.24 | 7.18 | 9.46 | 127.92 |
| *TYK2* | UPADACITINIB | 8.13 | 7.24 | 7.18 | 9.46 | 127.92 |
| *JAK1* | ZOTIRACICLIB | 8.45 | 9.22 | 6.71 | 7.75 | 127.79 |
| *LTA* | CARBAMAZEPINE | 7.37 | 8.23 | 7.76 | 8.56 | 127.36 |
| *PTGFR* | TRAVOPROST | 9.74 | 9.54 | 9.97 | 3.19 | 126.90 |
| *TYK2* | BARICITINIB | 8.13 | 7.24 | 6.95 | 9.46 | 126.13 |
| *PTGFR* | LATANOPROST | 9.74 | 9.54 | 9.84 | 3.19 | 125.62 |
| *PTGFR* | TAFLUPROST | 9.74 | 9.54 | 9.81 | 3.19 | 125.37 |
| *PTGFR* | BIMATOPROST | 9.74 | 9.54 | 9.79 | 3.19 | 125.14 |
| *TYK2* | ZOTIRACICLIB | 8.13 | 7.24 | 6.71 | 9.46 | 124.34 |
| *SPARC* | CALCIUM PHOSPHATE, TRIBASIC | 9.51 | 9.79 | 8.25 | 4.59 | 123.89 |
| *SPARC* | CALCIUM CITRATE | 9.51 | 9.79 | 8.25 | 4.59 | 123.89 |
| *TYK2* | RUXOLITINIB | 8.13 | 7.24 | 6.59 | 9.46 | 123.36 |
| *TYK2* | ALDESLEUKIN | 8.13 | 7.24 | 6.50 | 9.46 | 122.66 |
| *TYK2* | AT-9283 | 8.13 | 7.24 | 6.50 | 9.46 | 122.66 |
| *PTGFR* | LATANOPROSTENE BUNOD | 9.74 | 9.54 | 9.47 | 3.19 | 122.10 |
| *JAK1* | MYRICETIN | 8.45 | 9.22 | 6.05 | 7.75 | 121.92 |
